## Supplementary Materials for "Global seroprevalence of SARS-CoV-2 antibodies: a systematic review and meta-analysis"

### Supplementary Material

**Supplementary files**
**S1 File. PRISMA checklist**

| Section/topic | # | Checklist item | Reported on page # |
|---|---|---|---|
| **TITLE** | | | |
| Title | 1 | Identify the report as a systematic review, meta-analysis, or both. | 1 |
| **ABSTRACT** | | | |
| Structured summary | 2 | Provide a structured summary including, as applicable: background; objectives; data sources; study eligibility criteria, participants, and interventions; study appraisal and synthesis methods; results; limitations; conclusions and implications of key findings; systematic review registration number. | 2-3 |
| **INTRODUCTION** | | | |
| Rationale | 3 | Describe the rationale for the review in the context of what is already known. | 5-6 |
| Objectives | 4 | Provide an explicit statement of questions being addressed with reference to participants, interventions, comparisons, outcomes, and study design (PICOS). | 6 |
| **METHODS** | | | |
| Protocol and registration | 5 | Indicate if a review protocol exists, if and where it can be accessed (e.g., Web address), and, if available, provide registration information including registration number. | 6 |
| Eligibility criteria | 6 | Specify study characteristics (e.g., PICOS, length of follow-up) and report characteristics (e.g., years considered, language, publication status) used as criteria for eligibility, giving rationale. | 7-8 |
| Information sources | 7 | Describe all information sources (e.g., databases with dates of coverage, contact with study authors to identify additional studies) in the search and date last searched. | 6-7, S2 file, S4 file |
| Search | 8 | Present full electronic search strategy for at least one database, including any limits used, such that it could be repeated. | S2 Appendix file |
| Study selection | 9 | State the process for selecting studies (i.e., screening, eligibility, included in systematic review, and, if applicable, included in the meta-analysis). | 7-8 |
| Data collection process | 10 | Describe method of data extraction from reports (e.g., piloted forms, independently, in duplicate) and any processes for obtaining and confirming data from investigators. | 8-9, S4 file |
| Data items | 11 | List and define all variables for which data were sought (e.g., PICOS, funding sources) and any assumptions and simplifications made. | 9-11, S4 file |
| Risk of bias in individual studies | 12 | Describe methods used for assessing risk of bias of individual studies (including specification of whether this was done at the study or outcome level), and how this information is to be used in any data synthesis. | 8, S3 file |
| Summary measures | 13 | State the principal summary measures (e.g., risk ratio, difference in means). | 10-11 |
| Synthesis of results | 14 | Describe the methods of handling data and combining results of studies, if done, including measures of consistency (e.g., $I^2$) for each meta-analysis. | 9-11, S4 file |
| Risk of bias across studies | 15 | Specify any assessment of risk of bias that may affect the cumulative evidence (e.g., publication bias, selective reporting within studies). | 8, S3 file |
| Additional analyses | 16 | Describe methods of additional analyses (e.g., sensitivity or subgroup analyses, meta-regression), if done, indicating which were pre-specified. | 11, S4 file |
| **RESULTS** | | | |
| Study selection | 17 | Give numbers of studies screened, assessed for eligibility, and included in the review, with reasons for exclusions at each stage, ideally with a flow diagram. | 12, Fig 1 |
| Study characteristics | 18 | For each study, present characteristics for which data were extracted (e.g., study size, PICOS, follow-up period) and provide the citations. | 12, Table 1, Fig 2, Fig 3, S1 table, S2 table, S3 table |
| Risk of bias within studies | 19 | Present data on risk of bias of each study and, if available, any outcome level assessment (see item 12). | 12, Fig 3, S4 table |
| Results of individual studies | 20 | For all outcomes considered (benefits or harms), present, for each study: (a) simple summary data for each intervention group (b) effect estimates and confidence intervals, ideally with a forest plot. | S1 table, S2 table |

| | | | |
|---|---|---|---|
| Synthesis of results | 21 | Present results of each meta-analysis done, including confidence intervals and measures of consistency. | 20, 21, Table 5, S4 table, S6 table |
| Risk of bias across studies | 22 | Present results of any assessment of risk of bias across studies (see Item 15). | Fig 3 |
| Additional analysis | 23 | Give results of additional analyses, if done (e.g., sensitivity or subgroup analyses, meta-regression [see Item 16]). | 21, 22 Table 6 |
| **DISCUSSION** | | | |
| Summary of evidence | 24 | Summarize the main findings including the strength of evidence for each main outcome; consider their relevance to key groups (e.g., healthcare providers, users, and policy makers). | 23 |
| Limitations | 25 | Discuss limitations at study and outcome level (e.g., risk of bias), and at review-level (e.g., incomplete retrieval of identified research, reporting bias). | 27, 28 |
| Conclusions | 26 | Provide a general interpretation of the results in the context of other evidence, and implications for future research. | 29, 30 |
| **FUNDING** | | | |
| Funding | 27 | Describe sources of funding for the systematic review and other support (e.g., supply of data); role of funders for the systematic review. | 11 |

#### S2 File. Search strategy

Database: Ovid MEDLINE(R) and Epub Ahead of Print, In-Process & Other Non-Indexed Citations and Daily
Dates: January 1, 2020 to December 31, 2020
Notes: Covid-19 search terms were adapted from Ovid Expert Searches

| # | Search terms |
|---|---|
| 1 | exp Coronavirus/ |
| 2 | exp Coronavirus Infections/ |
| 3 | (coronavirus* or corona virus* or OC43 or NL63 or 229E or HKU1 or HCoV* or ncov* or covid* or sars-cov* or sarscov* or Sars-coronavirus* or Severe Acute Respiratory Syndrome Coronavirus*).tw,kf.[EB2] |
| 4 | or/1-3 |
| 5 | 4 not ((MERS or MERS-CoV or Middle East respiratory syndrome or camel* or dromedar* or equine or coronary or coronal or covidence* or covidien or influenza virus or HIV or bovine or calves or TGEV or feline or porcine or BCoV or PED or PEDV or PDCoV or FIPV or FCoV or SADS-CoV or canine or CCov or zoonotic or avian influenza or H1N1 or H5N1 or H5N6 or IBV).mp. or (animals/ not humans/)) |
| 6 | ((pneumonia or covid* or coronavirus* or corona virus* or ncov* or 2019-ncov or sars* or virus).tw,kf. or exp pneumonia/) and Wuhan.tw,kf. |
| 7 | (2019-ncov* or 2019nCov* or ncov19 or ncov-19 or 2019-novel CoV or sars-cov2* or sars-cov-2* or sarscov2* or sarscov-2* or Sars-coronavirus2 or Sars-coronavirus-2 or SARS-like coronavirus* or coronavirus 2 or coronavirus2* or corona or coronavirus-19 or covid19 or covid-19 or covid 2019 or ((novel or new or nouveau) adj2 (CoV or nCoV or covid or coronavirus* or corona virus or Pandemi*2)) or ((covid or covid19* or covid-19) and pandemic*2) or (coronavirus* and pneumonia)).tw,kf. |
| 8 | COVID-19.rx,px,ox. or severe acute respiratory syndrome coronavirus 2.os. |
| 9 | or/6-8 |
| 10 | 5 or 9 |
| 11 | immunoglobulins/ or antibodies/ or antibodies, blocking/ or exp antibodies, neutralizing/ or antibodies, viral/ or antigen-antibody complex/ or immune sera/ or exp immunoglobulin isotypes/ or immunoglobulin a/ or immunoglobulin d/ or immunoglobulin e/ or immunoglobulin g/ or immunoglobulin m/ |
| 12 | serologic tests/ or complement fixation tests/ or hemagglutination inhibition tests/ or neutralization tests/ |
| 13 | immunoassay/ or fluoroimmunoassay/ or exp immunoblotting/ or immunoenzyme techniques/ or exp enzyme-linked immunosorbent assay/ or exp enzyme-linked immunospot assay/ or immunosorbent techniques/ or serologic tests/ or complement fixation tests/ or hemagglutination inhibition tests/ or neutralization tests/ or Serology/di |
| 14 | (enzyme linked immunosorbent or enzyme-linked immunosorbent or ELISA or immunofluorescence or complement fixation or hemagglutination inhibition or immunoblot or western blot or neutrali*).tw,kf. |
| 15 | (antibod* or immunoglobulin* or immune globulin* or titer* or isotype* or IgG or IgM or IgA or neutrali* or sera or serum or serolog* or saliva).tw,kf. |
| 16 | or/11-14 |
| 17 | seroepidemiologic studies/ |
| 18 | incidence/ or prevalence/ |
| 19 | (seroconver* or seroprevalence or sero-prevalence or seroincidence or sero-incidence or seroepidemiolog* or sero-epidemiolog*).mp. |
| 20 | (inciden* or prevalen* or count* or rate*).mp. |
| 21 | (serosurvey or sero-survey or screen* or diagnostic).mp. |
| 22 | (seroconver* or seroprevalence or sero-prevalence or seroincidence or sero-incidence or seroepidemiolog* or sero-epidemiolog* or inciden* or prevalen* or silent or asymptomatic or serosurvey or sero-survey).tw,kf. |
| 23 | or/17-21 |
| 24 | 10 and (16 and 23) |
| 25 | 10 and 15 |
| 26 | 10 and 22 |
| 27 | or/24-26 |
| 28 | limit 27 to yr="2020-Current" |
| 29 | remove duplicates from 28 |

Database: Embase
Dates: January 1, 2020 to December 31, 2020
Notes: Covid-19 search terms were adapted from Ovid Expert Searches

| # | Searches |
|---|---|
| 1 | exp Coronavirus/ |
| 2 | exp Coronavirus Infections/ |
| 3 | (coronavirus* or corona virus* or OC43 or NL63 or 229E or HKU1 or HCoV* or ncov* or covid* or sars-cov* or sarscov* or Sars-coronavirus* or Severe Acute Respiratory Syndrome Coronavirus*).tw,kw. |
| 4 | or/1-3 |
| 5 | 4 not ((MERS or MERS-CoV or Middle East respiratory syndrome or camel* or dromedar* or equine or coronary or coronal or covidence* or covidien or influenza virus or HIV or bovine or calves or TGEV or feline or porcine or BCoV or PED or PEDV or PDCoV or FIPV or FCoV or SADS-CoV or canine or CCov or zoonotic or avian influenza or H1N1 or H5N1 or H5N6 or IBV).mp. or (animals/ not humans/)) |
| 6 | ((pneumonia or covid* or coronavirus* or corona virus* or ncov* or 2019-ncov or sars*).tw,kw. or exp pneumonia/) and Wuhan.tw,kw. |
| 7 | (2019-ncov or ncov19 or ncov-19 or 2019-novel CoV or sars-cov2 or sars-cov-2 or sarscov2 or sarscov-2 or Sars-coronavirus2 or Sars-coronavirus-2 or SARS-like coronavirus* or coronavirus-19 or covid19 or covid-19 or covid 2019 or ((novel or new or nouveau) adj2 (CoV or nCoV or covid or coronavirus* or corona virus or Pandemi*2)) or ((covid or covid19 or covid-19) and pandemic*2) or (coronavirus* and pneumonia)).tw,kw. |
| 8 | (coronavirus disease 2019 or severe acute respiratory syndrome coronavirus 2).sh,dj. |
| 9 | 6 or 7 or 8 |
| 10 | 5 or 9 |
| 11 | virus antibody/ec [Endogenous Compound] |
| 12 | neutralizing antibody/ec [Endogenous Compound] |
| 13 | exp immunoglobulin/ or exp immunoglobulin A antibody/ or exp immunoglobulin class/ or exp immunoglobulin M antibody/ or exp immunoglobulin G antibody/ or exp immunoglobulin antibody/ |
| 14 | 11 or 12 or 13 |
| 15 | serology/ |
| 16 | serodiagnosis/ or complement fixation test/ or hemagglutination inhibition test/ or hemolytic plaque assay/ |
| 17 | fluorescent antibody technique/ |
| 18 | immunofluorescence test/ or viral disease immunofluorescence assay/ |
| 19 | enzyme linked immunosorbent assay/ |
| 20 | western blotting/ |
| 21 | (enzyme linked immunosorbent or enzyme-linked immunosorbent or ELISA or immunoassay or immunofluorescence or fluorescent antibody or complement fixation or hemagglutination inhibition or hemolytic plaque assay or immunoblot or western blot or neutrali*).tw,kw. |
| 22 | (antibod* or immunoglobulin* or immune globulin* or titer* or isotype* or IgG or IgM or IgA or neutrali* or sera or serolog* or serum or saliva).tw,kw. |
| 23 | 15 or 16 or 17 or 18 or 19 or 20 or 21 |
| 24 | 14 or 23 |
| 25 | exp seroepidemiology/ |
| 26 | *prevalence/ |
| 27 | *incidence/ |
| 28 | (seroconver* or seroprevalence or sero-prevalence or seroincidence or sero-incidence or seroepidemiolog* or sero-epidemiolog* or inciden* or prevalen* or count* or rate* or serosurvey or sero-survey or screen* or diagnostic).mp. |
| 29 | (seroconver* or seroprevalence or sero-prevalence or seroincidence or sero-incidence or seroepidemiolog* or sero-epidemiolog* or inciden* or prevalen* or silent or asymptomatic or serosurvey or sero-survey).tw,kw. |
| 30 | 25 or 26 or 27 or 28 |
| 31 | 10 and (24 and 30) |
| 32 | 10 and 22 |
| 33 | 10 and 29 |
| 34 | 31 or 32 or 33 |
| 35 | limit 34 to yr="2020-Current" |
| 36 | remove duplicates from 35 |

Database: Web of Science Core Collection
Date: January 1, 2020 to December 31, 2020

| # | Searches |
|---|---|
| 1 | TS=(coronavirus* or corona virus* or OC43 or NL63 or 229E or HKU1 or HCoV* or ncov* or covid* or sars-cov* or sarscov* or Sars-coronavirus* or Severe Acute Respiratory Syndrome Coronavirus*) |
| 2 | TS=(MERS or MERS-CoV or Middle East respiratory syndrome or camel* or dromedar* or equine or coronary or coronal or covidence* or covidien or influenza virus or HIV or bovine or calves or TGEV or feline or porcine or BCoV or PED or PEDV or PDCoV or FIPV or FCoV or SADS-CoV or canine or CCov or zoonotic or avian influenza or H1N1 or H5N1 or H5N6 or IBV) |
| 3 | #1 NOT #2 |
| 4 | TS=((pneumonia or covid* or coronavirus* or corona virus* or ncov* or 2019-ncov or sars* or virus) AND Wuhan) |
| 5 | TS=(2019-ncov* or 2019nCov* or ncov19 or ncov-19 or 2019-novel CoV or sars-cov2* or sars-cov-2* or sarscov2* or sarscov-2* or Sars-coronavirus2 or Sars-coronavirus-2 or SARS-like coronavirus* or corona or coronavirus-19 or covid19 or covid-19 or covid 2019 or ((novel or new or nouveau) adj2 (CoV or nCoV or covid or coronavirus*) ) or (coronavirus* and pneumonia) ). |
| 6 | TS=(COVID-19 or "severe acute respiratory syndrome coronavirus") |
| 7 | #6 OR #5 OR #4 OR #3 |
| 8 | TS=(antibod* or immunoglobulin* or immune globulin* or titer* or isotype* or IgG or IgM or IgA or neutralization or sera or serolog* or saliva or serum). |
| 9 | TS=("enzyme linked immunosorbent assay" or "enzyme-linked immunosorbent assay" or "immunoenzyme" or ELISA or "lateral flow immunoassay" or LFIA or "immunofluorescence assay" or immunochromatography or "complement fixation test" or "hemagglutination inhibition" or immunoblot or "western blot" or "neutralization assay") |
| 10 | #9 OR #8 |
| 11 | TI=(seroconversion or seroprevalence or seroincidence or seroepidemiolog* or incidence or prevalence or asymptomatic or sero-survey*) or AK=(seroconversion or seroprevalence or seroincidence or seroepidemiolog* or incidence or prevalence or asymptomatic or sero-survey*) |
| 12 | ALL=(prevalence or incidence or seroconversion or seroconvert or seroprevalence or seroincidence or seroepidemiolog* or serosurvey or sero-survey or survey or screen* or diagnostic test) |
| 13 | #12 AND #10 AND #7 |
| 14 | #11 AND #7 |
| 15 | TI=(antibod* or immunoglobulin* or immune globulin* or titer* or isotype* or IgG or IgM or IgA or neutralization or sera or serolog* or saliva or serum). |
| 16 | #15 AND #7 |
| 17 | #16 OR #14 OR #13 |

Database: Europe PMC [Secondary search for pre-prints]
Dates: January 1, 2020 to December 31, 2020

| # | Searches |
|---|---|
| | ("2019-nCoV" OR "2019nCoV" OR "COVID-19" OR "SARS-CoV-2" OR "COVID19" OR "COVID" OR "SARS-nCoV" OR ("wuhan" AND "coronavirus") OR "Coronavirus" OR "Corona virus" OR "corona-virus" OR "corona viruses" OR "coronaviruses" OR "SARS-CoV" OR "Severe Acute Respiratory Syndrome Coronavirus" OR ("SARS" AND "coronavirus")) AND ABSTRACT:(sera* OR sero* OR immun* OR Ig* OR “enzyme-linked immunosorbent assay” OR ELISA OR “neutralization assay" OR seroprevalence) AND (SRC:"PPR") |

**Sources: Health organizations**
Dates: January 1, 2020 to December 31, 2020

| Source | Search strategy | |
|---|---|---|
| WHO Situation Reports | 1 | “antibod”, “sero”, “immun”, “ELISA” |
| National Institutes of Health | 1 | ("COVID" OR "SARS-CoV-2") |
| | 2 | ("sero*" OR "antibod*" OR "immun*" OR "RDT" OR "ELISA" OR "LFIA") |
| | 3 | allintext:(1 AND 2) site:nih.gov -site:ncbi.nlm.nih.gov |
| | 3 | 2 AND 3 |
| United States Centres for Disease Control and Prevention | 1 | ("COVID" OR "SARS-CoV-2") |
| | 2 | ("sero*" OR "antibod*" OR "immun*" OR "RDT" OR "ELISA" OR "LFIA") |
| | 3 | allintext:(1 AND 2) site:cdc.gov |
| | 5 | 2 AND 3 |
| European Centres for Disease Control and Prevention | 1 | ("COVID" OR "SARS-CoV-2") |
| | 2 | ("sero*" OR "antibod*" OR "immun*" OR "RDT" OR "ELISA" OR "LFIA") |
| | 3 | allintext:(1 AND 2) site:ecdc.europa.eu |
| | 5 | 2 AND 3 |

**Sources: Google News**
Dates: January 1, 2020 to December 31, 2020

| Source | Search strategy | |
|---|---|---|
| Google news | 1 | (antibody OR antibodies OR surveillance OR screen OR serology OR serological OR serosurvey OR ELISA OR LFIA OR assay OR blood OR serum OR immune OR immunity OR herd immunity OR random test) |

#### S3 File. Tool for assessing study risk of bias

| **Item 1: Was the sample frame appropriate to address the target population?** | |
|---|---|
| Yes | Sample frame described and it approximated the target population |
| No | Sample frame did not approximate the target population (e.g., blood donors do not represent general population, doctors do not represent all health care providers) |
| Exclude | Sample frame not described |
| *Notes | The term "target population" should not be taken to infer every individual from everywhere or with similar disease or exposure characteristics. Instead, give consideration to specific population characteristics in the study, including age range, gender, morbidities, medications, and other potentially influential factors. For example, a sample frame may not be appropriate to address the target population if a certain group has been used (such as those working for one organisation, or one profession) and the results then inferred to the target population (i.e. working adults). A sample frame may be appropriate when it includes almost all the members of the target population (i.e. a census, or a complete list of participants or complete registry data). |

| **Item 2: Were study participants recruited in an appropriate way?** | |
|---|---|
| Yes | Probability sampling method (simple or stratified random) or entire sample (e.g., an entire town) was used |
| No | Non-probability sampling |
| Exclude | Sampling method not reported |

| **Item 3: Was the sample size adequate?** | |
|---|---|
| Yes | $\geq$599 |
| No | <599 |
| Exclude | Sample size not reported |
| *Notes | To calculate the required sample size we used an assumed prevalence of 2.5%, which was the global average estimated by the WHO in April, 2020.[1] Based on guidance by the Joanna Briggs Institute and published medical statistical recommendations we selected a precision value that was half the assumed prevalence (1.25%)[2,3] We calculated a minimum sample size of 599 using these inputs:<br>Sample size calculation: $n = \frac{Z^2 P(1-P)}{d^2}$<br>Where n = sample size;<br>Z = Z statistic for level of confidence (95%);<br>P = expected prevalence (2.5% WHO global estimate);<br>d = precision (1.25%)<br><br>In cases where the sample size calculation was provided and the required sample for 80% power was below our threshold (n<599), this item was marked as yes. |

| **Item 4: Were the study subjects and setting described in detail?** | |
|---|---|
| Yes | Average age and distribution of gender/sex provided |
| No | Neither age or gender/sex is provided, or only one of age and gender/sex is provided |

| Item 5: Was data analysis conducted with sufficient coverage of the identified sample? | |
|---|---|
| Yes | The demographic characteristics (gender/sex, age, and ethnicity) of the sample is at least somewhat representative of the population |
| No | The demographic characteristics (gender/sex, age, and ethnicity) of the sample is not representative of the population |
| Unclear | Information is not provided about demographic characteristics of the sample (gender/sex, age, and ethnicity) |

| Item 6: Were valid methods used for the identification of the condition? | |
|---|---|
| Yes | The test used met the FDA standards for Emergency Use Authorizations for COVID-19 serological tests: sensitivity minimum 90%, specificity minimum 95%, as reported in the study.[4] |
| No | The test used did not meet the FDA standards for Emergency Use Authorizations for COVID-19 serological tests: sensitivity minimum 90%, specificity minimum 95%. |
| Exclude | Test sensitivity and specificity not reported |

| Item 7: Was the condition measured in a standard, reliable way for all participants? | |
|---|---|
| Yes | The same serology test was used for all participants |
| No | Different serology tests were used for participants |
| Unclear | No details were provided about which participants received which serology tests |

| Item 8: Was there appropriate statistical analysis? | |
|---|---|
| Yes | Does all of the following: corrects for population characteristics or the sample is somewhat representative of the population (probability sampling), corrects for test characteristics), and provides the information necessary to determine the numerator, denominator, prevalence estimate, and confidence interval. |
| No | Does not correct for population characteristics and the sample is not likely representative of the population (non-probability sampling), does not correct for test or provide the information necessary to correct for test characteristics, or does not provide the information necessary to determine the numerator, denominator, prevalence estimate, and confidence interval. |

| Item 9: Was the response rate adequate, and if not, was the low response rate managed appropriately? | |
|---|---|
| Yes | Response rate > 60% or the demographics of the sample were a reasonable match to those of the target population[5] |
| No | Response rate < 60% and the demographics of the sample were not a reasonable match to those of the target population |
| Unclear | Response rate not provided and it was unclear if the demographics of the sample differed from the target population |

| Item 10: Overall risk of bias | |
|---|---|
| Low | The estimates are very likely correct for the target population. To obtain a low risk of bias classification, all criteria must be met or departures from the criteria must be minimal and unlikely to impact on the validity and reliability of the prevalence estimate. These include sample sizes that are just below the threshold when all other criteria are met, reporting only some of characteristics of the sample, test characteristics below the threshold but corrections for the test performance, and response rates that are just below the threshold in the context of probability based sampling of an appropriate sampling frame with population weighted seroprevalence estimates. |
| Moderate | The estimates are likely correct for the target population. To obtain a moderate risk of bias classification, most criteria must be met and departures from the criteria are likely to have only a small impact on the validity and reliability of the prevalence estimates. |
| High | The estimates are not likely correct for the target population. To obtain a high risk of bias, many criteria must not be met or departures from criteria are likely to have a major impact on the validity and reliability of the prevalence estimates. |
| Unclear | There was insufficient information to assess the risk of bias. |

#### S4 File. Additional data analysis details

**Data Processing and Descriptive Statistics**
Data processing and descriptive statistics were conducted in Python. The packages used were as follows: data manipulation: numpy, pandas, geopandas; Bayesian analysis: PyStan, arviz; regression modelling: statsmodels; data visualization: matplotlib, seaborn.

**Grouping variables for analysis**
Each study was grouped according to Global Burden of Disease (GBD) region[6,7]

**Age**
Estimates were classified into four groups based on the reported ages of the included study participants: children and youth (0-17), adults (18-64), and seniors (65+). The following rules were used to categorize estimates when studies included participants of different ages:

- Participant age mean/median: select the category that it falls within
    - Participant age mean +/- SD: selected all categories within one standard deviation on either side of the mean
    - Participant age median and interquartile range: selected all categories that were included in the interquartile range
- Total age ranges: The age range needed to cover 30% or more of an age category to be classified in that category. As such, thresholds defining 30% for each category were defined.
    - Children and youth: Include "Children and youth" tag if age minimum was 12 or lower.
    - Adults: Include "Adults" tag if age minimum was 57 of lower
    - Adults: Include "Adults" tag if age maximum was 25 or higher.
    - Adults: Include "Adults" tag if age not reported but sample frame was health care workers
    - Seniors: Include "Seniors" tag if age maximum was 70 or higher.
        - For the seniors category, an upper limit of 72 was used (WHO reported world-wide life expectancy) in order to determine a 30% overlap[8]

**Dates**
When explicit dates were not provided, we inferred dates based on the reporting sampling where possible using the following rules:

- Description of a single month: selected first and last date of month
    - Example: “Samples were gathered in April” = April 1 - 30
- Description of a full month as part of a range: selected mid-month
    - Example: “Samples were gathered from April to June” = April 15 to June 15
- Description of early month: selected first two weeks as start and end end (14 days)
    - Example: “Samples were gathered in early April” = April 1 to April 13
- Description of mid-month: selected middle two weeks (14 days)
    - Example: “Samples were gathered in mid April” = April 6 to April 19
- Description of late month: selected the last two weeks as start and end date (14 days)
    - Example: “Samples were gathered in late April” = April 17 to April 30

**Test correction**
The sensitivity and specificity values for correction were derived, in order of preference, from: (i) the FINDDx-McGill database of independent evaluations of serological tests[9]; (ii) independent test evaluations conducted by serosurvey investigators and reported alongside serosurvey findings; (iii) manufacturer-reported sensitivity and specificity; (iv) published pooled sensitivity and specificity by immunoassay type.[10] If uncorrected estimates were unavailable, we used author-reported corrected seroprevalence estimates in lieu of performing our own correction. When none of the above corrections were possible, we excluded estimates from further analysis. Details of this order of priority follow:

1. The FINDDx-McGill database of independent evaluations of serological tests.[9] We only considered evaluations reporting both sensitivity and specificity for test performance across all sickness days (as opposed to day 1, day 5, etc). Where multiple evaluations were available, we prioritized in the following order:
    a. The evaluation needed to match the test name, manufacturer, and target isotype used in the study

b. The evaluation needed to match the sample specimen type used in the study
    i. For sample types that were not reported in either the test evaluation or the serosurvey study we assumed whole blood was used for LFIA tests and serum/plasma was used for non-LFIA tests
    ii. Plasma/Serum were used interchangeable when no direct match for index sample type was available
c. Prioritized reference specimen type which yield the most virus according to a systematic review and meta-analysis published in July 2020 that compared RT-PCR positivity of different specimens[11]
    i. It was assumed that "respiratory specimen" was referring to upper respiratory specimen as a conservative assumption as these viral loads are lower than bronchoalveolar lavage or sputum. We ranked it along side throat swab.
    ii. "Lower respiratory specimen" was ranked with with bronchoalveolar lavage fluid
    iii. "Upper respiratory specimen" was ranked with throat swab
    iv. If a mixed reference sample was used in the independent evaluation then an even distribution of sample types was assumed; the average % yield of the viral load was calculated and the sample type was ranked accordingly
d. Largest sample size

2. Serological test evaluations conducted by study authors, where those authors were at arms-length from the design of the study in questions
3. Manufacturer-reported sensitivity and specificity, which includes evaluations of in-house serological tests published by the research group that developed the same test
4. Published pooled sensitivity and specificity results for ELISAs, LFIAs, and CLIAs, based on the test type known to have been used, and using the definitions for these test types provided in the cited article.[10]

Where evaluation sample sizes were not reported (primarily for manufacturer-conducted evaluations), we used the minimum required sample sizes for US Food and Drug Administration Emergency Use Authorization approval (30 true positive samples, 80 true negative samples).

Using the best available sensitivity and specificity values and the corresponding denominators, we corrected seroprevalence estimates for test sensitivity and specificity using Bayesian measurement error models, with binomial test evaluation data for test sensitivity and specificity, as described in Section 2 of Gelman and Carpenter (2020).[12] We built the Bayesian model described therein using the statistical package PyStan.

For each seroprevalence estimate, we used the matching seroprevalence, sensitivity, and specificity, and the corresponding denominators, to fit the specified model. The model was fit with four chains and 2000 iterations per chain (1000 warmup, 1000 sampling iterations). A model fit was considered "good" if said fit met all five of PyStan's Hamiltonian Monte Carlo diagnostic criteria (no iterations ending with a divergence, no iterations saturating the max tree depth of 10, effective Bayesian fraction of missing information over 0.2 for all chains, a R-hat value between 0.9 and 1.1 for all parameters, the effective sample size n_eff being at least 0.001x the number of iterations).[13] Model fits not meeting these criteria were discarded, and the model was rerun until five good fits were obtained for each seroprevalence estimate.

Corrected seroprevalence values were obtained for each model fit by taking the expectation of the true prevalence parameter across all sampling iterations. For each seroprevalence estimate, the corrected seroprevalence was taken to be the median corrected seroprevalence across the five good model fits. The credible interval was defined as the highest posterior density interval (equivalent to the shortest credible interval for these unimodal distributions), and was calculated using the hpd function from the arviz package.

**Selecting summary seroprevalence estimates**

Some studies did not report a single overall seroprevalence estimate. In most cases, this was because the study reported different sets of results for different non-overlapping subsets of the data: e.g., for multiple non-overlapping time points (e.g., May and June 2020), multiple distinct populations (e.g., teachers and schoolchildren), or multiple non-overlapping regions (e.g., two regions of the same city). Articles that provided information on two or more distinct cohorts (different sample frames or different samples at different time points) without a pooled estimate were considered to be multiple studies.

In some cases, studies did not report a single overall seroprevalence estimate because they reported different sets of results for different assays. When studies reported estimates derived from multiple assays used in combination, we prioritized a sub-group estimate based on a single test using the following prioritization (i) independent evaluation available; (ii) pan-Ig; IgG +/- IgM/IgA; IgM+/- IgA; IgA; (iii); highest combined sensitivity and specificity value. These prioritization criteria were designed to minimize bias: using data from independent evaluations is the most reliable way to correct seroprevalence estimates for test sensitivity and specificity; non-IgG antibodies appear to decline over time, especially among individuals without repeated exposure, while IgG antibodies against the spike protein appear to persist for at least three to five months after infection; and estimates that use tests with the highest sensitivity and specificity likely exhibit the least bias to begin with.[14–17]

**Population differences in seroprevalence**
Male and female estimates were included in the sex/gender analysis, with Female as the reference group. Estimates from Caucasian, Black, Asian, Indigenous, and Multi-racial/other racial groups were included for the race/ethnicity analysis, with Caucasian as the reference group. Children and youth (0-17), adult (18-64), and older adult (65+) estimates were included for the age analysis, with adults as the reference group. Health care workers and caregivers and the general population were included in the occupation analysis. Known and no known contact with a positive COVID-19 case were included in the exposure level analysis.

#### S5 File. Methods for selecting and gathering data on cumulative incidence and population size

**Evidence-based timeline for COVID-19 clinical course and cumulative case data collection**

- Day 0: Symptom onset
  - FYI: Exposure occurs a mean of -5 days from symptom onset[15]
- Day 5: Confirmation on pneumonia
  - Median of 5 days from symptom onset to confirmation of pneumonia[18,19]
  - This is the assumed date of diagnosis
- Day 7: Presentation to hospital
  - Median of 7 days from symptom onset to presentation to hospital[20]
  - Alternative assumption for day of diagnosis
- Day 14: Seroconversion
  - Median of 14 days from symptom onset to seroconversion (IgG)[21]

**Data collection decisions**

The cumulative case date of -9 days before serosurvey sampling end date was selected for analyses based on the evidence above. This assumes that the day of radiological confirmation of pneumonia is similar to the reporting date for laboratory confirmed cases of COVID-19, accounting for reporting delays. This corresponds to a median of 5 days after symptom onset and assumes a median of 14 days from symptom onset to seroconversion

**Sources of data on cumulative incidence and population size**

For studies reporting population-wide seroprevalence estimates, we gathered the number of total confirmed SARS-CoV-2 infections at nine days before the study end-date. We extracted case and population data at the country level for national studies. Our sources were the Johns Hopkins University (JHU) COVID-19[22] tracker with population denominators from the UN Population Division estimates.[23] Using these data we calculated cases per 100,000 population for each country.

#### Supplementary tables

#### S1 Table. Characteristics and primary results of studies reporting population-wide seroprevalence estimates

| Article Author (Study Organization) | Sampling Dates (YMD) | Location | Geographic Scope | Overall Risk of Bias | Sample Size | Age | Female (%) | Sampling method | Sample frame | Uncorrected Seroprevalence (95% Confidence Interval) | Test Manufacturers \| Names \| Isotypes | Test Sens, Spec (Source) | Corrected Seroprevalence (95% Credible Interval) |
|---|---|---|---|---|---|---|---|---|---|---|---|---|---|
| **Central Europe, Eastern Europe, and Central Asia** | | | | | | | | | | | | | |
| **Bulgaria** | | | | | | | | | | | | | |
| Tsaneva-Damyanova (Medical Diagnostic Laboratory "STATUS")[2] | 2020-03-26 to 2020-04-20 | Varna \| Varna | Local | High | 586 | NR | 56.1 | Non-probability | Residual sera | 4.8% (3.2-6.8%) | "Zhejiang Orient Gene Biotech Co.; LTD" \| COVID-19 IgG / IgM Rapid Test Cassette \| IgG, IgM | 0.4%, 1.0% (FINDDx / MUHC independent evaluation) | - |
| **Czechia** | | | | | | | | | | | | | |
| Bloomfield (Charles University and Thomayer's Hospital)[7] | 2020-07-03 to 2020-08-19 | Prague \| Prague | Sublocal | High | 200 | 0-12 months (n=13); 1-5 years (n=52); 6-11 years (n=65); 12-18 years (n=70) | 45.5 | Non-probability | Residual sera | 0.0% (0.0-0.5%) | Roche Diagnostics \| Elecsys Anti-SARS-CoV-2 assay \| NR | 99.5%, 99.8% (Used author-adjusted estimate) | 0.0% (0.0-1.2%) |
| **Estonia** | | | | | | | | | | | | | |
| Jogi (University of Tartu)[9] | 2020-05-08 to 2020-07-31 | Saaremaa | Local | Moderate | 954 | 0-9 (n=101); 10-19 (N=110); 20-29 (n=104); 30-39 (n=121); 40-49 (n=114); 50-59 (n=113); 60-69 (n=112); 70-79 (n=112); 80-100 (n=67) | 61.7 | Probability | Household and community samples | 6.3% (5.0-7.9%) | Abbott; DiaSorin; SD Biosensor \| Abbott Architect SARS-CoV-2 IgG, Liaison DiaSorin IgG, Biosensor IgM/IgG test, Neutralization assays \| IgG, IgM | No data | - |
| Jogi (University of Tartu)[9] | 2020-05-08 to 2020-07-31 | Tallinn | Local | Moderate | 1006 | 0-9 (n=106); 10-19 (n=107); 20-29 (n=111); 30-39 (n=117); 40-49 (n=120); 50-59 (n=117); | 55.9 | Probability | Household and community samples | 1.5% (0.9-2.5%) | Abbott; DiaSorin; SD Biosensor \| Abbott Architect SARS-CoV-2 | No data | - |

| Article Author (Study Organization) | Sampling Dates (YMD) | Location | Geographic Scope | Overall Risk of Bias | Sample Size | Age | Female (%) | Sampling method | Sample frame | Uncorrected Seroprevalence (95% Confidence Interval) | Test Manufacturers \| Names \| Isotypes | Test Sens, Spec (Source) | Corrected Seroprevalence (95% Credible Interval) |
|---|---|---|---|---|---|---|---|---|---|---|---|---|---|
| | | | | | | 60-69 (n=116); 70-79 (n=114); 80-100 (n=98) | | | | | IgG, Liaison DiaSorin IgG, Biosensor IgM/IgG test, Neutralization assays \| IgG, IgM | | |
| **Georgia** | | | | | | | | | | | | | |
| Tsertsvadze (Infectious Diseases, AIDS and Clinical Immunology Research Center)[11] | 2020-05-18 to 2020-05-27 | Tbilisi \| Tbilisi | Local | Moderate | 1086 | 18-64 (n=964); 65+ (n=104) | 62.8 | Non-probability | Household and community samples | 1.0% (0.4-2.2%) | Zhejiang Orient Gene Biotech Co Ltd \| COVID 19 IgG/IgM Rapid Test Cassette \| IgG | 93.1%, 99.2% (Used author-adjusted estimate) | 1.0% (0.5-1.7%) |
| **Hungary** | | | | | | | | | | | | | |
| Merkely (Semmelweis University)[12] | 2020-05-01 to 2020-05-16 | | National | Moderate | 10474 | Mean 48.7 (SD 18) | 53.6 | Probability | Household and community samples | 0.7% (0.5-0.9%) | Abbott \| Abbott Architect SARS-CoV-2 IgG assay \| IgG | 0.7%, 1.0% (FINDDx / MUHC independent evaluation) | - |
| **Russian Federation** | | | | | | | | | | | | | |
| Popova (Saint-Petersburg Pasteur Research Institute)[17] | 2020-06-28 to 2020-07-19 | Irkutsk Region | Regional | Moderate | 2674 | 1-6 (n=102), 7-13 (n=191), 14-17 (n=94), 18-29 (n=385), 30-39 (n=373), 40-49 (n=374), 50-59 (n=386), 60-69 (n=390), Above 70 (n=379) | 67.5 | Probability | Household and community samples | 5.8% (5.3-6.2%) | State Scientific Center Applied Microbiology and Biotechnology \| IFa anti-SARS-CoV-2 IgG \| IgG | 75.6%, 97.6% (Used Bastos SR/MA data; no sens, spec, or author adjustment available) | 4.5% (2.8-6.3%) |
| Popova (Moscow Research Insistute of Epidemiology and Microbiology named after V.I G.N Gabrichevsky)[18] | 2020-06-22 to 2020-07-11 | Central Russia \| Moscow Region | Regional | Unclear | 2688 | 1-6 (n=111); 7-13 (n=185); 14-17 (n=88); 18-29 (n=384); 30-39 (n=384); 40-49 (n=384); 50-59 (n=384); 60-69 (n=384); 70+ (n=384) | 71.0 | Probability | Household and community samples | 21.0% | FBSI "State scientific center for applied microbiology and biotechnology" \| ELISA anti-SARS-CoV-2 IgG \| IgG | 75.6%, 97.6% (Used Bastos SR/MA data; no sens, spec, or author adjustment available) | 25.4% (23.0-27.8%) |

| Article Author (Study Organization) | Sampling Dates (YMD) | Location | Geographic Scope | Overall Risk of Bias | Sample Size | Age | Female (%) | Sampling method | Sample frame | Uncorrected Seroprevalence (95% Confidence Interval) | Test Manufacturers \| Names \| Isotypes | Test Sens, Spec (Source) | Corrected Seroprevalence (95% Credible Interval) |
|---|---|---|---|---|---|---|---|---|---|---|---|---|---|
| Popova (Saint-Petersburg Pasteur Research Institute of Epidemiology and Microbiology) [19] | 2020-06-23 to 2020-06-26 | Northwestern Federal District | Regional | High | 3130 | 1-17 (n=401), 18-29 (n=464), 30-39 (n=474), 40-49 (n=510), 50-50 (n=480), 60-69 (n=422), 70+ (n=379) | NR | Probability | Household and community samples | 20.7% | State Scientific Center of Applied Microbiology and Biotechnology (Obolensk) \| NR \| IgG | 75.6%, 97.6% (Used Bastos SR/MA data; no sens, spec, or author adjustment available) | 24.9% (22.5-27.3%) |
| Barchuk (European University At St Petersburg)[20] | 2020-05-27 to 2020-06-26 | Saint Petersburg | Local | Low | 988 | 18-34 (n=396); 35-49 (n=357); 50-64 (n=218); 65+ (n=67) | 66.0 | Probability | Household and community samples | 9.1% (7.2-10.9%) | Genetico \| CoronaPass total antibodies test \| NR | 98.7%, 100.0% (Used author-adjusted estimate) | 9.1% (7.3-10.9%) |
| Popova (Saint-Petersburg Pasteur Research Institute of Epidemiology and Microbiology) [21] | 2020-06-15 to 2020-06-20 | Saint-Petersburg | Local | High | 2713 | 1-17 (n=377); 18=29 (n=390); 30-39 (n=416); 40-49 (n=395); 50-59 (n=413); 60-69 (n=442); >=70 (n=280) | NR | Probability | Household and community samples | 26.0% (24.3-27.7%) | State Scientific Center for Applied Microbiology and Biotechnologies (Obolensk) \| NR \| IgG | 75.6%, 97.6% (Used Bastos SR/MA data; no sens, spec, or author adjustment available) | 32.2% (29.5-34.9%) |
| Popova (Saint Petersburg Pasteur Institute)[22] | 2020-06-07 to 2020-06-20 | Tyumen region | Regional | High | 2758 | 1-17 (n=356), 18-29 (n=485), 30-39 (n=452), 40-49 (n=447), 50-59 (n=482), 60-69 (n=387), >70 (n=149) | NR | Probability | Household and community samples | 24.5% (22.9-26.1%) | State Research Center for Applied Microbiology and Biotechnology of Rospotrebnadzor \| NR \| IgG | 75.6%, 97.6% (Used Bastos SR/MA data; no sens, spec, or author adjustment available) | 30.1% (27.6-32.9%) |
| Tickle (City of Moscow)[23] | 2020-06-05 to 2020-06-18 | Central Russia \| Moscow | Local | High | 90000 | NR | NR | Probability | Household and community samples | 19.9% | NR \| NR \| NR | No data | - |
| Kiselyov (Russian National Council for Combating Coronavirus)[25] | Until 2020-06-10 | | National | Moderate | 650000 | NR | NR | Non-probability | Household and community samples | 14.0% | Invitro \| NR \| NR | 95.0%, 98.0% (Test developer / manufacturer evaluation) | 12.2% (6.9-16.0%) |
| **High-income** | | | | | | | | | | | | | |
| **Andorra** | | | | | | | | | | | | | |

| Article Author (Study Organization) | Sampling Dates (YMD) | Location | Geographic Scope | Overall Risk of Bias | Sample Size | Age | Female (%) | Sampling method | Sample frame | Uncorrected Seroprevalence (95% Confidence Interval) | Test Manufacturers \| Names \| Isotypes | Test Sens, Spec (Source) | Corrected Seroprevalence (95% Credible Interval) |
|---|---|---|---|---|---|---|---|---|---|---|---|---|---|
| Royo-Cebrecos (ISGlobal)[26] | 2020-05-04 to 2020-05-28 | | National | Moderate | 70494 | Mean 40.4 (SD 20) | 44.6 | Non-probability | Household and community samples | 11.0% | nCOV 2019 IgG/IgM- Zhuhai Livzon Diagnostics; Inc. \| Livzon® rapid test \| IgG, IgM | 0.7%, 0.6% (FINDDx / MUHC independent evaluation) | - |
| **Argentina** | | | | | | | | | | | | | |
| Muñoz (Ministerio de Salud de la Provincia de Buenos Aires)[27] | 2020-07-15 to 2020-07-16 | Buenos Aires \| Quilmes | Sublocal | Moderate | 284 | Mean 40 (NR) | 60.9 | Non-probability | Household and community samples | 14.8% (11.1-19.5%) | COVIDAR \| SEROKIT-ELISA COVIDAR IgG \| IgG | 95.0%, 100.0% (Test developer / manufacturer evaluation) | 15.5% (10.1-21.0%) |
| **Austria** | | | | | | | | | | | | | |
| Ladage (Danube Private University)[33] | 2020-06-01 to 2020-06-15 | Wösendorf, Weißenkirchen, Joching, St. Michael | Local | High | 835 | NR | 51.5 | Probability | Household and community samples | 8.5% | N/A - Author Designed \| N/A - Author Designed \| IgG | 75.6%, 97.6% (Used Bastos SR/MA data; no sens, spec, or author adjustment available) | 8.1% (5.4-10.9%) |
| Knabl ( Medical University of Innsbruck )[36] | 2020-04-21 to 2020-04-27 | Tyrol \| Ischgl | Local | Moderate | 1473 | Mean 40.3 (Male); 40.9 (Female); 10.2 (Children); 45.8 (Adults) (SD 19.5 (Male); 20.1 (Female); 4.9 (Children); 16.5 (Adults)) | 51.5 | Probability | Household and community samples | 42.4% (39.8-44.7%) | EUROIMMUN ; Abbott \| Abbott SARS-CoV-2 IgG immunoassay, Euroimmun Anti-SARS-CoV-2 IgG IgA ELISA \| IgG | No data | - |
| Wagner (Medical University of Vienna)[37] | 2020-04-02 to 2020-04-17 | Vienna | Sublocal | High | 1655 | 15-25 (n = 226) ; 25-50 (n = 1031) ; over 50 (n=395) ; ns (n=3) | 53.5 | Non-probability | Household and community samples | 10.2% | Euroimmun Medizinische Labordiagnostika \| Euroimmune® \| IgG, IgA | 0.9%, 0.8% (FINDDx / MUHC independent evaluation) | - |
| **Belgium** | | | | | | | | | | | | | |
| Herzog (University of Antwerp)[41] | 2020-06-29 to 2020-07-04 | Wallonia, Brussels, Flanders | National | Moderate | 3023 | 0-10 (n=110); 10-20 (n=413); 20-30 (n=394); | 51.3 | Non-probability | Household and | 4.5% (3.7-5.4%) | EUROIMMUN \| Euroimmun Anti-SARS- | 0.8%, 1.0% (FINDDx / MUHC | - |

| Article Author (Study Organization) | Sampling Dates (YMD) | Location | Geographic Scope | Overall Risk of Bias | Sample Size | Age | Female (%) | Sampling method | Sample frame | Uncorrected Seroprevalence (95% Confidence Interval) | Test Manufacturers \| Names \| Isotypes | Test Sens, Spec (Source) | Corrected Seroprevalence (95% Credible Interval) |
|---|---|---|---|---|---|---|---|---|---|---|---|---|---|
| | | | | | | 30-40 (n=396); 40-50 (n=403); 50-60 (n=400); 60-70 (n=403); 70-80 (n=204), 80-90 (n=160); >90 (n=140) | | | community samples | | CoV-2 ELISA IgG \| IgG | independent evaluation) | |
| Herzog (University of Antwerp)[41] | 2020-06-08 to 2020-06-13 | Wallonia, Brussels, Flanders | National | Moderate | 2960 | 0-10 (n=124); 10-20 (n=375); 20-30 (n=383); 30-40 (n=395); 40-50 (n=394); 50-60 (n=393); 60-70 (n=399); 70-80 (n=201), 80-90 (n=166); >90 (n=130) | 51.9 | Non-probability | Household and community samples | 5.5% (4.7-6.5%) | EUROIMMUN \| Euroimmun Anti-SARS-CoV-2 ELISA IgG \| IgG | 0.8%, 1.0% (FINDDx / MUHC independent evaluation) | - |
| Herzog (University of Antwerp)[41] | 2020-05-18 to 2020-05-25 | Wallonia, Brussels, Flanders | National | Moderate | 3242 | 0-10 (n=174); 10-20 (n=431); 20-30 (n=414); 30-40 (n=424); 40-50 (n=411); 50-60 (n=419); 60-70 (n=417); 70-80 (n=236), 80-90 (n=163); >90 (n=153) | 51.0 | Non-probability | Household and community samples | 6.9% (5.9-8.0%) | EUROIMMUN \| Euroimmun Anti-SARS-CoV-2 ELISA IgG \| IgG | 0.8%, 1.0% (FINDDx / MUHC independent evaluation) | - |
| Herzog (University of Antwerp)[41] | 2020-04-20 to 2020-04-26 | Wallonia, Brussels, Flanders | National | Moderate | 3397 | 0-10 (n=85); 10-20 (n=442); 20-30 (n=375); 30-40 (n=407); 40-50 (n=406); 50-60 (n=430); 60-70 (n=426); 70-80 (n=316), 80-90 (n=315); >90 (n=195) | 52.9 | Non-probability | Household and community samples | 6.0% (5.1-7.1%) | EUROIMMUN \| Euroimmun Anti-SARS-CoV-2 ELISA IgG \| IgG | 0.8%, 1.0% (FINDDx / MUHC independent evaluation) | - |
| Herzog (University of Antwerp)[41] | 2020-03-30 to 2020-04-05 | Wallonia, Brussels, Flanders | National | Moderate | 3910 | 0-10 (n=36); 10-20 (n=294); 20-30 (n=436); 30-40 (n=461); 40-50 (n=468); 50-60 (n=498); 60-70 (n=507); 70-80 (n=506), 80-90 (n=493); >90 (n=211) | 54.0 | Non-probability | Household and community samples | 2.9% (2.3-3.4%) | EUROIMMUN \| Euroimmun Anti-SARS-CoV-2 ELISA IgG \| IgG | 0.8%, 1.0% (FINDDx / MUHC independent evaluation) | - |

| Article Author (Study Organization) | Sampling Dates (YMD) | Location | Geographic Scope | Overall Risk of Bias | Sample Size | Age | Female (%) | Sampling method | Sample frame | Uncorrected Seroprevalence (95% Confidence Interval) | Test Manufacturers \| Names \| Isotypes | Test Sens, Spec (Source) | Corrected Seroprevalence (95% Credible Interval) |
|---|---|---|---|---|---|---|---|---|---|---|---|---|---|
| **Canada** | | | | | | | | | | | | | |
| Canadian Blood Services (Canadian Blood Services)[47] | 2020-11-07 to 2020-11-25 | | National | Moderate | 17049 | 17-24 (n=1,631); 25-39 (n=4644); 40-59 (n=6382); 60+ (n=4392) | 43.7 | Non-probability | Blood donors | 1.5% (1.3-1.7%) | Abbott \| Abbott Architect SARS-CoV-2 IgG assay \| IgG | 92.7%, 99.9% (Used author-adjusted estimate) | 1.5% (1.3-1.7%) |
| Canadian Blood Services (Canadian Blood Services)[48] | 2020-10-12 to 2020-10-31 | | National | Moderate | 16811 | 17-24 (n=1,491); 25-39 (n=4,535); 40-59 (n=6,446); 60+ (n=4,339) | 43.2 | Non-probability | Blood donors | 0.9% (0.7-1.0%) | Abbott \| Abbott Architect SARS-CoV-2 IgG assay \| IgG | 92.7%, 99.9% (Used author-adjusted estimate) | 0.9% (0.7-1.0%) |
| Manny (University of Alberta)[49] | 2020-08-14 to 2020-10-23 | Alberta \| Edmonton | Local | High | 565 | Mean 10.5 (SD 1.6) | 49.2 | Non-probability | Household and community samples | 1.6% | Abbott Laboratories \| ARCHITECT i2000SR \| IgG | 0.4%, 1.0% (FINDDx / MUHC independent evaluation) | - |
| Bolotin (Public Health Ontario)[51] | 2020-08-01 to 2020-08-21 | Ontario | Regional | High | 5764 | NR | NR | Non-probability | Residual sera | 1.6% | Abbott Laboratories \| Abbott Architecht Anti-SARS-CoV-2 IgG test \| IgG | 0.4%, 1.0% (FINDDx / MUHC independent evaluation) | - |
| Bolotin (Public Health Ontario)[51] | 2020-07-04 to 2020-07-31 | Ontario | Regional | High | 7001 | NR | NR | Non-probability | Residual sera | 1.4% | Abbott Laboratories \| Abbott Architecht Anti-SARS-CoV-2 IgG test \| IgG | 0.4%, 1.0% (FINDDx / MUHC independent evaluation) | - |
| Héma-Québec (Héma-Québec)[53] | 2020-05-25 to 2020-07-09 | Quebec | Regional | Unclear | 7691 | NR | NR | Probability | Blood donors | 2.2% | NR \| NR \| NR | No data | - |
| Bolotin (Public health Ontario)[51] | 2020-06-05 to 2020-06-30 | Ontario | Regional | High | 7023 | NR | NR | Non-probability | Residual sera | 1.5% | USA; Abbott Laboratories \| Abbott architect SARSCoV-2 IgG test \| IgG | 0.4%, 1.0% (FINDDx / MUHC independent evaluation) | - |

| Article Author (Study Organization) | Sampling Dates (YMD) | Location | Geographic Scope | Overall Risk of Bias | Sample Size | Age | Female (%) | Sampling method | Sample frame | Uncorrected Seroprevalence (95% Confidence Interval) | Test Manufacturers \| Names \| Isotypes | Test Sens, Spec (Source) | Corrected Seroprevalence (95% Credible Interval) |
|---|---|---|---|---|---|---|---|---|---|---|---|---|---|
| Canadian Blood Services (Canadian Blood Services)[55] | 2020-05-09 to 2020-06-18 | | National | Moderate | 37737 | 17-25 (n=3581); 25-39 (n=10781); 40-59 (14147); 60+(n=9228) | 46.9 | Non-probability | Blood donors | 0.7% (0.6-0.8%) | Abbott \| Abbott Architect SARS-CoV-2 IgG assay \| IgG | 0.4%, 1.0% (FINDDx / MUHC independent evaluation) | - |
| Bolotin (Public health Ontario)[51] | 2020-05-05 to 2020-05-30 | Ontario | Regional | High | 1061 | NR | NR | Non-probability | Residual sera | 1.9% | USA; Abbott Laboratories \| Abbott architect SARSCoV-2 IgG test \| IgG | 0.4%, 1.0% (FINDDx / MUHC independent evaluation) | - |
| Skowronski (British Columbia Centre for Disease Control)[56] | 2020-05-15 to 2020-05-27 | British Columbia | Regional | Moderate | 885 | Median 45 (Range <10-80+) | 50.5 | Probability | Residual sera | 0.6% (0.1-1.4%) | Ortho-Clinical Diagnostics; Abbott; Siemens Healthineers \| VITROS anti-SARS-CoV-2 total antibody, Abbott Architect SARS-CoV-2 IgG assay \| IgG, IgA, IgM | 97.8%, 97.8% (Used Bastos SR/MA data; no sens, spec, or author adjustment available) | 0.1% (0.0-0.4%) |
| Bolotin (Public health Ontario)[51] | 2020-03-27 to 2020-04-30 | Ontario | Regional | High | 827 | NR | NR | Non-probability | Residual sera | 0.6% | USA; Abbott Laboratories \| Abbott architect SARSCoV-2 IgG test \| IgG | 0.4%, 1.0% (FINDDx / MUHC independent evaluation) | - |
| Skowronski (British Columbia Centre for Disease Control)[56] | 2020-03-05 to 2020-03-13 | British Columbia | Regional | Moderate | 869 | Median 45 (Range <10-80+) | 51.1 | Probability | Residual sera | 0.3% (0.0-0.9%) | Ortho-Clinical Diagnostics; Abbott \| VITROS anti-SARS-CoV-2 total antibody, Abbott Architect SARS-CoV-2 IgG assay \| IgG, IgA, IgM | 97.8%, 97.8% (Used Bastos SR/MA data; no sens, spec, or author adjustment available) | 0.1% (0.0-0.4%) |
| **Denmark** | | | | | | | | | | | | | |
| Jespersen (Aarhus | 2020-06-17 to 2020-06-30 | Eastern Central | Regional | High | 180 | NR | NR | Non-probability | Blood donors | 0.6% (0.0-3.5%) | Wantai Biological | 96.7%, 99.5% (Used author- | 0.6% (0.1-2.6%) |

| Article Author (Study Organization) | Sampling Dates (YMD) | Location | Geographic Scope | Overall Risk of Bias | Sample Size | Age | Female (%) | Sampling method | Sample frame | Uncorrected Seroprevalence (95% Confidence Interval) | Test Manufacturers \| Names \| Isotypes | Test Sens, Spec (Source) | Corrected Seroprevalence (95% Credible Interval) |
|---|---|---|---|---|---|---|---|---|---|---|---|---|---|
| University Hospital)[61] | | Denmark Region | | | | | | | | | Pharmacy Enterprise \| Wantai SARS CoV-2 Total Antibody ELISA \| IgG, IgA, IgM | adjusted estimate) | |
| Jespersen (Aarhus University Hospital)[61] | 2020-06-17 to 2020-06-30 | Western Central Denmark Region | Regional | High | 180 | NR | NR | Non-probability | Blood donors | 1.2% (0.0-4.4%) | Wantai Biological Pharmacy Enterprise \| Wantai SARS CoV-2 Total Antibody ELISA \| IgG, IgA, IgM | 96.7%, 99.5% (Used author-adjusted estimate) | 1.2% (0.2-3.5%) |
| Pedersen (Copenhagen University Hospital Rigshospitalet)[62] | 2020-06-01 to 2020-06-26 | | National | Moderate | 1110 | NR | NR | Non-probability | Blood donors | 2.5% (1.3-3.8%) | Beijing Wantai Biological Pharmacy Enterprise \| WANTAI SARS-CoV-2 Ab ELISA \| NR | 96.7%, 99.5% (Used author-adjusted estimate) | 2.5% (1.6-3.5%) |
| Pedersen (Copenhagen University Hospital Rigshospitalet)[62] | 2020-06-02 to 2020-06-19 | | National | Moderate | 1200 | Median 73 (IQR 71-76) | 43.0 | Probability | Blood donors | 1.4% (0.3-2.5%) | Beijing Wantai Biological Pharmacy Enterprise \| WANTAI SARS-CoV-2 Ab ELISA \| NR | 96.7%, 99.5% (Used author-adjusted estimate) | 1.4% (0.8-2.1%) |
| Erikstrup (Copenhagen University Hospital)[65] | 2020-04-06 to 2020-05-03 | | National | Moderate | 20640 | 17-69 (n=20640) | 50.5 | Probability | Blood donors | 1.9% (0.8-2.3%) | Livzon Diagnostics \| 2019-nCoV IgG/IgM Antibody Detection Kit \| IgG, IgM | 82.6%, 99.5% (Used author-adjusted estimate) | 1.9% (1.7-2.1%) |
| Iversen (University of Copenhagen)[66] | 2020-04-15 to 2020-04-22 | Capital Region of Denmark | Regional | High | 4672 | NR | NR | Non-probability | Blood donors | 3.0% (2.6-3.6%) | Livzon Diagnostics \| NR \| IgG, IgM | 73.4%, 99.9% (Used author-adjusted estimate) | 3.0% (2.6-3.6%) |
| **Faroe Islands** | | | | | | | | | | | | | |

| Article Author (Study Organization) | Sampling Dates (YMD) | Location | Geographic Scope | Overall Risk of Bias | Sample Size | Age | Female (%) | Sampling method | Sample frame | Uncorrected Seroprevalence (95% Confidence Interval) | Test Manufacturers \| Names \| Isotypes | Test Sens, Spec (Source) | Corrected Seroprevalence (95% Credible Interval) |
|---|---|---|---|---|---|---|---|---|---|---|---|---|---|
| Petersen (University of the Faroe Islands)[67] | 2020-04-27 to 2020-05-01 | Faroe Islands | Regional | Low | 1075 | Mean 42.1 (SD 23.1) | 50.0 | Probability | Household and community samples | 0.7% | Beijing Wantai Biological Pharmacy Enterprise \| Wantai SARS CoV-2 Total Antibody ELISA \| IgG, IgM | 94.4%, 100.0% (Used author-adjusted estimate) | 0.7% (0.3-1.3%) |
| **France** | | | | | | | | | | | | | |
| Carrat (Paris University)[75] | 2020-05-04 to 2020-06-23 | | National | Moderate | 14628 | <40 (n=2262), 40-50 (n=2897), 50-60 (n=3019), 60-70 (3272), 70+ (n=3175) | 60.3 | Probability | Household and community samples | 6.7% | Euroimmun \| ELISA-S Anti-Sars-CoV-2 IgG \| IgG | 0.6%, 0.9% (FINDDx / MUHC independent evaluation) | - |
| Capai (Université de Corse)[78] | 2020-04-16 to 2020-06-15 | Corsica \| Ajaccio Corte | Regional | Moderate | 1973 | Median 52 (IQR 34-70) | 60.2 | Non-probability | Residual sera | 5.5% (4.5-6.6%) | Bussy-Saint Martin; EUROIMMUN ; France \| EUROIMMUN ELISA IgG enzyme immunoassay kit \| IgG | 0.6%, 0.9% (FINDDx / MUHC independent evaluation) | - |
| Vu (Santé publique France)[81] | 2020-05-11 to 2020-05-17 | Auvergne-Rhône-Alpes, Occitanie, Martinique, Bourgogne-Franche Comté, Île-de-France, Corse, Hauts-de-France, Guadeloupe, French Guiana, Bretagne, Pays de la Loire, Centre-Val-de-Loire, | National | Moderate | 3592 | 0-9 (n=255); 10-19 (n=336); 20-29 (n=318); 30-39 (n=409); 40-49 (n=390); 50-59 (n=469); 60-69 (n=496); 70-79 (n=480); =>80 (n=439) | 57.0 | Non-probability | Residual sera | 13.0% | N/A - Author Designed \| Luciferase-Linked ImmunoSorbent Assay N, Luciferase-Linked ImmunoSorbent Assay S, Pseudo-neutralisation assay \| IgG | 100.0%, 100.0% (Test developer / manufacturer evaluation) | 12.9% (11.5-14.4%) |

| Article Author (Study Organization) | Sampling Dates (YMD) | Location | Geographic Scope | Overall Risk of Bias | Sample Size | Age | Female (%) | Sampling method | Sample frame | Uncorrected Seroprevalence (95% Confidence Interval) | Test Manufacturers \| Names \| Isotypes | Test Sens, Spec (Source) | Corrected Seroprevalence (95% Credible Interval) |
|---|---|---|---|---|---|---|---|---|---|---|---|---|---|
| | | Grand-Est, Nouvelle-Aquitaine, La Reunion, Provence-Alpes-Côte d'Azur, Normandie | | | | | | | | | | | |
| Cohen (Centre Hospitalier Intercommunal de Créteil)[82] | 2020-04-14 to 2020-05-12 | Île-de-France \| Paris | Local | High | 605 | Mean 4.9 (SD 3.9) | 46.8 | Non-probability | Residual sera | 10.7% (8.4-13.5%) | Biosynex \| Biosynex COVID-19 BSS test \| IgG, IgM | 91.8%, 99.2% (Test developer / manufacturer evaluation) | 9.9% (4.6-14.2%) |
| Fontanet (Institut Pasteur)[84] | 2020-04-28 to 2020-04-30 | Hauts-de-France \| Crepy-en-Valois | Local | Moderate | 1340 | Median NR (Pupils); 40 (parents); 47.5 (teachers); 47.5 (non-teaching staff) (IQR TR: 6-11 Pupils; 37-44 (Parents); 40-51 (Teachers); 32-54 (non-teaching staff)) | 57.4 | Non-probability | Household and community samples | 10.4% | Institut Pasteur \| S-Flow assay \| NR | 99.4%, 100.0% (Test developer / manufacturer evaluation) | 10.0% (7.9-12.1%) |
| Vu (Santé publique France)[81] | 2020-04-06 to 2020-04-12 | Auvergne-Rhône-Alpes, Occitanie, Martinique, Bourgogne-Franche Comté, Île-de-France, Corse, Hauts-de-France, Pays de la Loire, Nouvelle-Aquitaine, Bretagne, French Guiana, | National | Moderate | 3595 | 0-9 (n=167); 10-19 (n=298); 20-29 (n=355); 30-39 (n=461); 40-49 (n=455); 50-59 (n=489); 60-69 (n=487); 70-79 (n=455); =>80 (n=428) | 56.7 | Non-probability | Residual sera | 10.7% | N/A - Author Designed \| Luciferase-Linked ImmunoSorbent Assay N, Luciferase-Linked ImmunoSorbent Assay S, Pseudo-neutralisation assay \| IgG | 100.0%, 100.0% (Test developer / manufacturer evaluation) | 10.5% (9.1-11.9%) |

| Article Author (Study Organization) | Sampling Dates (YMD) | Location | Geographic Scope | Overall Risk of Bias | Sample Size | Age | Female (%) | Sampling method | Sample frame | Uncorrected Seroprevalence (95% Confidence Interval) | Test Manufacturers \| Names \| Isotypes | Test Sens, Spec (Source) | Corrected Seroprevalence (95% Credible Interval) |
|---|---|---|---|---|---|---|---|---|---|---|---|---|---|
| | | Centre-Val-de-Loire, Grand-Est, Guadeloupe, La Reunion, Provence-Alpes-Côte d'Azur, Normandie | | | | | | | | | | | |
| Gallian (Unite des Virus Emergents)[86] | 2020-03-25 to 2020-04-07 | Hauts-de-France, Provence-Alpes-Côte d'Azur, Île-de-France, Grand-Est | National | High | 998 | Median 41 (NR) | 49.0 | Non-probability | Blood donors | 2.7% | Author-designed \| N/A - Author-designed \| NR | No data | - |
| Grzelak (Institut Pasteur)[87] | 2020-03-20 to 2020-03-24 | Hauts-de-France \| Clermont, Lille, Noyon | Local | High | 200 | NR | NR | Non-probability | Blood donors | 0.0% | Author-designed \| N/A - Author-designed \| IgG, IgA, IgM | 75.6%, 97.6% (Used Bastos SR/MA data; no sens, spec, or author adjustment available) | 0.5% (0.0-2.0%) |
| Vu (Santé publique France)[81] | 2020-03-09 to 2020-03-15 | Auvergne-Rhône-Alpes, Occitanie, Martinique, Bourgogne-Franche Comté, Corse, Île-de-France, Hauts-de-France, Pays de la Loire, Nouvelle-Aquitaine, Bretagne, French Guiana, | National | Moderate | 3834 | 0-9 (n=359); 10-19 (n=420); 20-29 (n=420); 30-39 (n=500); 40-49 (n=487); 50-59 (n=461); 60-69 (n=455); 70-79 (n=382); =>80 (n=347) | 55.9 | Non-probability | Residual sera | 3.8% | N/A - Author Designed \| Luciferase-Linked ImmunoSorbent Assay N, Luciferase-Linked ImmunoSorbent Assay S, Pseudo-neutralisation assay \| IgG | 100.0%, 100.0% (Test developer / manufacturer evaluation) | 3.5% (2.3-4.4%) |

| Article Author (Study Organization) | Sampling Dates (YMD) | Location | Geographic Scope | Overall Risk of Bias | Sample Size | Age | Female (%) | Sampling method | Sample frame | Uncorrected Seroprevalence (95% Confidence Interval) | Test Manufacturers \| Names \| Isotypes | Test Sens, Spec (Source) | Corrected Seroprevalence (95% Credible Interval) |
|---|---|---|---|---|---|---|---|---|---|---|---|---|---|
| | | Grand-Est, Centre-Val-de-Loire, Guadeloupe, La Reunion, Provence-Alpes-Côte d'Azur, Normandie | | | | | | | | | | | |
| **Germany** | | | | | | | | | | | | | |
| Fuest (ifo Institute)[89] | 2020-10-26 to 2020-11-18 | | National | Unclear | 9929 | 18+ | NR | Non-probability | Household and community samples | 1.1% (0.9-1.3%) | Roche \| Elecsys \| NR | 0.9%, 1.0% (FINDDx / MUHC independent evaluation) | - |
| Robert Koch Institute (Robert Koch Institute)[90] | 2020-04-15 to 2020-11-05 | | National | Moderate | 48976 | NR | NR | Probability | Blood donors | 1.8% | NR \| NR \| IgG | 75.6%, 97.6% (Used Bastos SR/MA data; no sens, spec, or author adjustment available) | 0.2% (0.0-0.5%) |
| Schwarz (University of Witten)[93] | 2020-06-15 to 2020-09-01 | | National | High | 731 | NR | NR | Non-probability | Residual sera | 11.2% | Cleartest IgM; Euroimmun IgA; Diasorin IgG; Euroimmun IgG; Nadal IgM; Microgen IgG; Roche IgM \| Multiple \| IgG, IgA, IgM | No data | - |
| Laub (University of Regensburg)[97] | 2020-05-22 to 2020-07-22 | Bavaria | Regional | Moderate | 2832 | NR | 42.7 | Non-probability | Household and community samples | 4.9% | Roche Diagnostics \| Elecsys Anti-SARS-CoV-2 \| IgG, IgA, IgM | 0.6%, 1.0% (FINDDx / MUHC independent evaluation) | - |
| Aziz (University of Bonn)[101] | 2020-04-24 to 2020-06-30 | North Rhine- | Local | High | 4771 | 30-100 | 56.9 | Non-probability | Household and | 1.0% (0.7-1.3%) | EUROIMMUN \| Euroimmun Anti-SARS- | 0.6%, 0.9% (FINDDx / MUHC | - |

| Article Author (Study Organization) | Sampling Dates (YMD) | Location | Geographic Scope | Overall Risk of Bias | Sample Size | Age | Female (%) | Sampling method | Sample frame | Uncorrected Seroprevalence (95% Confidence Interval) | Test Manufacturers \| Names \| Isotypes | Test Sens, Spec (Source) | Corrected Seroprevalence (95% Credible Interval) |
|---|---|---|---|---|---|---|---|---|---|---|---|---|---|
| | | Westphalia \| Bonn | | | | | | | community samples | | CoV-2 ELISA IgG \| IgG | independent evaluation) | |
| Aziz (University of Bonn)[101] | 2020-04-24 to 2020-06-30 | North Rhine-Westphalia \| Bonn | Local | High | 360 | 30-100 | 60.6 | Non-probability | Household and community samples | 1.9% (0.8-4.4%) | EUROIMMUN \| Euroimmun Anti-SARS-CoV-2 ELISA IgG \| IgG | 0.6%, 0.9% (FINDDx / MUHC independent evaluation) | - |
| Runkel (University Medical Center of The Johannes Gutenberg-University Mainz)[104] | 2020-03-15 to 2020-06-15 | Southwest | Regional | High | 3880 | 18-29 (n=1292); 30-39 (n=772); 40-49 (n=537); 50-59 (n=797); 60-71 (n=282) | 45.3 | Non-probability | Blood donors | 0.9% | Abbott Laboratories \| ARCHITECT Anti-SARS-CoV-2 IgG Assay \| IgG | 0.4%, 1.0% (FINDDx / MUHC independent evaluation) | - |
| Santos-Hovener (Robert Koch Institute)[105] | 2020-05-20 to 2020-06-09 | Baden-Württemberg \| Kupferzell | Local | Low | 2203 | 18-34 (n= 713); 35-49 (n=533); 50-64 (n=572); =>65 (n= 385) | 51.9 | Probability | Household and community samples | 12.0% (10.4-14.0%) | Euroimmun \| SARS-CoV-2-S1 ELISA \| IgG | 88.3%, 99.2% (Used author-adjusted estimate) | 12.0% (10.7-13.4%) |
| Herrmann (Internal medicine and laboratory medicine specialist practice and laboratory)[107] | 2020-03-26 to 2020-06-04 | North Rhine-Westphalia | Regional | High | 415 | Mean 42 (SD 27) | NR | Non-probability | Residual sera | 1.2% | Euroimmun \| anti-SARS-CoV-2-ELISA-IgG \| IgG | 0.6%, 0.9% (FINDDx / MUHC independent evaluation) | - |
| Fischer (Heart and Diabetes Center NRW)[108] | 2020-03-09 to 2020-06-03 | Lower Saxony, North Rhine-Westphalia, Hesse | Regional | High | 3186 | NR | 70.8 | Non-probability | Residual sera | 0.9% (0.6-1.2%) | DiaSorin; EUROIMMUN; Abbott \| Abbott Architect SARS-CoV-2 IgG assay, Euroimmun Anti-SARS-CoV-2 ELISA IgG, LIAISON SARS-CoV-2 S1/S2 IgG \| IgG | 75.6%, 97.6% (Used Bastos SR/MA data; no sens, spec, or author adjustment available) | 0.1% (0.0-0.3%) |
| Weis (Jena University Hospital)[111] | 2020-05-12 to 2020-05-22 | Thuringia | Local | Moderate | 620 | Mean 58.1 (adults); 9.6 (children) (SD 16.5 (adults); | 51.3 | Probability | Household and community samples | 8.4% | Snibe Diagnostic; EUROIMMUN; Roche; | No data | - |

| Article Author (Study Organization) | Sampling Dates (YMD) | Location | Geographic Scope | Overall Risk of Bias | Sample Size | Age | Female (%) | Sampling method | Sample frame | Uncorrected Seroprevalence (95% Confidence Interval) | Test Manufacturers \| Names \| Isotypes | Test Sens, Spec (Source) | Corrected Seroprevalence (95% Credible Interval) |
|---|---|---|---|---|---|---|---|---|---|---|---|---|---|
| | | | | | | 4.38 (children) ) | | | | | DiaSorin; Epitope Diagnostics; Abbott \| EDI Novel Coronavirus SARS-CoV-2 IgG ELISA kit, SARS-CoV-2 IgG ELISA kit, SARS-CoV-2 S1/S2 IgG CLIA kit, 2019-nCoV IgG kit, SARS-CoV-2 IgG CMIA kit, Elecsys Anti-SARS-CoV-2 kit \| IgG | | |
| Weis (Friedrich Schiller University)[113] | 2020-05-13 to 2020-05-16 | Thuringia | Sublocal | Moderate | 620 | Mean 58.1 (adults); 9.62 (children and adolescents) (SD 16.5 (adults); 4.38 (children and adolescents) ) | 51.3 | Non-probability | Household and community samples | 8.4% | USA Euroimmun; Lübeck; USA Roche; Italy Snibe Co.; Ltd.; Basel Switzerland; China Abbott; Shenzhen; Saluggia; Germany DiaSorin; Epitope Diagnostics Inc.; San Diego; Chicago \| EDI Novel Coronavirus SARS-CoV-2 IgG ELISA kit, SARS-CoV-2 IgG ELISA kit, SARS-CoV-2 S1/S2 IgG CLIA kit, 2019-nCoV IgG kit, SARS-CoV-2 IgG CMIA kit and Elecsys Anti-SARS-CoV-2 kit. \| IgG | No data | - |

| Article Author (Study Organization) | Sampling Dates (YMD) | Location | Geographic Scope | Overall Risk of Bias | Sample Size | Age | Female (%) | Sampling method | Sample frame | Uncorrected Seroprevalence (95% Confidence Interval) | Test Manufacturers \| Names \| Isotypes | Test Sens, Spec (Source) | Corrected Seroprevalence (95% Credible Interval) |
|---|---|---|---|---|---|---|---|---|---|---|---|---|---|
| Reisinger (Universitätsmedizin Rostock)[118] | 2020-04-20 to 2020-04-22 | Mecklenburg-Westpomerania \| Rostock | Sublocal | High | 401 | Mean 36.9 (NR) | 100.0 | Non-probability | Household and community samples | 3.0% | EUROIMMUN \| Euroimmun Anti-SARS-CoV-2 IgG IgA ELISA \| IgG, IgA | 0.9%, 0.8% (FINDDx / MUHC independent evaluation) | - |
| Streeck (University of Bonn)[125] | 2021-03-31 to 2020-04-06 | Heinsberg \| Gangelt | Local | Low | 919 | Median 53 (Range 1-90) | 50.8 | Probability | Household and community samples | 14.1% (11.2-17.3%) | EUROIMMUN \| Euroimmun Anti-SARS-CoV-2 ELISA IgG \| IgG | 90.9%, 99.1% (Used author-adjusted estimate) | 14.1% (11.9-16.4%) |
| **Greece** | | | | | | | | | | | | | |
| Bogogiannidou (University of Thessaly)[129] | 2020-03-01 to 2020-04-30 | Epirus, Western Macedonia, Thessaly, Central Macedonia, Attica, Peloponnese, Western Greece, Central Greece | National | Moderate | 6586 | 0-29 (n=1464); 30-49 (n=2066); 50-69 (n=1762); >=70 (n=1294) | 55.0 | Non-probability | Residual sera | 0.4% | Abbott \| Abbott Architect SARS-CoV-2 IgG assay \| IgG | 0.4%, 1.0% (FINDDx / MUHC independent evaluation) | - |
| **Iceland** | | | | | | | | | | | | | |
| Gudbjartsson (deCODE Genetics)[130] | 2020-04-27 to 2020-06-12 | | National | Moderate | 23452 | Mean 56 (SD 20) | NR | Non-probability | Household and community samples | 0.3% | Wantai/Nordic BioSite; Roche International \| Roche Elecsys chemiluminescence assay, Wantai SARS CoV-2 Total Antibody ELISA \| IgG, IgA, IgM | 91.1%, 99.8% (Test developer / manufacturer evaluation) | 0.1% (0.0-0.3%) |
| **Ireland** | | | | | | | | | | | | | |
| Igoe (HSE Health Protection Surveillance | 2020-06-22 to 2020-07-16 | Leinster, Connaught \| Dublin, Sligo | National | Moderate | 1733 | 12-19 (n=3); 20-29 (n=7); 30-39 (n=5); 40-49 (n=7); | NR | Probability | Household and community samples | 1.7% (1.1-2.4%) | FORTRESS; Abbott \| Abbott Architect SARS-CoV-2 IgG assay, | 97.8%, 97.8% (Used Bastos SR/MA data; no sens, spec, or author | 0.2% (0.0-0.6%) |

| Article Author (Study Organization) | Sampling Dates (YMD) | Location | Geographic Scope | Overall Risk of Bias | Sample Size | Age | Female (%) | Sampling method | Sample frame | Uncorrected Seroprevalence (95% Confidence Interval) | Test Manufacturers \| Names \| Isotypes | Test Sens, Spec (Source) | Corrected Seroprevalence (95% Credible Interval) |
|---|---|---|---|---|---|---|---|---|---|---|---|---|---|
| Centre (HPSC) )[131] | | | | | | 50-59 (n=5); 60-69 (n=6) | | | | | FORTRESS (Wantai)-2 Assay \| IgG | adjustment available) | |
| **Israel** | | | | | | | | | | | | | |
| Rabinowitz (The Central Bureau of Statistics )[132] | 2020-06-15 to 2020-08-15 | Bnei Brak | Local | Moderate | 3500 | NR | NR | Probability | Household and community samples | 14.6% | NR \| NR \| NR | No data | - |
| Efrati (Israeli Pandemic Task Force)[133] | Until 2020-06-02 | Israel | National | Moderate | 1700 | NR | NR | Probability | Household and community samples | 2.5% | NR \| NR \| IgG | No data | - |
| **Italy** | | | | | | | | | | | | | |
| Italian Ministry of Health (Italian Ministry of Health)[137] | 2020-05-25 to 2020-07-15 | Molise, Campania, Friuli-Venezia Giulia, Abruzzo, Lazio, Liguria, Basilicata, Lombardy, Piemonte, Umbria, Sicily, Calabria, Sardegna, Valle D'Aosta, Veneto, Emilia Romagna, Puglia, Trentino-Alto Adige, Marche, Tuscany | National | Unclear | 64660 | NR | NR | Non-probability | Household and community samples | 2.5% | Abbott \| Abbott Architect SARS-CoV-2 IgG assay \| IgG | 0.4%, 1.0% (FINDDx / MUHC independent evaluation) | - |
| Cento (Universita degli Studi di Milano)[139] | 2020-05-11 to 2020-07-05 | Lombardy \| Milan | Sublocal | Moderate | 2753 | =<10 (n=8); 11–24 (n=132); 25–49 years (n=786); 50-64 (n=545); 65–84 | 49.5 | Non-probability | Residual sera | 3.3% | Abbott Park; Abbott; Saluggia; Italy; Illinois; US; DiaSorin | 97.8%, 97.8% (Used Bastos SR/MA data; no sens, spec, or author | 1.2% (0.2-2.1%) |

| Article Author (Study Organization) | Sampling Dates (YMD) | Location | Geographic Scope | Overall Risk of Bias | Sample Size | Age | Female (%) | Sampling method | Sample frame | Uncorrected Seroprevalence (95% Confidence Interval) | Test Manufacturers \| Names \| Isotypes | Test Sens, Spec (Source) | Corrected Seroprevalence (95% Credible Interval) |
|---|---|---|---|---|---|---|---|---|---|---|---|---|---|
| | | | | | | (n=984); =>85 (n=316) | | | | | S.p.A. \| Abbott ARCHITECT SARS-CoV-2 IgG assay; LIAISON1 SARS-CoV-2 S1/S2 IgG assay \| IgG | adjustment available) | |
| Berte (University of Cagliari)[141] | 2020-04-15 to 2020-06-15 | Lombardy \| Milan | Local | High | 129 | Median 45 (IQR 35-60) | 34.9 | Non-probability | Household and community samples | 7.0% (4.4-9.6%) | University of Cagliari \| N/A - Author Designed \| IgG, IgA | 97.6%, 95.2% (Test developer / manufacturer evaluation) | 3.3% (0.0-8.5%) |
| Pagani (University of Milan)[143] | 2020-05-18 to 2020-06-07 | Lombardy \| Castiglion e D'Adda | Local | Moderate | 509 | Median 46 (IgG negative); 55.4 (IgG positive) (SD 20.6 (IgG negative); 19.5 (IgG positive)) | 48.9 | Probability | Household and community samples | 22.6% (17.2-29.1%) | Abbott \| IgG Anti-SARS-CoV-2 CLIA \| IgG | 0.4%, 1.0% (FINDDx / MUHC independent evaluation) | - |
| Amante (Bergamo Health Agency)[145] | 2020-04-23 to 2020-06-03 | Lombardy | Regional | Moderate | 9965 | NR | NR | Probability | Household and community samples | 57.0% | NR \| NR \| NR | No data | - |
| Fiore (University of Foggia)[149] | 2020-05-01 to 2020-05-31 | Apulia \| Foggia | Local | High | 904 | 18 to 25 (n=112, 12.4%), 26 to 35 (n=195, 21.6%), 36 to 45 (n=202, 22.3%), 46 to 55 (n=149, 27.2%), 56 to 65 (n=149, 16.5%) | 26.4 | Non-probability | Blood donors | 1.0% | "New Industries Biomedical Engineering Co.; Ltd" \| NR \| IgG, IgM | 97.8%, 97.8% (Used Bastos SR/MA data; no sens, spec, or author adjustment available) | 0.1% (0.0-0.5%) |
| Alessi (ASL Vercelli)[154] | 2020-05-23 to 2020-05-24 | Piedmont \| Borgosesi a | Local | High | 4987 | Mean 55 years (Range 18-97) | 54.2 | Non-probability | Household and community samples | 4.9% (4.3-5.5%) | Prima Lab SA \| COVID-19 IgG/IgM Rapid Test Cassette \| IgG, IgM | 0.8%, 0.9% (FINDDx / MUHC independent evaluation) | - |
| Stefanelli (Azienda Provinciale per i Servizi Sanitari)[161] | 2020-05-05 to 2020-05-15 | Trento | Regional | Low | 6075 | Median 50 (Range 10-98) | 51.3 | Probability | Household and community samples | 25.7% | Abbott Laboratories \| Abbott SARS-CoV-2 IgG assay \| IgG | 90.0%, 100.0% (Used author-adjusted estimate) | 25.7% (24.6-26.8%) |

| Article Author (Study Organization) | Sampling Dates (YMD) | Location | Geographic Scope | Overall Risk of Bias | Sample Size | Age | Female (%) | Sampling method | Sample frame | Uncorrected Seroprevalence (95% Confidence Interval) | Test Manufacturers \| Names \| Isotypes | Test Sens, Spec (Source) | Corrected Seroprevalence (95% Credible Interval) |
|---|---|---|---|---|---|---|---|---|---|---|---|---|---|
| Guerriero (Instituto di Ricovero e Cura A Caratterre)[163] | 2020-04-25 to 2020-05-08 | Veneto \| Verona | Local | Moderate | 1515 | Mean 52.1 (SD 20) | 53.9 | Probability | Household and community samples | 2.7% | Abbott Laboratories \| Abbott test \| IgG | 0.4%, 1.0% (FINDDx / MUHC independent evaluation) | - |
| Baracco (A.S.S.T.Lodi)[164] | 2020-04-23 to 2020-05-05 | Lombardy region \| Lodi | Local | Moderate | 1792 | Mean 44 (SD 16.0) | 64.7 | Probability | Household and community samples | 29.8% | DiaSorin \| The Liaison® DiaSorin® SARS-CoV-2 S1/S2 IgG test \| IgG | 0.8%, 1.0% (FINDDx / MUHC independent evaluation) | - |
| Vena (University of Genoa)[167] | 2020-03-01 to 2020-04-30 | Lombardy , Liguria | Regional | High | 3609 | Median 51 (IQR 41-63) | 55.6 | Non-probability | Residual sera | 11.0% (10.0-12.1%) | Biosynex; Realy-Tech; Maglumi \| MAGLUMI800TM 2019-nCoV IgG (Cat. Ref. 130219015M) and IgM (130219016M), Realy-Tech 2019 nCOV/COVID-19 IgG/IgM Rapid Test, Biosynex COVID-19 BSS test \| IgG, IgM | No data | - |
| Landro (ATS Bergamo)[170] | 2020-04-23 to 2020-04-28 | Lombardy \| Nembro, Alzano | Local | Unclear | 750 | NR | NR | Non-probability | Household and community samples | 61.0% | NR \| NR \| NR | No data | - |
| Loconsole (Policlinico Hospital)[173] | 2020-03-23 to 2020-04-21 | Apulia \| Bari | Local | High | 819 | Median 66 (IQR 52-80) | 44.6 | Non-probability | Residual sera | 8.6% | VivaChek Biotech \| VivaDiag COVID-19 IgM/IgG Rapid Test \| IgG, IgM | 0.8%, 0.7% (FINDDx / MUHC independent evaluation) | - |
| Cito (Istituto Zooprofilattic o Sperimentale dell'Abruzzo e del Molise G. Caporale )[174] | 2020-04-18 to 2020-04-19 | Abruzzo \| Villa Caldari | Local | Moderate | 667 | Mean 58.6 (seropositive); 47.3 (seronegative) (IQR 28.5 (seropositive); | 50.2 | Probability | Household and community samples | 10.9% (8.8-13.5%) | China; Beijing; Beijing Wantai Biological Pharmacy Enterprise Co. \| WANTAI SARS-CoV-2 | 1.0%, 1.0% (FINDDx / MUHC independent evaluation) | - |

| Article Author (Study Organization) | Sampling Dates (YMD) | Location | Geographic Scope | Overall Risk of Bias | Sample Size | Age | Female (%) | Sampling method | Sample frame | Uncorrected Seroprevalence (95% Confidence Interval) | Test Manufacturers \| Names \| Isotypes | Test Sens, Spec (Source) | Corrected Seroprevalence (95% Credible Interval) |
|---|---|---|---|---|---|---|---|---|---|---|---|---|---|
| | | | | | | 28.1 (seronegative)) | | | | | Total Ab ELISA \| NR | | |
| Valenti ( Fondazione IRCCS Ca' Granda Ospedale Maggiore )[179] | 2020-02-24 to 2020-04-08 | Lombardy \| Milan | Local | High | 789 | Mean 42.6 (SARS-CoV-2 antibodies present); 40.7 (SARS-CoV-2 antibodies absent); (SD 13.4 (SARS-CoV-2 antibodies present); 13.2 (SARS-CoV-2 antibodies absent); ) | 35.0 | Non-probability | Blood donors | 5.1% | Prima Lab \| PRIMA COVID-19 IgG/IgM Rapid Test \| IgG, IgM | 0.8%, 0.9% (FINDDx / MUHC independent evaluation) | - |
| Percivalle (IRCCS Policlinico San Matteo)[180] | 2020-03-18 to 2020-04-06 | Lombardy \| Lodi | Local | High | 390 | Median 46 (Range 19-70) | 30.3 | Non-probability | Blood donors | 23.3% | Author-designed \| N/A - Author-designed \| NR | 95.0%, 100.0% (Test developer / manufacturer evaluation) | 22.7% (15.0-30.6%) |
| Milani (University of Milan)[182] | 2020-03-30 to 2020-03-31 | Lombardy \| Milan | Local | High | 197 | NR | NR | Non-probability | Household and community samples | 10.2% | EUROIMMUN ; Beijing Wantai Biological Pharmacy Enterprise \| Wantai SARS-CoV-2 Ab ELISA, Euroimmun Anti-SARS-CoV-2 ELISA IgG, Wantai Anti-SARS-CoV-2 IgM ELISA \| IgG, IgA, IgM | 75.6%, 97.6% (Used Bastos SR/MA data; no sens, spec, or author adjustment available) | 10.1% (4.8-16.0%) |
| Pancrazzi (Ospedale San Donato)[183] | 2020-03-17 to 2020-03-21 | Tuscany \| Arezzo | Local | High | 516 | Mean 53.7 years (NR) | 54.5 | Non-probability | Residual sera | 13.0% | Acro Biotech Inc. \| Acro Biotech COVID-19 Rapid Test \| IgG, IgM | 0.4%, 0.9% (FINDDx / MUHC independent evaluation) | - |
| **Japan** | | | | | | | | | | | | | |

| Article Author (Study Organization) | Sampling Dates (YMD) | Location | Geographic Scope | Overall Risk of Bias | Sample Size | Age | Female (%) | Sampling method | Sample frame | Uncorrected Seroprevalence (95% Confidence Interval) | Test Manufacturers \| Names \| Isotypes | Test Sens, Spec (Source) | Corrected Seroprevalence (95% Credible Interval) |
|---|---|---|---|---|---|---|---|---|---|---|---|---|---|
| Nawa (Tokyo Medical and Dental University)[190] | 2020-06-14 to 2020-07-05 | Kantō \| Utsunomiya City | Local | Moderate | 742 | <10 (n=56) ; 10-17 (n=42) ; 18-65 (n=463) ; >65 (n=181) | 52.6 | Probability | Household and community samples | 1.2% (0.2-1.2%) | Shenzhen YHLO Biotech Co \| NR \| IgG | 0.7%, 1.0% (FINDDx / MUHC independent evaluation) | - |
| Okuda (Health and Welfare Department)[191] | 2020-06-15 to 2020-06-28 | prefecture 3 | Regional | Unclear | 3000 | NR | NR | Non-probability | Residual sera | 0.0% | NR \| NR \| IgG | No data | - |
| Takita (Navitas Clinic Tachikawa)[197] | 2020-04-20 to 2020-05-20 | Kantō, Chūbu \| Yamanashi, Chiba, Kangawa, Saitama, Tokyo | Local | High | 1071 | <=17 (n=13); 18-34 (n=134); 35-54 (n=653); =>55 (n=271) | 46.2 | Non-probability | Household and community samples | 3.8% (2.8-5.2%) | Kurabo Industries \| SARS-CoV-2 Antibody Testing Kit IgG (RC - NC002) \| IgG | 76.4%, 100.0% (Test developer / manufacturer evaluation) | 4.3% (0.2-6.9%) |
| Doi (Kobe City Medical Center General Hospital)[200] | 2020-03-31 to 2020-04-07 | Hyogo Prefecture \| Kobe | Local | High | 1000 | 0-10 (n=81); 0-19 (n=27); 20-29 (n=36); 30-39 (n=90); 40-49 (n=155); 50-59 (n=164); 60-69 (n=171); 70-79 (n=166); 80-89 (n=164); >=90 (n=19) | 51.1 | Probability | Residual sera | 2.7% (1.8-3.9%) | Kurabo Industries \| SARS-CoV-2 Antibody Testing Kit IgG (RC - NC002) \| IgG | 76.4%, 100.0% (Test developer / manufacturer evaluation) | 2.8% (0.0-4.8%) |
| **Luxembourg** | | | | | | | | | | | | | |
| Snoeck (Luxembourg Institute of Health)[203] | 2020-04-15 to 2020-05-05 | | National | Moderate | 1820 | Mean 47 (SD 15) | 52.1 | Probability | Household and community samples | 2.1% (1.4-2.8%) | EUROIMMUN \| Euroimmun Anti-SARS-CoV-2 ELISA IgG \| IgG | 0.8%, 1.0% (FINDDx / MUHC independent evaluation) | - |
| **Monaco** | | | | | | | | | | | | | |
| Binacchi (Principality of Monaco)[204] | 2020-05-26 to 2020-06-16 | | National | Unclear | 35000 | NR | NR | Non-probability | Household and community samples | 2.8% | NR \| NR \| NR | 66.0%, 96.6% (Used Bastos SR/MA data; no sens, spec, or author adjustment available) | 0.1% (0.0-0.5%) |
| **Netherlands** | | | | | | | | | | | | | |

| Article Author (Study Organization) | Sampling Dates (YMD) | Location | Geographic Scope | Overall Risk of Bias | Sample Size | Age | Female (%) | Sampling method | Sample frame | Uncorrected Seroprevalence (95% Confidence Interval) | Test Manufacturers \| Names \| Isotypes | Test Sens, Spec (Source) | Corrected Seroprevalence (95% Credible Interval) |
|---|---|---|---|---|---|---|---|---|---|---|---|---|---|
| Vos (Centre for Infectious Disease Control)[205] | 2020-03-31 to 2020-05-11 | | National | Low | 570 | 2-17 (n = 93) ; 18-39 (n =196) ; 40-64 ( n=198) ; 65-90 (n = 83) | 40.9 | Probability | Household and community samples | 2.9% (1.4-6.3%) | NR \| fluorescent bead-based multiplex-immunoassay \| IgG | 84.4%, 99.0% (Used author-adjusted estimate) | 2.9% (1.7-4.4%) |
| Vos (Centre for Infectious Disease Control)[205] | 2020-03-31 to 2020-05-11 | | National | Low | 2637 | 2-17 (n = 507) ; 18-39 (n = 735) ; 40-64 ( n=919) ; 65-90 (n = 476) | 55.1 | Probability | Household and community samples | 2.8% (2.1-3.7%) | NR \| fluorescent bead-based multiplex-immunoassay \| IgG | 84.4%, 99.0% (Used author-adjusted estimate) | 2.8% (2.2-3.4%) |
| Slot (Amsterdam UMC)[206] | 2020-04-01 to 2020-04-15 | | National | Moderate | 7361 | 18-30 (n=1251); 31-40 (n=882); 41-50 (n=1354); 51-60 (n=2132); 60-72 (n=1742) | 47.8 | Non-probability | Blood donors | 3.4% | Wantai Biological Pharmacy Enterprise \| Wantai SARS CoV-2 Total Antibody ELISA \| IgG, IgA, IgM | 0.7%, 1.0% (FINDDx / MUHC independent evaluation) | - |
| Westerhuis (Erasmus Medical Centre)[207] | 2020-04-03 to 2020-04-07 | Rotterdam | Sublocal | High | 729 | 6-12 months (n=17); 1-2 (n=14); 2-5 (n=24); 5-10 (n=34); 10-20 (n=139); 20-40 (n=140); 40-60 (n=140); 60-80 (n=139); 80+ (n=82) | NR | Non-probability | Residual sera | 2.3% | Author-designed \| N/A - Author-designed \| IgG | No data | - |
| Westerhuis (Erasmus Medical Centre)[207] | 2020-03-02 to 2020-03-06 | Rotterdam | Sublocal | High | 879 | 6-12 months (n=41); 1-2 (n=75); 2-5 (n=42); 5-10 (n=96); 10-20 (n=131); 20-40 (n=131); 40-60 (n=134); 60-80 (n=124); 80+ (n=105) | NR | Non-probability | Residual sera | 0.1% | Author-designed \| N/A - Author-designed \| IgG | No data | - |
| **Norway** | | | | | | | | | | | | | |
| Tunheim (Norwegian Institute of | 2020-04-20 to 2020-05-17 | | National | Moderate | 900 | 0-4 (n=41); 5-14 (n=115);15-24 (n=166); 25- | 56.6 | Non-probability | Residual sera | 1.1% (0.5-2.0%) | Oslo University Hospital"; " Department of | 86.0%, 100.0% (Test developer | 0.8% (0.0-1.8%) |

| Article Author (Study Organization) | Sampling Dates (YMD) | Location | Geographic Scope | Overall Risk of Bias | Sample Size | Age | Female (%) | Sampling method | Sample frame | Uncorrected Seroprevalence (95% Confidence Interval) | Test Manufacturers \| Names \| Isotypes | Test Sens, Spec (Source) | Corrected Seroprevalence (95% Credible Interval) |
|---|---|---|---|---|---|---|---|---|---|---|---|---|---|
| Public Health)[209] | | | | | | 59 (n=372); ≥60 (n=206) | | | | | Immunology \| N/A - Author-designed \| IgG | / manufacturer evaluation) | |
| **Portugal** | | | | | | | | | | | | | |
| Kislaya ( Instituto Nacional de Saúde Doutor Ricardo Jorge)[215] | 2020-05-21 to 2020-07-08 | Alentejo Region, Centro Region, Algarve Region, Madeira, Lisboa Region, Norte Region, Azores | National | Moderate | 2301 | 1-9 (n = 404) 10-19 (n = 377) 20-39 (n = 377) 40-59 (n = 479) >= 60 ( n = 664) | 54.1 | Non-probability | Residual sera | 2.9% (2.0-4.2%) | 1) Wantai 2) EUROIMMUN \| 1) Wantai SARS-CoV-2 enzyme- linked immunosorbent assay (ELISA), 2) EUROIMMUN anti-SARS-CoV-2 assay \| IgG, IgM | 96.0%, 98.7% (Test developer / manufacturer evaluation) | 1.1% (0.0-2.6%) |
| **Republic of Korea** | | | | | | | | | | | | | |
| KBS World (Korea Disease Control and Prevention Agency)[217] | 2020-08-15 to 2020-10-15 | | National | Unclear | 1379 | NR | NR | Non-probability | Household and community samples | 0.2% | NR \| NR \| NR | No data | - |
| Ji-Ho (Korea Centers for Disease Control and Prevention)[218] | Until 2020-07-09 | Korea | National | Unclear | 3055 | NR | NR | Non-probability | Household and community samples | 0.0% | NR \| NR \| NR | No data | - |
| Noh (Korea University)[220] | 2020-05-25 to 2020-05-29 | Seoul | Local | Moderate | 1500 | 0-19 (n=226); 20-29 (n=258); 30-39 (n=261); 40-49 (n=254); 50-59 (n=255); >60 (n=246) | 51.3 | Non-probability | Residual sera | 0.1% | Roche elecsys \| Elecsys Anti-SARS-CoV-2 kit \| NR | 0.9%, 1.0% (FINDDx / MUHC independent evaluation) | - |
| **Singapore** | | | | | | | | | | | | | |
| Kurohi (Singapore National Centre for Infectious Diseases)[222] | 2020-02-15 to 2020-04-15 | Singapore | Local | Unclear | 774 | NR | NR | Non-probability | Household and community samples | 0.0% | NR \| NR \| NR | No data | - |

| Article Author (Study Organization) | Sampling Dates (YMD) | Location | Geographic Scope | Overall Risk of Bias | Sample Size | Age | Female (%) | Sampling method | Sample frame | Uncorrected Seroprevalence (95% Confidence Interval) | Test Manufacturers \| Names \| Isotypes | Test Sens, Spec (Source) | Corrected Seroprevalence (95% Credible Interval) |
|---|---|---|---|---|---|---|---|---|---|---|---|---|---|
| **Spain** | | | | | | | | | | | | | |
| Flores (Municipality of Torrejon de Ardoz)[230] | 2020-05-29 to 2020-06-03 | Torrejon de Ardoz | Local | Moderate | 104299 | >=1 | NR | Probability | Household and community samples | 23.1% | NR \| NR \| IgG, IgM | No data | - |
| Pollán (Institute of Health Carlos III)[236] | 2020-04-27 to 2020-05-11 | | National | Low | 51958 | 0–19 (n=11422); 20–34 (n=8469); 35–49 (n=14532); 50–64 (n=15094); >=65 (n=11558) | 61.1 | Probability | Household and community samples | 4.6% (4.3-5.0%) | Abbott \| Abbott Architect SARS-CoV-2 IgG assay \| IgG | 0.4%, 1.0% (FINDDx / MUHC independent evaluation) | - |
| Montenegro ( Sardenya Primary Health Care Center)[243] | 2020-04-21 to 2020-04-24 | Catalonia \| Barcelona | Local | High | 311 | Mean 43.7 (SD 21.79) | 55.0 | Probability | Household and community samples | 5.5% (3.4-8.6%) | Lysine and Sure Screen; Livzlon \| Livzlon, Lysine, Sure Screen \| IgG, IgM | 66.0%, 96.6% (Used Bastos SR/MA data; no sens, spec, or author adjustment available) | 3.6% (0.0-7.5%) |
| **Sweden** | | | | | | | | | | | | | |
| The Local (Werlabs)[250] | 2020-07-01 to 2020-07-22 | Uppland, Skåne, Västra Götaland \| Gothenburg, Stockholm, Malmo | Local | High | 83000 | NR | NR | Non-probability | Household and community samples | 14.4% | Werlabs \| NR \| NR | No data | - |
| Region (Sweden Public Health Authority)[251] | 2020-06-15 to 2020-07-15 | Uppland \| Stockholm | Local | High | 149616 | NR | NR | Non-probability | Household and community samples | 17.6% | NR \| NR \| NR | No data | - |
| Lundkvist (Uppsala University )[253] | 2020-06-17 to 2020-06-18 | Uppland, Södermanland \| Stockholm | Sublocal | High | 123 | Mean 37 (NR) | 57.7 | Probability | Household and community samples | 4.1% (0.6-7.6%) | Zhejiang Orient Gene Biotech Co Ltd \| COVID-19 IgG / IgM Rapid Test Cassette \| IgG, IgM | 0.4%, 1.0% (FINDDx / MUHC independent evaluation) | - |

| Article Author (Study Organization) | Sampling Dates (YMD) | Location | Geographic Scope | Overall Risk of Bias | Sample Size | Age | Female (%) | Sampling method | Sample frame | Uncorrected Seroprevalence (95% Confidence Interval) | Test Manufacturers \| Names \| Isotypes | Test Sens, Spec (Source) | Corrected Seroprevalence (95% Credible Interval) |
|---|---|---|---|---|---|---|---|---|---|---|---|---|---|
| Lundkvist (Uppsala University )[253] | 2020-06-17 to 2020-06-18 | Uppland, Södermanland \| Stockholm | Sublocal | High | 90 | Mean 50 (NR) | 28.9 | Probability | Household and community samples | 30.0% (20.3-39.7%) | Zhejiang Orient Gene Biotech Co Ltd \| COVID-19 IgG / IgM Rapid Test Cassette \| IgG, IgM | 0.4%, 1.0% (FINDDx / MUHC independent evaluation) | - |
| Swedish Public Health Agency (Sweden Public Health Authority)[255] | 2020-06-08 to 2020-06-14 | Örebro, Västerbotten Västra Götaland, Skåne, Jönköping, Kalmar, Uppsala, Stockholm, Jämtland-Härjedalen | National | High | 1200 | NR | NR | Non-probability | Residual sera | 5.2% (3.7-7.1%) | Sci LifeLab / KTH \| NR \| NR | 99.4%, 0.0% (Used author-adjusted estimate) | 5.2% (4.0-6.5%) |
| Swedish Public Health Agency (Sweden Public Health Authority)[255] | 2020-06-01 to 2020-06-07 | Örebro, Västerbotten Västra Götaland, Skåne, Jönköping, Kalmar, Uppsala, Stockholm, Jämtland-Härjedalen | National | High | 1200 | NR | NR | Non-probability | Residual sera | 6.8% (5.1-8.8%) | Sci LifeLab / KTH \| NR \| NR | 99.4%, 0.0% (Used author-adjusted estimate) | 6.8% (5.4-8.3%) |
| Swedish Public Health Agency (Sweden Public Health Authority)[255] | 2020-05-25 to 2020-05-31 | Örebro, Västerbotten Västra Götaland, Skåne, Jönköping, Kalmar, Uppsala, Stockholm, Jämtland-Härjedalen | National | High | 1200 | NR | NR | Non-probability | Residual sera | 5.7% (4.0-7.7%) | Sci LifeLab / KTH \| NR \| NR | 99.4%, 0.0% (Used author-adjusted estimate) | 5.7% (4.5-7.1%) |
| Swedish Public Health | 2020-05-18 to 2020-05-24 | Örebro, Skåne, | National | High | 1200 | NR | NR | Non-probability | Residual sera | 5.2% (3.7-7.0%) | Sci LifeLab / KTH \| NR \| NR | 99.4%, 0.0% (Used author- | 5.2% (4.0-6.5%) |

| Article Author (Study Organization) | Sampling Dates (YMD) | Location | Geographic Scope | Overall Risk of Bias | Sample Size | Age | Female (%) | Sampling method | Sample frame | Uncorrected Seroprevalence (95% Confidence Interval) | Test Manufacturers \| Names \| Isotypes | Test Sens, Spec (Source) | Corrected Seroprevalence (95% Credible Interval) |
|---|---|---|---|---|---|---|---|---|---|---|---|---|---|
| Agency (Sweden Public Health Authority)[255] | | Stockholm, Jönköping, Kalmar, Uppsala, Västerbotten Västra Götaland, Jämtland-Härjedalen | | | | | | | | | | adjusted estimate) | |
| Roxhed (KTH Royal Institute of Technology)[256] | 2020-04-01 to 2020-05-21 | Uppland \| Stockholm | Local | Moderate | 878 | 20-29 (n=135); 30-39 (n=174); 40-49 (n=165); 50-59 (n=151); 60-69 (n=130); 70-74 (n=79); Missing (n=44) | 55.0 | Probability | Household and community samples | 5.4% | Author-designed \| N/A - Author-designed \| IgG, IgM | 100.0%, 100.0% (Test developer / manufacturer evaluation) | 5.4% (3.9-7.1%) |
| Swedish Public Health Agency (Sweden Public Health Authority)[255] | 2020-05-11 to 2020-05-17 | Örebro, Västerbotten Västra Götaland, Skåne, Jönköping, Kalmar, Uppsala, Stockholm, Jämtland-Härjedalen | National | High | 1200 | NR | NR | Non-probability | Residual sera | 4.5% (3.1-6.2%) | Sci LifeLab / KTH \| NR \| NR | 99.4%, 0.0% (Used author-adjusted estimate) | 4.5% (3.4-5.8%) |
| Swedish Public Health Agency (Sweden Public Health Authority)[255] | 2020-05-04 to 2020-05-10 | Örebro, Västerbotten Västra Götaland, Skåne, Jönköping, Kalmar, Uppsala, Stockholm, Jämtland-Härjedalen | National | High | 1200 | NR | NR | Non-probability | Residual sera | 3.9% (2.6-5.4%) | Sci LifeLab / KTH \| NR \| NR | 99.4%, 0.0% (Used author-adjusted estimate) | 3.9% (2.9-5.0%) |
| Swedish Public Health Agency (Sweden | 2020-04-27 to 2020-05-03 | Örebro, Västerbotten Västra Götaland, | National | High | 1200 | NR | NR | Non-probability | Residual sera | 4.0% (2.7-5.7%) | Sci LifeLab / KTH \| NR \| NR | 99.4%, 0.0% (Used author-adjusted estimate) | 4.0% (3.0-5.2%) |

| Article Author (Study Organization) | Sampling Dates (YMD) | Location | Geographic Scope | Overall Risk of Bias | Sample Size | Age | Female (%) | Sampling method | Sample frame | Uncorrected Seroprevalence (95% Confidence Interval) | Test Manufacturers \| Names \| Isotypes | Test Sens, Spec (Source) | Corrected Seroprevalence (95% Credible Interval) |
|---|---|---|---|---|---|---|---|---|---|---|---|---|---|
| Public Health Authority)[255] | | Skåne, Jönköping, Kalmar, Uppsala, Stockholm, Jämtland-Härjedalen | | | | | | | | | | | |
| Swedish Public Health Agency (Sweden Public Health Authority)[255] | 2020-04-20 to 2020-04-26 | Örebro, Västerbotten Västra Götaland, Skåne, Jönköping, Kalmar, Uppsala, Stockholm, Jämtland-Härjedalen | National | High | 1200 | NR | NR | Non-probability | Residual sera | 5.3% (3.8-7.1%) | Sci LifeLab / KTH \| NR \| NR | 99.4%, 0.0% (Used author-adjusted estimate) | 5.3% (4.1-6.6%) |
| **Switzerland** | | | | | | | | | | | | | |
| Ulyte (University of Zurich)[259] | 2020-06-16 to 2020-11-19 | Zurich \| Zurich | Regional | Low | 2507 | Median 8 (lower school); 11 (middle school); 14 (upper school) (Range 6-10 (lower school); 8-13 (middle school); 11-16 (upper school)) | NR | Probability | Household and community samples | 8.2% | Institute of Medical Virology (IMV) of the University of Zurich \| ABCORA 2.0 binding assay \| IgG, IgA, IgM | 94.3%, 99.0% (Test developer / manufacturer evaluation) | 6.8% (2.5-9.8%) |
| Anker (Université de Fribourg)[260] | 2020-07-08 to 2020-10-14 | Canton of Fribourg | Regional | Moderate | 418 | Mean 58 (SD 17) | 54.1 | Probability | Household and community samples | 8.0% (4.0-12.0%) | N/A - institutionally designed (Centre Hospitalier Universitaire Vaudois (CHUV); l'École Polytechnique Fédérale de Lausanne (EPFL) et l'Institut Suisse | 96.6%, 99.7% (Used author-adjusted estimate) | 8.0% (5.6-10.8%) |

| Article Author (Study Organization) | Sampling Dates (YMD) | Location | Geographic Scope | Overall Risk of Bias | Sample Size | Age | Female (%) | Sampling method | Sample frame | Uncorrected Seroprevalence (95% Confidence Interval) | Test Manufacturers \| Names \| Isotypes | Test Sens, Spec (Source) | Corrected Seroprevalence (95% Credible Interval) |
|---|---|---|---|---|---|---|---|---|---|---|---|---|---|
| | | | | | | | | | | | de Recherche sur les Vaccins) \| SenASTrIS (Sensitive Anti-SARS-CoV-2 Spike Trimer Immunoglobulin Serological) \| IgG, IgA | | |
| Universita della Svizzera Italiana (The Institute of Public Health on the Università della Svizzera italiana)[262] | 2020-07-01 to 2020-07-31 | | Local | Unclear | 647 | 20-64 | NR | Probability | Household and community samples | 11.0% | Centre Hospitalier Universitaire Vaudois (CHUV) in collaboration with the Swiss Federal Institute of Technology in Lausanne (EFPL) and the Swiss Centre for Vaccine Research. \| NR \| NR | No data | - |
| Emmenegger (University of Zurich)[263] | 2020-07-01 to 2020-07-15 | Zurich \| Zurich | Local | Moderate | 1067 | NR | NR | Non-probability | Blood donors | 0.7% (0.3-1.2%) | Author-designed \| Tripartite Automated Blood Immunoassay \| IgG | 100.0%, 100.0% (Test developer / manufacturer evaluation) | 0.4% (0.0-1.0%) |
| Emmenegger (University of Zurich)[263] | 2020-05-15 to 2020-07-15 | Zurich \| Zurich | Local | Moderate | 12831 | NR | NR | Non-probability | Residual sera | 0.9% | Author-designed \| Tripartite Automated Blood Immunoassay \| IgG | 100.0%, 100.0% (Test developer / manufacturer evaluation) | 0.5% (0.0-0.9%) |
| Ulyte (University of Zurich)[264] | 2020-06-16 to 2020-07-09 | Zurich \| Zurich | Local | Moderate | 577 | NR | NR | Probability | Household and community samples | 3.3% (1.4-5.5%) | Swiss Federal Institute of Technology in Lausanne; Swiss Vaccine Center; Centre Hospitalier Universitaire | 98.3%, 98.4% (Used author-adjusted estimate) | 3.3% (2.1-5.0%) |

| Article Author (Study Organization) | Sampling Dates (YMD) | Location | Geographic Scope | Overall Risk of Bias | Sample Size | Age | Female (%) | Sampling method | Sample frame | Uncorrected Seroprevalence (95% Confidence Interval) | Test Manufacturers \| Names \| Isotypes | Test Sens, Spec (Source) | Corrected Seroprevalence (95% Credible Interval) |
|---|---|---|---|---|---|---|---|---|---|---|---|---|---|
| | | | | | | | | | | | Vaudois \| SenASTrIS (Sensitive Anti-SARS-COV-2 Spike Trimer Immunoglobulin Serological) \| IgG, IgA | | |
| Bi (Johns Hopkins Bloomberg School of Public Health)[265] | 2020-04-03 to 2020-06-30 | Geneva | Regional | Moderate | 4534 | Median 53 (IQR 34-65) | 53.6 | Non-probability | Household and community samples | 6.6% (6.0-7.0%) | Euroimmun \| NR \| IgG | 0.6%, 0.9% (FINDDx / MUHC independent evaluation) | - |
| Richard (Geneva University Hospital)[266] | 2020-04-06 to 2020-06-30 | Canton of Geneva | Regional | Low | 8344 | Mean 46.9 (Range 5-94) | 53.5 | Probability | Household and community samples | 7.8% (6.8-8.9%) | Euroimmun \| SARS-CoV-2 IgG ELISA \| IgG | 93.0%, 100.0% (Used author-adjusted estimate) | 7.8% (7.2-8.4%) |
| Fenwick (Lausanne University Hospital and University of Lausanne)[267] | 2020-05-04 to 2020-06-27 | Vaud | Regional | High | 311 | NR | NR | Probability | Household and community samples | 3.9% | Roche \| Elecsys anti-SARS-CoV-2 assay \| NR | 0.9%, 1.0% (FINDDx / MUHC independent evaluation) | - |
| Posfay-Barbe (Geneva University Hospital)[269] | 2020-04-01 to 2020-04-30 | Geneva | Sublocal | High | 208 | Median 9 (IQR 3.6-13.3) | 44.7 | Non-probability | Residual sera | 9.1% | Euroimmun \| Euroimmun IgG assay \| IgG | 0.8%, 1.0% (FINDDx / MUHC independent evaluation) | - |
| Emmenegger (University of Zurich)[263] | 2020-04-01 to 2020-04-30 | Zurich \| Zurich | Local | Moderate | 1469 | NR | NR | Non-probability | Blood donors | 1.2% | Author-designed \| Tripartite Automated Blood Immunoassay \| IgG | 100.0%, 100.0% (Test developer / manufacturer evaluation) | 0.8% (0.0-1.5%) |
| Emmenegger (University of Zurich)[263] | 2020-04-01 to 2020-04-30 | Zurich \| Zurich | Local | Moderate | 4275 | NR | NR | Non-probability | Residual sera | 1.4% (1.0-1.7%) | Author-designed \| Tripartite Automated Blood Immunoassay \| IgG | 100.0%, 100.0% (Test developer / manufacturer evaluation) | 0.9% (0.0-1.5%) |

| Article Author (Study Organization) | Sampling Dates (YMD) | Location | Geographic Scope | Overall Risk of Bias | Sample Size | Age | Female (%) | Sampling method | Sample frame | Uncorrected Seroprevalence (95% Confidence Interval) | Test Manufacturers \| Names \| Isotypes | Test Sens, Spec (Source) | Corrected Seroprevalence (95% Credible Interval) |
|---|---|---|---|---|---|---|---|---|---|---|---|---|---|
| Emmenegger (University of Zurich)[263] | 2020-03-01 to 2020-03-31 | Zurich \| Zurich | Local | Moderate | 3806 | NR | NR | Non-probability | Residual sera | 0.3% (0.1-0.5%) | Author-designed \| Tripartite Automated Blood Immunoassay \| IgG | 100.0%, 100.0% (Test developer / manufacturer evaluation) | 0.2% (0.0-0.4%) |
| **The United Kingdom** | | | | | | | | | | | | | |
| Office for National Statistics (Office of National Statistics)[271] | 2020-12-18 to 2021-01-14 | England | Regional | Moderate | 24150 | older | NR | Probability | Household and community samples | 15.3% (14.7-15.9%) | University of Oxford \| Oxford immunoassay \| IgG | 99.0%, 99.1% (Author-reported independent evaluation) | 14.6% (13.9-15.4%) |
| Office for National Statistics (Office of National Statistics)[271] | 2020-12-18 to 2021-01-14 | Northern Ireland | Regional | High | 313 | older | NR | Probability | Household and community samples | 12.0% (7.8-17.4%) | University of Oxford \| Oxford immunoassay \| IgG | 99.0%, 99.1% (Author-reported independent evaluation) | 11.2% (7.8-15.1%) |
| Office for National Statistics (Office of National Statistics)[271] | 2020-12-18 to 2021-01-14 | Scotland | Regional | Moderate | 1832 | older | NR | Probability | Household and community samples | 10.7% (9.1-12.4%) | University of Oxford \| Oxford immunoassay \| IgG | 99.0%, 99.1% (Author-reported independent evaluation) | 9.9% (8.4-11.5%) |
| Office for National Statistics (Office of National Statistics)[271] | 2020-12-18 to 2021-01-14 | Wales | Regional | Moderate | 732 | older | NR | Probability | Household and community samples | 11.5% (8.6-14.9%) | University of Oxford \| Oxford immunoassay \| IgG | 99.0%, 99.1% (Author-reported independent evaluation) | 10.8% (8.4-13.3%) |
| Office for National Statistics (Office of National Statistics)[272] | 2020-11-20 to 2020-12-17 | England | Regional | Moderate | 21684 | older | NR | Probability | Household and community samples | 10.1% (9.6-10.6%) | University of Oxford \| Oxford immunoassay \| IgG | 99.0%, 99.1% (Author-reported independent evaluation) | 9.3% (8.5-10.0%) |
| Office for National Statistics (Office of National Statistics)[272] | 2020-11-20 to 2020-12-17 | Northern Ireland | Regional | High | 248 | older | NR | Probability | Household and community samples | 3.7% (1.7-7.1%) | University of Oxford \| Oxford immunoassay \| IgG | 99.0%, 99.1% (Author-reported independent evaluation) | 3.0% (0.7-5.6%) |

| Article Author (Study Organization) | Sampling Dates (YMD) | Location | Geographic Scope | Overall Risk of Bias | Sample Size | Age | Female (%) | Sampling method | Sample frame | Uncorrected Seroprevalence (95% Confidence Interval) | Test Manufacturers \| Names \| Isotypes | Test Sens, Spec (Source) | Corrected Seroprevalence (95% Credible Interval) |
|---|---|---|---|---|---|---|---|---|---|---|---|---|---|
| Office for National Statistics (Office of National Statistics)[272] | 2020-11-20 to 2020-12-17 | Scotland | Regional | Moderate | 1034 | older | NR | Probability | Household and community samples | 5.9% (4.4-7.6%) | University of Oxford \| Oxford immunoassay \| IgG | 99.0%, 99.1% (Author-reported independent evaluation) | 5.1% (3.5-6.7%) |
| Office for National Statistics (Office of National Statistics)[272] | 2020-11-20 to 2020-12-17 | Wales | Regional | Moderate | 631 | older | NR | Probability | Household and community samples | 7.2% (5.1-9.9%) | University of Oxford \| Oxford immunoassay \| IgG | 99.0%, 99.1% (Author-reported independent evaluation) | 6.3% (4.3-8.6%) |
| Office for National Statistics (Office of National Statistics)[275] | 2020-10-23 to 2020-11-19 | Scotland | Regional | High | 397 | older | NR | Probability | Household and community samples | 7.8% (5.0-11.7%) | University of Oxford \| Oxford immunoassay \| IgG | 99.0%, 99.1% (Author-reported independent evaluation) | 6.8% (4.2-9.8%) |
| Office for National Statistics (Office of National Statistics)[275] | 2020-10-23 to 2020-11-19 | Northern Ireland | Regional | High | 333 | older | NR | Probability | Household and community samples | 3.5% (1.6-6.4%) | University of Oxford \| Oxford immunoassay \| IgG | 99.0%, 99.1% (Author-reported independent evaluation) | 2.5% (0.7-4.9%) |
| Office for National Statistics (Office of National Statistics)[275] | 2020-10-23 to 2020-11-19 | Wales | Regional | High | 497 | older | NR | Probability | Household and community samples | 6.1% (4.0-9.0%) | University of Oxford \| Oxford immunoassay \| IgG | 99.0%, 99.1% (Author-reported independent evaluation) | 5.3% (3.2-7.6%) |
| Office for National Statistics (Office of National Statistics)[275] | 2020-10-23 to 2020-11-19 | England | Regional | Moderate | 15834 | older | NR | Probability | Household and community samples | 8.7% (8.1-9.4%) | University of Oxford \| Oxford immunoassay \| IgG | 99.0%, 99.1% (Author-reported independent evaluation) | 7.9% (7.1-8.6%) |
| Office for National Statistics (Office of National Statistics)[276] | 2020-09-25 to 2020-10-22 | England | Regional | Moderate | 15687 | older | NR | Probability | Household and community samples | 6.9% (6.4-7.5%) | University of Oxford \| Oxford immunoassay \| IgG | 99.0%, 99.1% (Author-reported independent evaluation) | 6.0% (5.3-6.7%) |
| Office for National | 2020-09-25 to 2020-10-22 | Northern Ireland | Regional | High | 249 | older | NR | Probability | Household and | 2.8% (1.2-5.5%) | University of Oxford \| | 99.0%, 99.1% (Author- | 1.7% (0.0-3.9%) |

| Article Author (Study Organization) | Sampling Dates (YMD) | Location | Geographic Scope | Overall Risk of Bias | Sample Size | Age | Female (%) | Sampling method | Sample frame | Uncorrected Seroprevalence (95% Confidence Interval) | Test Manufacturers \| Names \| Isotypes | Test Sens, Spec (Source) | Corrected Seroprevalence (95% Credible Interval) |
|---|---|---|---|---|---|---|---|---|---|---|---|---|---|
| Statistics (Office of National Statistics)[276] | | | | | | | | | community samples | | Oxford immunoassay \| IgG | reported independent evaluation) | |
| Office for National Statistics (Office of National Statistics)[276] | 2020-09-25 to 2020-10-22 | Scotland | Regional | High | 383 | older | NR | Probability | Household and community samples | 6.8% (4.3-10.2%) | University of Oxford \| Oxford immunoassay \| IgG | 99.0%, 99.1% (Author-reported independent evaluation) | 6.0% (3.6-8.8%) |
| Office for National Statistics (Office of National Statistics)[276] | 2020-09-25 to 2020-10-22 | Wales | Regional | High | 473 | older | NR | Probability | Household and community samples | 4.7% (2.9-7.3%) | University of Oxford \| Oxford immunoassay \| IgG | 99.0%, 99.1% (Author-reported independent evaluation) | 3.9% (1.9-6.1%) |
| Ward (Imperial College London)[277] | 2020-09-15 to 2020-09-28 | | National | Moderate | 159367 | 18-24 (n= 8763); 25-34 (n= 20212); 35-44 (n=26687); 45-54 (n=32403); 55-64 (n=32870); 65-74 (n=26542); 75+ (n=11890) | 56.4 | Probability | Household and community samples | 4.4% (4.3-4.5%) | Fortress Diagnostics; Northern Ireland \| LFIA IgG Fortress Diagnostics \| IgG | 84.4%, 98.6% (Test developer / manufacturer evaluation) | 2.3% (0.0-4.5%) |
| Office for National Statistics (Office of National Statistics)[278] | 2020-08-28 to 2020-09-24 | Wales | Regional | High | 236 | older | NR | Probability | Household and community samples | 4.7% (2.0-9.2%) | University of Oxford \| Oxford immunoassay \| IgG | 99.0%, 99.1% (Author-reported independent evaluation) | 4.0% (1.3-7.2%) |
| Office for National Statistics (Office of National Statistics)[278] | 2020-08-28 to 2020-09-24 | England | Regional | Moderate | 8742 | older | NR | Probability | Household and community samples | 5.8% (5.1-6.5%) | University of Oxford \| Oxford immunoassay \| IgG | 99.0%, 99.1% (Author-reported independent evaluation) | 4.9% (4.1-5.7%) |
| Public Health England (Public Health England)[279] | 2020-08-24 to 2020-09-18 | England | National | Moderate | 8230 | >16 | NR | Probability | Blood donors | 6.1% (5.4-6.8%) | EUROIMMUN \| Euroimmun Anti-SARS-CoV-2 ELISA IgG \| IgG | 83.0%, 99.3% (Used author-adjusted estimate) | 6.1% (5.6-6.6%) |

| Article Author (Study Organization) | Sampling Dates (YMD) | Location | Geographic Scope | Overall Risk of Bias | Sample Size | Age | Female (%) | Sampling method | Sample frame | Uncorrected Seroprevalence (95% Confidence Interval) | Test Manufacturers \| Names \| Isotypes | Test Sens, Spec (Source) | Corrected Seroprevalence (95% Credible Interval) |
|---|---|---|---|---|---|---|---|---|---|---|---|---|---|
| Abo-Leyah (Ninewells Hospital and Medical School)[280] | 2020-05-28 to 2020-09-02 | Scotland | Regional | High | 231 | NR | NR | Probability | Residual sera | 4.8% | Siemens \| Siemens SARS-CoV-2 antibody assay \| NR | 0.9%, 1.0% (FINDDx / MUHC independent evaluation) | - |
| Public Health England (Public Health England)[281] | 2020-07-27 to 2020-08-30 | England | National | Moderate | 8764 | >16 | NR | Probability | Blood donors | 5.3% (4.8-5.8%) | EUROIMMUN \| Euroimmun Anti-SARS-CoV-2 ELISA IgG \| IgG | 83.0%, 99.3% (Used author-adjusted estimate) | 5.3% (4.8-5.8%) |
| Office for National Statistics (Office of National Statistics)[282] | 2020-07-31 to 2020-08-27 | England | Regional | Moderate | 6189 | older | NR | Probability | Household and community samples | 5.6% (4.7-6.5%) | University of Oxford \| Oxford immunoassay \| IgG | 99.0%, 99.1% (Author-reported independent evaluation) | 4.7% (3.8-5.5%) |
| Public Health England (Public Health England)[279] | 2020-06-15 to 2020-08-16 | England | National | Moderate | 4315 | None | NR | Probability | Residual sera | 6.5% (5.5-7.6%) | Abbott \| Abbott Architect SARS-CoV-2 IgG assay \| IgG | 0.4%, 1.0% (FINDDx / MUHC independent evaluation) | - |
| Public Health England (Public Health England)[279] | 2020-06-15 to 2020-08-16 | England | National | Low | 1212 | <30 | NR | Probability | Residual sera | 5.2% (3.9-7.0%) | Abbott \| Abbott Architect SARS-CoV-2 IgG assay \| IgG | 0.4%, 1.0% (FINDDx / MUHC independent evaluation) | - |
| Ward (Imperial College London)[277] | 2020-07-31 to 2020-08-13 | | National | Moderate | 105829 | 18-24 (n= 6493); 25-34 (n= 13573); 35-44 (n=17130); 45-54 (n=21487); 55-64 (n=21840); 65-74 (n=17617); 75+ (n=7689) | 56.3 | Probability | Household and community samples | 4.7% (4.6-4.8%) | Fortress Diagnostics; Northern Ireland \| LFIA IgG Fortress Diagnostics \| IgG | 84.4%, 98.6% (Test developer / manufacturer evaluation) | 2.6% (0.0-4.9%) |
| Public Health Scotland (Public Health Scotland)[285] | 2020-06-29 to 2020-08-09 | Scotland | National | Moderate | 3220 | NR | NR | Non-probability | Blood donors | 3.1% (2.5-3.8%) | NR \| NR \| NR | 87.5%, 98.6% (Used author-adjusted estimate) | 3.1% (2.5-3.7%) |
| Public Health Scotland (Public Health Scotland)[285] | 2020-04-20 to 2020-08-09 | Scotland | National | Moderate | 9802 | 0-19 (n=2341); 20-39 (n=2407); 40- | 50.5 | Non-probability | Residual sera | 4.9% | NR \| NR \| NR | 87.5%, 98.6% (Test developer / manufacturer evaluation) | 2.6% (0.1-5.0%) |

| Article Author (Study Organization) | Sampling Dates (YMD) | Location | Geographic Scope | Overall Risk of Bias | Sample Size | Age | Female (%) | Sampling method | Sample frame | Uncorrected Seroprevalence (95% Confidence Interval) | Test Manufacturers \| Names \| Isotypes | Test Sens, Spec (Source) | Corrected Seroprevalence (95% Credible Interval) |
|---|---|---|---|---|---|---|---|---|---|---|---|---|---|
| | | | | | | 59 (n=2171); 60+ (n=2883) | | | | | | | |
| Public Health England (Public Health England)[281] | 2020-05-01 to 2020-08-05 | England | National | Moderate | 568 | <20 | NR | Probability | Household and community samples | 5.2% (3.2-7.8%) | Abbott \| Abbott Architect SARS-CoV-2 IgG assay \| IgG | 0.4%, 1.0% (FINDDx / MUHC independent evaluation) | - |
| Office for National Statistics (Office of National Statistics)[282] | 2020-07-03 to 2020-07-30 | England | Regional | Moderate | 3236 | older | NR | Probability | Household and community samples | 6.5% (5.2-8.1%) | University of Oxford \| Oxford immunoassay \| IgG | 99.0%, 99.1% (Author-reported independent evaluation) | 5.6% (4.5-6.6%) |
| Public Health England (Public Health England)[288] | 2020-06-29 to 2020-07-28 | England | National | Moderate | 9388 | >16 | NR | Probability | Blood donors | 5.9% (5.3-6.5%) | EUROIMMUN \| Euroimmun Anti-SARS-CoV-2 ELISA IgG \| IgG | 83.0%, 99.3% (Used author-adjusted estimate) | 5.9% (5.4-6.4%) |
| Ward (Imperial College London)[290] | 2020-06-20 to 2020-07-13 | England | National | Low | 99908 | 18-24 (n=6499); 25-34 (n=13366); 35-44 (n=17052); 45-54 (n=20634); 55-64 (n=20404); 65-74 (n=15543); 75+ (n=6410) | 56.1 | Probability | Household and community samples | 5.6% (5.4-5.7%) | Fortress Diagnostics; Northern Ireland \| Fortress Diagnostics LFIA \| IgG | 84.4%, 98.6% (Test developer / manufacturer evaluation) | 3.5% (0.2-6.1%) |
| Office for National Statistics (Office of National Statistics)[295] | 2020-06-05 to 2020-07-02 | England | Regional | Moderate | 2392 | older | NR | Probability | Household and community samples | 4.2% | University of Oxford \| Oxford immunoassay \| IgG | 99.0%, 99.1% (Author-reported independent evaluation) | 3.2% (2.2-4.3%) |
| Public Health England (Public Health England)[288] | 2020-03-16 to 2020-06-30 | England | National | Moderate | 6000 | >64 | NR | Probability | Residual sera | 3.9% | EUROIMMUN \| Euroimmun Anti-SARS-CoV-2 ELISA IgG \| IgG | 0.0%, 0.0% (Used author-adjusted estimate) | 3.9% (3.4-4.4%) |
| Public Health England (Public Health England)[299] | 2020-06-01 to 2020-06-28 | England | National | Moderate | 10318 | 17-69 | NR | Probability | Blood donors | 7.2% (6.6-7.8%) | EUROIMMUN \| Euroimmun Anti-SARS-CoV-2 ELISA IgG \| IgG | 82.5%, 99.1% (Used author-adjusted estimate) | 7.2% (6.7-7.7%) |

| Article Author (Study Organization) | Sampling Dates (YMD) | Location | Geographic Scope | Overall Risk of Bias | Sample Size | Age | Female (%) | Sampling method | Sample frame | Uncorrected Seroprevalence (95% Confidence Interval) | Test Manufacturers \| Names \| Isotypes | Test Sens, Spec (Source) | Corrected Seroprevalence (95% Credible Interval) |
|---|---|---|---|---|---|---|---|---|---|---|---|---|---|
| Arif (Watford General Hospital)[306] | 2020-03-27 to 2020-06-08 | Watford | Sublocal | High | 443 | Mean 50.51 (NR) | 50.6 | Non-probability | Residual sera | 1.6% | NR \| NR \| NR | No data | - |
| Office for National Statistics (Office of National Statistics)[312] | 2020-05-08 to 2020-06-04 | England | Regional | Moderate | 1575 | older | NR | Probability | Household and community samples | 6.3% (4.5-8.7%) | University of Oxford \| Oxford immunoassay \| IgG | 99.0%, 99.1% (Author-reported independent evaluation) | 5.4% (4.1-6.9%) |
| Wells (King's College London)[313] | 2020-04-27 to 2020-06-02 | South East England | Regional | High | 431 | Mean 48.38 (SD 28) | 85.2 | Non-probability | Household and community samples | 12.0% (9.1-15.2%) | Imperial College London \| NR \| IgG, IgM | 90.0%, 100.0% (Author-reported independent evaluation) | 12.6% (4.3-21.6%) |
| Public Health England (Public Health England)[299] | 2020-05-04 to 2020-05-31 | England | National | Moderate | 7686 | 17-69 | NR | Probability | Blood donors | 8.3% (7.5-9.2%) | EUROIMMUN \| Euroimmun Anti-SARS-CoV-2 ELISA IgG \| IgG | 82.5%, 99.1% (Used author-adjusted estimate) | 8.3% (7.7-8.9%) |
| Public Health England (Public Health England)[281] | 2020-05-01 to 2020-05-31 | England | National | Moderate | 1061 | <30 | NR | Probability | Residual sera | 7.3% (5.6-9.2%) | Abbott \| Abbott Architect SARS-CoV-2 IgG assay \| IgG | 95.7%, 99.1% (Used author-adjusted estimate) | 7.3% (5.8-8.9%) |
| Hughes (The Francis Crick Institute)[316] | 2020-03-16 to 2020-05-24 | Scotland \| Glasgow | Local | Moderate | 6635 | 18-44 (n=1,662); 45-64 (n=2202); 65-74 (n=1244); >75 (n=1527) | 53.4 | Non-probability | Residual sera | 7.8% (7.2-8.5%) | Francis Crick Institute \| In house ELISA \| IgG | 95.3%, 97.2% (Test developer / manufacturer evaluation) | 5.3% (2.9-7.0%) |
| Thompson (University of Oxford)[317] | 2020-03-17 to 2020-05-18 | Scotland | Regional | Moderate | 3500 | Median 47 (IQR 34-56) | 53.4 | Probability | Blood donors | 3.2% | N/A - Author designed \| N/A - Author designed \| NR | 94.0%, 100.0% (Test developer / manufacturer evaluation) | 2.9% (0.5-4.7%) |
| Public Health England (Public Health England)[319] | 2020-04-01 to 2020-04-30 | England | National | Moderate | 647 | <30 | NR | Probability | Residual sera | 6.1% (4.1-8.4%) | Abbott \| Abbott Architect SARS-CoV-2 IgG assay \| IgG | 95.7%, 99.1% (Used author-adjusted estimate) | 6.1% (4.4-8.1%) |
| Public Health England (Public Health England)[319] | 2020-04-01 to 2020-04-30 | England | National | Moderate | 201 | <20 | NR | Probability | Household and community samples | 3.8% (1.2-7.5%) | Abbott \| Abbott Architect SARS-CoV-2 IgG assay \| IgG | 95.7%, 99.1% (Used author-adjusted estimate) | 3.8% (1.6-6.7%) |

| Article Author (Study Organization) | Sampling Dates (YMD) | Location | Geographic Scope | Overall Risk of Bias | Sample Size | Age | Female (%) | Sampling method | Sample frame | Uncorrected Seroprevalence (95% Confidence Interval) | Test Manufacturers \| Names \| Isotypes | Test Sens, Spec (Source) | Corrected Seroprevalence (95% Credible Interval) |
|---|---|---|---|---|---|---|---|---|---|---|---|---|---|
| Public Health England (Public Health England)[320] | 2020-04-24 to 2020-04-26 | England | Regional | Moderate | 865 | >16 | NR | Probability | Blood donors | 5.0% (3.2-7.0%) | EUROIMMUN \| Euroimmun Anti-SARS-CoV-2 ELISA IgG \| IgG | 82.5%, 99.1% (Used author-adjusted estimate) | 5.0% (3.7-6.6%) |
| Public Health England (Public Health England)[320] | 2020-04-23 to 2020-04-24 | England | Regional | Moderate | 1043 | >16 | NR | Probability | Blood donors | 7.3% (5.4-9.4%) | EUROIMMUN \| Euroimmun Anti-SARS-CoV-2 ELISA IgG \| IgG | 82.5%, 99.1% (Used author-adjusted estimate) | 7.3% (5.8-9.0%) |
| Public Health England (Public Health England)[320] | 2020-04-15 to 2020-04-20 | England | Regional | Moderate | 936 | >16 | NR | Probability | Blood donors | 6.2% (4.4-8.4%) | EUROIMMUN \| Euroimmun Anti-SARS-CoV-2 ELISA IgG \| IgG | 79.0%, 99.0% (Used author-adjusted estimate) | 6.2% (4.8-7.9%) |
| Public Health England (Public Health England)[320] | 2020-04-14 to 2020-04-16 | England | Regional | Moderate | 1017 | >16 | NR | Probability | Blood donors | 4.6% (3.0-6.4%) | EUROIMMUN \| Euroimmun Anti-SARS-CoV-2 ELISA IgG \| IgG | 79.0%, 99.0% (Used author-adjusted estimate) | 4.6% (3.4-5.9%) |
| Public Health England (Public Health England)[320] | 2020-04-09 to 2020-04-13 | England \| London | Regional | Moderate | 1085 | >16 | NR | Probability | Blood donors | 11.1% (8.9-13.6%) | EUROIMMUN \| Euroimmun Anti-SARS-CoV-2 ELISA IgG \| IgG | 82.5%, 99.1% (Used author-adjusted estimate) | 11.1% (9.3-13.0%) |
| Public Health England (Public Health England)[320] | 2020-04-02 to 2020-04-03 | England | Regional | Moderate | 916 | >16 | NR | Probability | Blood donors | 2.4% (1.0-4.0%) | EUROIMMUN \| Euroimmun Anti-SARS-CoV-2 ELISA IgG \| IgG | 82.5%, 99.1% (Used author-adjusted estimate) | 2.4% (1.5-3.4%) |
| Public Health England (Public Health England)[319] | 2020-02-01 to 2020-03-31 | England | National | Moderate | 443 | <30 | NR | Probability | Residual sera | 0.8% (0.0-2.5%) | Abbott \| Abbott Architect SARS-CoV-2 IgG assay \| IgG | 95.7%, 99.1% (Used author-adjusted estimate) | 0.8% (0.2-1.8%) |
| Public Health England (Public Health England)[319] | 2020-02-01 to 2020-03-31 | England | National | Moderate | 106 | <20 | NR | Probability | Household and community samples | 0.5% (0.0-3.6%) | Abbott \| Abbott Architect SARS-CoV-2 IgG assay \| IgG | 95.7%, 99.1% (Used author-adjusted estimate) | 0.5% (0.0-2.3%) |
| Public Health England (Public Health England)[320] | 2020-03-26 to 2020-03-27 | England \| London | Regional | Moderate | 757 | >16 | NR | Probability | Blood donors | 2.6% (1.1-4.4%) | EUROIMMUN \| Euroimmun Anti-SARS- | 82.5%, 99.1% (Used author-adjusted estimate) | 2.6% (1.6-3.8%) |

| Article Author (Study Organization) | Sampling Dates (YMD) | Location | Geographic Scope | Overall Risk of Bias | Sample Size | Age | Female (%) | Sampling method | Sample frame | Uncorrected Seroprevalence (95% Confidence Interval) | Test Manufacturers \| Names \| Isotypes | Test Sens, Spec (Source) | Corrected Seroprevalence (95% Credible Interval) |
|---|---|---|---|---|---|---|---|---|---|---|---|---|---|
| | | | | | | | | | | | CoV-2 ELISA IgG \| IgG | | |
| **United States of America** | | | | | | | | | | | | | |
| Maleck (University of Wisconsin)[325] | 2020-10-18 to 2020-12-14 | Wisconsin | Regional | Moderate | 994 | NR | NR | Probability | Household and community samples | 6.8% (4.3-9.4%) | Abbott \| Abbott Architect SARS-CoV- 2 IgG antibody test \| IgG | 0.0%, 99.6% (Used author-adjusted estimate) | 6.8% (5.3-8.4%) |
| Craver (Wake Forest Baptist Medical Center)[326] | 2020-04-15 to 2020-12-09 | North Carolina | Regional | Unclear | 8006 | 16-95 | NR | Probability | Household and community samples | 9.3% | NR \| NR \| NR | 66.0%, 96.6% (Used Bastos SR/MA data; no sens, spec, or author adjustment available) | 9.4% (8.0-10.8%) |
| Stout (Clinical Reference Laboratory Inc.)[327] | 2020-05-12 to 2020-12-06 | | National | High | 126587 | Median 44 (SARS-CoV-2 Negative); 41 (SARS-CoV-2 Positive) (IQR 34-54 (SARS-CoV-2 Negative); 32-50 (SARS-CoV-2 Positive)) | 44.8 | Non-probability | Residual sera | 5.9% | Roche \| Roche Elecsys Anti-SARS-CoV-2 Kit \| NR | 0.9%, 1.0% (FINDDx / MUHC independent evaluation) | - |
| Jolley (Florida Department of Health)[328] | 2020-05-01 to 2020-11-30 | Florida | Regional | Unclear | 630838 | NR | NR | Non-probability | Household and community samples | 11.0% | NR \| NR \| IgG | No data | - |
| Coffman (University of Louisville )[329] | 2020-11-09 to 2020-11-16 | Kentucky | Local | Unclear | 1091 | NR | NR | Probability | Household and community samples | 2.0% | NR \| NR \| NR | No data | - |
| Coffman (University of Louisville )[329] | 2020-11-09 to 2020-11-16 | Kentucky | Local | Unclear | 1709 | NR | NR | Non-probability | Household and community samples | 3.3% | NR \| NR \| NR | No data | - |
| Bajema (CDC COVID-19 Response)[331] | 2020-09-09 to 2020-09-24 | Delaware | Regional | Moderate | 980 | 0-17 (n=6612); 18-49 (n=15157); 50- | NR | Non-probability | Residual sera | 7.5% (4.9-10.5%) | Ortho-Clinical Diagnostics; Abbott; Roche \| Elecsys Anti- | 0.0%, 0.0% (Used author-adjusted estimate) | 7.5% (5.9-9.2%) |

| Article Author (Study Organization) | Sampling Dates (YMD) | Location | Geographic Scope | Overall Risk of Bias | Sample Size | Age | Female (%) | Sampling method | Sample frame | Uncorrected Seroprevalence (95% Confidence Interval) | Test Manufacturers \| Names \| Isotypes | Test Sens, Spec (Source) | Corrected Seroprevalence (95% Credible Interval) |
|---|---|---|---|---|---|---|---|---|---|---|---|---|---|
| | | | | | | 64 (n=13207); ≥65 (n=12933) | | | | | SARS-CoV-2 pan-immunoglobulin immunoassay, ARCHITECT SARS-CoV-2 IgG immunoassay, VIT- ROS SARS-CoV-2 IgG immunoassay \| IgG | | |
| Bajema (CDC COVID-19 Response)[331] | 2020-09-09 to 2020-09-24 | Iowa | Regional | Moderate | 977 | 0-17 (n=6612); 18-49 (n=15157); 50-64 (n=13207); ≥65 (n=12933) | NR | Non-probability | Residual sera | 7.6% (5.6-10.0%) | Ortho-Clinical Diagnostics; Abbott; Roche \| Elecsys Anti-SARS-CoV-2 pan-immunoglobulin immunoassay, ARCHITECT SARS-CoV-2 IgG immunoassay, VIT- ROS SARS-CoV-2 IgG immunoassay \| IgG | 0.0%, 0.0% (Used author-adjusted estimate) | 7.6% (6.0-9.4%) |
| Bajema (CDC COVID-19 Response)[331] | 2020-09-08 to 2020-09-24 | Maine | Regional | Moderate | 987 | 0-17 (n=6612); 18-49 (n=15157); 50-64 (n=13207); ≥65 (n=12933) | NR | Non-probability | Residual sera | 0.5% (0.1-0.9%) | Ortho-Clinical Diagnostics; Abbott; Roche \| Elecsys Anti-SARS-CoV-2 pan-immunoglobulin immunoassay, ARCHITECT SARS-CoV-2 IgG immunoassay, VIT- ROS SARS-CoV-2 IgG immunoassay \| IgG | 0.0%, 0.0% (Used author-adjusted estimate) | 0.5% (0.1-1.0%) |

| Article Author (Study Organization) | Sampling Dates (YMD) | Location | Geographic Scope | Overall Risk of Bias | Sample Size | Age | Female (%) | Sampling method | Sample frame | Uncorrected Seroprevalence (95% Confidence Interval) | Test Manufacturers \| Names \| Isotypes | Test Sens, Spec (Source) | Corrected Seroprevalence (95% Credible Interval) |
|---|---|---|---|---|---|---|---|---|---|---|---|---|---|
| Bajema (CDC COVID-19 Response)[331] | 2020-09-08 to 2020-09-24 | Missouri | Regional | Moderate | 1031 | 0-17 (n=6612); 18-49 (n=15157); 50-64 (n=13207); ≥65 (n=12933) | NR | Non-probability | Residual sera | 3.5% (2.1-5.0%) | Ortho-Clinical Diagnostics; Abbott; Roche \| Elecsys Anti-SARS-CoV-2 pan-immunoglobulin immunoassay, ARCHITECT SARS-CoV-2 IgG immunoassay, VIT- ROS SARS-CoV-2 IgG immunoassay \| IgG | 0.0%, 0.0% (Used author-adjusted estimate) | 3.5% (2.5-4.7%) |
| Bajema (CDC COVID-19 Response)[331] | 2020-09-08 to 2020-09-24 | New Mexico | Regional | Moderate | 1014 | 0-17 (n=6612); 18-49 (n=15157); 50-64 (n=13207); ≥65 (n=12933) | NR | Non-probability | Residual sera | 2.4% (1.2-3.6%) | Ortho-Clinical Diagnostics; Abbott; Roche \| Elecsys Anti-SARS-CoV-2 pan-immunoglobulin immunoassay, ARCHITECT SARS-CoV-2 IgG immunoassay, VIT- ROS SARS-CoV-2 IgG immunoassay \| IgG | 0.0%, 0.0% (Used author-adjusted estimate) | 2.4% (1.6-3.4%) |
| Bajema (CDC COVID-19 Response)[331] | 2020-09-11 to 2020-09-24 | New York | Regional | Moderate | 1168 | 0-17 (n=6612); 18-49 (n=15157); 50-64 (n=13207); ≥65 (n=12933) | NR | Non-probability | Residual sera | 17.0% (14.7-19.2%) | Ortho-Clinical Diagnostics; Abbott; Roche \| Elecsys Anti-SARS-CoV-2 pan-immunoglobulin immunoassay, ARCHITECT SARS-CoV-2 IgG immunoassay, VIT- ROS SARS-CoV-2 IgG | 0.0%, 0.0% (Used author-adjusted estimate) | 17.0% (14.9-19.2%) |

| Article Author (Study Organization) | Sampling Dates (YMD) | Location | Geographic Scope | Overall Risk of Bias | Sample Size | Age | Female (%) | Sampling method | Sample frame | Uncorrected Seroprevalence (95% Confidence Interval) | Test Manufacturers \| Names \| Isotypes | Test Sens, Spec (Source) | Corrected Seroprevalence (95% Credible Interval) |
|---|---|---|---|---|---|---|---|---|---|---|---|---|---|
| | | | | | | | | | | | immunoassay \| IgG | | |
| Bajema (CDC COVID-19 Response)[331] | 2020-09-09 to 2020-09-24 | North Dakota | Regional | High | 113 | 0-17 (n=6612); 18-49 (n=15157); 50-64 (n=13207); ≥65 (n=12933) | NR | Non-probability | Residual sera | 1.2% (0.0-3.4%) | Ortho-Clinical Diagnostics; Abbott; Roche \| Elecsys Anti-SARS-CoV-2 pan-immunoglobulin immunoassay, ARCHITECT SARS-CoV-2 IgG immunoassay, VIT- ROS SARS-CoV-2 IgG immunoassay \| IgG | 0.0%, 0.0% (Used author-adjusted estimate) | 1.2% (0.1-4.1%) |
| Bajema (CDC COVID-19 Response)[331] | 2020-09-09 to 2020-09-24 | Rhode Island | Regional | Moderate | 795 | 0-17 (n=6612); 18-49 (n=15157); 50-64 (n=13207); ≥65 (n=12933) | NR | Non-probability | Residual sera | 2.7% (1.1-4.9%) | Ortho-Clinical Diagnostics; Abbott; Roche \| Elecsys Anti-SARS-CoV-2 pan-immunoglobulin immunoassay, ARCHITECT SARS-CoV-2 IgG immunoassay, VIT- ROS SARS-CoV-2 IgG immunoassay \| IgG | 0.0%, 0.0% (Used author-adjusted estimate) | 2.7% (1.7-3.9%) |
| Bajema (CDC COVID-19 Response)[331] | 2020-09-09 to 2020-09-24 | Texas | Regional | Moderate | 988 | 0-17 (n=6612); 18-49 (n=15157); 50-64 (n=13207); ≥65 (n=12933) | NR | Non-probability | Residual sera | 8.2% (6.2-10.5%) | Ortho-Clinical Diagnostics; Abbott; Roche \| Elecsys Anti-SARS-CoV-2 pan-immunoglobulin immunoassay, ARCHITECT SARS-CoV-2 IgG | 0.0%, 0.0% (Used author-adjusted estimate) | 8.2% (6.6-10.0%) |

| Article Author (Study Organization) | Sampling Dates (YMD) | Location | Geographic Scope | Overall Risk of Bias | Sample Size | Age | Female (%) | Sampling method | Sample frame | Uncorrected Seroprevalence (95% Confidence Interval) | Test Manufacturers \| Names \| Isotypes | Test Sens, Spec (Source) | Corrected Seroprevalence (95% Credible Interval) |
|---|---|---|---|---|---|---|---|---|---|---|---|---|---|
| | | | | | | | | | | | immunoassay, VIT- ROS SARS-CoV-2 IgG immunoassay \| IgG | | |
| Bajema (CDC COVID-19 Response)[331] | 2020-09-09 to 2020-09-24 | Vermont | Regional | High | 497 | 0-17 (n=6612); 18-49 (n=15157); 50-64 (n=13207); ≥65 (n=12933) | NR | Non-probability | Residual sera | 1.7% (0.3-4.2%) | Ortho-Clinical Diagnostics; Abbott; Roche \| Elecsys Anti-SARS-CoV-2 pan-immunoglobulin immunoassay, ARCHITECT SARS-CoV-2 IgG immunoassay, VIT- ROS SARS-CoV-2 IgG immunoassay \| IgG | 0.0%, 0.0% (Used author-adjusted estimate) | 1.7% (0.8-3.0%) |
| Bajema (CDC COVID-19 Response)[331] | 2020-09-08 to 2020-09-24 | Washington D.C | Regional | Moderate | 976 | 0-17 (n=6612); 18-49 (n=15157); 50-64 (n=13207); ≥65 (n=12933) | NR | Non-probability | Residual sera | 6.5% (4.5-8.3%) | Ortho-Clinical Diagnostics; Abbott; Roche \| Elecsys Anti-SARS-CoV-2 pan-immunoglobulin immunoassay, ARCHITECT SARS-CoV-2 IgG immunoassay, VIT- ROS SARS-CoV-2 IgG immunoassay \| IgG | 0.0%, 0.0% (Used author-adjusted estimate) | 6.5% (5.0-8.1%) |
| Bajema (CDC COVID-19 Response)[331] | 2020-09-09 to 2020-09-24 | Wisconsin | Regional | Moderate | 976 | 0-17 (n=6612); 18-49 (n=15157); 50-64 (n=13207); ≥65 (n=12933) | NR | Non-probability | Residual sera | 3.8% (2.2-5.7%) | Ortho-Clinical Diagnostics; Abbott; Roche \| Elecsys Anti-SARS-CoV-2 pan-immunoglobulin | 0.0%, 0.0% (Used author-adjusted estimate) | 3.8% (2.7-5.1%) |

| Article Author (Study Organization) | Sampling Dates (YMD) | Location | Geographic Scope | Overall Risk of Bias | Sample Size | Age | Female (%) | Sampling method | Sample frame | Uncorrected Seroprevalence (95% Confidence Interval) | Test Manufacturers \| Names \| Isotypes | Test Sens, Spec (Source) | Corrected Seroprevalence (95% Credible Interval) |
|---|---|---|---|---|---|---|---|---|---|---|---|---|---|
| | | | | | | | | | | | immunoassay, ARCHITECT SARS-CoV-2 IgG immunoassay, VIT- ROS SARS-CoV-2 IgG immunoassay \| IgG | | |
| Bajema (CDC COVID-19 Response)[331] | 2020-09-09 to 2020-09-24 | Wyoming | Regional | High | 111 | 0-17 (n=6612); 18-49 (n=15157); 50-64 (n=13207); ≥65 (n=12933) | NR | Non-probability | Residual sera | 1.5% (0.0-4.0%) | Ortho-Clinical Diagnostics; Abbott; Roche \| Elecsys Anti-SARS-CoV-2 pan-immunoglobulin immunoassay, ARCHITECT SARS-CoV-2 IgG immunoassay, VIT- ROS SARS-CoV-2 IgG immunoassay \| IgG | 0.0%, 0.0% (Used author-adjusted estimate) | 1.5% (0.1-4.1%) |
| Bajema (CDC COVID-19 Response)[331] | 2020-09-09 to 2020-09-23 | Arizona | Regional | Moderate | 1208 | 0-17 (n=6612); 18-49 (n=15157); 50-64 (n=13207); ≥65 (n=12933) | NR | Non-probability | Residual sera | 5.4% (3.1-8.5%) | Ortho-Clinical Diagnostics; Abbott; Roche \| Elecsys Anti-SARS-CoV-2 pan-immunoglobulin immunoassay, ARCHITECT SARS-CoV-2 IgG immunoassay, VIT- ROS SARS-CoV-2 IgG immunoassay \| IgG | 0.0%, 0.0% (Used author-adjusted estimate) | 5.4% (4.2-6.8%) |
| Bajema (CDC COVID-19 Response)[331] | 2020-09-09 to 2020-09-23 | Indiana | Regional | Moderate | 1155 | 0-17 (n=6612); 18-49 (n=15157); 50- | NR | Non-probability | Residual sera | 4.0% (1.4-8.8%) | Ortho-Clinical Diagnostics; Abbott; Roche \| Elecsys Anti- | 0.0%, 0.0% (Used author-adjusted estimate) | 4.0% (3.0-5.2%) |

| Article Author (Study Organization) | Sampling Dates (YMD) | Location | Geographic Scope | Overall Risk of Bias | Sample Size | Age | Female (%) | Sampling method | Sample frame | Uncorrected Seroprevalence (95% Confidence Interval) | Test Manufacturers \| Names \| Isotypes | Test Sens, Spec (Source) | Corrected Seroprevalence (95% Credible Interval) |
|---|---|---|---|---|---|---|---|---|---|---|---|---|---|
| | | | | | | 64 (n=13207); ≥65 (n=12933) | | | | | SARS-CoV-2 pan-immunoglobulin immunoassay, ARCHITECT SARS-CoV-2 IgG immunoassay, VIT- ROS SARS-CoV-2 IgG immunoassay \| IgG | | |
| Bajema (CDC COVID-19 Response)[331] | 2020-09-10 to 2020-09-23 | Maryland | Regional | Moderate | 1242 | 0-17 (n=6612); 18-49 (n=15157); 50-64 (n=13207); ≥65 (n=12933) | NR | Non-probability | Residual sera | 10.2% (7.9-12.7%) | Ortho-Clinical Diagnostics; Abbott; Roche \| Elecsys Anti-SARS-CoV-2 pan-immunoglobulin immunoassay, ARCHITECT SARS-CoV-2 IgG immunoassay, VIT- ROS SARS-CoV-2 IgG immunoassay \| IgG | 0.0%, 0.0% (Used author-adjusted estimate) | 10.2% (8.6-11.9%) |
| Bajema (CDC COVID-19 Response)[331] | 2020-09-09 to 2020-09-23 | Minnesota | Regional | Moderate | 1033 | 0-17 (n=6612); 18-49 (n=15157); 50-64 (n=13207); ≥65 (n=12933) | NR | Non-probability | Residual sera | 8.0% (4.7-11.4%) | Ortho-Clinical Diagnostics; Abbott; Roche \| Elecsys Anti-SARS-CoV-2 pan-immunoglobulin immunoassay, ARCHITECT SARS-CoV-2 IgG immunoassay, VIT- ROS SARS-CoV-2 IgG immunoassay \| IgG | 0.0%, 0.0% (Used author-adjusted estimate) | 8.0% (6.4-9.7%) |

| Article Author (Study Organization) | Sampling Dates (YMD) | Location | Geographic Scope | Overall Risk of Bias | Sample Size | Age | Female (%) | Sampling method | Sample frame | Uncorrected Seroprevalence (95% Confidence Interval) | Test Manufacturers \| Names \| Isotypes | Test Sens, Spec (Source) | Corrected Seroprevalence (95% Credible Interval) |
|---|---|---|---|---|---|---|---|---|---|---|---|---|---|
| Bajema (CDC COVID-19 Response)[331] | 2020-09-09 to 2020-09-23 | Montana | Regional | High | 508 | 0-17 (n=6612); 18-49 (n=15157); 50-64 (n=13207); ≥65 (n=12933) | NR | Non-probability | Residual sera | 2.2% (0.7-4.4%) | Ortho-Clinical Diagnostics; Abbott; Roche \| Elecsys Anti-SARS-CoV-2 pan-immunoglobulin immunoassay, ARCHITECT SARS-CoV-2 IgG immunoassay, VIT- ROS SARS-CoV-2 IgG immunoassay \| IgG | 0.0%, 0.0% (Used author-adjusted estimate) | 2.2% (1.2-3.7%) |
| Bajema (CDC COVID-19 Response)[331] | 2020-09-09 to 2020-09-23 | New Jersey | Regional | Moderate | 1050 | 0-17 (n=6612); 18-49 (n=15157); 50-64 (n=13207); ≥65 (n=12933) | NR | Non-probability | Residual sera | 15.1% (12.7-17.6%) | Ortho-Clinical Diagnostics; Abbott; Roche \| Elecsys Anti-SARS-CoV-2 pan-immunoglobulin immunoassay, ARCHITECT SARS-CoV-2 IgG immunoassay, VIT- ROS SARS-CoV-2 IgG immunoassay \| IgG | 0.0%, 0.0% (Used author-adjusted estimate) | 15.1% (13.0-17.3%) |
| Bajema (CDC COVID-19 Response)[331] | 2020-09-09 to 2020-09-23 | Pennsylvania | Regional | Moderate | 1341 | 0-17 (n=6612); 18-49 (n=15157); 50-64 (n=13207); ≥65 (n=12933) | NR | Non-probability | Residual sera | 11.1% (8.8-13.7%) | Ortho-Clinical Diagnostics; Abbott; Roche \| Elecsys Anti-SARS-CoV-2 pan-immunoglobulin immunoassay, ARCHITECT SARS-CoV-2 IgG immunoassay, VIT- ROS SARS-CoV-2 IgG | 0.0%, 0.0% (Used author-adjusted estimate) | 11.1% (9.4-12.8%) |

| Article Author (Study Organization) | Sampling Dates (YMD) | Location | Geographic Scope | Overall Risk of Bias | Sample Size | Age | Female (%) | Sampling method | Sample frame | Uncorrected Seroprevalence (95% Confidence Interval) | Test Manufacturers \| Names \| Isotypes | Test Sens, Spec (Source) | Corrected Seroprevalence (95% Credible Interval) |
|---|---|---|---|---|---|---|---|---|---|---|---|---|---|
| | | | | | | | | | | | immunoassay \| IgG | | |
| Bajema (CDC COVID-19 Response)[331] | 2020-09-09 to 2020-09-23 | South Dakota | Regional | High | 99 | 0-17 (n=6612); 18-49 (n=15157); 50-64 (n=13207); ≥65 (n=12933) | NR | Non-probability | Residual sera | 1.8% (0.0-4.8%) | Ortho-Clinical Diagnostics; Abbott; Roche \| Elecsys Anti-SARS-CoV-2 pan-immunoglobulin immunoassay, ARCHITECT SARS-CoV-2 IgG immunoassay, VIT- ROS SARS-CoV-2 IgG immunoassay \| IgG | 0.0%, 0.0% (Used author-adjusted estimate) | 1.8% (0.1-4.6%) |
| Bajema (CDC COVID-19 Response)[331] | 2020-09-10 to 2020-09-23 | Virginia | Regional | Moderate | 1111 | 0-17 (n=6612); 18-49 (n=15157); 50-64 (n=13207); ≥65 (n=12933) | NR | Non-probability | Residual sera | 3.2% (1.8-5.1%) | Ortho-Clinical Diagnostics; Abbott; Roche \| Elecsys Anti-SARS-CoV-2 pan-immunoglobulin immunoassay, ARCHITECT SARS-CoV-2 IgG immunoassay, VIT- ROS SARS-CoV-2 IgG immunoassay \| IgG | 0.0%, 0.0% (Used author-adjusted estimate) | 3.2% (2.2-4.3%) |
| Bajema (CDC COVID-19 Response)[331] | 2020-09-08 to 2020-09-22 | Hawaii | Regional | High | 516 | 0-17 (n=6612); 18-49 (n=15157); 50-64 (n=13207); ≥65 (n=12933) | NR | Non-probability | Residual sera | 0.8% (0.0-2.2%) | Ortho-Clinical Diagnostics; Abbott; Roche \| Elecsys Anti-SARS-CoV-2 pan-immunoglobulin immunoassay, ARCHITECT SARS-CoV-2 IgG | 0.0%, 0.0% (Used author-adjusted estimate) | 0.8% (0.3-1.8%) |

| Article Author (Study Organization) | Sampling Dates (YMD) | Location | Geographic Scope | Overall Risk of Bias | Sample Size | Age | Female (%) | Sampling method | Sample frame | Uncorrected Seroprevalence (95% Confidence Interval) | Test Manufacturers \| Names \| Isotypes | Test Sens, Spec (Source) | Corrected Seroprevalence (95% Credible Interval) |
|---|---|---|---|---|---|---|---|---|---|---|---|---|---|
| | | | | | | | | | | | immunoassay, VIT- ROS SARS-CoV-2 IgG immunoassay \| IgG | | |
| Bajema (CDC COVID-19 Response)[331] | 2020-09-08 to 2020-09-22 | Michigan | Regional | Moderate | 980 | 0-17 (n=6612); 18-49 (n=15157); 50-64 (n=13207); ≥65 (n=12933) | NR | Non-probability | Residual sera | 3.7% (2.5-4.9%) | Ortho-Clinical Diagnostics; Abbott; Roche \| Elecsys Anti-SARS-CoV-2 pan-immunoglobulin immunoassay, ARCHITECT SARS-CoV-2 IgG immunoassay, VIT- ROS SARS-CoV-2 IgG immunoassay \| IgG | 0.0%, 0.0% (Used author-adjusted estimate) | 3.7% (2.6-5.0%) |
| Bajema (CDC COVID-19 Response)[331] | 2020-09-09 to 2020-09-22 | Mississippi | Regional | Moderate | 980 | 0-17 (n=6612); 18-49 (n=15157); 50-64 (n=13207); ≥65 (n=12933) | NR | Non-probability | Residual sera | 7.9% (5.3-10.9%) | Ortho-Clinical Diagnostics; Abbott; Roche \| Elecsys Anti-SARS-CoV-2 pan-immunoglobulin immunoassay, ARCHITECT SARS-CoV-2 IgG immunoassay, VIT- ROS SARS-CoV-2 IgG immunoassay \| IgG | 0.0%, 0.0% (Used author-adjusted estimate) | 7.9% (6.3-9.7%) |
| Bajema (CDC COVID-19 Response)[331] | 2020-09-10 to 2020-09-22 | Ohio | Regional | Moderate | 978 | 0-17 (n=6612); 18-49 (n=15157); 50-64 (n=13207); ≥65 (n=12933) | NR | Non-probability | Residual sera | 2.8% (1.7-4.2%) | Ortho-Clinical Diagnostics; Abbott; Roche \| Elecsys Anti-SARS-CoV-2 pan-immunoglobulin | 0.0%, 0.0% (Used author-adjusted estimate) | 2.8% (1.9-3.9%) |

| Article Author (Study Organization) | Sampling Dates (YMD) | Location | Geographic Scope | Overall Risk of Bias | Sample Size | Age | Female (%) | Sampling method | Sample frame | Uncorrected Seroprevalence (95% Confidence Interval) | Test Manufacturers \| Names \| Isotypes | Test Sens, Spec (Source) | Corrected Seroprevalence (95% Credible Interval) |
|---|---|---|---|---|---|---|---|---|---|---|---|---|---|
| | | | | | | | | | | | immunoassay, ARCHITECT SARS-CoV-2 IgG immunoassay, VIT- ROS SARS-CoV-2 IgG immunoassay \| IgG | | |
| Bajema (CDC COVID-19 Response)[331] | 2020-09-08 to 2020-09-22 | Oregon | Regional | Moderate | 971 | 0-17 (n=6612); 18-49 (n=15157); 50-64 (n=13207); ≥65 (n=12933) | NR | Non-probability | Residual sera | 2.6% (1.5-4.1%) | Ortho-Clinical Diagnostics; Abbott; Roche \| Elecsys Anti-SARS-CoV-2 pan-immunoglobulin immunoassay, ARCHITECT SARS-CoV-2 IgG immunoassay, VIT- ROS SARS-CoV-2 IgG immunoassay \| IgG | 0.0%, 0.0% (Used author-adjusted estimate) | 2.6% (1.7-3.7%) |
| Bajema (CDC COVID-19 Response)[331] | 2020-09-09 to 2020-09-22 | Washington | Regional | Moderate | 968 | 0-17 (n=6612); 18-49 (n=15157); 50-64 (n=13207); ≥65 (n=12933) | NR | Non-probability | Residual sera | 2.5% (1.3-3.8%) | Ortho-Clinical Diagnostics; Abbott; Roche \| Elecsys Anti-SARS-CoV-2 pan-immunoglobulin immunoassay, ARCHITECT SARS-CoV-2 IgG immunoassay, VIT- ROS SARS-CoV-2 IgG immunoassay \| IgG | 0.0%, 0.0% (Used author-adjusted estimate) | 2.5% (1.6-3.6%) |
| Bajema (CDC COVID-19 Response)[331] | 2020-09-09 to 2020-09-22 | West Virginia | Regional | Moderate | 968 | 0-17 (n=6612); 18-49 (n=15157); 50- | NR | Non-probability | Residual sera | 1.3% (0.5-2.2%) | Ortho-Clinical Diagnostics; Abbott; Roche \| Elecsys Anti- | 0.0%, 0.0% (Used author-adjusted estimate) | 1.3% (0.7-2.1%) |

| Article Author (Study Organization) | Sampling Dates (YMD) | Location | Geographic Scope | Overall Risk of Bias | Sample Size | Age | Female (%) | Sampling method | Sample frame | Uncorrected Seroprevalence (95% Confidence Interval) | Test Manufacturers \| Names \| Isotypes | Test Sens, Spec (Source) | Corrected Seroprevalence (95% Credible Interval) |
|---|---|---|---|---|---|---|---|---|---|---|---|---|---|
| | | | | | | 64 (n=13207); ≥65 (n=12933) | | | | | SARS-CoV-2 pan-immunoglobulin immunoassay, ARCHITECT SARS-CoV-2 IgG immunoassay, VIT- ROS SARS-CoV-2 IgG immunoassay \| IgG | | |
| Fountain Hills Times (Maricopa County Department of Public Health)[332] | 2020-09-11 to 2020-09-21 | Arizona | Local | Unclear | 260 | NR | NR | Probability | Household and community samples | 10.7% | NR \| NR \| NR | No data | - |
| Bajema (CDC COVID-19 Response)[331] | 2020-09-09 to 2020-09-19 | Kansas | Regional | Moderate | 1007 | 0-17 (n=6612); 18-49 (n=15157); 50-64 (n=13207); ≥65 (n=12933) | NR | Non-probability | Residual sera | 3.5% (2.3-4.8%) | Ortho-Clinical Diagnostics; Abbott; Roche \| Elecsys Anti-SARS-CoV-2 pan-immunoglobulin immunoassay, ARCHITECT SARS-CoV-2 IgG immunoassay, VIT- ROS SARS-CoV-2 IgG immunoassay \| IgG | 0.0%, 0.0% (Used author-adjusted estimate) | 3.5% (2.5-4.7%) |
| Bajema (CDC COVID-19 Response)[331] | 2020-09-09 to 2020-09-19 | Louisiana | Regional | Moderate | 952 | 0-17 (n=6612); 18-49 (n=15157); 50-64 (n=13207); ≥65 (n=12933) | NR | Non-probability | Residual sera | 12.5% (10.0-15.6%) | Ortho-Clinical Diagnostics; Abbott; Roche \| Elecsys Anti-SARS-CoV-2 pan-immunoglobulin immunoassay, ARCHITECT SARS-CoV-2 | 0.0%, 0.0% (Used author-adjusted estimate) | 12.5% (10.5-14.7%) |

| Article Author (Study Organization) | Sampling Dates (YMD) | Location | Geographic Scope | Overall Risk of Bias | Sample Size | Age | Female (%) | Sampling method | Sample frame | Uncorrected Seroprevalence (95% Confidence Interval) | Test Manufacturers \| Names \| Isotypes | Test Sens, Spec (Source) | Corrected Seroprevalence (95% Credible Interval) |
|---|---|---|---|---|---|---|---|---|---|---|---|---|---|
| | | | | | | | | | | | IgG immunoassay, VIT- ROS SARS-CoV-2 IgG immunoassay \| IgG | | |
| Bajema (CDC COVID-19 Response)[331] | 2020-09-09 to 2020-09-18 | Alabama | Regional | Moderate | 905 | 0-17 (n=6612); 18-49 (n=15157); 50-64 (n=13207); ≥65 (n=12933) | NR | Non-probability | Residual sera | 8.7% (6.2-11.3%) | Ortho-Clinical Diagnostics; Abbott; Roche \| Elecsys Anti-SARS-CoV-2 pan-immunoglobulin immunoassay, ARCHITECT SARS-CoV-2 IgG immunoassay, VIT- ROS SARS-CoV-2 IgG immunoassay \| IgG | 0.0%, 0.0% (Used author-adjusted estimate) | 8.7% (6.9-10.6%) |
| Bajema (CDC COVID-19 Response)[331] | 2020-09-09 to 2020-09-18 | Alaska | Regional | Moderate | 681 | 0-17 (n=6612); 18-49 (n=15157); 50-64 (n=13207); ≥65 (n=12933) | NR | Non-probability | Residual sera | 0.4% (0.1-0.8%) | Ortho-Clinical Diagnostics; Abbott; Roche \| Elecsys Anti-SARS-CoV-2 pan-immunoglobulin immunoassay, ARCHITECT SARS-CoV-2 IgG immunoassay, VIT- ROS SARS-CoV-2 IgG immunoassay \| IgG | 0.0%, 0.0% (Used author-adjusted estimate) | 0.4% (0.1-0.9%) |
| Bajema (CDC COVID-19 Response)[331] | 2020-09-09 to 2020-09-18 | Arkansas | Regional | Moderate | 990 | 0-17 (n=6612); 18-49 (n=15157); 50-64 (n=13207); ≥65 (n=12933) | NR | Non-probability | Residual sera | 6.3% (4.6-8.0%) | Ortho-Clinical Diagnostics; Abbott; Roche \| Elecsys Anti-SARS-CoV-2 pan-immunoglobuli | 0.0%, 0.0% (Used author-adjusted estimate) | 6.3% (4.9-7.9%) |

| Article Author (Study Organization) | Sampling Dates (YMD) | Location | Geographic Scope | Overall Risk of Bias | Sample Size | Age | Female (%) | Sampling method | Sample frame | Uncorrected Seroprevalence (95% Confidence Interval) | Test Manufacturers \| Names \| Isotypes | Test Sens, Spec (Source) | Corrected Seroprevalence (95% Credible Interval) |
|---|---|---|---|---|---|---|---|---|---|---|---|---|---|
| | | | | | | | | | | | n immunoassay, ARCHITECT SARS-CoV-2 IgG immunoassay, VIT- ROS SARS-CoV-2 IgG immunoassay \| IgG | | |
| Bajema (CDC COVID-19 Response)[331] | 2020-09-09 to 2020-09-18 | Colorado | Regional | Moderate | 1032 | 0-17 (n=6612); 18-49 (n=15157); 50-64 (n=13207); ≥65 (n=12933) | NR | Non-probability | Residual sera | 3.3% (1.9-4.9%) | Ortho-Clinical Diagnostics; Abbott; Roche \| Elecsys Anti-SARS-CoV-2 pan-immunoglobulin immunoassay, ARCHITECT SARS-CoV-2 IgG immunoassay, VIT- ROS SARS-CoV-2 IgG immunoassay \| IgG | 0.0%, 0.0% (Used author-adjusted estimate) | 3.3% (2.3-4.5%) |
| Bajema (CDC COVID-19 Response)[331] | 2020-09-09 to 2020-09-18 | Georgia | Regional | Moderate | 1100 | 0-17 (n=6612); 18-49 (n=15157); 50-64 (n=13207); ≥65 (n=12933) | NR | Non-probability | Residual sera | 13.0% (10.5-15.8%) | Ortho-Clinical Diagnostics; Abbott; Roche \| Elecsys Anti-SARS-CoV-2 pan-immunoglobulin immunoassay, ARCHITECT SARS-CoV-2 IgG immunoassay, VIT- ROS SARS-CoV-2 IgG immunoassay \| IgG | 0.0%, 0.0% (Used author-adjusted estimate) | 13.0% (11.1-15.1%) |
| Bajema (CDC COVID-19 Response)[331] | 2020-09-08 to 2020-09-18 | Idaho | Regional | Moderate | 732 | 0-17 (n=6612); 18-49 (n=15157); 50- | NR | Non-probability | Residual sera | 5.2% (2.7-8.5%) | Ortho-Clinical Diagnostics; Abbott; Roche \| | 0.0%, 0.0% (Used author- | 5.2% (3.8-7.0%) |

| Article Author (Study Organization) | Sampling Dates (YMD) | Location | Geographic Scope | Overall Risk of Bias | Sample Size | Age | Female (%) | Sampling method | Sample frame | Uncorrected Seroprevalence (95% Confidence Interval) | Test Manufacturers \| Names \| Isotypes | Test Sens, Spec (Source) | Corrected Seroprevalence (95% Credible Interval) |
|---|---|---|---|---|---|---|---|---|---|---|---|---|---|
| | | | | | | 64 (n=13207); ≥65 (n=12933) | | | | | Elecsys Anti-SARS-CoV-2 pan-immunoglobulin immunoassay, ARCHITECT SARS-CoV-2 IgG immunoassay, VIT- ROS SARS-CoV-2 IgG immunoassay \| IgG | adjusted estimate) | |
| Bajema (CDC COVID-19 Response)[331] | 2020-09-09 to 2020-09-18 | Kentucky | Regional | Moderate | 980 | 0-17 (n=6612); 18-49 (n=15157); 50-64 (n=13207); ≥65 (n=12933) | NR | Non-probability | Residual sera | 3.6% (2.3-5.2%) | Ortho-Clinical Diagnostics; Abbott; Roche \| Elecsys Anti-SARS-CoV-2 pan-immunoglobulin immunoassay, ARCHITECT SARS-CoV-2 IgG immunoassay, VIT- ROS SARS-CoV-2 IgG immunoassay \| IgG | 0.0%, 0.0% (Used author-adjusted estimate) | 3.6% (2.5-4.9%) |
| Bajema (CDC COVID-19 Response)[331] | 2020-09-09 to 2020-09-18 | Nebraska | Regional | Moderate | 976 | 0-17 (n=6612); 18-49 (n=15157); 50-64 (n=13207); ≥65 (n=12933) | NR | Non-probability | Residual sera | 6.7% (4.8-8.9%) | Ortho-Clinical Diagnostics; Abbott; Roche \| Elecsys Anti-SARS-CoV-2 pan-immunoglobulin immunoassay, ARCHITECT SARS-CoV-2 IgG immunoassay, VIT- ROS SARS-CoV-2 IgG immunoassay \| IgG | 0.0%, 0.0% (Used author-adjusted estimate) | 6.7% (5.2-8.4%) |

| Article Author (Study Organization) | Sampling Dates (YMD) | Location | Geographic Scope | Overall Risk of Bias | Sample Size | Age | Female (%) | Sampling method | Sample frame | Uncorrected Seroprevalence (95% Confidence Interval) | Test Manufacturers \| Names \| Isotypes | Test Sens, Spec (Source) | Corrected Seroprevalence (95% Credible Interval) |
|---|---|---|---|---|---|---|---|---|---|---|---|---|---|
| Bajema (CDC COVID-19 Response)[331] | 2020-09-09 to 2020-09-18 | Oklahoma | Regional | Moderate | 980 | 0-17 (n=6612); 18-49 (n=15157); 50-64 (n=13207); ≥65 (n=12933) | NR | Non-probability | Residual sera | 5.0% (3.4-6.5%) | Ortho-Clinical Diagnostics; Abbott; Roche \| Elecsys Anti-SARS-CoV-2 pan-immunoglobulin immunoassay, ARCHITECT SARS-CoV-2 IgG immunoassay, VIT- ROS SARS-CoV-2 IgG immunoassay \| IgG | 0.0%, 0.0% (Used author-adjusted estimate) | 5.0% (3.8-6.5%) |
| Bajema (CDC COVID-19 Response)[331] | 2020-09-09 to 2020-09-18 | South Carolina | Regional | Moderate | 975 | 0-17 (n=6612); 18-49 (n=15157); 50-64 (n=13207); ≥65 (n=12933) | NR | Non-probability | Residual sera | 7.8% (5.6-10.2%) | Ortho-Clinical Diagnostics; Abbott; Roche \| Elecsys Anti-SARS-CoV-2 pan-immunoglobulin immunoassay, ARCHITECT SARS-CoV-2 IgG immunoassay, VIT- ROS SARS-CoV-2 IgG immunoassay \| IgG | 0.0%, 0.0% (Used author-adjusted estimate) | 7.8% (6.2-9.6%) |
| Bajema (CDC COVID-19 Response)[331] | 2020-09-09 to 2020-09-18 | Utah | Regional | Moderate | 1022 | 0-17 (n=6612); 18-49 (n=15157); 50-64 (n=13207); ≥65 (n=12933) | NR | Non-probability | Residual sera | 5.1% (3.3-7.9%) | Ortho-Clinical Diagnostics; Abbott; Roche \| Elecsys Anti-SARS-CoV-2 pan-immunoglobulin immunoassay, ARCHITECT SARS-CoV-2 IgG immunoassay, VIT- ROS SARS-CoV-2 IgG | 0.0%, 0.0% (Used author-adjusted estimate) | 5.1% (3.9-6.6%) |

| Article Author (Study Organization) | Sampling Dates (YMD) | Location | Geographic Scope | Overall Risk of Bias | Sample Size | Age | Female (%) | Sampling method | Sample frame | Uncorrected Seroprevalence (95% Confidence Interval) | Test Manufacturers \| Names \| Isotypes | Test Sens, Spec (Source) | Corrected Seroprevalence (95% Credible Interval) |
|---|---|---|---|---|---|---|---|---|---|---|---|---|---|
| | | | | | | | | | | | immunoassay \| IgG | | |
| Bajema (CDC COVID-19 Response)[331] | 2020-09-08 to 2020-09-17 | Illinois | Regional | Moderate | 992 | 0-17 (n=6612); 18-49 (n=15157); 50-64 (n=13207); ≥65 (n=12933) | NR | Non-probability | Residual sera | 4.5% (3.1-6.1%) | Ortho-Clinical Diagnostics; Abbott; Roche \| Elecsys Anti-SARS-CoV-2 pan-immunoglobulin immunoassay, ARCHITECT SARS-CoV-2 IgG immunoassay, VIT- ROS SARS-CoV-2 IgG immunoassay \| IgG | 0.0%, 0.0% (Used author-adjusted estimate) | 4.5% (3.3-5.9%) |
| Bajema (CDC COVID-19 Response)[331] | 2020-09-09 to 2020-09-17 | New Hampshire | Regional | Moderate | 1034 | 0-17 (n=6612); 18-49 (n=15157); 50-64 (n=13207); ≥65 (n=12933) | NR | Non-probability | Residual sera | 0.7% (0.3-1.3%) | Ortho-Clinical Diagnostics; Abbott; Roche \| Elecsys Anti-SARS-CoV-2 pan-immunoglobulin immunoassay, ARCHITECT SARS-CoV-2 IgG immunoassay, VIT- ROS SARS-CoV-2 IgG immunoassay \| IgG | 0.0%, 0.0% (Used author-adjusted estimate) | 0.7% (0.3-1.3%) |
| Bajema (CDC COVID-19 Response)[331] | 2020-09-09 to 2020-09-17 | North Carolina | Regional | Moderate | 925 | 0-17 (n=6612); 18-49 (n=15157); 50-64 (n=13207); ≥65 (n=12933) | NR | Non-probability | Residual sera | 6.8% (4.8-8.9%) | Ortho-Clinical Diagnostics; Abbott; Roche \| Elecsys Anti-SARS-CoV-2 pan-immunoglobulin immunoassay, ARCHITECT SARS-CoV-2 IgG | 0.0%, 0.0% (Used author-adjusted estimate) | 6.8% (5.2-8.5%) |

| Article Author (Study Organization) | Sampling Dates (YMD) | Location | Geographic Scope | Overall Risk of Bias | Sample Size | Age | Female (%) | Sampling method | Sample frame | Uncorrected Seroprevalence (95% Confidence Interval) | Test Manufacturers \| Names \| Isotypes | Test Sens, Spec (Source) | Corrected Seroprevalence (95% Credible Interval) |
|---|---|---|---|---|---|---|---|---|---|---|---|---|---|
| | | | | | | | | | | | immunoassay, VIT- ROS SARS-CoV-2 IgG immunoassay \| IgG | | |
| Bajema (CDC COVID-19 Response)[331] | 2020-09-10 to 2020-09-16 | California | Regional | Moderate | 979 | 0-17 (n=6612); 18-49 (n=15157); 50-64 (n=13207); ≥65 (n=12933) | NR | Non-probability | Residual sera | 4.9% (3.2-6.8%) | Ortho-Clinical Diagnostics; Abbott; Roche \| Elecsys Anti-SARS-CoV-2 pan-immunoglobulin immunoassay, ARCHITECT SARS-CoV-2 IgG immunoassay, VIT- ROS SARS-CoV-2 IgG immunoassay \| IgG | 0.0%, 0.0% (Used author-adjusted estimate) | 4.9% (3.6-6.3%) |
| Bajema (CDC COVID-19 Response)[331] | 2020-09-08 to 2020-09-16 | Tennessee | Regional | Moderate | 1028 | 0-17 (n=6612); 18-49 (n=15157); 50-64 (n=13207); ≥65 (n=12933) | NR | Non-probability | Residual sera | 6.7% (5.0-8.6%) | Ortho-Clinical Diagnostics; Abbott; Roche \| Elecsys Anti-SARS-CoV-2 pan-immunoglobulin immunoassay, ARCHITECT SARS-CoV-2 IgG immunoassay, VIT- ROS SARS-CoV-2 IgG immunoassay \| IgG | 0.0%, 0.0% (Used author-adjusted estimate) | 6.7% (5.2-8.3%) |
| Bajema (CDC COVID-19 Response)[331] | 2020-09-09 to 2020-09-15 | Massachusetts | Regional | Moderate | 988 | 0-17 (n=6612); 18-49 (n=15157); 50-64 (n=13207); ≥65 (n=12933) | NR | Non-probability | Residual sera | 3.7% (2.2-5.2%) | Ortho-Clinical Diagnostics; Abbott; Roche \| Elecsys Anti-SARS-CoV-2 pan-immunoglobulin | 0.0%, 0.0% (Used author-adjusted estimate) | 3.7% (2.6-5.0%) |

| Article Author (Study Organization) | Sampling Dates (YMD) | Location | Geographic Scope | Overall Risk of Bias | Sample Size | Age | Female (%) | Sampling method | Sample frame | Uncorrected Seroprevalence (95% Confidence Interval) | Test Manufacturers \| Names \| Isotypes | Test Sens, Spec (Source) | Corrected Seroprevalence (95% Credible Interval) |
|---|---|---|---|---|---|---|---|---|---|---|---|---|---|
| | | | | | | | | | | | immunoassay, ARCHITECT SARS-CoV-2 IgG immunoassay, VIT- ROS SARS-CoV-2 IgG immunoassay \| IgG | | |
| Bajema (CDC COVID-19 Response)[331] | 2020-09-08 to 2020-09-15 | Puerto Rico | Regional | Moderate | 970 | 0-17 (n=6612); 18-49 (n=15157); 50-64 (n=13207); ≥65 (n=12933) | NR | Non-probability | Residual sera | 3.0% (1.8-4.3%) | Ortho-Clinical Diagnostics; Abbott; Roche \| Elecsys Anti-SARS-CoV-2 pan-immunoglobulin immunoassay, ARCHITECT SARS-CoV-2 IgG immunoassay, VIT- ROS SARS-CoV-2 IgG immunoassay \| IgG | 0.0%, 0.0% (Used author-adjusted estimate) | 3.0% (2.1-4.2%) |
| Bajema (CDC COVID-19 Response)[331] | 2020-09-09 to 2020-09-14 | Connecticut | Regional | Moderate | 982 | 0-17 (n=6612); 18-49 (n=15157); 50-64 (n=13207); ≥65 (n=12933) | NR | Non-probability | Residual sera | 3.1% (2.1-4.4%) | Ortho-Clinical Diagnostics; Abbott; Roche \| Elecsys Anti-SARS-CoV-2 pan-immunoglobulin immunoassay, ARCHITECT SARS-CoV-2 IgG immunoassay, VIT- ROS SARS-CoV-2 IgG immunoassay \| IgG | 0.0%, 0.0% (Used author-adjusted estimate) | 3.1% (2.1-4.3%) |
| Bajema (CDC COVID-19 Response)[331] | 2020-09-09 to 2020-09-12 | Nevada | Regional | Moderate | 979 | 0-17 (n=6612); 18-49 (n=15157); 50- | NR | Non-probability | Residual sera | 7.8% (5.9-10.0%) | Ortho-Clinical Diagnostics; Abbott; Roche \| Elecsys Anti- | 0.0%, 0.0% (Used author-adjusted estimate) | 7.8% (6.2-9.6%) |

| Article Author (Study Organization) | Sampling Dates (YMD) | Location | Geographic Scope | Overall Risk of Bias | Sample Size | Age | Female (%) | Sampling method | Sample frame | Uncorrected Seroprevalence (95% Confidence Interval) | Test Manufacturers \| Names \| Isotypes | Test Sens, Spec (Source) | Corrected Seroprevalence (95% Credible Interval) |
|---|---|---|---|---|---|---|---|---|---|---|---|---|---|
| | | | | | | 64 (n=13207); ≥65 (n=12933) | | | | | SARS-CoV-2 pan-immunoglobulin immunoassay, ARCHITECT SARS-CoV-2 IgG immunoassay, VIT- ROS SARS-CoV-2 IgG immunoassay \| IgG | | |
| Bajema (CDC COVID-19 Response)[331] | 2020-09-11 to 2020-09-11 | Florida | Regional | Moderate | 979 | 0-17 (n=6612); 18-49 (n=15157); 50-64 (n=13207); ≥65 (n=12933) | NR | Non-probability | Residual sera | 8.5% (6.6-10.7%) | Ortho-Clinical Diagnostics; Abbott; Roche \| Elecsys Anti-SARS-CoV-2 pan-immunoglobulin immunoassay, ARCHITECT SARS-CoV-2 IgG immunoassay, VIT- ROS SARS-CoV-2 IgG immunoassay \| IgG | 0.0%, 0.0% (Used author-adjusted estimate) | 8.5% (6.9-10.3%) |
| Bajema (CDC COVID-19 Response)[331] | 2020-08-26 to 2020-09-10 | Delaware | Regional | Moderate | 914 | 0-17 (n=6484); 18-49 (n=14079); 50-64 (n=12426); ≥65 (n=12316) | NR | Non-probability | Residual sera | 4.5% (2.4-7.1%) | Ortho-Clinical Diagnostics; Abbott; Roche \| Elecsys Anti-SARS-CoV-2 pan-immunoglobulin immunoassay, ARCHITECT SARS-CoV-2 IgG immunoassay, VIT- ROS SARS-CoV-2 IgG immunoassay \| IgG | 0.0%, 0.0% (Used author-adjusted estimate) | 4.5% (3.3-6.0%) |

| Article Author (Study Organization) | Sampling Dates (YMD) | Location | Geographic Scope | Overall Risk of Bias | Sample Size | Age | Female (%) | Sampling method | Sample frame | Uncorrected Seroprevalence (95% Confidence Interval) | Test Manufacturers \| Names \| Isotypes | Test Sens, Spec (Source) | Corrected Seroprevalence (95% Credible Interval) |
|---|---|---|---|---|---|---|---|---|---|---|---|---|---|
| Bajema (CDC COVID-19 Response)[331] | 2020-08-25 to 2020-09-10 | Georgia | Regional | Moderate | 1020 | 0-17 (n=6484); 18-49 (n=14079); 50-64 (n=12426); ≥65 (n=12316) | NR | Non-probability | Residual sera | 8.7% (6.7-11.1%) | Ortho-Clinical Diagnostics; Abbott; Roche \| Elecsys Anti-SARS-CoV-2 pan-immunoglobulin immunoassay, ARCHITECT SARS-CoV-2 IgG immunoassay, VIT- ROS SARS-CoV-2 IgG immunoassay \| IgG | 0.0%, 0.0% (Used author-adjusted estimate) | 8.7% (7.0-10.5%) |
| Bajema (CDC COVID-19 Response)[331] | 2020-08-26 to 2020-09-10 | Idaho | Regional | Moderate | 790 | 0-17 (n=6484); 18-49 (n=14079); 50-64 (n=12426); ≥65 (n=12316) | NR | Non-probability | Residual sera | 5.7% (2.9-9.7%) | Ortho-Clinical Diagnostics; Abbott; Roche \| Elecsys Anti-SARS-CoV-2 pan-immunoglobulin immunoassay, ARCHITECT SARS-CoV-2 IgG immunoassay, VIT- ROS SARS-CoV-2 IgG immunoassay \| IgG | 0.0%, 0.0% (Used author-adjusted estimate) | 5.7% (4.2-7.5%) |
| Bajema (CDC COVID-19 Response)[331] | 2020-08-25 to 2020-09-10 | Iowa | Regional | Moderate | 1006 | 0-17 (n=6484); 18-49 (n=14079); 50-64 (n=12426); ≥65 (n=12316) | NR | Non-probability | Residual sera | 8.4% (6.2-10.7%) | Ortho-Clinical Diagnostics; Abbott; Roche \| Elecsys Anti-SARS-CoV-2 pan-immunoglobulin immunoassay, ARCHITECT SARS-CoV-2 IgG immunoassay, VIT- ROS SARS-CoV-2 IgG | 0.0%, 0.0% (Used author-adjusted estimate) | 8.4% (6.8-10.2%) |

| Article Author (Study Organization) | Sampling Dates (YMD) | Location | Geographic Scope | Overall Risk of Bias | Sample Size | Age | Female (%) | Sampling method | Sample frame | Uncorrected Seroprevalence (95% Confidence Interval) | Test Manufacturers \| Names \| Isotypes | Test Sens, Spec (Source) | Corrected Seroprevalence (95% Credible Interval) |
|---|---|---|---|---|---|---|---|---|---|---|---|---|---|
| | | | | | | | | | | | immunoassay \| IgG | | |
| Bajema (CDC COVID-19 Response)[331] | 2020-08-26 to 2020-09-10 | Maine | Regional | Moderate | 759 | 0-17 (n=6484); 18-49 (n=14079); 50-64 (n=12426); ≥65 (n=12316) | NR | Non-probability | Residual sera | 1.5% (0.4-3.2%) | Ortho-Clinical Diagnostics; Abbott; Roche \| Elecsys Anti-SARS-CoV-2 pan-immunoglobulin immunoassay, ARCHITECT SARS-CoV-2 IgG immunoassay, VIT- ROS SARS-CoV-2 IgG immunoassay \| IgG | 0.0%, 0.0% (Used author-adjusted estimate) | 1.5% (0.8-2.5%) |
| Bajema (CDC COVID-19 Response)[331] | 2020-08-26 to 2020-09-10 | Mississippi | Regional | Moderate | 987 | 0-17 (n=6484); 18-49 (n=14079); 50-64 (n=12426); ≥65 (n=12316) | NR | Non-probability | Residual sera | 8.4% (4.9-12.7%) | Ortho-Clinical Diagnostics; Abbott; Roche \| Elecsys Anti-SARS-CoV-2 pan-immunoglobulin immunoassay, ARCHITECT SARS-CoV-2 IgG immunoassay, VIT- ROS SARS-CoV-2 IgG immunoassay \| IgG | 0.0%, 0.0% (Used author-adjusted estimate) | 8.4% (6.7-10.2%) |
| Bajema (CDC COVID-19 Response)[331] | 2020-08-24 to 2020-09-10 | Missouri | Regional | Moderate | 1024 | 0-17 (n=6484); 18-49 (n=14079); 50-64 (n=12426); ≥65 (n=12316) | NR | Non-probability | Residual sera | 2.9% (1.7-4.2%) | Ortho-Clinical Diagnostics; Abbott; Roche \| Elecsys Anti-SARS-CoV-2 pan-immunoglobulin immunoassay, ARCHITECT SARS-CoV-2 IgG | 0.0%, 0.0% (Used author-adjusted estimate) | 2.9% (1.9-4.0%) |

| Article Author (Study Organization) | Sampling Dates (YMD) | Location | Geographic Scope | Overall Risk of Bias | Sample Size | Age | Female (%) | Sampling method | Sample frame | Uncorrected Seroprevalence (95% Confidence Interval) | Test Manufacturers \| Names \| Isotypes | Test Sens, Spec (Source) | Corrected Seroprevalence (95% Credible Interval) |
|---|---|---|---|---|---|---|---|---|---|---|---|---|---|
| | | | | | | | | | | | immunoassay, VIT- ROS SARS-CoV-2 IgG immunoassay \| IgG | | |
| Bajema (CDC COVID-19 Response)[331] | 2020-08-24 to 2020-09-10 | Nebraska | Regional | Moderate | 938 | 0-17 (n=6484); 18-49 (n=14079); 50-64 (n=12426); ≥65 (n=12316) | NR | Non-probability | Residual sera | 6.3% (4.5-8.3%) | Ortho-Clinical Diagnostics; Abbott; Roche \| Elecsys Anti-SARS-CoV-2 pan-immunoglobulin immunoassay, ARCHITECT SARS-CoV-2 IgG immunoassay, VIT- ROS SARS-CoV-2 IgG immunoassay \| IgG | 0.0%, 0.0% (Used author-adjusted estimate) | 6.3% (4.9-8.0%) |
| Bajema (CDC COVID-19 Response)[331] | 2020-08-26 to 2020-09-10 | New York | Regional | Moderate | 1043 | 0-17 (n=6484); 18-49 (n=14079); 50-64 (n=12426); ≥65 (n=12316) | NR | Non-probability | Residual sera | 19.5% (16.9-22.4%) | Ortho-Clinical Diagnostics; Abbott; Roche \| Elecsys Anti-SARS-CoV-2 pan-immunoglobulin immunoassay, ARCHITECT SARS-CoV-2 IgG immunoassay, VIT- ROS SARS-CoV-2 IgG immunoassay \| IgG | 0.0%, 0.0% (Used author-adjusted estimate) | 19.5% (17.1-21.9%) |
| Bajema (CDC COVID-19 Response)[331] | 2020-08-27 to 2020-09-10 | Ohio | Regional | Moderate | 983 | 0-17 (n=6484); 18-49 (n=14079); 50-64 (n=12426); ≥65 (n=12316) | NR | Non-probability | Residual sera | 5.0% (3.2-7.1%) | Ortho-Clinical Diagnostics; Abbott; Roche \| Elecsys Anti-SARS-CoV-2 pan-immunoglobulin | 0.0%, 0.0% (Used author-adjusted estimate) | 5.0% (3.8-6.5%) |

| Article Author (Study Organization) | Sampling Dates (YMD) | Location | Geographic Scope | Overall Risk of Bias | Sample Size | Age | Female (%) | Sampling method | Sample frame | Uncorrected Seroprevalence (95% Confidence Interval) | Test Manufacturers \| Names \| Isotypes | Test Sens, Spec (Source) | Corrected Seroprevalence (95% Credible Interval) |
|---|---|---|---|---|---|---|---|---|---|---|---|---|---|
| | | | | | | | | | | | immunoassay, ARCHITECT SARS-CoV-2 IgG immunoassay, VIT- ROS SARS-CoV-2 IgG immunoassay \| IgG | | |
| Bajema (CDC COVID-19 Response)[331] | 2020-08-26 to 2020-09-10 | Oregon | Regional | Moderate | 968 | 0-17 (n=6484); 18-49 (n=14079); 50-64 (n=12426); ≥65 (n=12316) | NR | Non-probability | Residual sera | 1.9% (0.9-2.9%) | Ortho-Clinical Diagnostics; Abbott; Roche \| Elecsys Anti-SARS-CoV-2 pan-immunoglobulin immunoassay, ARCHITECT SARS-CoV-2 IgG immunoassay, VIT- ROS SARS-CoV-2 IgG immunoassay \| IgG | 0.0%, 0.0% (Used author-adjusted estimate) | 1.9% (1.1-2.9%) |
| Bajema (CDC COVID-19 Response)[331] | 2020-08-27 to 2020-09-10 | Rhode Island | Regional | Moderate | 786 | 0-17 (n=6484); 18-49 (n=14079); 50-64 (n=12426); ≥65 (n=12316) | NR | Non-probability | Residual sera | 3.1% (1.7-4.6%) | Ortho-Clinical Diagnostics; Abbott; Roche \| Elecsys Anti-SARS-CoV-2 pan-immunoglobulin immunoassay, ARCHITECT SARS-CoV-2 IgG immunoassay, VIT- ROS SARS-CoV-2 IgG immunoassay \| IgG | 0.0%, 0.0% (Used author-adjusted estimate) | 3.1% (2.0-4.4%) |
| Bajema (CDC COVID-19 Response)[331] | 2020-08-26 to 2020-09-10 | South Carolina | Regional | Moderate | 927 | 0-17 (n=6484); 18-49 (n=14079); 50- | NR | Non-probability | Residual sera | 7.2% (5.1-9.6%) | Ortho-Clinical Diagnostics; Abbott; Roche \| Elecsys Anti- | 0.0%, 0.0% (Used author-adjusted estimate) | 7.2% (5.6-8.9%) |

| Article Author (Study Organization) | Sampling Dates (YMD) | Location | Geographic Scope | Overall Risk of Bias | Sample Size | Age | Female (%) | Sampling method | Sample frame | Uncorrected Seroprevalence (95% Confidence Interval) | Test Manufacturers \| Names \| Isotypes | Test Sens, Spec (Source) | Corrected Seroprevalence (95% Credible Interval) |
|---|---|---|---|---|---|---|---|---|---|---|---|---|---|
| | | | | | | 64 (n=12426); ≥65 (n=12316) | | | | | SARS-CoV-2 pan-immunoglobulin immunoassay, ARCHITECT SARS-CoV-2 IgG immunoassay, VIT- ROS SARS-CoV-2 IgG immunoassay \| IgG | | |
| Bajema (CDC COVID-19 Response)[331] | 2020-08-26 to 2020-09-10 | South Dakota | Regional | High | 97 | 0-17 (n=6484); 18-49 (n=14079); 50-64 (n=12426); ≥65 (n=12316) | NR | Non-probability | Residual sera | 0.7% (0.0-2.5%) | Ortho-Clinical Diagnostics; Abbott; Roche \| Elecsys Anti-SARS-CoV-2 pan-immunoglobulin immunoassay, ARCHITECT SARS-CoV-2 IgG immunoassay, VIT- ROS SARS-CoV-2 IgG immunoassay \| IgG | 0.0%, 0.0% (Used author-adjusted estimate) | 0.7% (0.0-2.5%) |
| Bajema (CDC COVID-19 Response)[331] | 2020-08-26 to 2020-09-10 | Vermont | Regional | High | 485 | 0-17 (n=6484); 18-49 (n=14079); 50-64 (n=12426); ≥65 (n=12316) | NR | Non-probability | Residual sera | 0.4% (0.0-0.9%) | Ortho-Clinical Diagnostics; Abbott; Roche \| Elecsys Anti-SARS-CoV-2 pan-immunoglobulin immunoassay, ARCHITECT SARS-CoV-2 IgG immunoassay, VIT- ROS SARS-CoV-2 IgG immunoassay \| IgG | 0.0%, 0.0% (Used author-adjusted estimate) | 0.4% (0.0-1.0%) |

| Article Author (Study Organization) | Sampling Dates (YMD) | Location | Geographic Scope | Overall Risk of Bias | Sample Size | Age | Female (%) | Sampling method | Sample frame | Uncorrected Seroprevalence (95% Confidence Interval) | Test Manufacturers \| Names \| Isotypes | Test Sens, Spec (Source) | Corrected Seroprevalence (95% Credible Interval) |
|---|---|---|---|---|---|---|---|---|---|---|---|---|---|
| Bajema (CDC COVID-19 Response)[331] | 2020-08-25 to 2020-09-10 | Washington | Regional | Moderate | 820 | 0-17 (n=6484); 18-49 (n=14079); 50-64 (n=12426); ≥65 (n=12316) | NR | Non-probability | Residual sera | 4.5% (2.6-6.9%) | Ortho-Clinical Diagnostics; Abbott; Roche \| Elecsys Anti-SARS-CoV-2 pan-immunoglobulin immunoassay, ARCHITECT SARS-CoV-2 IgG immunoassay, VIT- ROS SARS-CoV-2 IgG immunoassay \| IgG | 0.0%, 0.0% (Used author-adjusted estimate) | 4.5% (3.1-6.0%) |
| Bajema (CDC COVID-19 Response)[331] | 2020-08-27 to 2020-09-10 | Washington D.C | Regional | Moderate | 643 | 0-17 (n=6484); 18-49 (n=14079); 50-64 (n=12426); ≥65 (n=12316) | NR | Non-probability | Residual sera | 5.0% (2.8-7.5%) | Ortho-Clinical Diagnostics; Abbott; Roche \| Elecsys Anti-SARS-CoV-2 pan-immunoglobulin immunoassay, ARCHITECT SARS-CoV-2 IgG immunoassay, VIT- ROS SARS-CoV-2 IgG immunoassay \| IgG | 0.0%, 0.0% (Used author-adjusted estimate) | 5.0% (3.5-6.9%) |
| Bajema (CDC COVID-19 Response)[331] | 2020-08-26 to 2020-09-10 | West Virginia | Regional | Moderate | 1128 | 0-17 (n=6484); 18-49 (n=14079); 50-64 (n=12426); ≥65 (n=12316) | NR | Non-probability | Residual sera | 1.5% (0.8-2.4%) | Ortho-Clinical Diagnostics; Abbott; Roche \| Elecsys Anti-SARS-CoV-2 pan-immunoglobulin immunoassay, ARCHITECT SARS-CoV-2 IgG immunoassay, VIT- ROS SARS-CoV-2 IgG | 0.0%, 0.0% (Used author-adjusted estimate) | 1.5% (0.8-2.2%) |

| Article Author (Study Organization) | Sampling Dates (YMD) | Location | Geographic Scope | Overall Risk of Bias | Sample Size | Age | Female (%) | Sampling method | Sample frame | Uncorrected Seroprevalence (95% Confidence Interval) | Test Manufacturers \| Names \| Isotypes | Test Sens, Spec (Source) | Corrected Seroprevalence (95% Credible Interval) |
|---|---|---|---|---|---|---|---|---|---|---|---|---|---|
| | | | | | | | | | | | immunoassay \| IgG | | |
| Bajema (CDC COVID-19 Response)[331] | 2020-08-25 to 2020-09-10 | Wisconsi n | Regional | Moderate | 976 | 0-17 (n=6484); 18-49 (n=14079); 50-64 (n=12426); ≥65 (n=12316) | NR | Non-probability | Residual sera | 2.6% (1.5-4.0%) | Ortho-Clinical Diagnostics; Abbott; Roche \| Elecsys Anti-SARS-CoV-2 pan-immunoglobuli n immunoassay, ARCHITECT SARS-CoV-2 IgG immunoassay, VIT- ROS SARS-CoV-2 IgG immunoassay \| IgG | 0.0%, 0.0% (Used author-adjusted estimate) | 2.6% (1.7-3.7%) |
| Bajema (CDC COVID-19 Response)[331] | 2020-08-26 to 2020-09-09 | Alaska | Regional | Moderate | 610 | 0-17 (n=6484); 18-49 (n=14079); 50-64 (n=12426); ≥65 (n=12316) | NR | Non-probability | Residual sera | 1.0% (0.2-2.2%) | Ortho-Clinical Diagnostics; Abbott; Roche \| Elecsys Anti-SARS-CoV-2 pan-immunoglobuli n immunoassay, ARCHITECT SARS-CoV-2 IgG immunoassay, VIT- ROS SARS-CoV-2 IgG immunoassay \| IgG | 0.0%, 0.0% (Used author-adjusted estimate) | 1.0% (0.4-2.0%) |
| Bajema (CDC COVID-19 Response)[331] | 2020-08-26 to 2020-09-09 | Arizona | Regional | Moderate | 1030 | 0-17 (n=6484); 18-49 (n=14079); 50-64 (n=12426); ≥65 (n=12316) | NR | Non-probability | Residual sera | 4.9% (3.0-7.1%) | Ortho-Clinical Diagnostics; Abbott; Roche \| Elecsys Anti-SARS-CoV-2 pan-immunoglobuli n immunoassay, ARCHITECT SARS-CoV-2 IgG | 0.0%, 0.0% (Used author-adjusted estimate) | 4.9% (3.7-6.3%) |

| Article Author (Study Organization) | Sampling Dates (YMD) | Location | Geographic Scope | Overall Risk of Bias | Sample Size | Age | Female (%) | Sampling method | Sample frame | Uncorrected Seroprevalence (95% Confidence Interval) | Test Manufacturers \| Names \| Isotypes | Test Sens, Spec (Source) | Corrected Seroprevalence (95% Credible Interval) |
|---|---|---|---|---|---|---|---|---|---|---|---|---|---|
| | | | | | | | | | | | immunoassay, VIT- ROS SARS-CoV-2 IgG immunoassay \| IgG | | |
| Bajema (CDC COVID-19 Response)[331] | 2020-08-28 to 2020-09-09 | California | Regional | Moderate | 983 | 0-17 (n=6484); 18-49 (n=14079); 50-64 (n=12426); ≥65 (n=12316) | NR | Non-probability | Residual sera | 6.0% (4.2-7.8%) | Ortho-Clinical Diagnostics; Abbott; Roche \| Elecsys Anti-SARS-CoV-2 pan-immunoglobulin immunoassay, ARCHITECT SARS-CoV-2 IgG immunoassay, VIT- ROS SARS-CoV-2 IgG immunoassay \| IgG | 0.0%, 0.0% (Used author-adjusted estimate) | 6.0% (4.6-7.5%) |
| Bajema (CDC COVID-19 Response)[331] | 2020-08-27 to 2020-09-09 | Florida | Regional | Moderate | 981 | 0-17 (n=6484); 18-49 (n=14079); 50-64 (n=12426); ≥65 (n=12316) | NR | Non-probability | Residual sera | 5.7% (3.9-7.5%) | Ortho-Clinical Diagnostics; Abbott; Roche \| Elecsys Anti-SARS-CoV-2 pan-immunoglobulin immunoassay, ARCHITECT SARS-CoV-2 IgG immunoassay, VIT- ROS SARS-CoV-2 IgG immunoassay \| IgG | 0.0%, 0.0% (Used author-adjusted estimate) | 5.7% (4.3-7.2%) |
| Bajema (CDC COVID-19 Response)[331] | 2020-08-26 to 2020-09-09 | Indiana | Regional | Moderate | 984 | 0-17 (n=6484); 18-49 (n=14079); 50-64 (n=12426); ≥65 (n=12316) | NR | Non-probability | Residual sera | 2.4% (1.4-3.5%) | Ortho-Clinical Diagnostics; Abbott; Roche \| Elecsys Anti-SARS-CoV-2 pan-immunoglobulin | 0.0%, 0.0% (Used author-adjusted estimate) | 2.4% (1.5-3.4%) |

| Article Author (Study Organization) | Sampling Dates (YMD) | Location | Geographic Scope | Overall Risk of Bias | Sample Size | Age | Female (%) | Sampling method | Sample frame | Uncorrected Seroprevalence (95% Confidence Interval) | Test Manufacturers \| Names \| Isotypes | Test Sens, Spec (Source) | Corrected Seroprevalence (95% Credible Interval) |
|---|---|---|---|---|---|---|---|---|---|---|---|---|---|
| | | | | | | | | | | | immunoassay, ARCHITECT SARS-CoV-2 IgG immunoassay, VIT- ROS SARS-CoV-2 IgG immunoassay \| IgG | | |
| Bajema (CDC COVID-19 Response)[331] | 2020-08-24 to 2020-09-09 | Montana | Regional | High | 518 | 0-17 (n=6484); 18-49 (n=14079); 50-64 (n=12426); ≥65 (n=12316) | NR | Non-probability | Residual sera | 0.9% (0.2-1.8%) | Ortho-Clinical Diagnostics; Abbott; Roche \| Elecsys Anti-SARS-CoV-2 pan-immunoglobulin immunoassay, ARCHITECT SARS-CoV-2 IgG immunoassay, VIT- ROS SARS-CoV-2 IgG immunoassay \| IgG | 0.0%, 0.0% (Used author-adjusted estimate) | 0.9% (0.3-1.8%) |
| Bajema (CDC COVID-19 Response)[331] | 2020-08-26 to 2020-09-09 | North Carolina | Regional | Moderate | 982 | 0-17 (n=6484); 18-49 (n=14079); 50-64 (n=12426); ≥65 (n=12316) | NR | Non-probability | Residual sera | 3.8% (2.3-5.3%) | Ortho-Clinical Diagnostics; Abbott; Roche \| Elecsys Anti-SARS-CoV-2 pan-immunoglobulin immunoassay, ARCHITECT SARS-CoV-2 IgG immunoassay, VIT- ROS SARS-CoV-2 IgG immunoassay \| IgG | 0.0%, 0.0% (Used author-adjusted estimate) | 3.8% (2.7-5.1%) |
| Bajema (CDC COVID-19 Response)[331] | 2020-08-26 to 2020-09-09 | North Dakota | Regional | High | 109 | 0-17 (n=6484); 18-49 (n=14079); 50- | NR | Non-probability | Residual sera | 0.2% (0.0-1.0%) | Ortho-Clinical Diagnostics; Abbott; Roche \| Elecsys Anti- | 0.0%, 0.0% (Used author-adjusted estimate) | 0.2% (0.0-2.3%) |

| Article Author (Study Organization) | Sampling Dates (YMD) | Location | Geographic Scope | Overall Risk of Bias | Sample Size | Age | Female (%) | Sampling method | Sample frame | Uncorrected Seroprevalence (95% Confidence Interval) | Test Manufacturers \| Names \| Isotypes | Test Sens, Spec (Source) | Corrected Seroprevalence (95% Credible Interval) |
|---|---|---|---|---|---|---|---|---|---|---|---|---|---|
| | | | | | | 64 (n=12426); ≥65 (n=12316) | | | | | SARS-CoV-2 pan-immunoglobulin immunoassay, ARCHITECT SARS-CoV-2 IgG immunoassay, VIT- ROS SARS-CoV-2 IgG immunoassay \| IgG | | |
| Bajema (CDC COVID-19 Response)[331] | 2020-08-26 to 2020-09-09 | Pennsylvania | Regional | Moderate | 845 | 0-17 (n=6484); 18-49 (n=14079); 50-64 (n=12426); ≥65 (n=12316) | NR | Non-probability | Residual sera | 9.5% (6.6-13.6%) | Ortho-Clinical Diagnostics; Abbott; Roche \| Elecsys Anti-SARS-CoV-2 pan-immunoglobulin immunoassay, ARCHITECT SARS-CoV-2 IgG immunoassay, VIT- ROS SARS-CoV-2 IgG immunoassay \| IgG | 0.0%, 0.0% (Used author-adjusted estimate) | 9.5% (7.6-11.6%) |
| Bajema (CDC COVID-19 Response)[331] | 2020-08-26 to 2020-09-08 | Alabama | Regional | Moderate | 955 | 0-17 (n=6484); 18-49 (n=14079); 50-64 (n=12426); ≥65 (n=12316) | NR | Non-probability | Residual sera | 9.9% (7.2-12.8%) | Ortho-Clinical Diagnostics; Abbott; Roche \| Elecsys Anti-SARS-CoV-2 pan-immunoglobulin immunoassay, ARCHITECT SARS-CoV-2 IgG immunoassay, VIT- ROS SARS-CoV-2 IgG immunoassay \| IgG | 0.0%, 0.0% (Used author-adjusted estimate) | 9.9% (8.1-11.9%) |

| Article Author (Study Organization) | Sampling Dates (YMD) | Location | Geographic Scope | Overall Risk of Bias | Sample Size | Age | Female (%) | Sampling method | Sample frame | Uncorrected Seroprevalence (95% Confidence Interval) | Test Manufacturers \| Names \| Isotypes | Test Sens, Spec (Source) | Corrected Seroprevalence (95% Credible Interval) |
|---|---|---|---|---|---|---|---|---|---|---|---|---|---|
| Bajema (CDC COVID-19 Response)[331] | 2020-08-24 to 2020-09-08 | Arkansas | Regional | Moderate | 992 | 0-17 (n=6484); 18-49 (n=14079); 50-64 (n=12426); ≥65 (n=12316) | NR | Non-probability | Residual sera | 4.9% (3.5-6.4%) | Ortho-Clinical Diagnostics; Abbott; Roche \| Elecsys Anti-SARS-CoV-2 pan-immunoglobulin immunoassay, ARCHITECT SARS-CoV-2 IgG immunoassay, VIT- ROS SARS-CoV-2 IgG immunoassay \| IgG | 0.0%, 0.0% (Used author-adjusted estimate) | 4.9% (3.6-6.3%) |
| Bajema (CDC COVID-19 Response)[331] | 2020-08-26 to 2020-09-08 | Kentucky | Regional | Moderate | 983 | 0-17 (n=6484); 18-49 (n=14079); 50-64 (n=12426); ≥65 (n=12316) | NR | Non-probability | Residual sera | 3.1% (1.9-4.3%) | Ortho-Clinical Diagnostics; Abbott; Roche \| Elecsys Anti-SARS-CoV-2 pan-immunoglobulin immunoassay, ARCHITECT SARS-CoV-2 IgG immunoassay, VIT- ROS SARS-CoV-2 IgG immunoassay \| IgG | 0.0%, 0.0% (Used author-adjusted estimate) | 3.1% (2.1-4.3%) |
| Bajema (CDC COVID-19 Response)[331] | 2020-08-26 to 2020-09-08 | Louisiana | Regional | Moderate | 1001 | 0-17 (n=6484); 18-49 (n=14079); 50-64 (n=12426); ≥65 (n=12316) | NR | Non-probability | Residual sera | 8.6% (6.7-11.3%) | Ortho-Clinical Diagnostics; Abbott; Roche \| Elecsys Anti-SARS-CoV-2 pan-immunoglobulin immunoassay, ARCHITECT SARS-CoV-2 IgG immunoassay, VIT- ROS SARS-CoV-2 IgG | 0.0%, 0.0% (Used author-adjusted estimate) | 8.6% (7.0-10.4%) |

| Article Author (Study Organization) | Sampling Dates (YMD) | Location | Geographic Scope | Overall Risk of Bias | Sample Size | Age | Female (%) | Sampling method | Sample frame | Uncorrected Seroprevalence (95% Confidence Interval) | Test Manufacturers \| Names \| Isotypes | Test Sens, Spec (Source) | Corrected Seroprevalence (95% Credible Interval) |
|---|---|---|---|---|---|---|---|---|---|---|---|---|---|
| | | | | | | | | | | | immunoassay \| IgG | | |
| Bajema (CDC COVID-19 Response)[331] | 2020-08-26 to 2020-09-08 | Maryland | Regional | Moderate | 980 | 0-17 (n=6484); 18-49 (n=14079); 50-64 (n=12426); ≥65 (n=12316) | NR | Non-probability | Residual sera | 7.9% (5.7-10.3%) | Ortho-Clinical Diagnostics; Abbott; Roche \| Elecsys Anti-SARS-CoV-2 pan-immunoglobulin immunoassay, ARCHITECT SARS-CoV-2 IgG immunoassay, VIT- ROS SARS-CoV-2 IgG immunoassay \| IgG | 0.0%, 0.0% (Used author-adjusted estimate) | 7.9% (6.3-9.7%) |
| Bajema (CDC COVID-19 Response)[331] | 2020-08-26 to 2020-09-08 | Minnesota | Regional | Moderate | 974 | 0-17 (n=6484); 18-49 (n=14079); 50-64 (n=12426); ≥65 (n=12316) | NR | Non-probability | Residual sera | 9.2% (6.7-11.9%) | Ortho-Clinical Diagnostics; Abbott; Roche \| Elecsys Anti-SARS-CoV-2 pan-immunoglobulin immunoassay, ARCHITECT SARS-CoV-2 IgG immunoassay, VIT- ROS SARS-CoV-2 IgG immunoassay \| IgG | 0.0%, 0.0% (Used author-adjusted estimate) | 9.2% (7.4-11.1%) |
| Bajema (CDC COVID-19 Response)[331] | 2020-08-25 to 2020-09-08 | New Mexico | Regional | Moderate | 985 | 0-17 (n=6484); 18-49 (n=14079); 50-64 (n=12426); ≥65 (n=12316) | NR | Non-probability | Residual sera | 3.7% (2.3-5.5%) | Ortho-Clinical Diagnostics; Abbott; Roche \| Elecsys Anti-SARS-CoV-2 pan-immunoglobulin immunoassay, ARCHITECT SARS-CoV-2 IgG | 0.0%, 0.0% (Used author-adjusted estimate) | 3.7% (2.6-5.0%) |

| Article Author (Study Organization) | Sampling Dates (YMD) | Location | Geographic Scope | Overall Risk of Bias | Sample Size | Age | Female (%) | Sampling method | Sample frame | Uncorrected Seroprevalence (95% Confidence Interval) | Test Manufacturers \| Names \| Isotypes | Test Sens, Spec (Source) | Corrected Seroprevalence (95% Credible Interval) |
|---|---|---|---|---|---|---|---|---|---|---|---|---|---|
| | | | | | | | | | | | immunoassay, VIT- ROS SARS-CoV-2 IgG immunoassay \| IgG | | |
| Bajema (CDC COVID-19 Response)[331] | 2020-08-25 to 2020-09-08 | Tennesse e | Regional | Moderate | 950 | 0-17 (n=6484); 18-49 (n=14079); 50-64 (n=12426); ≥65 (n=12316) | NR | Non-probability | Residual sera | 5.4% (3.6-7.3%) | Ortho-Clinical Diagnostics; Abbott; Roche \| Elecsys Anti-SARS-CoV-2 pan-immunoglobuli n immunoassay, ARCHITECT SARS-CoV-2 IgG immunoassay, VIT- ROS SARS-CoV-2 IgG immunoassay \| IgG | 0.0%, 0.0% (Used author-adjusted estimate) | 5.4% (4.1-6.9%) |
| Bajema (CDC COVID-19 Response)[331] | 2020-08-24 to 2020-09-08 | Utah | Regional | Moderate | 1104 | 0-17 (n=6484); 18-49 (n=14079); 50-64 (n=12426); ≥65 (n=12316) | NR | Non-probability | Residual sera | 4.9% (2.8-7.7%) | Ortho-Clinical Diagnostics; Abbott; Roche \| Elecsys Anti-SARS-CoV-2 pan-immunoglobuli n immunoassay, ARCHITECT SARS-CoV-2 IgG immunoassay, VIT- ROS SARS-CoV-2 IgG immunoassay \| IgG | 0.0%, 0.0% (Used author-adjusted estimate) | 4.9% (3.7-6.3%) |
| Bajema (CDC COVID-19 Response)[331] | 2020-08-26 to 2020-09-08 | Virginia | Regional | Moderate | 1007 | 0-17 (n=6484); 18-49 (n=14079); 50-64 (n=12426); ≥65 (n=12316) | NR | Non-probability | Residual sera | 3.5% (2.0-5.3%) | Ortho-Clinical Diagnostics; Abbott; Roche \| Elecsys Anti-SARS-CoV-2 pan-immunoglobuli n | 0.0%, 0.0% (Used author-adjusted estimate) | 3.5% (2.5-4.7%) |

| Article Author (Study Organization) | Sampling Dates (YMD) | Location | Geographic Scope | Overall Risk of Bias | Sample Size | Age | Female (%) | Sampling method | Sample frame | Uncorrected Seroprevalence (95% Confidence Interval) | Test Manufacturers \| Names \| Isotypes | Test Sens, Spec (Source) | Corrected Seroprevalence (95% Credible Interval) |
|---|---|---|---|---|---|---|---|---|---|---|---|---|---|
| | | | | | | | | | | | immunoassay, ARCHITECT SARS-CoV-2 IgG immunoassay, VIT- ROS SARS-CoV-2 IgG immunoassay \| IgG | | |
| Bajema (CDC COVID-19 Response)[331] | 2020-08-26 to 2020-09-07 | New Jersey | Regional | Moderate | 944 | 0-17 (n=6484); 18-49 (n=14079); 50-64 (n=12426); ≥65 (n=12316) | NR | Non-probability | Residual sera | 12.8% (10.5-15.3%) | Ortho-Clinical Diagnostics; Abbott; Roche \| Elecsys Anti-SARS-CoV-2 pan-immunoglobulin immunoassay, ARCHITECT SARS-CoV-2 IgG immunoassay, VIT- ROS SARS-CoV-2 IgG immunoassay \| IgG | 0.0%, 0.0% (Used author-adjusted estimate) | 12.8% (10.7-15.0%) |
| Demonbreun (Northwestern University)[334] | 2020-06-24 to 2020-09-06 | Illinois \| Chicago | Local | High | 1545 | Median 37 (IQR 18) | 59.3 | Non-probability | Household and community samples | 19.8% | In-House \| NR \| IgG | 75.6%, 97.6% (Used Bastos SR/MA data; no sens, spec, or author adjustment available) | 23.6% (20.6-26.6%) |
| Bajema (CDC COVID-19 Response)[331] | 2020-08-27 to 2020-09-05 | Massachusetts | Regional | Moderate | 981 | 0-17 (n=6484); 18-49 (n=14079); 50-64 (n=12426); ≥65 (n=12316) | NR | Non-probability | Residual sera | 3.6% (2.4-5.1%) | Ortho-Clinical Diagnostics; Abbott; Roche \| Elecsys Anti-SARS-CoV-2 pan-immunoglobulin immunoassay, ARCHITECT SARS-CoV-2 IgG immunoassay, VIT- ROS SARS-CoV-2 | 0.0%, 0.0% (Used author-adjusted estimate) | 3.6% (2.5-4.9%) |

| Article Author (Study Organization) | Sampling Dates (YMD) | Location | Geographic Scope | Overall Risk of Bias | Sample Size | Age | Female (%) | Sampling method | Sample frame | Uncorrected Seroprevalence (95% Confidence Interval) | Test Manufacturers \| Names \| Isotypes | Test Sens, Spec (Source) | Corrected Seroprevalence (95% Credible Interval) |
|---|---|---|---|---|---|---|---|---|---|---|---|---|---|
| | | | | | | | | | | | IgG immunoassay \| IgG | | |
| Bajema (CDC COVID-19 Response)[331] | 2020-08-24 to 2020-09-04 | Colorado | Regional | Moderate | 1025 | 0-17 (n=6484); 18-49 (n=14079); 50-64 (n=12426); ≥65 (n=12316) | NR | Non-probability | Residual sera | 3.6% (2.3-5.1%) | Ortho-Clinical Diagnostics; Abbott; Roche \| Elecsys Anti-SARS-CoV-2 pan-immunoglobulin immunoassay, ARCHITECT SARS-CoV-2 IgG immunoassay, VIT- ROS SARS-CoV-2 IgG immunoassay \| IgG | 0.0%, 0.0% (Used author-adjusted estimate) | 3.6% (2.5-4.8%) |
| Bajema (CDC COVID-19 Response)[331] | 2020-08-26 to 2020-09-04 | Connecticut | Regional | Moderate | 983 | 0-17 (n=6484); 18-49 (n=14079); 50-64 (n=12426); ≥65 (n=12316) | NR | Non-probability | Residual sera | 4.3% (2.9-5.7%) | Ortho-Clinical Diagnostics; Abbott; Roche \| Elecsys Anti-SARS-CoV-2 pan-immunoglobulin immunoassay, ARCHITECT SARS-CoV-2 IgG immunoassay, VIT- ROS SARS-CoV-2 IgG immunoassay \| IgG | 0.0%, 0.0% (Used author-adjusted estimate) | 4.3% (3.1-5.7%) |
| Bajema (CDC COVID-19 Response)[331] | 2020-08-26 to 2020-09-03 | Illinois | Regional | Moderate | 979 | 0-17 (n=6484); 18-49 (n=14079); 50-64 (n=12426); ≥65 (n=12316) | NR | Non-probability | Residual sera | 5.6% (3.9-7.7%) | Ortho-Clinical Diagnostics; Abbott; Roche \| Elecsys Anti-SARS-CoV-2 pan-immunoglobulin immunoassay, ARCHITECT SARS-CoV-2 | 0.0%, 0.0% (Used author-adjusted estimate) | 5.6% (4.2-7.1%) |

| Article Author (Study Organization) | Sampling Dates (YMD) | Location | Geographic Scope | Overall Risk of Bias | Sample Size | Age | Female (%) | Sampling method | Sample frame | Uncorrected Seroprevalence (95% Confidence Interval) | Test Manufacturers \| Names \| Isotypes | Test Sens, Spec (Source) | Corrected Seroprevalence (95% Credible Interval) |
|---|---|---|---|---|---|---|---|---|---|---|---|---|---|
| | | | | | | | | | | | IgG immunoassay, VIT- ROS SARS-CoV-2 IgG immunoassay \| IgG | | |
| Bajema (CDC COVID-19 Response)[331] | 2020-08-24 to 2020-09-03 | Kansas | Regional | Moderate | 1025 | 0-17 (n=6484); 18-49 (n=14079); 50-64 (n=12426); ≥65 (n=12316) | NR | Non-probability | Residual sera | 2.9% (1.6-4.3%) | Ortho-Clinical Diagnostics; Abbott; Roche \| Elecsys Anti-SARS-CoV-2 pan-immunoglobulin immunoassay, ARCHITECT SARS-CoV-2 IgG immunoassay, VIT- ROS SARS-CoV-2 IgG immunoassay \| IgG | 0.0%, 0.0% (Used author-adjusted estimate) | 2.9% (1.9-4.0%) |
| Bajema (CDC COVID-19 Response)[331] | 2020-08-26 to 2020-09-03 | Michigan | Regional | Moderate | 976 | 0-17 (n=6484); 18-49 (n=14079); 50-64 (n=12426); ≥65 (n=12316) | NR | Non-probability | Residual sera | 3.4% (2.2-5.0%) | Ortho-Clinical Diagnostics; Abbott; Roche \| Elecsys Anti-SARS-CoV-2 pan-immunoglobulin immunoassay, ARCHITECT SARS-CoV-2 IgG immunoassay, VIT- ROS SARS-CoV-2 IgG immunoassay \| IgG | 0.0%, 0.0% (Used author-adjusted estimate) | 3.4% (2.4-4.7%) |
| Bajema (CDC COVID-19 Response)[331] | 2020-08-26 to 2020-09-03 | New Hampshire | Regional | Moderate | 1147 | 0-17 (n=6484); 18-49 (n=14079); 50-64 (n=12426); ≥65 (n=12316) | NR | Non-probability | Residual sera | 1.6% (0.7-2.8%) | Ortho-Clinical Diagnostics; Abbott; Roche \| Elecsys Anti-SARS-CoV-2 pan-immunoglobuli | 0.0%, 0.0% (Used author-adjusted estimate) | 1.6% (1.0-2.4%) |

| Article Author (Study Organization) | Sampling Dates (YMD) | Location | Geographic Scope | Overall Risk of Bias | Sample Size | Age | Female (%) | Sampling method | Sample frame | Uncorrected Seroprevalence (95% Confidence Interval) | Test Manufacturers \| Names \| Isotypes | Test Sens, Spec (Source) | Corrected Seroprevalence (95% Credible Interval) |
|---|---|---|---|---|---|---|---|---|---|---|---|---|---|
| | | | | | | | | | | | n immunoassay, ARCHITECT SARS-CoV-2 IgG immunoassay, VIT- ROS SARS-CoV-2 IgG immunoassay \| IgG | | |
| Bajema (CDC COVID-19 Response)[331] | 2020-08-24 to 2020-09-03 | Puerto Rico | Regional | Moderate | 980 | 0-17 (n=6484); 18-49 (n=14079); 50-64 (n=12426); ≥65 (n=12316) | NR | Non-probability | Residual sera | 2.5% (1.4-3.8%) | Ortho-Clinical Diagnostics; Abbott; Roche \| Elecsys Anti-SARS-CoV-2 pan-immunoglobulin immunoassay, ARCHITECT SARS-CoV-2 IgG immunoassay, VIT- ROS SARS-CoV-2 IgG immunoassay \| IgG | 0.0%, 0.0% (Used author-adjusted estimate) | 2.5% (1.6-3.6%) |
| Bajema (CDC COVID-19 Response)[331] | 2020-08-25 to 2020-09-02 | Texas | Regional | Moderate | 979 | 0-17 (n=6484); 18-49 (n=14079); 50-64 (n=12426); ≥65 (n=12316) | NR | Non-probability | Residual sera | 5.8% (4.0-7.9%) | Ortho-Clinical Diagnostics; Abbott; Roche \| Elecsys Anti-SARS-CoV-2 pan-immunoglobulin immunoassay, ARCHITECT SARS-CoV-2 IgG immunoassay, VIT- ROS SARS-CoV-2 IgG immunoassay \| IgG | 0.0%, 0.0% (Used author-adjusted estimate) | 5.8% (4.4-7.3%) |
| Bajema (CDC COVID-19 Response)[331] | 2020-08-24 to 2020-09-01 | Oklahoma | Regional | Moderate | 979 | 0-17 (n=6484); 18-49 (n=14079); 50- | NR | Non-probability | Residual sera | 5.4% (3.8-7.3%) | Ortho-Clinical Diagnostics; Abbott; Roche \| | 0.0%, 0.0% (Used author- | 5.4% (4.0-6.9%) |

| Article Author (Study Organization) | Sampling Dates (YMD) | Location | Geographic Scope | Overall Risk of Bias | Sample Size | Age | Female (%) | Sampling method | Sample frame | Uncorrected Seroprevalence (95% Confidence Interval) | Test Manufacturers \| Names \| Isotypes | Test Sens, Spec (Source) | Corrected Seroprevalence (95% Credible Interval) |
|---|---|---|---|---|---|---|---|---|---|---|---|---|---|
| | | | | | | 64 (n=12426); ≥65 (n=12316) | | | | | Elecsys Anti-SARS-CoV-2 pan-immunoglobulin immunoassay, ARCHITECT SARS-CoV-2 IgG immunoassay, VIT- ROS SARS-CoV-2 IgG immunoassay \| IgG | adjusted estimate) | |
| Maleck (University of Wisconsin)[325] | 2020-07-01 to 2020-08-31 | Wisconsin | Regional | Moderate | 996 | NR | NR | Probability | Household and community samples | 1.6% (0.6-2.5%) | Abbott \| Abbott Architect SARS-CoV- 2 IgG antibody test \| IgG | 0.0%, 99.6% (Used author-adjusted estimate) | 1.6% (0.9-2.4%) |
| Bajema (CDC COVID-19 Response)[331] | 2020-08-27 to 2020-08-29 | Nevada | Regional | Moderate | 980 | 0-17 (n=6484); 18-49 (n=14079); 50-64 (n=12426); ≥65 (n=12316) | NR | Non-probability | Residual sera | 6.5% (4.6-8.6%) | Ortho-Clinical Diagnostics; Abbott; Roche \| Elecsys Anti-SARS-CoV-2 pan-immunoglobulin immunoassay, ARCHITECT SARS-CoV-2 IgG immunoassay, VIT- ROS SARS-CoV-2 IgG immunoassay \| IgG | 0.0%, 0.0% (Used author-adjusted estimate) | 6.5% (5.0-8.1%) |
| Bajema (CDC COVID-19 Response)[331] | 2020-08-12 to 2020-08-27 | Delaware | Regional | Moderate | 1004 | 0-17 (n=6920); 18-49 (n=14571); 50-64 (n=12514 ); ≥65 (n=11856) | NR | Non-probability | Residual sera | 8.5% (5.8-11.2%) | Ortho-Clinical Diagnostics; Abbott; Roche \| Elecsys Anti-SARS-CoV-2 pan-immunoglobulin immunoassay, ARCHITECT SARS-CoV-2 IgG | 0.0%, 0.0% (Used author-adjusted estimate) | 8.5% (6.9-10.3%) |

| Article Author (Study Organization) | Sampling Dates (YMD) | Location | Geographic Scope | Overall Risk of Bias | Sample Size | Age | Female (%) | Sampling method | Sample frame | Uncorrected Seroprevalence (95% Confidence Interval) | Test Manufacturers \| Names \| Isotypes | Test Sens, Spec (Source) | Corrected Seroprevalence (95% Credible Interval) |
|---|---|---|---|---|---|---|---|---|---|---|---|---|---|
| | | | | | | | | | | | immunoassay, VIT- ROS SARS-CoV-2 IgG immunoassay \| IgG | | |
| Bajema (CDC COVID-19 Response)[331] | 2020-08-12 to 2020-08-27 | Illinois | Regional | Moderate | 985 | 0-17 (n=6920); 18-49 (n=14571); 50-64 (n=12514 ); ≥65 (n=11856) | NR | Non-probability | Residual sera | 4.9% (3.4-6.7%) | Ortho-Clinical Diagnostics; Abbott; Roche \| Elecsys Anti-SARS-CoV-2 pan-immunoglobulin immunoassay, ARCHITECT SARS-CoV-2 IgG immunoassay, VIT- ROS SARS-CoV-2 IgG immunoassay \| IgG | 0.0%, 0.0% (Used author-adjusted estimate) | 4.9% (3.7-6.4%) |
| Bajema (CDC COVID-19 Response)[331] | 2020-08-12 to 2020-08-27 | Iowa | Regional | Moderate | 980 | 0-17 (n=6920); 18-49 (n=14571); 50-64 (n=12514 ); ≥65 (n=11856) | NR | Non-probability | Residual sera | 9.4% (7.1-11.6%) | Ortho-Clinical Diagnostics; Abbott; Roche \| Elecsys Anti-SARS-CoV-2 pan-immunoglobulin immunoassay, ARCHITECT SARS-CoV-2 IgG immunoassay, VIT- ROS SARS-CoV-2 IgG immunoassay \| IgG | 0.0%, 0.0% (Used author-adjusted estimate) | 9.4% (7.7-11.3%) |
| Bajema (CDC COVID-19 Response)[331] | 2020-08-12 to 2020-08-27 | Maine | Regional | Moderate | 609 | 0-17 (n=6920); 18-49 (n=14571); 50-64 (n=12514 ); ≥65 (n=11856) | NR | Non-probability | Residual sera | 0.6% (0.0-1.4%) | Ortho-Clinical Diagnostics; Abbott; Roche \| Elecsys Anti-SARS-CoV-2 pan-immunoglobulin | 0.0%, 0.0% (Used author-adjusted estimate) | 0.6% (0.1-1.3%) |

| Article Author (Study Organization) | Sampling Dates (YMD) | Location | Geographic Scope | Overall Risk of Bias | Sample Size | Age | Female (%) | Sampling method | Sample frame | Uncorrected Seroprevalence (95% Confidence Interval) | Test Manufacturers \| Names \| Isotypes | Test Sens, Spec (Source) | Corrected Seroprevalence (95% Credible Interval) |
|---|---|---|---|---|---|---|---|---|---|---|---|---|---|
| | | | | | | | | | | | immunoassay, ARCHITECT SARS-CoV-2 IgG immunoassay, VIT- ROS SARS-CoV-2 IgG immunoassay \| IgG | | |
| Bajema (CDC COVID-19 Response)[331] | 2020-08-12 to 2020-08-27 | Massachusetts | Regional | Moderate | 980 | 0-17 (n=6920); 18-49 (n=14571); 50-64 (n=12514 ); ≥65 (n=11856) | NR | Non-probability | Residual sera | 3.7% (2.3-5.2%) | Ortho-Clinical Diagnostics; Abbott; Roche \| Elecsys Anti-SARS-CoV-2 pan-immunoglobulin immunoassay, ARCHITECT SARS-CoV-2 IgG immunoassay, VIT- ROS SARS-CoV-2 IgG immunoassay \| IgG | 0.0%, 0.0% (Used author-adjusted estimate) | 3.7% (2.6-5.0%) |
| Bajema (CDC COVID-19 Response)[331] | 2020-08-11 to 2020-08-27 | Minnesota | Regional | Moderate | 978 | 0-17 (n=6920); 18-49 (n=14571); 50-64 (n=12514 ); ≥65 (n=11856) | NR | Non-probability | Residual sera | 4.5% (3.0-5.9%) | Ortho-Clinical Diagnostics; Abbott; Roche \| Elecsys Anti-SARS-CoV-2 pan-immunoglobulin immunoassay, ARCHITECT SARS-CoV-2 IgG immunoassay, VIT- ROS SARS-CoV-2 IgG immunoassay \| IgG | 0.0%, 0.0% (Used author-adjusted estimate) | 4.5% (3.3-5.9%) |
| Bajema (CDC COVID-19 Response)[331] | 2020-08-12 to 2020-08-27 | Mississippi | Regional | Moderate | 968 | 0-17 (n=6920); 18-49 (n=14571); 50- | NR | Non-probability | Residual sera | 10.0% (6.7-14.7%) | Ortho-Clinical Diagnostics; Abbott; Roche \| Elecsys Anti- | 0.0%, 0.0% (Used author-adjusted estimate) | 10.0% (8.2-11.9%) |

| Article Author (Study Organization) | Sampling Dates (YMD) | Location | Geographic Scope | Overall Risk of Bias | Sample Size | Age | Female (%) | Sampling method | Sample frame | Uncorrected Seroprevalence (95% Confidence Interval) | Test Manufacturers \| Names \| Isotypes | Test Sens, Spec (Source) | Corrected Seroprevalence (95% Credible Interval) |
|---|---|---|---|---|---|---|---|---|---|---|---|---|---|
| | | | | | | 64 (n=12514 ); ≥65 (n=11856) | | | | | SARS-CoV-2 pan-immunoglobulin immunoassay, ARCHITECT SARS-CoV-2 IgG immunoassay, VIT- ROS SARS-CoV-2 IgG immunoassay \| IgG | | |
| Bajema (CDC COVID-19 Response)[331] | 2020-08-10 to 2020-08-27 | New Mexico | Regional | Moderate | 976 | 0-17 (n=6920); 18-49 (n=14571); 50-64 (n=12514 ); ≥65 (n=11856) | NR | Non-probability | Residual sera | 2.5% (1.5-3.8%) | Ortho-Clinical Diagnostics; Abbott; Roche \| Elecsys Anti-SARS-CoV-2 pan-immunoglobulin immunoassay, ARCHITECT SARS-CoV-2 IgG immunoassay, VIT- ROS SARS-CoV-2 IgG immunoassay \| IgG | 0.0%, 0.0% (Used author-adjusted estimate) | 2.5% (1.6-3.6%) |
| Bajema (CDC COVID-19 Response)[331] | 2020-08-11 to 2020-08-27 | North Carolina | Regional | Moderate | 982 | 0-17 (n=6920); 18-49 (n=14571); 50-64 (n=12514 ); ≥65 (n=11856) | NR | Non-probability | Residual sera | 3.8% (2.3-5.3%) | Ortho-Clinical Diagnostics; Abbott; Roche \| Elecsys Anti-SARS-CoV-2 pan-immunoglobulin immunoassay, ARCHITECT SARS-CoV-2 IgG immunoassay, VIT- ROS SARS-CoV-2 IgG immunoassay \| IgG | 0.0%, 0.0% (Used author-adjusted estimate) | 3.8% (2.7-5.1%) |

| Article Author (Study Organization) | Sampling Dates (YMD) | Location | Geographic Scope | Overall Risk of Bias | Sample Size | Age | Female (%) | Sampling method | Sample frame | Uncorrected Seroprevalence (95% Confidence Interval) | Test Manufacturers \| Names \| Isotypes | Test Sens, Spec (Source) | Corrected Seroprevalence (95% Credible Interval) |
|---|---|---|---|---|---|---|---|---|---|---|---|---|---|
| Bajema (CDC COVID-19 Response)[331] | 2020-08-12 to 2020-08-27 | Ohio | Regional | Moderate | 985 | 0-17 (n=6920); 18-49 (n=14571); 50-64 (n=12514 ); ≥65 (n=11856) | NR | Non-probability | Residual sera | 2.1% (1.1-3.3%) | Ortho-Clinical Diagnostics; Abbott; Roche \| Elecsys Anti-SARS-CoV-2 pan-immunoglobulin immunoassay, ARCHITECT SARS-CoV-2 IgG immunoassay, VIT- ROS SARS-CoV-2 IgG immunoassay \| IgG | 0.0%, 0.0% (Used author-adjusted estimate) | 2.1% (1.3-3.1%) |
| Bajema (CDC COVID-19 Response)[331] | 2020-08-10 to 2020-08-27 | Oregon | Regional | Moderate | 1086 | 0-17 (n=6920); 18-49 (n=14571); 50-64 (n=12514 ); ≥65 (n=11856) | NR | Non-probability | Residual sera | 2.4% (1.4-3.6%) | Ortho-Clinical Diagnostics; Abbott; Roche \| Elecsys Anti-SARS-CoV-2 pan-immunoglobulin immunoassay, ARCHITECT SARS-CoV-2 IgG immunoassay, VIT- ROS SARS-CoV-2 IgG immunoassay \| IgG | 0.0%, 0.0% (Used author-adjusted estimate) | 2.4% (1.6-3.4%) |
| Bajema (CDC COVID-19 Response)[331] | 2020-08-12 to 2020-08-27 | Rhode Island | Regional | High | 570 | 0-17 (n=6920); 18-49 (n=14571); 50-64 (n=12514 ); ≥65 (n=11856) | NR | Non-probability | Residual sera | 3.6% (1.7-5.9%) | Ortho-Clinical Diagnostics; Abbott; Roche \| Elecsys Anti-SARS-CoV-2 pan-immunoglobulin immunoassay, ARCHITECT SARS-CoV-2 IgG immunoassay, VIT- ROS SARS-CoV-2 IgG | 0.0%, 0.0% (Used author-adjusted estimate) | 3.6% (2.2-5.3%) |

| Article Author (Study Organization) | Sampling Dates (YMD) | Location | Geographic Scope | Overall Risk of Bias | Sample Size | Age | Female (%) | Sampling method | Sample frame | Uncorrected Seroprevalence (95% Confidence Interval) | Test Manufacturers \| Names \| Isotypes | Test Sens, Spec (Source) | Corrected Seroprevalence (95% Credible Interval) |
|---|---|---|---|---|---|---|---|---|---|---|---|---|---|
| | | | | | | | | | | | immunoassay \| IgG | | |
| Bajema (CDC COVID-19 Response)[331] | 2020-08-12 to 2020-08-27 | South Carolina | Regional | Moderate | 1063 | 0-17 (n=6920); 18-49 (n=14571); 50-64 (n=12514 ); ≥65 (n=11856) | NR | Non-probability | Residual sera | 6.0% (4.4-8.0%) | Ortho-Clinical Diagnostics; Abbott; Roche \| Elecsys Anti-SARS-CoV-2 pan-immunoglobulin immunoassay, ARCHITECT SARS-CoV-2 IgG immunoassay, VIT- ROS SARS-CoV-2 IgG immunoassay \| IgG | 0.0%, 0.0% (Used author-adjusted estimate) | 6.0% (4.6-7.5%) |
| Bajema (CDC COVID-19 Response)[331] | 2020-08-13 to 2020-08-27 | Vermont | Regional | High | 382 | 0-17 (n=6920); 18-49 (n=14571); 50-64 (n=12514 ); ≥65 (n=11856) | NR | Non-probability | Residual sera | 0.3% (0.0-0.8%) | Ortho-Clinical Diagnostics; Abbott; Roche \| Elecsys Anti-SARS-CoV-2 pan-immunoglobulin immunoassay, ARCHITECT SARS-CoV-2 IgG immunoassay, VIT- ROS SARS-CoV-2 IgG immunoassay \| IgG | 0.0%, 0.0% (Used author-adjusted estimate) | 0.3% (0.0-1.2%) |
| Bajema (CDC COVID-19 Response)[331] | 2020-08-12 to 2020-08-27 | Washington | Regional | Moderate | 1238 | 0-17 (n=6920); 18-49 (n=14571); 50-64 (n=12514 ); ≥65 (n=11856) | NR | Non-probability | Residual sera | 3.0% (1.8-4.5%) | Ortho-Clinical Diagnostics; Abbott; Roche \| Elecsys Anti-SARS-CoV-2 pan-immunoglobulin immunoassay, ARCHITECT SARS-CoV-2 IgG | 0.0%, 0.0% (Used author-adjusted estimate) | 3.0% (2.1-4.0%) |

| Article Author (Study Organization) | Sampling Dates (YMD) | Location | Geographic Scope | Overall Risk of Bias | Sample Size | Age | Female (%) | Sampling method | Sample frame | Uncorrected Seroprevalence (95% Confidence Interval) | Test Manufacturers \| Names \| Isotypes | Test Sens, Spec (Source) | Corrected Seroprevalence (95% Credible Interval) |
|---|---|---|---|---|---|---|---|---|---|---|---|---|---|
| | | | | | | | | | | | immunoassay, VIT- ROS SARS-CoV-2 IgG immunoassay \| IgG | | |
| Bajema (CDC COVID-19 Response)[331] | 2020-08-13 to 2020-08-27 | Washington D.C | Regional | Moderate | 781 | 0-17 (n=6920); 18-49 (n=14571); 50-64 (n=12514 ); ≥65 (n=11856) | NR | Non-probability | Residual sera | 6.8% (4.6-9.2%) | Ortho-Clinical Diagnostics; Abbott; Roche \| Elecsys Anti-SARS-CoV-2 pan-immunoglobulin immunoassay, ARCHITECT SARS-CoV-2 IgG immunoassay, VIT- ROS SARS-CoV-2 IgG immunoassay \| IgG | 0.0%, 0.0% (Used author-adjusted estimate) | 6.8% (5.2-8.7%) |
| Bajema (CDC COVID-19 Response)[331] | 2020-08-13 to 2020-08-27 | West Virginia | Regional | Moderate | 637 | 0-17 (n=6920); 18-49 (n=14571); 50-64 (n=12514 ); ≥65 (n=11856) | NR | Non-probability | Residual sera | 2.2% (0.5-4.9%) | Ortho-Clinical Diagnostics; Abbott; Roche \| Elecsys Anti-SARS-CoV-2 pan-immunoglobulin immunoassay, ARCHITECT SARS-CoV-2 IgG immunoassay, VIT- ROS SARS-CoV-2 IgG immunoassay \| IgG | 0.0%, 0.0% (Used author-adjusted estimate) | 2.2% (1.3-3.6%) |
| Bajema (CDC COVID-19 Response)[331] | 2020-08-12 to 2020-08-27 | Wisconsin | Regional | Moderate | 838 | 0-17 (n=6920); 18-49 (n=14571); 50-64 (n=12514 ); ≥65 (n=11856) | NR | Non-probability | Residual sera | 3.3% (2.0-4.7%) | Ortho-Clinical Diagnostics; Abbott; Roche \| Elecsys Anti-SARS-CoV-2 pan-immunoglobulin | 0.0%, 0.0% (Used author-adjusted estimate) | 3.3% (2.2-4.6%) |

| Article Author (Study Organization) | Sampling Dates (YMD) | Location | Geographic Scope | Overall Risk of Bias | Sample Size | Age | Female (%) | Sampling method | Sample frame | Uncorrected Seroprevalence (95% Confidence Interval) | Test Manufacturers \| Names \| Isotypes | Test Sens, Spec (Source) | Corrected Seroprevalence (95% Credible Interval) |
|---|---|---|---|---|---|---|---|---|---|---|---|---|---|
| | | | | | | | | | | | immunoassay, ARCHITECT SARS-CoV-2 IgG immunoassay, VIT- ROS SARS-CoV-2 IgG immunoassay \| IgG | | |
| Bajema (CDC COVID-19 Response)[331] | 2020-08-12 to 2020-08-26 | Alabama | Regional | Moderate | 974 | 0-17 (n=6920); 18-49 (n=14571); 50-64 (n=12514 ); ≥65 (n=11856) | NR | Non-probability | Residual sera | 7.6% (5.3-9.8%) | Ortho-Clinical Diagnostics; Abbott; Roche \| Elecsys Anti-SARS-CoV-2 pan-immunoglobulin immunoassay, ARCHITECT SARS-CoV-2 IgG immunoassay, VIT- ROS SARS-CoV-2 IgG immunoassay \| IgG | 0.0%, 0.0% (Used author-adjusted estimate) | 7.6% (6.1-9.4%) |
| Bajema (CDC COVID-19 Response)[331] | 2020-08-12 to 2020-08-26 | Alaska | Regional | Moderate | 704 | 0-17 (n=6920); 18-49 (n=14571); 50-64 (n=12514 ); ≥65 (n=11856) | NR | Non-probability | Residual sera | 1.3% (0.5-2.3%) | Ortho-Clinical Diagnostics; Abbott; Roche \| Elecsys Anti-SARS-CoV-2 pan-immunoglobulin immunoassay, ARCHITECT SARS-CoV-2 IgG immunoassay, VIT- ROS SARS-CoV-2 IgG immunoassay \| IgG | 0.0%, 0.0% (Used author-adjusted estimate) | 1.3% (0.6-2.3%) |
| Bajema (CDC COVID-19 Response)[331] | 2020-08-12 to 2020-08-26 | Arizona | Regional | Moderate | 1304 | 0-17 (n=6920); 18-49 (n=14571); 50- | NR | Non-probability | Residual sera | 4.7% (2.7-7.6%) | Ortho-Clinical Diagnostics; Abbott; Roche \| Elecsys Anti- | 0.0%, 0.0% (Used author-adjusted estimate) | 4.7% (3.6-5.9%) |

| Article Author (Study Organization) | Sampling Dates (YMD) | Location | Geographic Scope | Overall Risk of Bias | Sample Size | Age | Female (%) | Sampling method | Sample frame | Uncorrected Seroprevalence (95% Confidence Interval) | Test Manufacturers \| Names \| Isotypes | Test Sens, Spec (Source) | Corrected Seroprevalence (95% Credible Interval) |
|---|---|---|---|---|---|---|---|---|---|---|---|---|---|
| | | | | | | 64 (n=12514 ); ≥65 (n=11856) | | | | | SARS-CoV-2 pan-immunoglobulin immunoassay, ARCHITECT SARS-CoV-2 IgG immunoassay, VIT- ROS SARS-CoV-2 IgG immunoassay \| IgG | | |
| Bajema (CDC COVID-19 Response)[331] | 2020-08-13 to 2020-08-26 | Georgia | Regional | Moderate | 1031 | 0-17 (n=6920); 18-49 (n=14571); 50-64 (n=12514 ); ≥65 (n=11856) | NR | Non-probability | Residual sera | 7.4% (5.5-9.5%) | Ortho-Clinical Diagnostics; Abbott; Roche \| Elecsys Anti-SARS-CoV-2 pan-immunoglobulin immunoassay, ARCHITECT SARS-CoV-2 IgG immunoassay, VIT- ROS SARS-CoV-2 IgG immunoassay \| IgG | 0.0%, 0.0% (Used author-adjusted estimate) | 7.4% (5.9-9.1%) |
| Bajema (CDC COVID-19 Response)[331] | 2020-08-12 to 2020-08-26 | Idaho | Regional | Moderate | 651 | 0-17 (n=6920); 18-49 (n=14571); 50-64 (n=12514 ); ≥65 (n=11856) | NR | Non-probability | Residual sera | 4.6% (1.7-9.1%) | Ortho-Clinical Diagnostics; Abbott; Roche \| Elecsys Anti-SARS-CoV-2 pan-immunoglobulin immunoassay, ARCHITECT SARS-CoV-2 IgG immunoassay, VIT- ROS SARS-CoV-2 IgG immunoassay \| IgG | 0.0%, 0.0% (Used author-adjusted estimate) | 4.6% (3.1-6.2%) |

| Article Author (Study Organization) | Sampling Dates (YMD) | Location | Geographic Scope | Overall Risk of Bias | Sample Size | Age | Female (%) | Sampling method | Sample frame | Uncorrected Seroprevalence (95% Confidence Interval) | Test Manufacturers \| Names \| Isotypes | Test Sens, Spec (Source) | Corrected Seroprevalence (95% Credible Interval) |
|---|---|---|---|---|---|---|---|---|---|---|---|---|---|
| Bajema (CDC COVID-19 Response)[331] | 2020-08-12 to 2020-08-26 | Indiana | Regional | Moderate | 906 | 0-17 (n=6920); 18-49 (n=14571); 50-64 (n=12514 ); ≥65 (n=11856) | NR | Non-probability | Residual sera | 3.1% (1.5-4.8%) | Ortho-Clinical Diagnostics; Abbott; Roche \| Elecsys Anti-SARS-CoV-2 pan-immunoglobulin immunoassay, ARCHITECT SARS-CoV-2 IgG immunoassay, VIT- ROS SARS-CoV-2 IgG immunoassay \| IgG | 0.0%, 0.0% (Used author-adjusted estimate) | 3.1% (2.1-4.4%) |
| Bajema (CDC COVID-19 Response)[331] | 2020-08-12 to 2020-08-26 | Kentucky | Regional | Moderate | 1126 | 0-17 (n=6920); 18-49 (n=14571); 50-64 (n=12514 ); ≥65 (n=11856) | NR | Non-probability | Residual sera | 3.1% (2.0-4.5%) | Ortho-Clinical Diagnostics; Abbott; Roche \| Elecsys Anti-SARS-CoV-2 pan-immunoglobulin immunoassay, ARCHITECT SARS-CoV-2 IgG immunoassay, VIT- ROS SARS-CoV-2 IgG immunoassay \| IgG | 0.0%, 0.0% (Used author-adjusted estimate) | 3.1% (2.1-4.1%) |
| Bajema (CDC COVID-19 Response)[331] | 2020-08-10 to 2020-08-26 | Maryland | Regional | Moderate | 1174 | 0-17 (n=6920); 18-49 (n=14571); 50-64 (n=12514 ); ≥65 (n=11856) | NR | Non-probability | Residual sera | 7.0% (4.9-9.3%) | Ortho-Clinical Diagnostics; Abbott; Roche \| Elecsys Anti-SARS-CoV-2 pan-immunoglobulin immunoassay, ARCHITECT SARS-CoV-2 IgG immunoassay, VIT- ROS SARS-CoV-2 IgG | 0.0%, 0.0% (Used author-adjusted estimate) | 7.0% (5.6-8.5%) |

| Article Author (Study Organization) | Sampling Dates (YMD) | Location | Geographic Scope | Overall Risk of Bias | Sample Size | Age | Female (%) | Sampling method | Sample frame | Uncorrected Seroprevalence (95% Confidence Interval) | Test Manufacturers \| Names \| Isotypes | Test Sens, Spec (Source) | Corrected Seroprevalence (95% Credible Interval) |
|---|---|---|---|---|---|---|---|---|---|---|---|---|---|
| | | | | | | | | | | | immunoassay \| IgG | | |
| Bajema (CDC COVID-19 Response)[331] | 2020-08-12 to 2020-08-26 | Nevada | Regional | Moderate | 972 | 0-17 (n=6920); 18-49 (n=14571); 50-64 (n=12514 ); ≥65 (n=11856) | NR | Non-probability | Residual sera | 7.9% (6.1-9.8%) | Ortho-Clinical Diagnostics; Abbott; Roche \| Elecsys Anti-SARS-CoV-2 pan-immunoglobulin immunoassay, ARCHITECT SARS-CoV-2 IgG immunoassay, VIT- ROS SARS-CoV-2 IgG immunoassay \| IgG | 0.0%, 0.0% (Used author-adjusted estimate) | 7.9% (6.3-9.6%) |
| Bajema (CDC COVID-19 Response)[331] | 2020-08-10 to 2020-08-26 | New Jersey | Regional | Moderate | 1127 | 0-17 (n=6920); 18-49 (n=14571); 50-64 (n=12514 ); ≥65 (n=11856) | NR | Non-probability | Residual sera | 12.2% (10.2-14.5%) | Ortho-Clinical Diagnostics; Abbott; Roche \| Elecsys Anti-SARS-CoV-2 pan-immunoglobulin immunoassay, ARCHITECT SARS-CoV-2 IgG immunoassay, VIT- ROS SARS-CoV-2 IgG immunoassay \| IgG | 0.0%, 0.0% (Used author-adjusted estimate) | 12.2% (10.3-14.2%) |
| Bajema (CDC COVID-19 Response)[331] | 2020-08-10 to 2020-08-26 | New York | Regional | Moderate | 1122 | 0-17 (n=6920); 18-49 (n=14571); 50-64 (n=12514 ); ≥65 (n=11856) | NR | Non-probability | Residual sera | 20.6% (18.0-23.1%) | Ortho-Clinical Diagnostics; Abbott; Roche \| Elecsys Anti-SARS-CoV-2 pan-immunoglobulin immunoassay, ARCHITECT SARS-CoV-2 IgG | 0.0%, 0.0% (Used author-adjusted estimate) | 20.6% (18.3-23.0%) |

| Article Author (Study Organization) | Sampling Dates (YMD) | Location | Geographic Scope | Overall Risk of Bias | Sample Size | Age | Female (%) | Sampling method | Sample frame | Uncorrected Seroprevalence (95% Confidence Interval) | Test Manufacturers \| Names \| Isotypes | Test Sens, Spec (Source) | Corrected Seroprevalence (95% Credible Interval) |
|---|---|---|---|---|---|---|---|---|---|---|---|---|---|
| | | | | | | | | | | | immunoassay, VIT- ROS SARS-CoV-2 IgG immunoassay \| IgG | | |
| Bajema (CDC COVID-19 Response)[331] | 2020-08-12 to 2020-08-26 | North Dakota | Regional | High | 135 | 0-17 (n=6920); 18-49 (n=14571); 50-64 (n=12514 ); ≥65 (n=11856) | NR | Non-probability | Residual sera | 0.6% (0.0-1.4%) | Ortho-Clinical Diagnostics; Abbott; Roche \| Elecsys Anti-SARS-CoV-2 pan-immunoglobulin immunoassay, ARCHITECT SARS-CoV-2 IgG immunoassay, VIT- ROS SARS-CoV-2 IgG immunoassay \| IgG | 0.0%, 0.0% (Used author-adjusted estimate) | 0.6% (0.0-1.8%) |
| Bajema (CDC COVID-19 Response)[331] | 2020-08-10 to 2020-08-26 | Pennsylvania | Regional | Moderate | 887 | 0-17 (n=6920); 18-49 (n=14571); 50-64 (n=12514 ); ≥65 (n=11856) | NR | Non-probability | Residual sera | 10.1% (7.5-13.7%) | Ortho-Clinical Diagnostics; Abbott; Roche \| Elecsys Anti-SARS-CoV-2 pan-immunoglobulin immunoassay, ARCHITECT SARS-CoV-2 IgG immunoassay, VIT- ROS SARS-CoV-2 IgG immunoassay \| IgG | 0.0%, 0.0% (Used author-adjusted estimate) | 10.1% (8.2-12.1%) |
| Bajema (CDC COVID-19 Response)[331] | 2020-08-12 to 2020-08-26 | South Dakota | Regional | High | 83 | 0-17 (n=6920); 18-49 (n=14571); 50-64 (n=12514 ); ≥65 (n=11856) | NR | Non-probability | Residual sera | 0.0% (0.0-4.3%) | Ortho-Clinical Diagnostics; Abbott; Roche \| Elecsys Anti-SARS-CoV-2 pan-immunoglobulin | 0.0%, 0.0% (Used author-adjusted estimate) | 0.0% (0.0-3.0%) |

| Article Author (Study Organization) | Sampling Dates (YMD) | Location | Geographic Scope | Overall Risk of Bias | Sample Size | Age | Female (%) | Sampling method | Sample frame | Uncorrected Seroprevalence (95% Confidence Interval) | Test Manufacturers \| Names \| Isotypes | Test Sens, Spec (Source) | Corrected Seroprevalence (95% Credible Interval) |
|---|---|---|---|---|---|---|---|---|---|---|---|---|---|
| | | | | | | | | | | | immunoassay, ARCHITECT SARS-CoV-2 IgG immunoassay, VIT- ROS SARS-CoV-2 IgG immunoassay \| IgG | | |
| Bajema (CDC COVID-19 Response)[331] | 2020-08-12 to 2020-08-26 | Tennesse e | Regional | Moderate | 1029 | 0-17 (n=6920); 18-49 (n=14571); 50-64 (n=12514 ); ≥65 (n=11856) | NR | Non-probability | Residual sera | 6.7% (5.1-8.6%) | Ortho-Clinical Diagnostics; Abbott; Roche \| Elecsys Anti-SARS-CoV-2 pan-immunoglobuli n immunoassay, ARCHITECT SARS-CoV-2 IgG immunoassay, VIT- ROS SARS-CoV-2 IgG immunoassay \| IgG | 0.0%, 0.0% (Used author-adjusted estimate) | 6.7% (5.2-8.2%) |
| Bajema (CDC COVID-19 Response)[331] | 2020-08-10 to 2020-08-26 | Virginia | Regional | Moderate | 1243 | 0-17 (n=6920); 18-49 (n=14571); 50-64 (n=12514 ); ≥65 (n=11856) | NR | Non-probability | Residual sera | 4.7% (2.5-7.2%) | Ortho-Clinical Diagnostics; Abbott; Roche \| Elecsys Anti-SARS-CoV-2 pan-immunoglobuli n immunoassay, ARCHITECT SARS-CoV-2 IgG immunoassay, VIT- ROS SARS-CoV-2 IgG immunoassay \| IgG | 0.0%, 0.0% (Used author-adjusted estimate) | 4.7% (3.6-5.9%) |
| Bajema (CDC COVID-19 Response)[331] | 2020-08-11 to 2020-08-25 | Arkansas | Regional | Moderate | 998 | 0-17 (n=6920); 18-49 (n=14571); 50- | NR | Non-probability | Residual sera | 3.0% (1.8-4.3%) | Ortho-Clinical Diagnostics; Abbott; Roche \| Elecsys Anti- | 0.0%, 0.0% (Used author-adjusted estimate) | 3.0% (2.0-4.1%) |

| Article Author (Study Organization) | Sampling Dates (YMD) | Location | Geographic Scope | Overall Risk of Bias | Sample Size | Age | Female (%) | Sampling method | Sample frame | Uncorrected Seroprevalence (95% Confidence Interval) | Test Manufacturers \| Names \| Isotypes | Test Sens, Spec (Source) | Corrected Seroprevalence (95% Credible Interval) |
|---|---|---|---|---|---|---|---|---|---|---|---|---|---|
| | | | | | | 64 (n=12514 ); ≥65 (n=11856) | | | | | SARS-CoV-2 pan-immunoglobulin immunoassay, ARCHITECT SARS-CoV-2 IgG immunoassay, VIT- ROS SARS-CoV-2 IgG immunoassay \| IgG | | |
| Bajema (CDC COVID-19 Response)[331] | 2020-08-10 to 2020-08-25 | Colorado | Regional | Moderate | 1019 | 0-17 (n=6920); 18-49 (n=14571); 50-64 (n=12514 ); ≥65 (n=11856) | NR | Non-probability | Residual sera | 3.3% (1.8-4.8%) | Ortho-Clinical Diagnostics; Abbott; Roche \| Elecsys Anti-SARS-CoV-2 pan-immunoglobulin immunoassay, ARCHITECT SARS-CoV-2 IgG immunoassay, VIT- ROS SARS-CoV-2 IgG immunoassay \| IgG | 0.0%, 0.0% (Used author-adjusted estimate) | 3.3% (2.3-4.5%) |
| Bajema (CDC COVID-19 Response)[331] | 2020-08-11 to 2020-08-25 | Kansas | Regional | Moderate | 1016 | 0-17 (n=6920); 18-49 (n=14571); 50-64 (n=12514 ); ≥65 (n=11856) | NR | Non-probability | Residual sera | 3.7% (2.0-5.5%) | Ortho-Clinical Diagnostics; Abbott; Roche \| Elecsys Anti-SARS-CoV-2 pan-immunoglobulin immunoassay, ARCHITECT SARS-CoV-2 IgG immunoassay, VIT- ROS SARS-CoV-2 IgG immunoassay \| IgG | 0.0%, 0.0% (Used author-adjusted estimate) | 3.7% (2.6-4.9%) |

| Article Author (Study Organization) | Sampling Dates (YMD) | Location | Geographic Scope | Overall Risk of Bias | Sample Size | Age | Female (%) | Sampling method | Sample frame | Uncorrected Seroprevalence (95% Confidence Interval) | Test Manufacturers \| Names \| Isotypes | Test Sens, Spec (Source) | Corrected Seroprevalence (95% Credible Interval) |
|---|---|---|---|---|---|---|---|---|---|---|---|---|---|
| Bajema (CDC COVID-19 Response)[331] | 2020-08-12 to 2020-08-25 | Louisiana | Regional | Moderate | 989 | 0-17 (n=6920); 18-49 (n=14571); 50-64 (n=12514 ); ≥65 (n=11856) | NR | Non-probability | Residual sera | 11.8% (9.3-14.8%) | Ortho-Clinical Diagnostics; Abbott; Roche \| Elecsys Anti-SARS-CoV-2 pan-immunoglobulin immunoassay, ARCHITECT SARS-CoV-2 IgG immunoassay, VIT- ROS SARS-CoV-2 IgG immunoassay \| IgG | 0.0%, 0.0% (Used author-adjusted estimate) | 11.8% (9.8-13.8%) |
| Bajema (CDC COVID-19 Response)[331] | 2020-08-12 to 2020-08-25 | Michigan | Regional | Moderate | 983 | 0-17 (n=6920); 18-49 (n=14571); 50-64 (n=12514 ); ≥65 (n=11856) | NR | Non-probability | Residual sera | 4.2% (2.9-5.8%) | Ortho-Clinical Diagnostics; Abbott; Roche \| Elecsys Anti-SARS-CoV-2 pan-immunoglobulin immunoassay, ARCHITECT SARS-CoV-2 IgG immunoassay, VIT- ROS SARS-CoV-2 IgG immunoassay \| IgG | 0.0%, 0.0% (Used author-adjusted estimate) | 4.2% (3.1-5.6%) |
| Bajema (CDC COVID-19 Response)[331] | 2020-08-11 to 2020-08-25 | Nebraska | Regional | Moderate | 990 | 0-17 (n=6920); 18-49 (n=14571); 50-64 (n=12514 ); ≥65 (n=11856) | NR | Non-probability | Residual sera | 7.9% (5.5-10.6%) | Ortho-Clinical Diagnostics; Abbott; Roche \| Elecsys Anti-SARS-CoV-2 pan-immunoglobulin immunoassay, ARCHITECT SARS-CoV-2 IgG immunoassay, VIT- ROS SARS-CoV-2 IgG | 0.0%, 0.0% (Used author-adjusted estimate) | 7.9% (6.3-9.7%) |

| Article Author (Study Organization) | Sampling Dates (YMD) | Location | Geographic Scope | Overall Risk of Bias | Sample Size | Age | Female (%) | Sampling method | Sample frame | Uncorrected Seroprevalence (95% Confidence Interval) | Test Manufacturers \| Names \| Isotypes | Test Sens, Spec (Source) | Corrected Seroprevalence (95% Credible Interval) |
|---|---|---|---|---|---|---|---|---|---|---|---|---|---|
| | | | | | | | | | | | immunoassay \| IgG | | |
| Bajema (CDC COVID-19 Response)[331] | 2020-08-13 to 2020-08-25 | New Hampshire | Regional | Moderate | 1125 | 0-17 (n=6920); 18-49 (n=14571); 50-64 (n=12514 ); ≥65 (n=11856) | NR | Non-probability | Residual sera | 0.8% (0.3-1.2%) | Ortho-Clinical Diagnostics; Abbott; Roche \| Elecsys Anti-SARS-CoV-2 pan-immunoglobulin immunoassay, ARCHITECT SARS-CoV-2 IgG immunoassay, VIT- ROS SARS-CoV-2 IgG immunoassay \| IgG | 0.0%, 0.0% (Used author-adjusted estimate) | 0.8% (0.4-1.5%) |
| Bajema (CDC COVID-19 Response)[331] | 2020-08-15 to 2020-08-25 | Utah | Regional | Moderate | 871 | 0-17 (n=6920); 18-49 (n=14571); 50-64 (n=12514 ); ≥65 (n=11856) | NR | Non-probability | Residual sera | 5.5% (2.9-8.7%) | Ortho-Clinical Diagnostics; Abbott; Roche \| Elecsys Anti-SARS-CoV-2 pan-immunoglobulin immunoassay, ARCHITECT SARS-CoV-2 IgG immunoassay, VIT- ROS SARS-CoV-2 IgG immunoassay \| IgG | 0.0%, 0.0% (Used author-adjusted estimate) | 5.5% (4.0-7.0%) |
| Bajema (CDC COVID-19 Response)[331] | 2020-08-11 to 2020-08-24 | Connecticut | Regional | Moderate | 983 | 0-17 (n=6920); 18-49 (n=14571); 50-64 (n=12514 ); ≥65 (n=11856) | NR | Non-probability | Residual sera | 2.4% (1.4-3.5%) | Ortho-Clinical Diagnostics; Abbott; Roche \| Elecsys Anti-SARS-CoV-2 pan-immunoglobulin immunoassay, ARCHITECT SARS-CoV-2 IgG | 0.0%, 0.0% (Used author-adjusted estimate) | 2.4% (1.5-3.4%) |

| Article Author (Study Organization) | Sampling Dates (YMD) | Location | Geographic Scope | Overall Risk of Bias | Sample Size | Age | Female (%) | Sampling method | Sample frame | Uncorrected Seroprevalence (95% Confidence Interval) | Test Manufacturers \| Names \| Isotypes | Test Sens, Spec (Source) | Corrected Seroprevalence (95% Credible Interval) |
|---|---|---|---|---|---|---|---|---|---|---|---|---|---|
| | | | | | | | | | | | immunoassay, VIT- ROS SARS-CoV-2 IgG immunoassay \| IgG | | |
| Bajema (CDC COVID-19 Response)[331] | 2020-08-12 to 2020-08-24 | Montana | Regional | High | 383 | 0-17 (n=6920); 18-49 (n=14571); 50-64 (n=12514 ); ≥65 (n=11856) | NR | Non-probability | Residual sera | 1.3% (0.4-2.5%) | Ortho-Clinical Diagnostics; Abbott; Roche \| Elecsys Anti-SARS-CoV-2 pan-immunoglobulin immunoassay, ARCHITECT SARS-CoV-2 IgG immunoassay, VIT- ROS SARS-CoV-2 IgG immunoassay \| IgG | 0.0%, 0.0% (Used author-adjusted estimate) | 1.3% (0.4-2.5%) |
| Bajema (CDC COVID-19 Response)[331] | 2020-08-12 to 2020-08-24 | Texas | Regional | Moderate | 1001 | 0-17 (n=6920); 18-49 (n=14571); 50-64 (n=12514 ); ≥65 (n=11856) | NR | Non-probability | Residual sera | 6.5% (4.7-8.5%) | Ortho-Clinical Diagnostics; Abbott; Roche \| Elecsys Anti-SARS-CoV-2 pan-immunoglobulin immunoassay, ARCHITECT SARS-CoV-2 IgG immunoassay, VIT- ROS SARS-CoV-2 IgG immunoassay \| IgG | 0.0%, 0.0% (Used author-adjusted estimate) | 6.5% (5.1-8.1%) |
| Bajema (CDC COVID-19 Response)[331] | 2020-08-13 to 2020-08-24 | Wyoming | Regional | High | 86 | 0-17 (n=6920); 18-49 (n=14571); 50-64 (n=12514 ); ≥65 (n=11856) | NR | Non-probability | Residual sera | 0.8% (0.0-3.1%) | Ortho-Clinical Diagnostics; Abbott; Roche \| Elecsys Anti-SARS-CoV-2 pan-immunoglobulin | 0.0%, 0.0% (Used author-adjusted estimate) | 0.8% (0.0-2.9%) |

| Article Author (Study Organization) | Sampling Dates (YMD) | Location | Geographic Scope | Overall Risk of Bias | Sample Size | Age | Female (%) | Sampling method | Sample frame | Uncorrected Seroprevalence (95% Confidence Interval) | Test Manufacturers \| Names \| Isotypes | Test Sens, Spec (Source) | Corrected Seroprevalence (95% Credible Interval) |
|---|---|---|---|---|---|---|---|---|---|---|---|---|---|
| | | | | | | | | | | | immunoassay, ARCHITECT SARS-CoV-2 IgG immunoassay, VIT- ROS SARS-CoV-2 IgG immunoassay \| IgG | | |
| Bajema (CDC COVID-19 Response)[331] | 2020-08-12 to 2020-08-21 | Missouri | Regional | Moderate | 1021 | 0-17 (n=6920); 18-49 (n=14571); 50-64 (n=12514 ); ≥65 (n=11856) | NR | Non-probability | Residual sera | 3.5% (2.2-5.1%) | Ortho-Clinical Diagnostics; Abbott; Roche \| Elecsys Anti-SARS-CoV-2 pan-immunoglobulin immunoassay, ARCHITECT SARS-CoV-2 IgG immunoassay, VIT- ROS SARS-CoV-2 IgG immunoassay \| IgG | 0.0%, 0.0% (Used author-adjusted estimate) | 3.5% (2.4-4.7%) |
| Bajema (CDC COVID-19 Response)[331] | 2020-08-13 to 2020-08-19 | California | Regional | Moderate | 980 | 0-17 (n=6920); 18-49 (n=14571); 50-64 (n=12514 ); ≥65 (n=11856) | NR | Non-probability | Residual sera | 4.3% (2.9-6.1%) | Ortho-Clinical Diagnostics; Abbott; Roche \| Elecsys Anti-SARS-CoV-2 pan-immunoglobulin immunoassay, ARCHITECT SARS-CoV-2 IgG immunoassay, VIT- ROS SARS-CoV-2 IgG immunoassay \| IgG | 0.0%, 0.0% (Used author-adjusted estimate) | 4.3% (3.1-5.7%) |
| Bajema (CDC COVID-19 Response)[331] | 2020-08-10 to 2020-08-18 | Oklahoma | Regional | Moderate | 979 | 0-17 (n=6920); 18-49 (n=14571); 50- | NR | Non-probability | Residual sera | 4.0% (2.7-5.5%) | Ortho-Clinical Diagnostics; Abbott; Roche \| Elecsys Anti- | 0.0%, 0.0% (Used author-adjusted estimate) | 4.0% (2.9-5.3%) |

| Article Author (Study Organization) | Sampling Dates (YMD) | Location | Geographic Scope | Overall Risk of Bias | Sample Size | Age | Female (%) | Sampling method | Sample frame | Uncorrected Seroprevalence (95% Confidence Interval) | Test Manufacturers \| Names \| Isotypes | Test Sens, Spec (Source) | Corrected Seroprevalence (95% Credible Interval) |
|---|---|---|---|---|---|---|---|---|---|---|---|---|---|
| | | | | | | 64 (n=12514 ); ≥65 (n=11856) | | | | | SARS-CoV-2 pan-immunoglobulin immunoassay, ARCHITECT SARS-CoV-2 IgG immunoassay, VIT- ROS SARS-CoV-2 IgG immunoassay \| IgG | | |
| Bajema (CDC COVID-19 Response)[331] | 2020-08-10 to 2020-08-17 | Puerto Rico | Regional | Moderate | 986 | 0-17 (n=6920); 18-49 (n=14571); 50-64 (n=12514 ); ≥65 (n=11856) | NR | Non-probability | Residual sera | 2.2% (1.2-3.3%) | Ortho-Clinical Diagnostics; Abbott; Roche \| Elecsys Anti-SARS-CoV-2 pan-immunoglobulin immunoassay, ARCHITECT SARS-CoV-2 IgG immunoassay, VIT- ROS SARS-CoV-2 IgG immunoassay \| IgG | 0.0%, 0.0% (Used author-adjusted estimate) | 2.2% (1.4-3.2%) |
| Bruckner (University of California Irvine)[336] | 2020-07-10 to 2020-08-16 | California | Local | Low | 2979 | 18-34 (n=673), 35-54 (n=1362), 55+ (n=944) | 56.4 | Probability | Household and community samples | 11.5% (10.5-12.4%) | NR \| coronavirus antigen microarray (CoVAM) \| IgG, IgM | 94.0%, 100.0% (Author-reported independent evaluation) | 11.5% (8.8-13.6%) |
| Heim (UW-Madison)[339] | 2020-07-15 to 2020-08-15 | Wisconsin | Regional | Unclear | 1056 | NR | NR | Probability | Household and community samples | 1.6% | NR \| NR \| NR | No data | - |
| Bajema (CDC COVID-19 Response)[331] | 2020-08-14 to 2020-08-14 | Florida | Regional | Moderate | 978 | 0-17 (n=6920); 18-49 (n=14571); 50-64 (n=12514 ); ≥65 (n=11856) | NR | Non-probability | Residual sera | 4.5% (3.1-6.1%) | Ortho-Clinical Diagnostics; Abbott; Roche \| Elecsys Anti-SARS-CoV-2 pan- | 0.0%, 0.0% (Used author-adjusted estimate) | 4.5% (3.3-5.9%) |

| Article Author (Study Organization) | Sampling Dates (YMD) | Location | Geographic Scope | Overall Risk of Bias | Sample Size | Age | Female (%) | Sampling method | Sample frame | Uncorrected Seroprevalence (95% Confidence Interval) | Test Manufacturers \| Names \| Isotypes | Test Sens, Spec (Source) | Corrected Seroprevalence (95% Credible Interval) |
|---|---|---|---|---|---|---|---|---|---|---|---|---|---|
| | | | | | | | | | | | immunoglobulin immunoassay, ARCHITECT SARS-CoV-2 IgG immunoassay, VIT- ROS SARS-CoV-2 IgG immunoassay \| IgG | | |
| Bajema (CDC COVID-19 Response)[331] | 2020-07-29 to 2020-08-13 | Alabama | Regional | Moderate | 938 | 0-17 (n=6700); 18-49 (n=11237); 50-64 (n=10367); ≥65 (n=10408) | NR | Non-probability | Residual sera | 5.8% (4.2-7.7%) | Ortho-Clinical Diagnostics; Abbott; Roche \| Elecsys Anti-SARS-CoV-2 pan-immunoglobulin immunoassay, ARCHITECT SARS-CoV-2 IgG immunoassay, VIT- ROS SARS-CoV-2 IgG immunoassay \| IgG | 0.0%, 0.0% (Used author-adjusted estimate) | 5.8% (4.4-7.4%) |
| Bajema (CDC COVID-19 Response)[331] | 2020-07-29 to 2020-08-13 | Arkansas | Regional | Moderate | 937 | 0-17 (n=6700); 18-49 (n=11237); 50-64 (n=10367); ≥65 (n=10408) | NR | Non-probability | Residual sera | 4.1% (2.7-5.6%) | Ortho-Clinical Diagnostics; Abbott; Roche \| Elecsys Anti-SARS-CoV-2 pan-immunoglobulin immunoassay, ARCHITECT SARS-CoV-2 IgG immunoassay, VIT- ROS SARS-CoV-2 IgG immunoassay \| IgG | 0.0%, 0.0% (Used author-adjusted estimate) | 4.1% (2.9-5.5%) |

| Article Author (Study Organization) | Sampling Dates (YMD) | Location | Geographic Scope | Overall Risk of Bias | Sample Size | Age | Female (%) | Sampling method | Sample frame | Uncorrected Seroprevalence (95% Confidence Interval) | Test Manufacturers \| Names \| Isotypes | Test Sens, Spec (Source) | Corrected Seroprevalence (95% Credible Interval) |
|---|---|---|---|---|---|---|---|---|---|---|---|---|---|
| Bajema (CDC COVID-19 Response)[331] | 2020-07-29 to 2020-08-13 | Delaware | Regional | Moderate | 804 | 0-17 (n=6700); 18-49 (n=11237); 50-64 (n=10367); ≥65 (n=10408) | NR | Non-probability | Residual sera | 5.7% (3.5-8.2%) | Ortho-Clinical Diagnostics; Abbott; Roche \| Elecsys Anti-SARS-CoV-2 pan-immunoglobulin immunoassay, ARCHITECT SARS-CoV-2 IgG immunoassay, VIT- ROS SARS-CoV-2 IgG immunoassay \| IgG | 0.0%, 0.0% (Used author-adjusted estimate) | 5.7% (4.2-7.3%) |
| Bajema (CDC COVID-19 Response)[331] | 2020-07-29 to 2020-08-13 | Iowa | Regional | Moderate | 811 | 0-17 (n=6700); 18-49 (n=11237); 50-64 (n=10367); ≥65 (n=10408) | NR | Non-probability | Residual sera | 8.6% (6.0-11.3%) | Ortho-Clinical Diagnostics; Abbott; Roche \| Elecsys Anti-SARS-CoV-2 pan-immunoglobulin immunoassay, ARCHITECT SARS-CoV-2 IgG immunoassay, VIT- ROS SARS-CoV-2 IgG immunoassay \| IgG | 0.0%, 0.0% (Used author-adjusted estimate) | 8.6% (6.7-10.6%) |
| Bajema (CDC COVID-19 Response)[331] | 2020-07-30 to 2020-08-13 | Kentucky | Regional | Moderate | 975 | 0-17 (n=6700); 18-49 (n=11237); 50-64 (n=10367); ≥65 (n=10408) | NR | Non-probability | Residual sera | 2.4% (1.3-3.6%) | Ortho-Clinical Diagnostics; Abbott; Roche \| Elecsys Anti-SARS-CoV-2 pan-immunoglobulin immunoassay, ARCHITECT SARS-CoV-2 IgG immunoassay, VIT- ROS SARS-CoV-2 IgG | 0.0%, 0.0% (Used author-adjusted estimate) | 2.4% (1.5-3.5%) |

| Article Author (Study Organization) | Sampling Dates (YMD) | Location | Geographic Scope | Overall Risk of Bias | Sample Size | Age | Female (%) | Sampling method | Sample frame | Uncorrected Seroprevalence (95% Confidence Interval) | Test Manufacturers \| Names \| Isotypes | Test Sens, Spec (Source) | Corrected Seroprevalence (95% Credible Interval) |
|---|---|---|---|---|---|---|---|---|---|---|---|---|---|
| | | | | | | | | | | | immunoassay \| IgG | | |
| Bajema (CDC COVID-19 Response)[331] | 2020-07-28 to 2020-08-13 | Louisiana | Regional | Moderate | 1002 | 0-17 (n=6700); 18-49 (n=11237); 50-64 (n=10367); ≥65 (n=10408) | NR | Non-probability | Residual sera | 9.6% (6.9-12.3%) | Ortho-Clinical Diagnostics; Abbott; Roche \| Elecsys Anti-SARS-CoV-2 pan-immunoglobulin immunoassay, ARCHITECT SARS-CoV-2 IgG immunoassay, VIT- ROS SARS-CoV-2 IgG immunoassay \| IgG | 0.0%, 0.0% (Used author-adjusted estimate) | 9.6% (7.9-11.5%) |
| Bajema (CDC COVID-19 Response)[331] | 2020-07-29 to 2020-08-13 | Minnesota | Regional | Moderate | 879 | 0-17 (n=6700); 18-49 (n=11237); 50-64 (n=10367); ≥65 (n=10408) | NR | Non-probability | Residual sera | 3.5% (2.1-5.0%) | Ortho-Clinical Diagnostics; Abbott; Roche \| Elecsys Anti-SARS-CoV-2 pan-immunoglobulin immunoassay, ARCHITECT SARS-CoV-2 IgG immunoassay, VIT- ROS SARS-CoV-2 IgG immunoassay \| IgG | 0.0%, 0.0% (Used author-adjusted estimate) | 3.5% (2.4-4.8%) |
| Bajema (CDC COVID-19 Response)[331] | 2020-07-30 to 2020-08-13 | Mississippi | Regional | Moderate | 769 | 0-17 (n=6700); 18-49 (n=11237); 50-64 (n=10367); ≥65 (n=10408) | NR | Non-probability | Residual sera | 7.1% (3.8-11.1%) | Ortho-Clinical Diagnostics; Abbott; Roche \| Elecsys Anti-SARS-CoV-2 pan-immunoglobulin immunoassay, ARCHITECT SARS-CoV-2 IgG | 0.0%, 0.0% (Used author-adjusted estimate) | 7.1% (5.4-9.0%) |

| Article Author (Study Organization) | Sampling Dates (YMD) | Location | Geographic Scope | Overall Risk of Bias | Sample Size | Age | Female (%) | Sampling method | Sample frame | Uncorrected Seroprevalence (95% Confidence Interval) | Test Manufacturers \| Names \| Isotypes | Test Sens, Spec (Source) | Corrected Seroprevalence (95% Credible Interval) |
|---|---|---|---|---|---|---|---|---|---|---|---|---|---|
| | | | | | | | | | | | immunoassay, VIT- ROS SARS-CoV-2 IgG immunoassay \| IgG | | |
| Bajema (CDC COVID-19 Response)[331] | 2020-07-28 to 2020-08-13 | Nebraska | Regional | Moderate | 954 | 0-17 (n=6700); 18-49 (n=11237); 50-64 (n=10367); ≥65 (n=10408) | NR | Non-probability | Residual sera | 7.4% (5.2-9.8%) | Ortho-Clinical Diagnostics; Abbott; Roche \| Elecsys Anti-SARS-CoV-2 pan-immunoglobulin immunoassay, ARCHITECT SARS-CoV-2 IgG immunoassay, VIT- ROS SARS-CoV-2 IgG immunoassay \| IgG | 0.0%, 0.0% (Used author-adjusted estimate) | 7.4% (5.8-9.1%) |
| Bajema (CDC COVID-19 Response)[331] | 2020-07-29 to 2020-08-13 | New Mexico | Regional | Moderate | 964 | 0-17 (n=6700); 18-49 (n=11237); 50-64 (n=10367); ≥65 (n=10408) | NR | Non-probability | Residual sera | 2.0% (1.1-3.0%) | Ortho-Clinical Diagnostics; Abbott; Roche \| Elecsys Anti-SARS-CoV-2 pan-immunoglobulin immunoassay, ARCHITECT SARS-CoV-2 IgG immunoassay, VIT- ROS SARS-CoV-2 IgG immunoassay \| IgG | 0.0%, 0.0% (Used author-adjusted estimate) | 2.0% (1.2-3.0%) |
| Bajema (CDC COVID-19 Response)[331] | 2020-07-30 to 2020-08-13 | South Carolina | Regional | Moderate | 840 | 0-17 (n=6700); 18-49 (n=11237); 50-64 (n=10367); ≥65 (n=10408) | NR | Non-probability | Residual sera | 8.1% (5.8-10.7%) | Ortho-Clinical Diagnostics; Abbott; Roche \| Elecsys Anti-SARS-CoV-2 pan-immunoglobulin | 0.0%, 0.0% (Used author-adjusted estimate) | 8.1% (6.4-10.1%) |

| Article Author (Study Organization) | Sampling Dates (YMD) | Location | Geographic Scope | Overall Risk of Bias | Sample Size | Age | Female (%) | Sampling method | Sample frame | Uncorrected Seroprevalence (95% Confidence Interval) | Test Manufacturers \| Names \| Isotypes | Test Sens, Spec (Source) | Corrected Seroprevalence (95% Credible Interval) |
|---|---|---|---|---|---|---|---|---|---|---|---|---|---|
| | | | | | | | | | | | immunoassay, ARCHITECT SARS-CoV-2 IgG immunoassay, VIT- ROS SARS-CoV-2 IgG immunoassay \| IgG | | |
| Bajema (CDC COVID-19 Response)[331] | 2020-07-30 to 2020-08-13 | Washington D.C | Regional | Moderate | 717 | 0-17 (n=6700); 18-49 (n=11237); 50-64 (n=10367); ≥65 (n=10408) | NR | Non-probability | Residual sera | 3.9% (2.2-5.8%) | Ortho-Clinical Diagnostics; Abbott; Roche \| Elecsys Anti-SARS-CoV-2 pan-immunoglobulin immunoassay, ARCHITECT SARS-CoV-2 IgG immunoassay, VIT- ROS SARS-CoV-2 IgG immunoassay \| IgG | 0.0%, 0.0% (Used author-adjusted estimate) | 3.9% (2.6-5.4%) |
| Bajema (CDC COVID-19 Response)[331] | 2020-07-30 to 2020-08-13 | West Virginia | Regional | High | 317 | 0-17 (n=6700); 18-49 (n=11237); 50-64 (n=10367); ≥65 (n=10408) | NR | Non-probability | Residual sera | 1.2% (0.2-2.5%) | Ortho-Clinical Diagnostics; Abbott; Roche \| Elecsys Anti-SARS-CoV-2 pan-immunoglobulin immunoassay, ARCHITECT SARS-CoV-2 IgG immunoassay, VIT- ROS SARS-CoV-2 IgG immunoassay \| IgG | 0.0%, 0.0% (Used author-adjusted estimate) | 1.2% (0.3-2.5%) |
| Bajema (CDC COVID-19 Response)[331] | 2020-07-30 to 2020-08-13 | Wisconsin | Regional | Moderate | 814 | 0-17 (n=6700); 18-49 (n=11237); 50- | NR | Non-probability | Residual sera | 1.8% (0.8-3.2%) | Ortho-Clinical Diagnostics; Abbott; Roche \| Elecsys Anti- | 0.0%, 0.0% (Used author-adjusted estimate) | 1.8% (1.0-2.8%) |

| Article Author (Study Organization) | Sampling Dates (YMD) | Location | Geographic Scope | Overall Risk of Bias | Sample Size | Age | Female (%) | Sampling method | Sample frame | Uncorrected Seroprevalence (95% Confidence Interval) | Test Manufacturers \| Names \| Isotypes | Test Sens, Spec (Source) | Corrected Seroprevalence (95% Credible Interval) |
|---|---|---|---|---|---|---|---|---|---|---|---|---|---|
| | | | | | | 64 (n=10367); ≥65 (n=10408) | | | | | SARS-CoV-2 pan-immunoglobulin immunoassay, ARCHITECT SARS-CoV-2 IgG immunoassay, VIT- ROS SARS-CoV-2 IgG immunoassay \| IgG | | |
| Bajema (CDC COVID-19 Response)[331] | 2020-07-29 to 2020-08-12 | North Dakota | Regional | High | 121 | 0-17 (n=6700); 18-49 (n=11237); 50-64 (n=10367); ≥65 (n=10408) | NR | Non-probability | Residual sera | 7.3% (1.3-14.3%) | Ortho-Clinical Diagnostics; Abbott; Roche \| Elecsys Anti-SARS-CoV-2 pan-immunoglobulin immunoassay, ARCHITECT SARS-CoV-2 IgG immunoassay, VIT- ROS SARS-CoV-2 IgG immunoassay \| IgG | 0.0%, 0.0% (Used author-adjusted estimate) | 7.3% (3.2-12.1%) |
| Bajema (CDC COVID-19 Response)[331] | 2020-08-06 to 2020-08-11 | Alaska | Regional | High | 242 | 0-17 (n=6700); 18-49 (n=11237); 50-64 (n=10367); ≥65 (n=10408) | NR | Non-probability | Residual sera | 0.3% (0.0-1.1%) | Ortho-Clinical Diagnostics; Abbott; Roche \| Elecsys Anti-SARS-CoV-2 pan-immunoglobulin immunoassay, ARCHITECT SARS-CoV-2 IgG immunoassay, VIT- ROS SARS-CoV-2 IgG immunoassay \| IgG | 0.0%, 0.0% (Used author-adjusted estimate) | 0.3% (0.0-1.0%) |

| Article Author (Study Organization) | Sampling Dates (YMD) | Location | Geographic Scope | Overall Risk of Bias | Sample Size | Age | Female (%) | Sampling method | Sample frame | Uncorrected Seroprevalence (95% Confidence Interval) | Test Manufacturers \| Names \| Isotypes | Test Sens, Spec (Source) | Corrected Seroprevalence (95% Credible Interval) |
|---|---|---|---|---|---|---|---|---|---|---|---|---|---|
| Bajema (CDC COVID-19 Response)[331] | 2020-07-31 to 2020-08-11 | Arizona | Regional | High | 591 | 0-17 (n=6700); 18-49 (n=11237); 50-64 (n=10367); ≥65 (n=10408) | NR | Non-probability | Residual sera | 8.2% (4.1-13.6%) | Ortho-Clinical Diagnostics; Abbott; Roche \| Elecsys Anti-SARS-CoV-2 pan-immunoglobulin immunoassay, ARCHITECT SARS-CoV-2 IgG immunoassay, VIT- ROS SARS-CoV-2 IgG immunoassay \| IgG | 0.0%, 0.0% (Used author-adjusted estimate) | 8.2% (6.1-10.5%) |
| Bajema (CDC COVID-19 Response)[331] | 2020-08-02 to 2020-08-11 | Georgia | Regional | Moderate | 985 | 0-17 (n=6700); 18-49 (n=11237); 50-64 (n=10367); ≥65 (n=10408) | NR | Non-probability | Residual sera | 6.8% (4.8-8.8%) | Ortho-Clinical Diagnostics; Abbott; Roche \| Elecsys Anti-SARS-CoV-2 pan-immunoglobulin immunoassay, ARCHITECT SARS-CoV-2 IgG immunoassay, VIT- ROS SARS-CoV-2 IgG immunoassay \| IgG | 0.0%, 0.0% (Used author-adjusted estimate) | 6.8% (5.3-8.4%) |
| Bajema (CDC COVID-19 Response)[331] | 2020-08-04 to 2020-08-11 | Idaho | Regional | High | 246 | 0-17 (n=6700); 18-49 (n=11237); 50-64 (n=10367); ≥65 (n=10408) | NR | Non-probability | Residual sera | 4.5% (1.8-8.0%) | Ortho-Clinical Diagnostics; Abbott; Roche \| Elecsys Anti-SARS-CoV-2 pan-immunoglobulin immunoassay, ARCHITECT SARS-CoV-2 IgG immunoassay, VIT- ROS SARS-CoV-2 IgG | 0.0%, 0.0% (Used author-adjusted estimate) | 4.5% (2.4-7.6%) |

| Article Author (Study Organization) | Sampling Dates (YMD) | Location | Geographic Scope | Overall Risk of Bias | Sample Size | Age | Female (%) | Sampling method | Sample frame | Uncorrected Seroprevalence (95% Confidence Interval) | Test Manufacturers \| Names \| Isotypes | Test Sens, Spec (Source) | Corrected Seroprevalence (95% Credible Interval) |
|---|---|---|---|---|---|---|---|---|---|---|---|---|---|
| | | | | | | | | | | | immunoassay \| IgG | | |
| Bajema (CDC COVID-19 Response)[331] | 2020-07-31 to 2020-08-11 | Indiana | Regional | High | 579 | 0-17 (n=6700); 18-49 (n=11237); 50-64 (n=10367); ≥65 (n=10408) | NR | Non-probability | Residual sera | 2.2% (1.1-3.6%) | Ortho-Clinical Diagnostics; Abbott; Roche \| Elecsys Anti-SARS-CoV-2 pan-immunoglobulin immunoassay, ARCHITECT SARS-CoV-2 IgG immunoassay, VIT- ROS SARS-CoV-2 IgG immunoassay \| IgG | 0.0%, 0.0% (Used author-adjusted estimate) | 2.2% (1.1-3.5%) |
| Bajema (CDC COVID-19 Response)[331] | 2020-07-30 to 2020-08-11 | Maine | Regional | High | 598 | 0-17 (n=6700); 18-49 (n=11237); 50-64 (n=10367); ≥65 (n=10408) | NR | Non-probability | Residual sera | 0.5% (0.0-1.2%) | Ortho-Clinical Diagnostics; Abbott; Roche \| Elecsys Anti-SARS-CoV-2 pan-immunoglobulin immunoassay, ARCHITECT SARS-CoV-2 IgG immunoassay, VIT- ROS SARS-CoV-2 IgG immunoassay \| IgG | 0.0%, 0.0% (Used author-adjusted estimate) | 0.5% (0.1-1.1%) |
| Bajema (CDC COVID-19 Response)[331] | 2020-07-31 to 2020-08-11 | Maryland | Regional | Moderate | 783 | 0-17 (n=6700); 18-49 (n=11237); 50-64 (n=10367); ≥65 (n=10408) | NR | Non-probability | Residual sera | 9.7% (7.3-12.9%) | Ortho-Clinical Diagnostics; Abbott; Roche \| Elecsys Anti-SARS-CoV-2 pan-immunoglobulin immunoassay, ARCHITECT SARS-CoV-2 IgG | 0.0%, 0.0% (Used author-adjusted estimate) | 9.7% (7.7-11.8%) |

| Article Author (Study Organization) | Sampling Dates (YMD) | Location | Geographic Scope | Overall Risk of Bias | Sample Size | Age | Female (%) | Sampling method | Sample frame | Uncorrected Seroprevalence (95% Confidence Interval) | Test Manufacturers \| Names \| Isotypes | Test Sens, Spec (Source) | Corrected Seroprevalence (95% Credible Interval) |
|---|---|---|---|---|---|---|---|---|---|---|---|---|---|
| | | | | | | | | | | | immunoassay, VIT- ROS SARS-CoV-2 IgG immunoassay \| IgG | | |
| Bajema (CDC COVID-19 Response)[331] | 2020-07-30 to 2020-08-11 | Michigan | Regional | Moderate | 982 | 0-17 (n=6700); 18-49 (n=11237); 50-64 (n=10367); ≥65 (n=10408) | NR | Non-probability | Residual sera | 3.4% (2.2-4.8%) | Ortho-Clinical Diagnostics; Abbott; Roche \| Elecsys Anti-SARS-CoV-2 pan-immunoglobulin immunoassay, ARCHITECT SARS-CoV-2 IgG immunoassay, VIT- ROS SARS-CoV-2 IgG immunoassay \| IgG | 0.0%, 0.0% (Used author-adjusted estimate) | 3.4% (2.4-4.6%) |
| Bajema (CDC COVID-19 Response)[331] | 2020-07-30 to 2020-08-11 | New Hampshire | Regional | Moderate | 809 | 0-17 (n=6700); 18-49 (n=11237); 50-64 (n=10367); ≥65 (n=10408) | NR | Non-probability | Residual sera | 0.8% (0.2-1.5%) | Ortho-Clinical Diagnostics; Abbott; Roche \| Elecsys Anti-SARS-CoV-2 pan-immunoglobulin immunoassay, ARCHITECT SARS-CoV-2 IgG immunoassay, VIT- ROS SARS-CoV-2 IgG immunoassay \| IgG | 0.0%, 0.0% (Used author-adjusted estimate) | 0.8% (0.3-1.5%) |
| Bajema (CDC COVID-19 Response)[331] | 2020-07-31 to 2020-08-11 | New Jersey | Regional | Moderate | 871 | 0-17 (n=6700); 18-49 (n=11237); 50-64 (n=10367); ≥65 (n=10408) | NR | Non-probability | Residual sera | 14.8% (12.2-17.6%) | Ortho-Clinical Diagnostics; Abbott; Roche \| Elecsys Anti-SARS-CoV-2 pan-immunoglobulin | 0.0%, 0.0% (Used author-adjusted estimate) | 14.8% (12.5-17.2%) |

| Article Author (Study Organization) | Sampling Dates (YMD) | Location | Geographic Scope | Overall Risk of Bias | Sample Size | Age | Female (%) | Sampling method | Sample frame | Uncorrected Seroprevalence (95% Confidence Interval) | Test Manufacturers \| Names \| Isotypes | Test Sens, Spec (Source) | Corrected Seroprevalence (95% Credible Interval) |
|---|---|---|---|---|---|---|---|---|---|---|---|---|---|
| | | | | | | | | | | | immunoassay, ARCHITECT SARS-CoV-2 IgG immunoassay, VIT- ROS SARS-CoV-2 IgG immunoassay \| IgG | | |
| Bajema (CDC COVID-19 Response)[331] | 2020-07-31 to 2020-08-11 | New York | Regional | Moderate | 846 | 0-17 (n=6700); 18-49 (n=11237); 50-64 (n=10367); ≥65 (n=10408) | NR | Non-probability | Residual sera | 23.3% (20.1-26.3%) | Ortho-Clinical Diagnostics; Abbott; Roche \| Elecsys Anti-SARS-CoV-2 pan-immunoglobulin immunoassay, ARCHITECT SARS-CoV-2 IgG immunoassay, VIT- ROS SARS-CoV-2 IgG immunoassay \| IgG | 0.0%, 0.0% (Used author-adjusted estimate) | 23.3% (20.5-26.2%) |
| Bajema (CDC COVID-19 Response)[331] | 2020-07-29 to 2020-08-11 | Ohio | Regional | Moderate | 786 | 0-17 (n=6700); 18-49 (n=11237); 50-64 (n=10367); ≥65 (n=10408) | NR | Non-probability | Residual sera | 2.3% (1.1-3.6%) | Ortho-Clinical Diagnostics; Abbott; Roche \| Elecsys Anti-SARS-CoV-2 pan-immunoglobulin immunoassay, ARCHITECT SARS-CoV-2 IgG immunoassay, VIT- ROS SARS-CoV-2 IgG immunoassay \| IgG | 0.0%, 0.0% (Used author-adjusted estimate) | 2.3% (1.4-3.5%) |
| Bajema (CDC COVID-19 Response)[331] | 2020-08-05 to 2020-08-11 | Oregon | Regional | Moderate | 658 | 0-17 (n=6700); 18-49 (n=11237); 50- | NR | Non-probability | Residual sera | 2.3% (1.0-3.8%) | Ortho-Clinical Diagnostics; Abbott; Roche \| Elecsys Anti- | 0.0%, 0.0% (Used author-adjusted estimate) | 2.3% (1.3-3.6%) |

| Article Author (Study Organization) | Sampling Dates (YMD) | Location | Geographic Scope | Overall Risk of Bias | Sample Size | Age | Female (%) | Sampling method | Sample frame | Uncorrected Seroprevalence (95% Confidence Interval) | Test Manufacturers \| Names \| Isotypes | Test Sens, Spec (Source) | Corrected Seroprevalence (95% Credible Interval) |
|---|---|---|---|---|---|---|---|---|---|---|---|---|---|
| | | | | | | 64 (n=10367); ≥65 (n=10408) | | | | | SARS-CoV-2 pan-immunoglobulin immunoassay, ARCHITECT SARS-CoV-2 IgG immunoassay, VIT- ROS SARS-CoV-2 IgG immunoassay \| IgG | | |
| Bajema (CDC COVID-19 Response)[331] | 2020-07-31 to 2020-08-11 | Pennsylvania | Regional | High | 575 | 0-17 (n=6700); 18-49 (n=11237); 50-64 (n=10367); ≥65 (n=10408) | NR | Non-probability | Residual sera | 10.2% (5.7-17.2%) | Ortho-Clinical Diagnostics; Abbott; Roche \| Elecsys Anti-SARS-CoV-2 pan-immunoglobulin immunoassay, ARCHITECT SARS-CoV-2 IgG immunoassay, VIT- ROS SARS-CoV-2 IgG immunoassay \| IgG | 0.0%, 0.0% (Used author-adjusted estimate) | 10.2% (7.8-12.7%) |
| Bajema (CDC COVID-19 Response)[331] | 2020-07-30 to 2020-08-11 | Rhode Island | Regional | Moderate | 684 | 0-17 (n=6700); 18-49 (n=11237); 50-64 (n=10367); ≥65 (n=10408) | NR | Non-probability | Residual sera | 3.0% (1.2-5.5%) | Ortho-Clinical Diagnostics; Abbott; Roche \| Elecsys Anti-SARS-CoV-2 pan-immunoglobulin immunoassay, ARCHITECT SARS-CoV-2 IgG immunoassay, VIT- ROS SARS-CoV-2 IgG immunoassay \| IgG | 0.0%, 0.0% (Used author-adjusted estimate) | 3.0% (1.9-4.4%) |

| Article Author (Study Organization) | Sampling Dates (YMD) | Location | Geographic Scope | Overall Risk of Bias | Sample Size | Age | Female (%) | Sampling method | Sample frame | Uncorrected Seroprevalence (95% Confidence Interval) | Test Manufacturers \| Names \| Isotypes | Test Sens, Spec (Source) | Corrected Seroprevalence (95% Credible Interval) |
|---|---|---|---|---|---|---|---|---|---|---|---|---|---|
| Bajema (CDC COVID-19 Response)[331] | 2020-07-30 to 2020-08-11 | Tennessee | Regional | Moderate | 1003 | 0-17 (n=6700); 18-49 (n=11237); 50-64 (n=10367); ≥65 (n=10408) | NR | Non-probability | Residual sera | 6.3% (4.4-8.2%) | Ortho-Clinical Diagnostics; Abbott; Roche \| Elecsys Anti-SARS-CoV-2 pan-immunoglobulin immunoassay, ARCHITECT SARS-CoV-2 IgG immunoassay, VIT- ROS SARS-CoV-2 IgG immunoassay \| IgG | 0.0%, 0.0% (Used author-adjusted estimate) | 6.3% (4.9-7.9%) |
| Bajema (CDC COVID-19 Response)[331] | 2020-07-30 to 2020-08-11 | Utah | Regional | Moderate | 880 | 0-17 (n=6700); 18-49 (n=11237); 50-64 (n=10367); ≥65 (n=10408) | NR | Non-probability | Residual sera | 3.2% (1.7-5.0%) | Ortho-Clinical Diagnostics; Abbott; Roche \| Elecsys Anti-SARS-CoV-2 pan-immunoglobulin immunoassay, ARCHITECT SARS-CoV-2 IgG immunoassay, VIT- ROS SARS-CoV-2 IgG immunoassay \| IgG | 0.0%, 0.0% (Used author-adjusted estimate) | 3.2% (2.2-4.5%) |
| Bajema (CDC COVID-19 Response)[331] | 2020-07-30 to 2020-08-11 | Vermont | Regional | High | 346 | 0-17 (n=6700); 18-49 (n=11237); 50-64 (n=10367); ≥65 (n=10408) | NR | Non-probability | Residual sera | 0.5% (0.0-1.8%) | Ortho-Clinical Diagnostics; Abbott; Roche \| Elecsys Anti-SARS-CoV-2 pan-immunoglobulin immunoassay, ARCHITECT SARS-CoV-2 IgG immunoassay, VIT- ROS SARS-CoV-2 IgG | 0.0%, 0.0% (Used author-adjusted estimate) | 0.5% (0.0-1.3%) |

| Article Author (Study Organization) | Sampling Dates (YMD) | Location | Geographic Scope | Overall Risk of Bias | Sample Size | Age | Female (%) | Sampling method | Sample frame | Uncorrected Seroprevalence (95% Confidence Interval) | Test Manufacturers \| Names \| Isotypes | Test Sens, Spec (Source) | Corrected Seroprevalence (95% Credible Interval) |
|---|---|---|---|---|---|---|---|---|---|---|---|---|---|
| | | | | | | | | | | | immunoassay \| IgG | | |
| Bajema (CDC COVID-19 Response)[331] | 2020-07-31 to 2020-08-11 | Virginia | Regional | Moderate | 730 | 0-17 (n=6700); 18-49 (n=11237); 50-64 (n=10367); ≥65 (n=10408) | NR | Non-probability | Residual sera | 4.1% (1.8-6.9%) | Ortho-Clinical Diagnostics; Abbott; Roche \| Elecsys Anti-SARS-CoV-2 pan-immunoglobulin immunoassay, ARCHITECT SARS-CoV-2 IgG immunoassay, VIT- ROS SARS-CoV-2 IgG immunoassay \| IgG | 0.0%, 0.0% (Used author-adjusted estimate) | 4.1% (2.7-5.6%) |
| Bajema (CDC COVID-19 Response)[331] | 2020-07-29 to 2020-08-11 | Washington | Regional | Moderate | 684 | 0-17 (n=6700); 18-49 (n=11237); 50-64 (n=10367); ≥65 (n=10408) | NR | Non-probability | Residual sera | 2.1% (0.8-3.7%) | Ortho-Clinical Diagnostics; Abbott; Roche \| Elecsys Anti-SARS-CoV-2 pan-immunoglobulin immunoassay, ARCHITECT SARS-CoV-2 IgG immunoassay, VIT- ROS SARS-CoV-2 IgG immunoassay \| IgG | 0.0%, 0.0% (Used author-adjusted estimate) | 2.1% (1.2-3.3%) |
| Bajema (CDC COVID-19 Response)[331] | 2020-07-29 to 2020-08-10 | Illinois | Regional | Moderate | 1004 | 0-17 (n=6700); 18-49 (n=11237); 50-64 (n=10367); ≥65 (n=10408) | NR | Non-probability | Residual sera | 3.9% (2.5-5.2%) | Ortho-Clinical Diagnostics; Abbott; Roche \| Elecsys Anti-SARS-CoV-2 pan-immunoglobulin immunoassay, ARCHITECT SARS-CoV-2 IgG | 0.0%, 0.0% (Used author-adjusted estimate) | 3.9% (2.8-5.2%) |

| Article Author (Study Organization) | Sampling Dates (YMD) | Location | Geographic Scope | Overall Risk of Bias | Sample Size | Age | Female (%) | Sampling method | Sample frame | Uncorrected Seroprevalence (95% Confidence Interval) | Test Manufacturers \| Names \| Isotypes | Test Sens, Spec (Source) | Corrected Seroprevalence (95% Credible Interval) |
|---|---|---|---|---|---|---|---|---|---|---|---|---|---|
| | | | | | | | | | | | immunoassay, VIT- ROS SARS-CoV-2 IgG immunoassay \| IgG | | |
| Bajema (CDC COVID-19 Response)[331] | 2020-07-30 to 2020-08-10 | Massachusetts | Regional | Moderate | 978 | 0-17 (n=6700); 18-49 (n=11237); 50-64 (n=10367); ≥65 (n=10408) | NR | Non-probability | Residual sera | 4.2% (2.9-5.8%) | Ortho-Clinical Diagnostics; Abbott; Roche \| Elecsys Anti-SARS-CoV-2 pan-immunoglobulin immunoassay, ARCHITECT SARS-CoV-2 IgG immunoassay, VIT- ROS SARS-CoV-2 IgG immunoassay \| IgG | 0.0%, 0.0% (Used author-adjusted estimate) | 4.2% (3.1-5.6%) |
| Bajema (CDC COVID-19 Response)[331] | 2020-07-28 to 2020-08-10 | Missouri | Regional | Moderate | 979 | 0-17 (n=6700); 18-49 (n=11237); 50-64 (n=10367); ≥65 (n=10408) | NR | Non-probability | Residual sera | 2.5% (1.4-3.6%) | Ortho-Clinical Diagnostics; Abbott; Roche \| Elecsys Anti-SARS-CoV-2 pan-immunoglobulin immunoassay, ARCHITECT SARS-CoV-2 IgG immunoassay, VIT- ROS SARS-CoV-2 IgG immunoassay \| IgG | 0.0%, 0.0% (Used author-adjusted estimate) | 2.5% (1.6-3.6%) |
| Bajema (CDC COVID-19 Response)[331] | 2020-07-29 to 2020-08-10 | Montana | Regional | High | 199 | 0-17 (n=6700); 18-49 (n=11237); 50-64 (n=10367); ≥65 (n=10408) | NR | Non-probability | Residual sera | 0.5% (0.0-1.7%) | Ortho-Clinical Diagnostics; Abbott; Roche \| Elecsys Anti-SARS-CoV-2 pan-immunoglobulin | 0.0%, 0.0% (Used author-adjusted estimate) | 0.5% (0.0-1.3%) |

| Article Author (Study Organization) | Sampling Dates (YMD) | Location | Geographic Scope | Overall Risk of Bias | Sample Size | Age | Female (%) | Sampling method | Sample frame | Uncorrected Seroprevalence (95% Confidence Interval) | Test Manufacturers \| Names \| Isotypes | Test Sens, Spec (Source) | Corrected Seroprevalence (95% Credible Interval) |
|---|---|---|---|---|---|---|---|---|---|---|---|---|---|
| | | | | | | | | | | | immunoassay, ARCHITECT SARS-CoV-2 IgG immunoassay, VIT- ROS SARS-CoV-2 IgG immunoassay \| IgG | | |
| Bajema (CDC COVID-19 Response)[331] | 2020-07-29 to 2020-08-10 | North Carolina | Regional | Moderate | 928 | 0-17 (n=6700); 18-49 (n=11237); 50-64 (n=10367); ≥65 (n=10408) | NR | Non-probability | Residual sera | 2.5% (1.3-3.7%) | Ortho-Clinical Diagnostics; Abbott; Roche \| Elecsys Anti-SARS-CoV-2 pan-immunoglobulin immunoassay, ARCHITECT SARS-CoV-2 IgG immunoassay, VIT- ROS SARS-CoV-2 IgG immunoassay \| IgG | 0.0%, 0.0% (Used author-adjusted estimate) | 2.5% (1.6-3.6%) |
| Bajema (CDC COVID-19 Response)[331] | 2020-07-29 to 2020-08-08 | Kansas | Regional | Moderate | 962 | 0-17 (n=6700); 18-49 (n=11237); 50-64 (n=10367); ≥65 (n=10408) | NR | Non-probability | Residual sera | 1.6% (0.7-2.8%) | Ortho-Clinical Diagnostics; Abbott; Roche \| Elecsys Anti-SARS-CoV-2 pan-immunoglobulin immunoassay, ARCHITECT SARS-CoV-2 IgG immunoassay, VIT- ROS SARS-CoV-2 IgG immunoassay \| IgG | 0.0%, 0.0% (Used author-adjusted estimate) | 1.6% (0.9-2.5%) |
| Bajema (CDC COVID-19 Response)[331] | 2020-07-30 to 2020-08-07 | Colorado | Regional | Moderate | 1001 | 0-17 (n=6700); 18-49 (n=11237); 50- | NR | Non-probability | Residual sera | 2.4% (0.9-4.5%) | Ortho-Clinical Diagnostics; Abbott; Roche \| Elecsys Anti- | 0.0%, 0.0% (Used author-adjusted estimate) | 2.4% (1.6-3.5%) |

| Article Author (Study Organization) | Sampling Dates (YMD) | Location | Geographic Scope | Overall Risk of Bias | Sample Size | Age | Female (%) | Sampling method | Sample frame | Uncorrected Seroprevalence (95% Confidence Interval) | Test Manufacturers \| Names \| Isotypes | Test Sens, Spec (Source) | Corrected Seroprevalence (95% Credible Interval) |
|---|---|---|---|---|---|---|---|---|---|---|---|---|---|
| | | | | | | 64 (n=10367); ≥65 (n=10408) | | | | | SARS-CoV-2 pan-immunoglobulin immunoassay, ARCHITECT SARS-CoV-2 IgG immunoassay, VIT- ROS SARS-CoV-2 IgG immunoassay \| IgG | | |
| Bajema (CDC COVID-19 Response)[331] | 2020-07-27 to 2020-08-07 | Puerto Rico | Regional | Moderate | 984 | 0-17 (n=6700); 18-49 (n=11237); 50-64 (n=10367); ≥65 (n=10408) | NR | Non-probability | Residual sera | 1.1% (0.4-1.8%) | Ortho-Clinical Diagnostics; Abbott; Roche \| Elecsys Anti-SARS-CoV-2 pan-immunoglobulin immunoassay, ARCHITECT SARS-CoV-2 IgG immunoassay, VIT- ROS SARS-CoV-2 IgG immunoassay \| IgG | 0.0%, 0.0% (Used author-adjusted estimate) | 1.1% (0.5-1.8%) |
| Bajema (CDC COVID-19 Response)[331] | 2020-07-30 to 2020-08-05 | California | Regional | Moderate | 980 | 0-17 (n=6700); 18-49 (n=11237); 50-64 (n=10367); ≥65 (n=10408) | NR | Non-probability | Residual sera | 5.7% (4.0-7.8%) | Ortho-Clinical Diagnostics; Abbott; Roche \| Elecsys Anti-SARS-CoV-2 pan-immunoglobulin immunoassay, ARCHITECT SARS-CoV-2 IgG immunoassay, VIT- ROS SARS-CoV-2 IgG immunoassay \| IgG | 0.0%, 0.0% (Used author-adjusted estimate) | 5.7% (4.3-7.2%) |

| Article Author (Study Organization) | Sampling Dates (YMD) | Location | Geographic Scope | Overall Risk of Bias | Sample Size | Age | Female (%) | Sampling method | Sample frame | Uncorrected Seroprevalence (95% Confidence Interval) | Test Manufacturers \| Names \| Isotypes | Test Sens, Spec (Source) | Corrected Seroprevalence (95% Credible Interval) |
|---|---|---|---|---|---|---|---|---|---|---|---|---|---|
| Bajema (CDC COVID-19 Response)[331] | 2020-07-29 to 2020-08-05 | Texas | Regional | Moderate | 986 | 0-17 (n=6700); 18-49 (n=11237); 50-64 (n=10367); ≥65 (n=10408) | NR | Non-probability | Residual sera | 5.9% (4.0-8.0%) | Ortho-Clinical Diagnostics; Abbott; Roche \| Elecsys Anti-SARS-CoV-2 pan-immunoglobulin immunoassay, ARCHITECT SARS-CoV-2 IgG immunoassay, VIT- ROS SARS-CoV-2 IgG immunoassay \| IgG | 0.0%, 0.0% (Used author-adjusted estimate) | 5.9% (4.5-7.5%) |
| Bajema (CDC COVID-19 Response)[331] | 2020-07-28 to 2020-08-04 | Oklahoma | Regional | Moderate | 979 | 0-17 (n=6700); 18-49 (n=11237); 50-64 (n=10367); ≥65 (n=10408) | NR | Non-probability | Residual sera | 1.6% (0.9-2.5%) | Ortho-Clinical Diagnostics; Abbott; Roche \| Elecsys Anti-SARS-CoV-2 pan-immunoglobulin immunoassay, ARCHITECT SARS-CoV-2 IgG immunoassay, VIT- ROS SARS-CoV-2 IgG immunoassay \| IgG | 0.0%, 0.0% (Used author-adjusted estimate) | 1.6% (0.9-2.5%) |
| Bajema (CDC COVID-19 Response)[331] | 2020-07-30 to 2020-08-03 | Connecticut | Regional | Moderate | 994 | 0-17 (n=6700); 18-49 (n=11237); 50-64 (n=10367); ≥65 (n=10408) | NR | Non-probability | Residual sera | 3.4% (2.0-4.7%) | Ortho-Clinical Diagnostics; Abbott; Roche \| Elecsys Anti-SARS-CoV-2 pan-immunoglobulin immunoassay, ARCHITECT SARS-CoV-2 IgG immunoassay, VIT- ROS SARS-CoV-2 IgG | 0.0%, 0.0% (Used author-adjusted estimate) | 3.4% (2.3-4.6%) |

| Article Author (Study Organization) | Sampling Dates (YMD) | Location | Geographic Scope | Overall Risk of Bias | Sample Size | Age | Female (%) | Sampling method | Sample frame | Uncorrected Seroprevalence (95% Confidence Interval) | Test Manufacturers \| Names \| Isotypes | Test Sens, Spec (Source) | Corrected Seroprevalence (95% Credible Interval) |
|---|---|---|---|---|---|---|---|---|---|---|---|---|---|
| | | | | | | | | | | | immunoassay \| IgG | | |
| Bajema (CDC COVID-19 Response)[331] | 2020-07-31 to 2020-08-03 | Florida | Regional | Moderate | 980 | 0-17 (n=6700); 18-49 (n=11237); 50-64 (n=10367); ≥65 (n=10408) | NR | Non-probability | Residual sera | 4.3% (2.8-5.9%) | Ortho-Clinical Diagnostics; Abbott; Roche \| Elecsys Anti-SARS-CoV-2 pan-immunoglobulin immunoassay, ARCHITECT SARS-CoV-2 IgG immunoassay, VIT- ROS SARS-CoV-2 IgG immunoassay \| IgG | 0.0%, 0.0% (Used author-adjusted estimate) | 4.3% (3.1-5.7%) |
| Bajema (CDC COVID-19 Response)[331] | 2020-07-30 to 2020-08-02 | Nevada | Regional | Moderate | 979 | 0-17 (n=6700); 18-49 (n=11237); 50-64 (n=10367); ≥65 (n=10408) | NR | Non-probability | Residual sera | 5.1% (3.6-6.7%) | Ortho-Clinical Diagnostics; Abbott; Roche \| Elecsys Anti-SARS-CoV-2 pan-immunoglobulin immunoassay, ARCHITECT SARS-CoV-2 IgG immunoassay, VIT- ROS SARS-CoV-2 IgG immunoassay \| IgG | 0.0%, 0.0% (Used author-adjusted estimate) | 5.1% (3.8-6.5%) |
| Feehan (Louisiana State University)[341] | 2020-07-15 to 2020-07-31 | Louisiana \| Baton Rouge | Local | Moderate | 2138 | Mean 48.7 (Range 18-91) | 63.6 | Probability | Household and community samples | 3.6% (2.8-4.4%) | Abbott \| Abbott ARCHITECT i2000SR \| IgG | 0.4%, 1.0% (FINDDx / MUHC independent evaluation) | - |
| Vassallo (Vitalant)[342] | 2020-06-01 to 2020-07-31 | Colorado, Mississippi, North Dakota, Nevada, | National | Moderate | 252882 | 16-17 (n=1812); 18-29 (n=22048); 30-39 (n=30956); 40- | 39.4 | Non-probability | Blood donors | 1.8% | Ortho-Clinical Diagnostics \| VITROS Anti-SARS-CoV-2 Total Reagent | 0.9%, 1.0% (FINDDx / MUHC independent evaluation) | - |

| Article Author (Study Organization) | Sampling Dates (YMD) | Location | Geographic Scope | Overall Risk of Bias | Sample Size | Age | Female (%) | Sampling method | Sample frame | Uncorrected Seroprevalence (95% Confidence Interval) | Test Manufacturers \| Names \| Isotypes | Test Sens, Spec (Source) | Corrected Seroprevalence (95% Credible Interval) |
|---|---|---|---|---|---|---|---|---|---|---|---|---|---|
| | | Louisiana, Tennessee, Washington, Wyoming, California, Montana, South Dakota, New Mexico, Texas, Arizona \| Reno, Las Vegas, Cheyenne, Phoenix, Lubbock, Billings, Albuquerque, Rapid City, Tupelo, San Francisco, Spokane, McAllen, El Paso, Fargo, Newark, ventura, Memphis, Lafayette, Sacramento, Denver | | | | 49 (n=34151); 50-64 (n=66030); 65-74 (n=28453); 75-84 (n=5848); >85 (n=357) | | | | | Pack \| IgG, IgA, IgM | | |
| Spencer (American Red Cross)[345] | 2020-06-15 to 2020-07-27 | | National | High | 573586 | =>18 | NR | Non-probability | Blood donors | 1.5% | Ortho \| Ortho Anti-SARS-CoV2 Total Ig Test \| IgG | 0.8%, 1.0% (FINDDx / MUHC independent evaluation) | - |
| Yamaki (St. Jude Medical Center)[346] | 2020-07-15 to 2020-07-27 | California \| Fullerton | Local | High | 865 | 10-19 (N=9), 20-29 (n=55), 30-39 (n=81), 40-49 (n=83), 50-59 (n=133), 60-69 (n=159), 70-79 (n=164), 80-89 (n=144), 90-99 (n=35), 100-109 (n=2) | 55.3 | Non-probability | Residual sera | 9.4% (8.0-12.0%) | Abbott Laboratories \| Abbott Architect \| IgG | 0.4%, 1.0% (FINDDx / MUHC independent evaluation) | - |

| Article Author (Study Organization) | Sampling Dates (YMD) | Location | Geographic Scope | Overall Risk of Bias | Sample Size | Age | Female (%) | Sampling method | Sample frame | Uncorrected Seroprevalence (95% Confidence Interval) | Test Manufacturers \| Names \| Isotypes | Test Sens, Spec (Source) | Corrected Seroprevalence (95% Credible Interval) |
|---|---|---|---|---|---|---|---|---|---|---|---|---|---|
| Mahajan (Yale School of Medicine)[349] | 2020-06-23 to 2020-07-22 | Connecticut | Regional | High | 148 | Mean 46.4 (SD 13.0) | 64.2 | Probability | Household and community samples | 6.4% (0.9-11.9%) | Ortho-Clinical Diagnostics \| Ortho-Diagnostics VITROS anti-SARS-CoV-2 IgG assay \| IgG | 0.8%, 1.0% (FINDDx / MUHC independent evaluation) | - |
| Mahajan (Yale School of Medicine)[349] | 2020-06-23 to 2020-07-22 | Connecticut | Regional | High | 171 | Mean 39.9 (SD 15.5) | 60.2 | Probability | Household and community samples | 19.9% (13.2-26.6%) | Ortho-Clinical Diagnostics \| Ortho-Diagnostics VITROS anti-SARS-CoV-2 IgG assay \| IgG | 0.8%, 1.0% (FINDDx / MUHC independent evaluation) | - |
| McMillan (Riverside University Health System)[350] | 2020-07-06 to 2020-07-20 | California | Local | Unclear | 1726 | NR | NR | Probability | Household and community samples | 5.9% | NR \| NR \| NR | No data | - |
| Zauzmer (Washington DC Public Health)[352] | 2020-06-15 to 2020-07-15 | Washington D.C \| Washington DC | Local | High | 13706 | NR | NR | Non-probability | Household and community samples | 5.9% | DiaSorin LIAISON; Abbot Alinity i \| NR \| NR | 97.8%, 97.8% (Used Bastos SR/MA data; no sens, spec, or author adjustment available) | 3.8% (3.2-4.6%) |
| Jin (New York Blood Centre)[353] | 2020-06-15 to 2020-07-15 | New York \| New York City | Local | High | 1000 | Median 48 (95% CI 46-49) | 38.5 | Non-probability | Blood donors | 12.8% | Ortho Clinical Diagnostics; Abbott Labs \| VITROS™ SARS-CoV-2 Total Ig assay, Abbott Labs Architect SARS-CoV-2 IgG \| IgG | 97.8%, 97.8% (Used Bastos SR/MA data; no sens, spec, or author adjustment available) | 11.2% (8.9-13.5%) |
| Kaufman (Quest Diagnostics)[355] | 2020-04-21 to 2020-07-10 | | National | High | 2402282 | NR | NR | Non-probability | Household and community samples | 11.8% | Ortho-Clinical Diagnostics; EUROIMMUN; Abbott Laboratories \| Architect, VITROS, EUROIMMUN Anti-SARS-CoV-2 ELISA (IgG) \| IgG | No data | - |

| Article Author (Study Organization) | Sampling Dates (YMD) | Location | Geographic Scope | Overall Risk of Bias | Sample Size | Age | Female (%) | Sampling method | Sample frame | Uncorrected Seroprevalence (95% Confidence Interval) | Test Manufacturers \| Names \| Isotypes | Test Sens, Spec (Source) | Corrected Seroprevalence (95% Credible Interval) |
|---|---|---|---|---|---|---|---|---|---|---|---|---|---|
| Kamath (Serimmune)[359] | 2020-03-01 to 2020-07-07 | New York \| New York City | Local | High | 1559 | 17-25 (n=395, 25%); 26-35 (n=200, 13%); 36-45 (n=164, 11%); 46-55 (n=206, 13%); 56-65 (n=370, 24%); 66-80 (n=224, 14%). | 48.4 | Non-probability | Blood donors | 4.4% | Serimmune \| Serum Epitope Repertoire Analysis (SERA) \| IgG | 91.0%, 99.3% (Test developer / manufacturer evaluation) | 4.0% (2.7-5.2%) |
| Childs (Oxnard Fire Department)[360] | 2020-06-22 to 2020-07-06 | California | Local | High | 6794 | 13+ | 60.7 | Non-probability | Household and community samples | 1.3% | 1) Abbott 2) Bio-Rad \| NR \| NR | No data | - |
| Stadlbauer (Icahn School of Medicine at Mount Sinai)[361] | 2020-02-02 to 2020-07-05 | New York \| New York City | Local | High | 6590 | 0-20 (n=358); 21-40 (n=2808); 41-60 (n=1381); >=61 (n=2043) | 67.6 | Non-probability | Residual sera | 11.1% | Mount Sinai Hospital New York \| N/A - Author-designed \| IgG | 95.0%, 100.0% (Test developer / manufacturer evaluation) | 11.0% (7.9-13.2%) |
| Stadlbauer (Icahn School of Medicine at Mount Sinai)[361] | 2020-02-02 to 2020-07-05 | New York \| New York City | Local | High | 4101 | 0-20 (n=115); 21-40 (n=616); 41-60 (n=1185); >=61 (n=2185) | 45.5 | Non-probability | Residual sera | 26.0% | Mount Sinai Hospital New York \| N/A - Author-designed \| IgG | 95.0%, 100.0% (Test developer / manufacturer evaluation) | 27.0% (23.1-30.7%) |
| Walker (DaVita Clinical Research)[363] | 2020-05-27 to 2020-07-01 | | National | Moderate | 4297 | <50 (n=2398), 50-59 (n=2669), 60-69 (n=3568), 70-79 (n=2873), 80+ (n=1424) | 125.5 | Probability | Residual sera | 4.8% (4.1-5.4%) | Diazyme Laboratories; Inc \| Indirect chemiluminescence immunoassays for anti-SARS-CoV2 IgG antibodies \| IgG | 97.8%, 97.8% (Used Bastos SR/MA data; no sens, spec, or author adjustment available) | 2.7% (2.2-3.2%) |
| Viglienzoni (University of Vermont)[367] | Until 2020-06-30 | Vermont | Local | High | 455 | NR | NR | Non-probability | Household and community samples | 2.2% | University of Vermont \| NR \| NR | No data | - |
| Samore (University of Utah)[368] | 2020-05-04 to 2020-06-30 | Utah | Regional | Moderate | 8108 | Median 44 (IQR 30-62) | 53.5 | Probability | Household and community samples | 0.8% (0.1-1.6%) | Abbott \| Abbott SARS-CoV-2 IgG \| IgG | 86.4%, 99.6% (Used author-adjusted estimate) | 0.8% (0.6-1.0%) |
| Mahajan (Yale School of Medicine)[349] | 2020-06-04 to 2020-06-23 | Connecticut | Regional | Moderate | 567 | Mean 50.1 (SD 17.2) | 57.0 | Probability | Household and community samples | 4.0% (2.0-6.0%) | Ortho-Clinical Diagnostics \| Ortho-Diagnostics | 0.8%, 1.0% (FINDDx / MUHC | - |

| Article Author (Study Organization) | Sampling Dates (YMD) | Location | Geographic Scope | Overall Risk of Bias | Sample Size | Age | Female (%) | Sampling method | Sample frame | Uncorrected Seroprevalence (95% Confidence Interval) | Test Manufacturers \| Names \| Isotypes | Test Sens, Spec (Source) | Corrected Seroprevalence (95% Credible Interval) |
|---|---|---|---|---|---|---|---|---|---|---|---|---|---|
| | | | | | | | | | | | VITROS anti-SARS-CoV-2 IgG assay \| IgG | independent evaluation) | |
| Barzin (University of North Carolina Chapel Hill)[374] | 2020-04-28 to 2020-06-19 | North Carolina | Regional | Moderate | 2973 | 20-29 (n=342), 30-39 (n=599), 40-49 (n=518), 50-59 (n=602), 60-69 (n=489), 70-79 (n=310) 80+ (n=77) | 65.8 | Non-probability | Residual sera | 0.0% (0.0-0.9%) | Abbott Laboratories \| Abbott SARS-CoV-2 IgG assay \| IgG | 100.0%, 98.9% (Used author-adjusted estimate) | 0.0% (0.0-0.1%) |
| Spectrum News Staff ( University of Louisville)[375] | 2020-06-10 to 2020-06-19 | Kentucky | Local | High | 2237 | 18-34 (n=470); 35-59 (n=895); =>60 (n= 895) | NR | Non-probability | Household and community samples | 4.0% | NR \| NR \| NR | No data | - |
| Dodd (American Red Cross Scientific Affairs)[377] | 2020-06-15 to 2020-06-15 | | National | Moderate | 953926 | 16-17 (n=8375); 18-24 (n=51763); 25-39 (n=204407); 40-54 (n=262912); 55+ (n=426469) | 55.0 | Non-probability | Blood donors | 1.8% | Ortho-Clinical Diagnostics \| Ortho VITROS anti-SARS-CoV-2 S1 Total Ig assay \| NR | 0.9%, 1.0% (FINDDx / MUHC independent evaluation) | - |
| Reifer (Sherman Abrams Laboratory )[378] | 2020-05-15 to 2020-06-15 | New York \| New York | Local | High | 28523 | >=0 | NR | Non-probability | Residual sera | 44.0% | DiaSorin LIAISON \| Liaison® SARS- CoV-2 S1/S2 IgG \| IgG | 0.8%, 1.0% (FINDDx / MUHC independent evaluation) | - |
| Sutton (Oregon Health Authority)[379] | 2020-05-11 to 2020-06-15 | Oregon | Regional | High | 897 | 0-4 (n=5); 5-17 (n=24); 18-49 (n=274); 50-64 (n=211); 65-74 (n=178); 75-84 (n=144); >=85 (n=61) | NR | Probability | Residual sera | 1.0% (0.2-1.8%) | Abbott \| Abbott Architect SARS-CoV-2 IgG assay \| IgG | 0.4%, 1.0% (FINDDx / MUHC independent evaluation) | - |
| Anand (Stanford University)[381] | 2020-06-15 to 2020-06-15 | | National | Moderate | 28503 | 18-44 (n=3303); 45-64 (n=11541); 65-79 (n=10220); 80+ (n=3439) | 42.6 | Probability | Residual sera | 9.3% (8.8-9.9%) | Siemens Healthineers \| SARS-CoV-2 spike protein receptor binding domain (S1RBD) total antibody | 0.9%, 1.0% (FINDDx / MUHC independent evaluation) | - |

| Article Author (Study Organization) | Sampling Dates (YMD) | Location | Geographic Scope | Overall Risk of Bias | Sample Size | Age | Female (%) | Sampling method | Sample frame | Uncorrected Seroprevalence (95% Confidence Interval) | Test Manufacturers \| Names \| Isotypes | Test Sens, Spec (Source) | Corrected Seroprevalence (95% Credible Interval) |
|---|---|---|---|---|---|---|---|---|---|---|---|---|---|
| | | | | | | | | | | | chemiluminescence assay \| NR | | |
| Hynes (Washoe County Health District)[384] | 2020-06-09 to 2020-06-10 | Nevada | Local | High | 234 | NR | NR | Probability | Household and community samples | 2.3% | Abbott \| NR \| NR | No data | - |
| Harden (Routt County Board of Health)[385] | Until 2020-06-10 | Colorado \| Steamboat Springs | Local | High | 649 | NR | NR | Non-probability | Household and community samples | 2.6% | NR \| NR \| NR | No data | - |
| Rudavsky (Indiana University Fairbanks School of Public Health)[388] | 2020-06-03 to 2020-06-08 | Indiana | Regional | Moderate | 3600 | NR | NR | Probability | Household and community samples | 1.5% | NR \| NR \| NR | No data | - |
| Barzin (University of North Carolina Chapel Hill)[374] | 2020-03-03 to 2020-06-04 | North Carolina | Regional | Moderate | 1449 | =>18 | NR | Non-probability | Residual sera | 0.7% | Abbott Laboratories \| Abbott SARS-CoV-2 IgG assay \| IgG | 0.4%, 1.0% (FINDDx / MUHC independent evaluation) | - |
| Ohnesorge (Psalms/Solid Rock Medicine Group)[393] | 2020-05-29 to 2020-05-29 | North Carolina | Local | Unclear | 329 | NR | NR | Non-probability | Household and community samples | 1.5% | NR \| NR \| NR | No data | - |
| Hayden (Desert Healthcare District)[395] | Until 2020-05-28 | California | Local | Unclear | 896 | NR | 50.0 | Non-probability | Household and community samples | 6.0% | Phamatech \| NR \| IgG, IgM | 0.9%, 0.9% (FINDDx / MUHC independent evaluation) | - |
| Liu (Yale School of Medicine)[396] | 2020-02-19 to 2020-05-26 | Connecticut \| New Haven | Sublocal | High | 3048 | NR | NR | Non-probability | Residual sera | 5.9% | In-house \| In-house ELISA test \| IgG | 75.6%, 97.6% (Used Bastos SR/MA data; no sens, spec, or author adjustment available) | 4.6% (2.9-6.3%) |
| Osher (BioSolutions | 2020-04-22 to 2020-05-26 | Illinois \| Chicago | Sublocal | High | 113 | 3 to 84 | NR | Non-probability | Residual sera | 15.9% | CLUNGENE \| CLUNGENE SARS-COV-2 | 66.0%, 96.6% (Used Bastos SR/MA data; | 19.3% (8.9-29.6%) |

| Article Author (Study Organization) | Sampling Dates (YMD) | Location | Geographic Scope | Overall Risk of Bias | Sample Size | Age | Female (%) | Sampling method | Sample frame | Uncorrected Seroprevalence (95% Confidence Interval) | Test Manufacturers \| Names \| Isotypes | Test Sens, Spec (Source) | Corrected Seroprevalence (95% Credible Interval) |
|---|---|---|---|---|---|---|---|---|---|---|---|---|---|
| Services LLC)[397] | | | | | | | | | | | VIRUS (COVID-19) IgG/IgM Rapid Test Cassette lateral flow immunoassay (LFI). \| IgG, IgM | no sens, spec, or author adjustment available) | |
| Mukherjee (Humboldt County Department of Health and Human Services)[398] | Until 2020-05-26 | California | Local | High | 1000 | NR | NR | Non-probability | Household and community samples | 1.0% | NR \| NR \| NR | No data | - |
| Alfini (UW Health)[407] | 2020-05-13 to 2020-05-20 | Wisconsin \| Madison | Local | Unclear | 433 | NR | NR | Non-probability | Household and community samples | 0.3% | NR \| NR \| NR | No data | - |
| Alfini (UW Health)[407] | 2020-05-13 to 2020-05-20 | Wisconsin \| Madison | Local | Unclear | 5303 | NR | NR | Non-probability | Household and community samples | 1.5% | NR \| NR \| NR | No data | - |
| Montgomery (Iowa Department of Public Health)[408] | 2021-04-01 to 2020-05-20 | Iowa | Regional | High | 18160 | NR | NR | Non-probability | Household and community samples | 10.0% | NR \| NR \| IgG | No data | - |
| McLaughlin ( University of Washington)[412] | 2020-05-04 to 2020-05-19 | Idaho | Local | Moderate | 917 | 18-29 (n=96); 30-39 (n=151); 40-49 (n=186); 50-59 (n=225); 60-69 (n=172); >=70 (n=87); | 52.2 | Non-probability | Household and community samples | 23.1% (19.8-26.5%) | Abbott \| Abbott Architect SARS-CoV-2 IgG assay \| IgG | 92.9%, 99.6% (Used author-adjusted estimate) | 23.1% (20.4-25.8%) |
| Feehan (Oschner Clinic Foundation)[414] | 2020-05-09 to 2020-05-15 | Louisiana | Local | Moderate | 2640 | Mean 50.6 (NR) | 63.5 | Probability | Household and community samples | 6.9% (6.8-6.9%) | Abbott \| Abbott Architect SARS-CoV-2 IgG assay \| IgG | 0.4%, 1.0% (FINDDx / MUHC independent evaluation) | - |
| Meyer (Winona Health)[415] | 2020-05-10 to 2020-05-15 | Minnesota \| Winona | Local | High | 1000 | NR | NR | Non-probability | Household and community samples | 7.0% | NR \| NR \| IgG | No data | - |

| Article Author (Study Organization) | Sampling Dates (YMD) | Location | Geographic Scope | Overall Risk of Bias | Sample Size | Age | Female (%) | Sampling method | Sample frame | Uncorrected Seroprevalence (95% Confidence Interval) | Test Manufacturers \| Names \| Isotypes | Test Sens, Spec (Source) | Corrected Seroprevalence (95% Credible Interval) |
|---|---|---|---|---|---|---|---|---|---|---|---|---|---|
| Dietrich (Tulane University)[416] | 2020-03-18 to 2020-05-15 | Louisiana \| Orleans | Local | High | 812 | Median 11.0 (IQR 4-15) | 50.4 | Non-probability | Residual sera | 7.6% | Author-designed \| N/A - Author-designed \| IgG | 75.6%, 97.6% (Used Bastos SR/MA data; no sens, spec, or author adjustment available) | 6.9% (4.1-9.6%) |
| City of Boston (Boston Public Health Commission )[417] | Until 2020-05-15 | Massachusetts \| Boston | Local | Moderate | 750 | Median 42.4 (NR) | 61.6 | Probability | Household and community samples | 9.9% | NR \| NR \| NR | No data | - |
| CBS Los Angeles (University of Southern California)[420] | 2020-05-08 to 2020-05-12 | California | Local | Unclear | 1014 | NR | NR | Non-probability | Household and community samples | 2.1% | NR \| NR \| NR | No data | - |
| Nesbitt (New York Blood Center)[421] | 2020-04-27 to 2020-05-11 | Rhode Island | Regional | Moderate | 1996 | Median 56 (NR) | 47.3 | Non-probability | Blood donors | 0.7% | Ortho-Clinical Diagnostics \| Ortho-Diagnostics VITROS anti-SARS-CoV-2 IgG assay \| IgG | 0.8%, 1.0% (FINDDx / MUHC independent evaluation) | - |
| Ripperger (University of Arizona)[424] | 2020-04-30 to 2020-05-07 | Arizona | Local | High | 5882 | Median NR40 (Range 18-85) | 52.4 | Non-probability | Household and community samples | 1.2% | NR \| NR \| IgG, IgM | 97.6%, 100.0% (Test developer / manufacturer evaluation) | 1.0% (0.2-1.5%) |
| Bajema (Centers for Disease Control and Prevention)[428] | 2020-04-28 to 2020-05-03 | Georgia \| Atlanta | Local | Low | 669 | Median 46 (IQR 36-73) | 54.9 | Probability | Household and community samples | 3.6% | Centers for Disease Control and Prevention (CDC) \| CDC SARS-CoV-2 ELISA \| NR | 96.0%, 99.3% (Test developer / manufacturer evaluation) | 2.1% (0.0-4.2%) |
| Biggs (CDC COVID-19 Response Team)[429] | 2020-04-28 to 2020-05-03 | Georgia | Local | Moderate | 696 | NR | 54.2 | Probability | Household and community samples | 2.7% | Ortho-Clinical Diagnostics \| VITROS anti-SARS-CoV-2 total antibody \| IgG, IgA, IgM | 0.9%, 1.0% (FINDDx / MUHC independent evaluation) | - |
| Menachemi (Indiana University | 2020-05-02 to 2020-05-03 | Indiana \| Indianapolis | Local | High | 898 | <20 (n=77); 20-39 (n=277); 40-59 (n=369); | 58.2 | Non-probability | Household and | 5.8% | Author derived \| NR \| IgG | 100.0%, 99.6% (Test developer | 4.1% (0.4-6.5%) |

| Article Author (Study Organization) | Sampling Dates (YMD) | Location | Geographic Scope | Overall Risk of Bias | Sample Size | Age | Female (%) | Sampling method | Sample frame | Uncorrected Seroprevalence (95% Confidence Interval) | Test Manufacturers \| Names \| Isotypes | Test Sens, Spec (Source) | Corrected Seroprevalence (95% Credible Interval) |
|---|---|---|---|---|---|---|---|---|---|---|---|---|---|
| Fairbanks School of Public Health)[430] | | | | | | 60-79 (n=169); >=80 (n=6) | | | community samples | | | / manufacturer evaluation) | |
| Bajema (Centers for Disease Control and Prevention)[428] | 2020-04-27 to 2020-05-01 | Georgia \| Atlanta | Local | Moderate | 1343 | Median 57 (IQR 36-73) | 55.2 | Non-probability | Residual sera | 4.2% | Centers for Disease Control and Prevention (CDC) \| CDC SARS-CoV-2 ELISA \| NR | 96.0%, 99.3% (Test developer / manufacturer evaluation) | 2.7% (0.1-4.8%) |
| Menachemi (Indiana University Fairbanks School of Public Health)[430] | 2020-04-25 to 2020-04-29 | Indiana | Regional | Moderate | 3518 | <40 (n=1017); 40 to 59 (n=1328); >=60 (n=1313) | 56.7 | Probability | Household and community samples | 1.1% (0.8-1.5%) | Author derived \| NR \| IgG | 100.0%, 99.6% (Test developer / manufacturer evaluation) | 0.4% (0.0-1.0%) |
| Chamie (University of California, San Francisco)[436] | 2020-04-25 to 2020-04-28 | California \| San Francisco | Local | Moderate | 3861 | 4-10 (n=109); 11-17 (n=135); 18-50 (n=2466); 51-70 (n=916); >70 (n=190) | 42.6 | Non-probability | Household and community samples | 3.4% (2.9-4.1%) | Abbott \| Abbott Architect SARS-CoV-2 IgG assay \| IgG | 0.4%, 1.0% (FINDDx / MUHC independent evaluation) | - |
| Rosenberg (New York State Department of Health)[437] | 2020-04-19 to 2020-04-28 | New York | Regional | Moderate | 15101 | 18-34 (n=3151); 35-44 (n=2628); 45-54 (n=3345); >=55 (n=5977) | 56.1 | Non-probability | Household and community samples | 14.0% (13.3-14.7%) | NYSDOH Wadsworth Center \| SARS-CoV IgG Microsphere Immunoassay \| IgG | 87.9%, 99.8% (Used author-adjusted estimate) | 14.0% (13.5-14.6%) |
| Zou (Georgia State University)[438] | 2020-04-17 to 2020-04-27 | Georgia \| Atlanta | Local | High | 127 | NR | NR | Non-probability | Household and community samples | 2.4% | Author-designed \| Krammer and colleagues ELISA S1 \| IgG | 75.6%, 97.6% (Used Bastos SR/MA data; no sens, spec, or author adjustment available) | 1.2% (0.0-4.7%) |
| Bryan (University of Washington School of Medicine)[439] | 2020-04-20 to 2020-04-26 | Idaho \| Boise | Local | High | 4856 | 0-19 (n=240); 20-29 (n=301); 30-39 (n=831); 40-49 (n=1102); 50-59 (n=1142); 60-69 (n=888); | 54.2 | Non-probability | Household and community samples | 1.8% | Abbott \| Abbott Architect SARS-CoV-2 IgG assay \| IgG | 0.4%, 1.0% (FINDDx / MUHC independent evaluation) | - |

| Article Author (Study Organization) | Sampling Dates (YMD) | Location | Geographic Scope | Overall Risk of Bias | Sample Size | Age | Female (%) | Sampling method | Sample frame | Uncorrected Seroprevalence (95% Confidence Interval) | Test Manufacturers \| Names \| Isotypes | Test Sens, Spec (Source) | Corrected Seroprevalence (95% Credible Interval) |
|---|---|---|---|---|---|---|---|---|---|---|---|---|---|
| | | | | | | 70-79 (n=327); >=80 (n=25) | | | | | | | |
| Appa (University of California San Francisco)[441] | 2020-04-20 to 2020-04-24 | California \| Bolinas | Local | Moderate | 1880 | NR | NR | Non-probability | Household and community samples | 0.2% | University of California \| In-House ELISA \| IgG | 96.2%, 99.9% (Test developer / manufacturer evaluation) | 0.1% (0.0-0.3%) |
| Sassoon (OMNI Healthcare)[443] | 2020-04-17 to 2020-04-24 | Florida | Local | High | 1000 | NR | NR | Non-probability | Household and community samples | 1.0% | NR \| NR \| IgG, IgM | 100.0%, 100.0% (Test developer / manufacturer evaluation) | 0.7% (0.0-1.4%) |
| Dingens (Fred Hutchinson Cancer Research Center)[444] | 2020-03-03 to 2020-04-24 | Washington \| Seattle | Sublocal | High | 1076 | 0-4 (n=192); 5-9 (n=214); 10-14 (n=301); >15 (n=369) | 49.7 | Non-probability | Residual sera | 0.9% | Abbott; N/A \| In-house ELISA assays, Abbott Architect SARS-CoV-2 IgG assay \| IgG | No data | - |
| Morrison (Crush The Curve Idaho)[446] | Until 2020-04-22 | Idaho | Regional | High | 1946 | NR | NR | Non-probability | Residual sera | 1.8% | University of Washington \| NR \| IgG | 100.0%, 99.6% (Test developer / manufacturer evaluation) | 0.8% (0.0-1.7%) |
| Paterson (Baton Rouge General Hospital)[449] | Until 2020-04-21 | Louisiana \| Baton Rouge | Local | Unclear | 432 | NR | NR | Non-probability | Household and community samples | 4.4% | Mayo Clinic \| NR \| NR | No data | - |
| Naranbhai (Massachusetts General Hospital)[453] | 2020-04-14 to 2020-04-15 | Massachusetts \| Chelsea | Local | Moderate | 200 | Median 46 (IQR 27-55) | 40.0 | Non-probability | Household and community samples | 32.8% (27.2-38.8%) | BioMedomics \| BioMedomics SARS-CoV-2 combined IgM/IgG LFA \| IgG, IgM | 90.0%, 99.0% (Used author-adjusted estimate) | 32.8% (26.3-39.2%) |
| Sood (Stanford University School of Medicine)[454] | 2020-04-10 to 2020-04-14 | California | Local | Moderate | 863 | 18-34 (n=191); 35-54 (n=475); >= 55 years (n=197) | 59.6 | Probability | Household and community samples | 4.7% (2.5-7.1%) | Premier Biotech \| NR \| IgG, IgM | 82.7%, 99.5% (Used author-adjusted estimate) | 4.7% (3.4-6.2%) |
| Ng (University of California, San Francisco)[455] | 2020-03-17 to 2020-04-13 | California \| San Francisco | Local | High | 387 | <=19 (n=15); 20-29 (n=28); 30-39 (n=41); 40-49 (n=54); 50-59 (n=68); | 48.1 | Non-probability | Residual sera | 0.3% (0.0-0.8%) | Abbott \| Abbott Architect SARS-CoV-2 IgG \| IgG | 0.4%, 1.0% (FINDDx / MUHC independent evaluation) | - |

| Article Author (Study Organization) | Sampling Dates (YMD) | Location | Geographic Scope | Overall Risk of Bias | Sample Size | Age | Female (%) | Sampling method | Sample frame | Uncorrected Seroprevalence (95% Confidence Interval) | Test Manufacturers \| Names \| Isotypes | Test Sens, Spec (Source) | Corrected Seroprevalence (95% Credible Interval) |
|---|---|---|---|---|---|---|---|---|---|---|---|---|---|
| | | | | | | 60-69 (n=71); 70-79 (n=56); 80-89 (n=20); 90-99 (n=5); unknown (n=29) | | | | | | | |
| Bendavid (Stanford University School of Medicine)[457] | 2020-04-03 to 2020-04-04 | California | Local | High | 3328 | 0-19 (n=637); 20-39 (n=907); 40-69 (n=1706); >= 70 (n=78) | 63.1 | Non-probability | Household and community samples | 2.8% (1.3-4.2%) | Premier Biotech \| NR \| IgG, IgM | 85.6%, 99.5% (Used author-adjusted estimate) | 2.8% (2.2-3.3%) |
| Ng (University of California, San Francisco)[455] | 2020-03-01 to 2020-03-31 | California \| San Francisco | Local | Moderate | 1000 | <=19 (n=18); 20-29 (n=152); 30-39 (n=205); 40-49 (n=172); 50-59 (n=185); 60-69 (n=197); 70-79 (n=63); 80-89 (n=7) | 49.1 | Non-probability | Blood donors | 0.1% (0.0-0.6%) | Ortho-Clinical Diagnostics; Abbott \| Abbott Architect SARS-CoV-2 IgG, VITROS anti-SARS-CoV-2 total antibody, SARS-CoV-2 neutralization assay. \| IgG | No data | - |
| Schuchat (University of Washington)[459] | 2020-03-01 to 2020-03-31 | Washington \| Seattle | Local | High | 221 | NR | NR | Non-probability | Residual sera | 0.4% | NR \| NR \| NR | No data | - |
| Basavaraju (American Red Cross)[462] | 2019-12-13 to 2020-01-17 | Wisconsin, Oregon, Massachusetts, Michigan, Connecticut, Washington, California, Iowa, Rhode Island | National | Moderate | 7389 | 16-29 (n = 895) ; 30-39 (n = 885) ; 40-49 (n = 1070) ; 50-59 (n = 1844) ; 60-69 (n = 1893) ; 70+ (n = 802) | 46.0 | Non-probability | Blood donors | 1.4% | CDC \| pan-IgG ELISA \| NR | 96.0%, 99.3% (Test developer / manufacturer evaluation) | 0.6% (0.0-1.1%) |
| **Latin America and Caribbean** | | | | | | | | | | | | | |
| **Brazil** | | | | | | | | | | | | | |

| Article Author (Study Organization) | Sampling Dates (YMD) | Location | Geographic Scope | Overall Risk of Bias | Sample Size | Age | Female (%) | Sampling method | Sample frame | Uncorrected Seroprevalence (95% Confidence Interval) | Test Manufacturers \| Names \| Isotypes | Test Sens, Spec (Source) | Corrected Seroprevalence (95% Credible Interval) |
|---|---|---|---|---|---|---|---|---|---|---|---|---|---|
| Buss (Universidade de Sao Paulo)[463] | 2020-10-12 to 2020-10-24 | Sao Paulo State \| Sao Paulo | Local | Moderate | 877 | NR | NR | Non-probability | Blood donors | 18.2% (15.0-21.7%) | Abbott Laboratories \| Abbott N IgG Assay \| IgG | 92.2%, 96.1% (Used author-adjusted estimate) | 18.2% (15.7-20.8%) |
| Buss (Universidade de Sao Paulo)[463] | 2020-10-10 to 2020-10-17 | Amazonas \| Manaus | Local | Moderate | 882 | NR | NR | Non-probability | Blood donors | 43.1% (38.3-47.9%) | Abbott Laboratories \| Abbott N IgG Assay \| IgG | 0.4%, 1.0% (FINDDx / MUHC independent evaluation) | 56.3% (52.5-59.6%) |
| Buss (Universidade de Sao Paulo)[463] | 2020-09-07 to 2020-09-29 | Sao Paulo State \| Sao Paulo | Local | Moderate | 933 | NR | NR | Non-probability | Blood donors | 15.1% (12.3-18.2%) | Abbott Laboratories \| Abbott N IgG Assay \| IgG | 92.2%, 96.1% (Used author-adjusted estimate) | 15.1% (12.8-17.4%) |
| Buss (Universidade de Sao Paulo)[463] | 2020-09-05 to 2020-09-14 | Amazonas \| Manaus | Local | Moderate | 868 | NR | NR | Non-probability | Blood donors | 45.4% (40.8-50.1%) | Abbott Laboratories \| Abbott N IgG Assay \| IgG | 0.4%, 1.0% (FINDDx / MUHC independent evaluation) | 54.0% (50.4-57.5%) |
| Buss (Universidade de Sao Paulo)[463] | 2020-08-10 to 2020-08-29 | Sao Paulo State \| Sao Paulo | Local | Moderate | 906 | NR | NR | Non-probability | Blood donors | 15.5% (12.3-18.2%) | Abbott Laboratories \| Abbott N IgG Assay \| IgG | 92.2%, 96.1% (Used author-adjusted estimate) | 15.5% (13.2-17.9%) |
| Buss (Universidade de Sao Paulo)[463] | 2020-08-08 to 2020-08-19 | Amazonas \| Manaus | Local | Moderate | 881 | NR | NR | Non-probability | Blood donors | 44.0% (40.0-48.0%) | Abbott Laboratories \| Abbott N IgG Assay \| IgG | 0.4%, 1.0% (FINDDx / MUHC independent evaluation) | 55.4% (51.8-58.8%) |
| daSilva (Federal University of Maranhao)[465] | 2020-07-27 to 2020-08-08 | Maranhao | Regional | Moderate | 3156 | 1-9 (n=124); 10-19 (n=10-19); 20-29 (n=427); 30-39 (n=475); 40-49 (n=502); 50-59 (n=501); 60-69 (n=409); >70 (n=386). | 62.0 | Probability | Household and community samples | 40.4% (35.6-45.3%) | Roche Diagnostics \| Elecsys Anti-SARS-CoV-2 assay \| IgG, IgM | 0.8%, 1.0% (FINDDx / MUHC independent evaluation) | 59.6% (57.8-61.5%) |
| Oliveira (Universidade de São Paulo)[466] | 2020-06-30 to 2020-08-04 | State of Sao Paulo \| Sao Paulo | Sublocal | High | 439 | 18-39 years old (n=121); 40-59 years (n=185); ≥ 60 years old (n=133) | 64.5 | Non-probability | Residual sera | 13.9% | Roche Diagnostics; DiaSorin S.p.A. \| LIAISON® SARS-CoV-2 S1/S2 IgG, Elecsys Anti- | 97.8%, 97.8% (Used Bastos SR/MA data; no sens, spec, or author adjustment available) | 12.4% (9.0-15.9%) |

| Article Author (Study Organization) | Sampling Dates (YMD) | Location | Geographic Scope | Overall Risk of Bias | Sample Size | Age | Female (%) | Sampling method | Sample frame | Uncorrected Seroprevalence (95% Confidence Interval) | Test Manufacturers \| Names \| Isotypes | Test Sens, Spec (Source) | Corrected Seroprevalence (95% Credible Interval) |
|---|---|---|---|---|---|---|---|---|---|---|---|---|---|
| | | | | | | | | | | | SARSCoV-2 Cobas® assay \| IgG | | |
| Buss (Universidade de Sao Paulo)[463] | 2020-07-01 to 2020-07-31 | Sao Paulo State \| Sao Paulo | Local | Unclear | 1000 | NR | NR | Non-probability | Blood donors | 13.3% | Abbott Laboratories; Roche Diagnostics \| Abbott N IgG Assay; Roche Elecsys SARS-CoV-2 \| IgG | 97.8%, 97.8% (Used Bastos SR/MA data; no sens, spec, or author adjustment available) | 11.7% (9.6-14.1%) |
| Paula (Universidade Federal de Mato Grosso (UFMT))[468] | 2020-04-01 to 2020-07-31 | Mato Grosso \| Cuiabá | Sublocal | High | 6392 | 0-9 (n=168); 10-19 (n= 220); 20-29 (n= 782); 30-39 (n= 1562); 40-49 (1472); 50-59 (n=1076); 60-69 (n=748); 70-79 (n=272); 80-89 (n= 74); 90-100 (n=14); 101-120 (n=4). | 59.7 | Probability | Residual sera | 22.1% | Carlos Chagas Laboratory \| SOROVID-19? \| IgG, IgM | No data | - |
| Buss (Universidade de Sao Paulo)[463] | 2020-07-13 to 2020-07-25 | Sao Paulo State \| Sao Paulo | Local | Moderate | 879 | NR | NR | Non-probability | Blood donors | 10.6% (8.1-13.5%) | Abbott Laboratories \| Abbott N IgG Assay \| IgG | 92.2%, 96.1% (Used author-adjusted estimate) | 10.6% (8.7-12.7%) |
| Buss (Universidade de Sao Paulo)[463] | 2020-07-06 to 2020-07-15 | Amazonas \| Manaus | Local | Moderate | 1147 | NR | NR | Non-probability | Blood donors | 46.9% (42.7-51.1%) | Abbott Laboratories \| Abbott N IgG Assay \| IgG | 0.4%, 1.0% (FINDDx / MUHC independent evaluation) | 52.5% (49.3-55.7%) |
| Horta (Universidade Federal de Pelotas)[470] | 2020-05-14 to 2020-06-24 | | National | Moderate | 89362 | ≤ 9 (n= 4 263) 10 – 19 (n = 8 024) 20 – 39 (n = 27 485) 40 – 59 (n = 28 402) 60+ (n = 21 188) | 58.2 | Probability | Household and community samples | 2.3% | China; Guangzhou; Wondfo Biotech; Co. \| WONDFO SARS-CoV-2 Antibody Test \| IgG, IgM | 0.7%, 1.0% (FINDDx / MUHC independent evaluation) | - |
| Buss (Universidade de Sao Paulo)[463] | 2020-06-08 to 2020-06-20 | Sao Paulo State \| Sao Paulo | Local | Moderate | 880 | NR | NR | Non-probability | Blood donors | 13.1% (10.3-16.3%) | Abbott Laboratories \| Abbott N IgG Assay \| IgG | 92.2%, 96.1% (Used author-adjusted estimate) | 13.1% (11.0-15.4%) |

| Article Author (Study Organization) | Sampling Dates (YMD) | Location | Geographic Scope | Overall Risk of Bias | Sample Size | Age | Female (%) | Sampling method | Sample frame | Uncorrected Seroprevalence (95% Confidence Interval) | Test Manufacturers \| Names \| Isotypes | Test Sens, Spec (Source) | Corrected Seroprevalence (95% Credible Interval) |
|---|---|---|---|---|---|---|---|---|---|---|---|---|---|
| Picon (Universidade do Vale do Taquari)[471] | 2020-06-13 to 2020-06-17 | Rio Grande do Sul \| Lajeado | Local | Moderate | 1127 | Median 49.9 (positive); 49.9 (negative) (IQR 31.1 (positive); 26.9 (negative)) | 84.5 | Probability | Household and community samples | 2.6% (1.8-3.7%) | Guangzhou Wondfo Biotech \| Wondfo Lateral Flow Immunoassay \| IgG, IgM | 76.2%, 99.6% (Used author-adjusted estimate) | 2.6% (1.8-3.6%) |
| Buss (Universidade de Sao Paulo)[463] | 2020-06-05 to 2020-06-15 | Amazonas \| Manaus | Local | Moderate | 911 | NR | NR | Non-probability | Blood donors | 52.7% (48.5-56.9%) | Abbott Laboratories \| Abbott N IgG Assay \| IgG | 0.4%, 1.0% (FINDDx / MUHC independent evaluation) | 46.5% (42.7-50.0%) |
| Picon (Universidade do Vale do Taquari)[471] | 2020-05-30 to 2020-06-04 | Rio Grande do Sul \| Lajeado | Local | Moderate | 1450 | Median 45.4 (positive); 49.6 (negative) (IQR 24.8 (positive); 27.8 (n egative)) | 49.9 | Probability | Household and community samples | 4.0% (3.1-5.2%) | Guangzhou Wondfo Biotech \| Wondfo Lateral Flow Immunoassay \| IgG, IgM | 76.2%, 99.6% (Used author-adjusted estimate) | 4.0% (3.1-5.1%) |
| Vieira ( Centro de Operações de Emergências em Saúde Pública)[472] | 2020-05-31 to 2020-05-31 | Piaui \| Teresina | Local | Moderate | 900 | NR | NR | Probability | Household and community samples | 8.3% (6.6-10.3%) | Wondfo Biotech Co. \| WONDFO SARS-CoV-2 Antibody Test \| IgG, IgM | 0.7%, 1.0% (FINDDx / MUHC independent evaluation) | - |
| Sales (Universidade Federal do Ceará)[473] | 2020-05-22 to 2020-05-31 | State of Pernambu co \| Fernando de Noronha | Local | Moderate | 904 | <10 (n =31), 10-18 (n=46), 19-39 (n=366), 40-58 (n=375), 59-68 (n=69), =>69 (n=17) | 52.1 | Probability | Household and community samples | 4.3% (3.2-5.9%) | Ltd.; Guangzhou Wondfo Biotech Co. \| SARS-CoV-2 Antibody Test Kit (IgG / IgM) \| IgG, IgM | 0.7%, 1.0% (FINDDx / MUHC independent evaluation) | - |
| Jr (Secretaria Municipal de Saude)[474] | 2020-05-30 to 2020-05-30 | Biguacu | Local | Unclear | 217 | <20 (n=39); 20-30 (n=29); 30-40 (n=46); 40-50 (n=36); 50-60 (n=29); 70-80 (n=19); >80 (n=18) | 58.1 | Probability | Household and community samples | 0.4% | NR \| NR \| NR | No data | - |
| Vieira ( Centro de Operações de Emergências | 2020-05-24 to 2020-05-24 | Piaui \| Teresina | Local | Moderate | 900 | NR | NR | Probability | Household and community samples | 5.8% (4.3-7.3%) | Wondfo Biotech Co. \| WONDFO SARS-CoV-2 | 0.7%, 1.0% (FINDDx / MUHC independent evaluation) | - |

| Article Author (Study Organization) | Sampling Dates (YMD) | Location | Geographic Scope | Overall Risk of Bias | Sample Size | Age | Female (%) | Sampling method | Sample frame | Uncorrected Seroprevalence (95% Confidence Interval) | Test Manufacturers \| Names \| Isotypes | Test Sens, Spec (Source) | Corrected Seroprevalence (95% Credible Interval) |
|---|---|---|---|---|---|---|---|---|---|---|---|---|---|
| em Saúde Pública)[472] | | | | | | | | | | | Antibody Test \| IgG, IgM | | |
| Buss (Universidade de Sao Paulo)[463] | 2020-05-08 to 2020-05-21 | Sao Paulo State \| Sao Paulo | Local | Moderate | 826 | NR | NR | Non-probability | Blood donors | 4.7% (2.6-7.3%) | Abbott Laboratories \| Abbott N IgG Assay \| IgG | 92.2%, 96.1% (Used author-adjusted estimate) | 4.7% (3.3-6.2%) |
| Vieira ( Centro de Operações de Emergências em Saúde Pública)[472] | 2020-05-17 to 2020-05-17 | Piaui \| Teresina | Local | Moderate | 900 | NR | NR | Probability | Household and community samples | 3.8% (2.6-5.2%) | Wondfo Biotech Co. \| WONDFO SARS-CoV-2 Antibody Test \| IgG, IgM | 0.7%, 1.0% (FINDDx / MUHC independent evaluation) | - |
| Chioro (Baixada Santista Metropolitan Region )[476] | 2020-05-13 to 2020-05-15 | São Paulo state \| Baixada Santista | Local | High | 2415 | Mean 37.55 (NR) | NR | Probability | Household and community samples | 2.2% | Wondfo \| Wondfo SARS-CoV-2 test \| NR | 0.8%, 1.0% (FINDDx / MUHC independent evaluation) | - |
| Gomes (Secretaria de Estado da Saúde do Espírito Santo)[477] | 2020-05-13 to 2020-05-15 | Espírito Santo | Regional | Moderate | 4612 | <=20 years (n=434); 21-40 (n=1367); 41-60 (n=1583); 61-80 (n=1081); >= 81 (n=143) | 60.9 | Probability | Household and community samples | 2.1% (1.7-2.5%) | Celer Technologies Inc. \| NR \| IgG, IgM | 86.4%, 97.6% (Test developer / manufacturer evaluation) | 0.8% (0.0-2.0%) |
| Gomes (Secretaria de Estado da Saúde do Espírito Santo)[477] | 2020-05-13 to 2020-05-15 | Espírito Santo | Regional | Moderate | 1163 | NR | NR | Probability | Household and community samples | 0.3% (0.1-0.8%) | Celer Technologies Inc. \| NR \| IgG, IgM | 86.4%, 97.6% (Test developer / manufacturer evaluation) | 0.1% (0.0-0.5%) |
| Buss (Universidade de Sao Paulo)[463] | 2020-05-05 to 2020-05-14 | Amazonas \| Manaus | Local | Moderate | 901 | NR | NR | Non-probability | Blood donors | 42.1% (37.5-46.7%) | Abbott Laboratories \| Abbott N IgG Assay \| IgG | 0.4%, 1.0% (FINDDx / MUHC independent evaluation) | 57.3% (53.6-60.6%) |
| Tess (Universidade de São Paulo)[478] | 2020-05-04 to 2020-05-12 | São Paulo \| São Paulo | Local | Moderate | 517 | 18-44 (n=269); >= 45 (n=248) | 54.9 | Probability | Household and community samples | 4.7% (3.0-6.6%) | SNIBE – Shenzhen New Industries Biomedical Engineering Co. \| MAGLUMI 2019-nCoV | 100.0%, 99.5% (Test developer / manufacturer evaluation) | 3.0% (0.0-5.3%) |

| Article Author (Study Organization) | Sampling Dates (YMD) | Location | Geographic Scope | Overall Risk of Bias | Sample Size | Age | Female (%) | Sampling method | Sample frame | Uncorrected Seroprevalence (95% Confidence Interval) | Test Manufacturers \| Names \| Isotypes | Test Sens, Spec (Source) | Corrected Seroprevalence (95% Credible Interval) |
|---|---|---|---|---|---|---|---|---|---|---|---|---|---|
| | | | | | | | | | | | IgG (CLIA) \| IgG, IgM | | |
| Watanabe (Grupo Fleury)[479] | 2020-05-04 to 2020-05-12 | São Paulo \| São Paulo | Sublocal | Unclear | 520 | NR | NR | Probability | Household and community samples | 5.2% | NR \| NR \| NR | No data | - |
| Silveira (Universidade Federal de Pelotas)[480] | 2020-05-09 to 2020-05-11 | Rio Grande do Sul \| Pelotas, Santa Maria, Canoas, Ijui, Uruguaiana, Porto Alegre, Passo Fundo, Santa Cruz do Sul, Caxias | Regional | Low | 4500 | 0-9 (n=113); 10-19 (n=266); 20-29 (n=581); 30-39 (n=684); 40-49 (n=689); 50-59 (n=769); 60-69 (n=734); 70-79 (n=464); >=80 (n=202); | 58.9 | Probability | Household and community samples | 0.2% (0.1-0.4%) | Wondfo Biotech Co. \| WONDFO SARS-CoV-2 Antibody Test \| IgG, IgM | 0.8%, 1.0% (FINDDx / MUHC independent evaluation) | - |
| Vieira ( Centro de Operações de Emergências em Saúde Pública)[472] | 2020-05-10 to 2020-05-10 | Piaui \| Teresina | Local | Moderate | 900 | NR | NR | Probability | Household and community samples | 2.0% (1.2-3.1%) | Wondfo Biotech Co. \| WONDFO SARS-CoV-2 Antibody Test \| IgG, IgM | 0.7%, 1.0% (FINDDx / MUHC independent evaluation) | - |
| Borges (Federal University of Fergipe )[481] | 2020-05-02 to 2020-05-09 | Sergipe \| Nossa Senhora do, São Cristóvão, Lagarto, Estância, Tobias Barreto, Itabaiana, Nossa Senhora da Glória, Itabaianinha, Simão Dias, Aracaju | Regional | High | 2635 | Mean 39.76 (SD 16.83) | 59.8 | Non-probability | Household and community samples | 8.3% (7.2-9.4%) | Boditech Med Inc. \| Ichroma2™ COVID-19 Ab \| IgG | 66.0%, 96.6% (Used Bastos SR/MA data; no sens, spec, or author adjustment available) | 7.8% (5.8-9.7%) |

| Article Author (Study Organization) | Sampling Dates (YMD) | Location | Geographic Scope | Overall Risk of Bias | Sample Size | Age | Female (%) | Sampling method | Sample frame | Uncorrected Seroprevalence (95% Confidence Interval) | Test Manufacturers \| Names \| Isotypes | Test Sens, Spec (Source) | Corrected Seroprevalence (95% Credible Interval) |
|---|---|---|---|---|---|---|---|---|---|---|---|---|---|
| Vieira ( Centro de Operações de Emergências em Saúde Pública)[472] | 2020-05-03 to 2020-05-03 | Piaui \| Teresina | Local | Moderate | 900 | NR | NR | Probability | Household and community samples | 1.4% (0.8-2.5%) | Wondfo Biotech Co. \| WONDFO SARS-CoV-2 Antibody Test \| IgG, IgM | 0.7%, 1.0% (FINDDx / MUHC independent evaluation) | - |
| Buss (Universidade de Sao Paulo)[463] | 2020-04-08 to 2020-04-30 | Sao Paulo State \| Sao Paulo | Local | Moderate | 900 | NR | NR | Non-probability | Blood donors | 4.0% (1.8-6.6%) | Abbott Laboratories \| Abbott N IgG Assay \| IgG | 92.2%, 96.1% (Used author-adjusted estimate) | 4.0% (2.9-5.4%) |
| Silveira (Universidade Federal de Pelotas)[480] | 2020-04-25 to 2020-04-27 | Rio Grande do Sul \| Pelotas, Canoas, Santa Maria, Ijui, Uruguaiana, Porto Alegre, Passo Fundo, Santa Cruz do Sul, Caxias | Regional | Low | 4501 | 0-9 (n=116); 10-19 (n=227); 20-29 (n=508); 30-39 (n=753); 40-49 (n=647); 50-59 (n=798); 60-69 (n=798); 70-79 (n=459); >=80 (n=156) | 58.9 | Probability | Household and community samples | 0.1% (0.0-0.3%) | Wondfo Biotech Co. \| WONDFO SARS-CoV-2 Antibody Test \| IgG, IgM | 0.8%, 1.0% (FINDDx / MUHC independent evaluation) | - |
| Filho (Universidade do Estado do Rio de Janeiro)[483] | 2020-04-14 to 2020-04-27 | Rio de Janeiro \| Rio de Janeiro | Local | Moderate | 2857 | 18-29 (n=870); 30-49 (n=1443); 50-69 (n=544) | 49.2 | Non-probability | Blood donors | 4.0% (3.3-4.7%) | MedLevensohn \| MedTest Coronavirus 2019-nCoV IgG/IgM \| IgG, IgM | 85.0%, 99.0% (Test developer / manufacturer evaluation) | 2.8% (0.0-4.9%) |
| Vieira ( Centro de Operações de Emergências em Saúde Pública)[472] | 2020-04-26 to 2020-04-26 | Piaui \| Teresina | Local | Moderate | 900 | NR | NR | Probability | Household and community samples | 0.9% (0.4-1.8%) | Wondfo Biotech Co. \| WONDFO SARS-CoV-2 Antibody Test \| IgG, IgM | 0.7%, 1.0% (FINDDx / MUHC independent evaluation) | - |
| Vieira ( Centro de Operações de Emergências em Saúde Pública)[472] | 2020-04-19 to 2020-04-19 | Piaui \| Teresina | Local | Moderate | 900 | NR | NR | Probability | Household and community samples | 0.6% (0.2-1.3%) | Wondfo Biotech Co. \| WONDFO SARS-CoV-2 Antibody Test \| IgG, IgM | 0.7%, 1.0% (FINDDx / MUHC independent evaluation) | - |

| Article Author (Study Organization) | Sampling Dates (YMD) | Location | Geographic Scope | Overall Risk of Bias | Sample Size | Age | Female (%) | Sampling method | Sample frame | Uncorrected Seroprevalence (95% Confidence Interval) | Test Manufacturers \| Names \| Isotypes | Test Sens, Spec (Source) | Corrected Seroprevalence (95% Credible Interval) |
|---|---|---|---|---|---|---|---|---|---|---|---|---|---|
| Buss (Universidade de Sao Paulo)[463] | 2020-04-06 to 2020-04-17 | Amazonas \| Manaus | Local | Moderate | 829 | NR | NR | Non-probability | Blood donors | 5.0% (2.9-7.4%) | Abbott Laboratories \| Abbott N IgG Assay \| IgG | 92.2%, 96.1% (Used author-adjusted estimate) | 5.0% (3.6-6.6%) |
| Gurgel (Federal University of Sergipe)[484] | 2020-01-15 to 2020-04-15 | Sergipe \| Aracaju | Local | Moderate | 987 | Mean 38.9 (SD 22.2) | 69.2 | Non-probability | Residual sera | 1.6% | Nantong Egens Biotechnology CO; Ltd.; China and Bodytech Med Inc. \| Nantong Egens Biotechnology CO, Ltd; iChroma2™ \| IgG, IgM | 66.0%, 96.6% (Used Bastos SR/MA data; no sens, spec, or author adjustment available) | 0.2% (0.0-0.9%) |
| Silveira (Universidade Federal de Pelotas)[480] | 2020-04-11 to 2020-04-13 | Rio Grande do Sul \| Pelotas, Santa Maria, Canoas, Ijui, Uruguaiana, Porto Alegre, Passo Fundo, Santa Cruz do Sul, Caxias | Regional | Low | 4151 | 0-9 (n=149); 10-19 (n=224); 20-29 (n=506); 30-39 (n=635); 40-49 (n=643); 50-59 (n=734); 60-69 (n=685); 70-79 (n=3985); >=80 (n=183) | 58.3 | Probability | Household and community samples | 0.0% (0.0-0.2%) | Wondfo Biotech Co. \| WONDFO SARS-CoV-2 Antibody Test \| IgG, IgM | 0.8%, 1.0% (FINDDx / MUHC independent evaluation) | - |
| Buss (Universidade de Sao Paulo)[463] | 2020-03-09 to 2020-03-21 | Sao Paulo State \| Sao Paulo | Local | Moderate | 2454 | NR | NR | Non-probability | Blood donors | 2.8% (1.8-3.9%) | Abbott Laboratories \| Abbott N IgG Assay \| IgG | 92.2%, 96.1% (Used author-adjusted estimate) | 2.8% (2.2-3.5%) |
| Buss (Universidade de Sao Paulo)[463] | 2020-03-06 to 2020-03-12 | Amazonas \| Manaus | Local | Moderate | 832 | NR | NR | Non-probability | Blood donors | 0.0% (0.0-0.8%) | Abbott Laboratories \| Abbott N IgG Assay \| IgG | 92.2%, 96.1% (Used author-adjusted estimate) | 0.0% (0.0-0.3%) |
| Buss (Universidade de Sao Paulo)[463] | 2020-02-08 to 2020-02-29 | Sao Paulo State \| Sao Paulo | Local | Moderate | 799 | NR | NR | Non-probability | Blood donors | 1.0% (0.0-2.9%) | Abbott Laboratories \| Abbott N IgG Assay \| IgG | 92.2%, 96.1% (Used author-adjusted estimate) | 1.0% (0.4-1.7%) |

| Article Author (Study Organization) | Sampling Dates (YMD) | Location | Geographic Scope | Overall Risk of Bias | Sample Size | Age | Female (%) | Sampling method | Sample frame | Uncorrected Seroprevalence (95% Confidence Interval) | Test Manufacturers \| Names \| Isotypes | Test Sens, Spec (Source) | Corrected Seroprevalence (95% Credible Interval) |
|---|---|---|---|---|---|---|---|---|---|---|---|---|---|
| Buss (Universidade de Sao Paulo)[463] | 2020-02-07 to 2020-02-13 | Amazonas \| Manaus | Local | Moderate | 821 | NR | NR | Non-probability | Blood donors | 0.4% (0.0-3.3%) | Abbott Laboratories \| Abbott N IgG Assay \| IgG | 92.2%, 96.1% (Used author-adjusted estimate) | 0.4% (0.1-1.0%) |
| **Columbia** | | | | | | | | | | | | | |
| Mattar (Universidad de Córdoba)[485] | 2020-08-04 to 2020-09-01 | Caribbean Region \| Monteria | Local | Moderate | 1368 | NR | 57.8 | Probability | Household and community samples | 55.3% (52.5-57.8%) | Ingenasa \| INgezim COVID 19 DR test \| IgG, IgA, IgM | 92.0%, 100.0% (Author-reported independent evaluation) | 59.4% (50.4-68.7%) |
| **Dominican Republic** | | | | | | | | | | | | | |
| Paulino-Ramirez (Universidad Iberoamericana)[489] | 2020-04-15 to 2020-06-15 | | National | High | 12897 | Mean 42 (NR) | 48.8 | Probability | Household and community samples | 5.5% | GenBody Inc. \| Confiscope G20 \| IgG | 56.7%, 100.0% (Test developer / manufacturer evaluation) | 8.3% (3.2-13.0%) |
| **Ecuador** | | | | | | | | | | | | | |
| Brutto (Universidad Peruana Cayetano Heredia)[490] | 2020-06-25 to 2020-06-30 | Atahualpa | Local | High | 362 | Mean 59.9 (SD 13) | 56.6 | Probability | Household and community samples | 7.7% | BIOHIT Health Care \| BIOHIT SARS-CoV-2 antibody test \| IgG, IgM | 66.0%, 96.6% (Used Bastos SR/MA data; no sens, spec, or author adjustment available) | 6.7% (2.3-11.2%) |
| Brutto (Universidad Peruana Cayetano Heredia)[491] | 2020-05-01 to 2020-05-25 | El Oro | Local | Moderate | 673 | Mean 59.2 (SD 12.8) | 56.6 | Probability | Household and community samples | 45.0% | Biohit Health Care Ltd \| BIOHIT SARS-CoV-2 antibody test kit (Colloidal gold method) \| IgG, IgM | 66.0%, 96.6% (Used Bastos SR/MA data; no sens, spec, or author adjustment available) | 66.3% (60.1-72.3%) |
| **French Guiana** | | | | | | | | | | | | | |
| Flamand (Institut Pasteur )[492] | 2020-07-15 to 2020-07-23 | | National | Moderate | 480 | Mean 38.3 (Range 0.2 - 87) | 68.1 | Non-probability | Residual sera | 15.4% (9.3-24.4%) | Euroimmun \| anti-SARS-CoV-2 IgG ELISA \| IgG | 0.6%, 0.9% (FINDDx / MUHC independent evaluation) | - |
| **Honduras** | | | | | | | | | | | | | |

| Article Author (Study Organization) | Sampling Dates (YMD) | Location | Geographic Scope | Overall Risk of Bias | Sample Size | Age | Female (%) | Sampling method | Sample frame | Uncorrected Seroprevalence (95% Confidence Interval) | Test Manufacturers \| Names \| Isotypes | Test Sens, Spec (Source) | Corrected Seroprevalence (95% Credible Interval) |
|---|---|---|---|---|---|---|---|---|---|---|---|---|---|
| Núñez (Colegio Médico)[493] | 2020-06-16 to 2020-06-23 | | National | Moderate | 792 | Median 37 (IQR 27-50) | 54.7 | Probability | Household and community samples | 6.2% | NR \| NR \| IgG, IgM | 93.4%, 97.7% (Test developer / manufacturer evaluation) | 3.7% (0.0-6.6%) |
| **Mexico** | | | | | | | | | | | | | |
| Martinez-Acuña (Autonomous University of Nuevo Leon)[495] | 2020-01-01 to 2020-08-30 | Nuevo Leon | Regional | High | 1968 | Mean 34.8 (Range 18 to 65) | 27.0 | Non-probability | Blood donors | 4.0% | Abbott Laboratories \| SARS-CoV-2 IgG ARCHITECT \| IgG | 0.4%, 1.0% (FINDDx / MUHC independent evaluation) | - |
| Remes-Troche (Universidad Veracruzana)[496] | 2020-06-01 to 2020-07-31 | Veracruz City | Sublocal | High | 2174 | Mean 41.8 (Range 18-98) | 53.6 | Non-probability | Residual sera | 29.5% (27.6-31.5%) | Ortho-Clinical Diagnostics; Abbott \| Abbott SARS- CoV-2 IgG assay \| IgG | 0.4%, 1.0% (FINDDx / MUHC independent evaluation) | - |
| **Panama** | | | | | | | | | | | | | |
| Villarreal (Instituto de Investigaciones Científicas y Servicios de Alta Tecnología (INDICASAT-AIP))[498] | 2020-04-30 to 2020-07-07 | Colon, Panama | National | High | 255 | Median 73.2 (SD 11.1) | 29.0 | Non-probability | Blood donors | 11.7% (8.3-16.3%) | N/A - Author Designed \| Chinese Academy of Science Test (CAST) \| IgG, IgM | 87.0%, 98.9% (Test developer / manufacturer evaluation) | 11.4% (4.5-17.6%) |
| **Paraguay** | | | | | | | | | | | | | |
| Ultimahora (Ministerio de Salud Pública)[499] | 2020-11-29 to 2020-12-15 | Ciudad del Este | Local | Unclear | 1675 | NR | NR | Non-probability | Household and community samples | 30.0% | NR \| NR \| NR | No data | - |
| **Peru** | | | | | | | | | | | | | |
| Peruvian Government (National Center for Epidemiology, Prevention and Control of Diseases)[501] | 2020-06-28 to 2020-07-09 | Callao, Lima \| Callao, Lima | Local | High | 3118 | NR | NR | Probability | Household and community samples | 21.6% (18.7-24.6%) | NR \| NR \| IgG, IgM | 66.0%, 96.6% (Used Bastos SR/MA data; no sens, spec, or author adjustment available) | 29.1% (26.4-31.4%) |

| Article Author (Study Organization) | Sampling Dates (YMD) | Location | Geographic Scope | Overall Risk of Bias | Sample Size | Age | Female (%) | Sampling method | Sample frame | Uncorrected Seroprevalence (95% Confidence Interval) | Test Manufacturers \| Names \| Isotypes | Test Sens, Spec (Source) | Corrected Seroprevalence (95% Credible Interval) |
|---|---|---|---|---|---|---|---|---|---|---|---|---|---|
| Angulo-Bazán (National Institute of Health (INS))[502] | 2020-04-23 to 2020-05-02 | Lima \| Lima | Local | Moderate | 236 | Mean 36.2 (SD 20.1) | 54.7 | Non-probability | Household and community samples | 53.0% | Core Technology Co. Ltd \| Coretests COVID-19 IgM/IgG Ab Test \| IgG, IgM | 97.6%, 100.0% (Test developer / manufacturer evaluation) | 55.8% (47.9-65.8%) |
| **North Africa and Middle East** | | | | | | | | | | | | | |
| **Iran** | | | | | | | | | | | | | |
| Poustchi (Tehran University of Medical Sciences)[505] | 2020-04-17 to 2020-06-02 | Kerman, Shiraz, Esfahan , Urmia, Tehran, Ardabil, Ahvaz, Sari, Qom, Zahedan, Hamedan, Sanandaj, Kermansh ah, Mashhad, Gorgan, Tabriz, Babol , Rasht | National | Low | 3530 | ≤19 (n = 140); 20–29 (n = 470); 30–39 (n = 992); 40–49 (n = 937); 50–59 (n =590); ≥60 (n = 401) | 49.2 | Probability | Household and community samples | 12.9% (11.3-14.5%) | Pishtaz Teb \| PT-SARS-COV-2.IgM-96 and PT-SARS-COV-2.IgG-96 \| IgG, IgM | 66.9%, 98.2% (Author-reported independent evaluation) | 16.2% (10.7-20.6%) |
| Shakiba (Tehran University of Medical Sciences)[509] | 2020-04-11 to 2020-04-19 | Guilan | Regional | Moderate | 528 | <5 (n = 26) ; 5-17 (n = 101) ; 18-59 (n = 329) ; 60+ (n=72) | 51.3 | Probability | Household and community samples | 22.2% (16.4-28.5%) | VivaChek \| VivaDiag Rapid test kit \| IgG, IgM | 83.3%, 99.0% (Used author-adjusted estimate) | 22.2% (18.8-25.8%) |
| **Libya** | | | | | | | | | | | | | |
| Elfakhri (The Center of Infectious Diseases-Benghazi)[513] | 2020-05-02 to 2020-05-07 | Benghazi | Local | Moderate | 600 | Mean 38.2 (SD 17.5) | 50.5 | Probability | Household and community samples | 0.0% | Biotest \| Right Sign (Lot No. COV20030003 ) \| IgG, IgM | 81.7%, 96.7% (Test developer / manufacturer evaluation) | 0.2% (0.0-0.7%) |
| **Saudi Arabia** | | | | | | | | | | | | | |

| Article Author (Study Organization) | Sampling Dates (YMD) | Location | Geographic Scope | Overall Risk of Bias | Sample Size | Age | Female (%) | Sampling method | Sample frame | Uncorrected Seroprevalence (95% Confidence Interval) | Test Manufacturers \| Names \| Isotypes | Test Sens, Spec (Source) | Corrected Seroprevalence (95% Credible Interval) |
|---|---|---|---|---|---|---|---|---|---|---|---|---|---|
| Mahallawi (Taibah University)[517] | 2020-05-15 to 2020-07-15 | Al-Madinah | Local | High | 1212 | Mean 31.73 (SD 9.05) | 0.0 | Non-probability | Blood donors | 19.3% (17.1-21.6%) | In-house; NR \| In-house ELISA, NR \| IgG | No data | - |
| **Turkey** | | | | | | | | | | | | | |
| Sozcu (Unclear (Turkey Ministry of Health))[520] | 2020-06-15 to 2020-06-25 | | National | Unclear | 132000 | NR | NR | Non-probability | Household and community samples | 0.8% | NR \| NR \| NR | 75.6%, 97.6% (Used Bastos SR/MA data; no sens, spec, or author adjustment available) | 0.0% (0.0-0.1%) |
| **South Asia** | | | | | | | | | | | | | |
| **India** | | | | | | | | | | | | | |
| Khan (Government Medical College Sringar)[522] | 2020-10-17 to 2020-11-04 | | Regional | Low | 6230 | 18-29 (n=1513); 30-49 (n=2672); 50-69 (n=1643); >70 (n=402) | 49.8 | Probability | Household and community samples | 36.7% (34.3-39.2%) | Abbott \| Abbott SARS-CoV-2 IgG assay \| IgG | 100.0%, 99.6% (Used author-adjusted estimate) | 36.7% (35.5-37.9%) |
| Kumar (Unclear)[523] | 2020-10-18 to 2020-10-24 | Haryan | Regional | Unclear | 16477 | NR | NR | Probability | Household and community samples | 14.8% | NR \| NR \| NR | No data | - |
| Sharma (Government of National Capital Territory of Delhi)[524] | 2020-10-15 to 2020-10-21 | NCT Delhi | Regional | Low | 15015 | <18 (n=2430); 18-49 (n=9420); =>50 (n=3043) | 52.9 | Probability | Household and community samples | 25.2% (24.5-25.9%) | NR \| ERBALISA COVID-19 IgG \| IgG | 99.1%, 99.3% (Test developer / manufacturer evaluation) | 25.0% (20.8-29.2%) |
| Ramanath (The Regional Medical Research Centre in Bhubaneswar)[525] | 2020-10-16 to 2020-10-18 | Bhubaneswar | Local | Unclear | 1403 | NR | NR | Probability | Household and community samples | 50.0% | NR \| NR \| NR | No data | - |
| Banerjee (Dr DY Patil Medical College )[526] | 2020-10-07 to 2020-10-17 | Maharashtra \| Pune | Local | Moderate | 5000 | NR | NR | Probability | Household and community samples | 34.0% (31.3-36.8%) | IL; Abbott Chicago; USA. \| Abbott IgG \| IgG | 92.7%, 99.9% (Used author-adjusted estimate) | 34.0% (32.7-35.4%) |

| Article Author (Study Organization) | Sampling Dates (YMD) | Location | Geographic Scope | Overall Risk of Bias | Sample Size | Age | Female (%) | Sampling method | Sample frame | Uncorrected Seroprevalence (95% Confidence Interval) | Test Manufacturers \| Names \| Isotypes | Test Sens, Spec (Source) | Corrected Seroprevalence (95% Credible Interval) |
|---|---|---|---|---|---|---|---|---|---|---|---|---|---|
| Babu (Indian Institute of Public Health)[527] | 2020-09-03 to 2020-09-16 | Karnatka | Regional | Moderate | 15939 | 18-29 (n=5184); 30-39 (n=3353); 40-49 (n=2447); 50-59 (n=1792); >=60 (n=2848) | 51.2 | Non-probability | Household and community samples | 16.1% | India; Zydus Cadila \| Covid Kavach Anti SARS-CoV-2 IgG antibody detection ELISA \| IgG | 92.1%, 99.8% (Test developer / manufacturer evaluation) | 16.3% (11.4-20.0%) |
| Sharma (Government of National Capital Territory of Delhi)[524] | 2020-09-01 to 2020-09-07 | NCT Delhi | Regional | Low | 17409 | <18 (n=3013); 18-49 (n=10544); =>50 (n=3392) | 52.3 | Probability | Household and community samples | 24.6% (23.9-25.2%) | NR \| ERBALISA COVID-19 IgG \| IgG | 99.1%, 99.3% (Test developer / manufacturer evaluation) | 24.3% (20.3-28.1%) |
| Kshatri (Indian Council of Medical Research-Regional Medical Research Centre in Bhubaneswar (ICMR-RMRC) Serosurvey group)[529] | 2020-08-01 to 2020-08-31 | Odisha \| Bhubaneswar, Berhampur, Rourkela | Local | High | 4146 | Mean 44.2 (NR) | 37.5 | Probability | Household and community samples | 20.8% (19.6-22.1%) | NR \| Elecsys® Anti-SARS-CoV-2 \| NR | 0.9%, 1.0% (FINDDx / MUHC independent evaluation) | - |
| Singh (The Times of India)[530] | 2020-08-27 to 2020-08-30 | Madhya Pradesh | Regional | Unclear | 3000 | NR | NR | Non-probability | Household and community samples | 11.9% | NR \| NR \| NR | No data | - |
| Mohanan (Mapmygenome)[531] | 2020-06-15 to 2020-08-29 | Karnataka | Regional | Moderate | 1197 | 12 or 15-24 (n = 192) ; 25-39 (n=363) ; 40-59 (n = 742) ; >= 60 (n = 111) | NR | Probability | Household and community samples | 39.6% (35.7-43.4%) | India; Translational Health Science and Technology Institute \| NR \| IgG | 84.7%, 100.0% (Test developer / manufacturer evaluation) | 48.1% (39.9-59.3%) |
| Dixit (Government Medical School, Aurangabad)[533] | 2020-08-10 to 2020-08-19 | Maharashtra | Local | Moderate | 6571 | 18-20 (n = 367); 21-30 (n = 1732); 31-40 (n = 1844); 41-50 (n = 1356); 51-60 (n = 761); 61-70 (n = 427); 71-80 | NR | Probability | Household and community samples | 5.8% | NR \| NR \| IgG | 75.6%, 97.6% (Used Bastos SR/MA data; no sens, spec, or author adjustment available) | 4.5% (2.9-5.8%) |

| Article Author (Study Organization) | Sampling Dates (YMD) | Location | Geographic Scope | Overall Risk of Bias | Sample Size | Age | Female (%) | Sampling method | Sample frame | Uncorrected Seroprevalence (95% Confidence Interval) | Test Manufacturers \| Names \| Isotypes | Test Sens, Spec (Source) | Corrected Seroprevalence (95% Credible Interval) |
|---|---|---|---|---|---|---|---|---|---|---|---|---|---|
| | | | | | | (n = 99): >80 (n = 23) | | | | | | | |
| Ray (All India Institute of Medical Sciences)[535] | 2020-06-08 to 2020-08-08 | Punjab \| New Delhi | Local | High | 212 | Mean 41.2 (SD 15.4) | 44.8 | Non-probability | Residual sera | 19.8% (14.7-25.8%) | "Translational Health Science and Technology Institute (THSTI); Faridabad" \| THSTI ELISA IgG \| IgG | 88.2%, 99.8% (Test developer / manufacturer evaluation) | 21.0% (13.8-30.1%) |
| Satpati (Medical College Kolkata)[536] | 2020-07-26 to 2020-08-08 | West Bengal | Regional | Moderate | 458 | 0-5 (n=1); 11-19 (n=28); 20-45 (n=290); 46-59 (n=101); 60 and above (n=38) | 37.3 | Probability | Household and community samples | 4.2% (2.5-6.4%) | Calbiotech Inc. \| ErbaLisa COVID-19 IgG ELISA Kit \| IgG | 98.3%, 98.1% (Test developer / manufacturer evaluation) | 2.0% (0.0-4.6%) |
| Bedi (National Centre for Disease Control)[537] | 2020-08-01 to 2020-08-07 | NCT Delhi | Regional | Unclear | 15289 | NR | NR | Non-probability | Blood donors | 28.3% | NR \| NR \| NR | No data | - |
| Sharma (Government of National Capital Territory of Delhi)[524] | 2020-08-01 to 2020-08-07 | NCT Delhi | Regional | Low | 15046 | <18 (n=2446); 18-49 (n=9191); =>50 (n=3036) | 49.5 | Non-probability | Household and community samples | 28.8% (28.1-29.6%) | NR \| ELISA COVID-Kawach IgG \| IgG | 92.1%, 97.7% (Test developer / manufacturer evaluation) | 30.1% (25.2-37.0%) |
| Dey (National Centre for Disease Control)[538] | 2020-08-01 to 2020-08-07 | NCT Delhi \| New Delhi | Local | Unclear | 15311 | NR | NR | Non-probability | Household and community samples | 29.1% (28.5-29.9%) | ICMR-National Institute of Virology \| KOVID KAVACH ELISA test \| IgG | 92.4%, 97.9% (Test developer / manufacturer evaluation) | 30.4% (24.1-36.7%) |
| Ghose (Indian Institute of Science Education and Research)[539] | 2020-07-20 to 2020-08-05 | Maharashtra \| Pune | Local | Moderate | 1659 | 18-30 (n=394); 31-50 (n=677); 51-65 (n=417); >65 (n=171) | 48.3 | Probability | Household and community samples | 51.3% (39.9-62.4%) | In-house developed. \| THSTI RBD-IgG ELISA \| IgG | 75.6%, 97.6% (Used Bastos SR/MA data; no sens, spec, or author adjustment available) | 66.8% (62.4-70.8%) |
| Malani (Tata Institute for | 2020-06-29 to 2020-07-19 | Mumbai | Local | Low | 2702 | 12-24 (n=176), 25-39 (n=733), 40-60 | 29.7 | Probability | Household and | 17.1% (15.8-18.4%) | Abbott Laboratories \| Abbott | 90.0%, 99.0% (Used author- | 17.1% (15.7-18.6%) |

| Article Author (Study Organization) | Sampling Dates (YMD) | Location | Geographic Scope | Overall Risk of Bias | Sample Size | Age | Female (%) | Sampling method | Sample frame | Uncorrected Seroprevalence (95% Confidence Interval) | Test Manufacturers \| Names \| Isotypes | Test Sens, Spec (Source) | Corrected Seroprevalence (95% Credible Interval) |
|---|---|---|---|---|---|---|---|---|---|---|---|---|---|
| Fundamental Research)[541] | | | | | | (n=1349), 61+ (n=444) | | | community samples | | Diagnostics Architect \| IgG | adjusted estimate) | |
| Husain (King George's Medical University)[542] | 2020-07-05 to 2020-07-19 | Uttar Pradesh \| Lucknow | Local | High | 1235 | NR | NR | Non-probability | Blood donors | 3.7% | NR \| NR \| IgG | No data | - |
| India.com News Desk (Thyrocare)[543] | 2020-06-28 to 2020-07-19 | | National | High | 53000 | NR | NR | Probability | Household and community samples | 15.0% | NR \| NR \| NR | No data | - |
| Khan (Government Medical College Srinagar)[544] | 2020-07-01 to 2020-07-15 | Srinagar \| District Srinagar | Regional | Moderate | 2906 | <30 (n=836); 30-49 (n=1424); 50-69 (n=556); >70 (n=90) | 49.7 | Non-probability | Residual sera | 3.6% (2.9-4.3%) | Abbott \| ARCHITECT i2000SR analyzer \| IgG | 0.4%, 1.0% (FINDDx / MUHC independent evaluation) | - |
| Mishra (Haryana Health Department)[545] | 2020-06-21 to 2020-07-15 | Haryana \| Panchkula , Sonipat, Faridabad, Gurugram , Ambala | Regional | High | 1544 | NR | NR | Probability | Household and community samples | 12.0% | NR \| NR \| IgG | 75.6%, 97.6% (Used Bastos SR/MA data; no sens, spec, or author adjustment available) | 13.0% (10.5-15.6%) |
| Saxena (Indian Center for Disease Control)[547] | 2020-06-27 to 2020-07-10 | NCT Delhi | Regional | Moderate | 21387 | NR | NR | Probability | Household and community samples | 23.5% | Zydus Cadila Healthcare \| COVID KAVACH \| IgG | 98.7%, 100.0% (Test developer / manufacturer evaluation) | 24.0% (20.9-27.9%) |
| Dey (National Centre for Disease Control)[538] | 2020-06-26 to 2020-07-10 | NCT Delhi \| New Delhi | Local | Unclear | 21387 | NR | NR | Non-probability | Household and community samples | 23.5% (23.0-24.0%) | ICMR-National Institute of Virology \| KOVID KAVACH ELISA test \| IgG | 92.4%, 97.9% (Test developer / manufacturer evaluation) | 23.8% (18.6-29.2%) |
| Prakash ( Municipal Medical College )[548] | 2020-06-16 to 2020-07-07 | Gujarat \| Ahmedabad | Local | High | 29891 | Mean 39.1 (SD 14.54) | 46.3 | Non-probability | Residual sera | 17.6% (17.2-18.0%) | Zydus Diagnostics \| ‘Covid Kavach \| IgG | 92.4%, 97.9% (Test developer / manufacturer evaluation) | 16.9% (12.0-21.6%) |
| Kumar (Thiruvanathapuram Health Department)[552] | 2020-06-09 to 2020-06-12 | Kerala \| Thiruvananthapuram | Local | Unclear | 450 | NR | NR | Non-probability | Household and community samples | 2.7% | NR \| NR \| IgG, IgM | No data | - |

| Article Author (Study Organization) | Sampling Dates (YMD) | Location | Geographic Scope | Overall Risk of Bias | Sample Size | Age | Female (%) | Sampling method | Sample frame | Uncorrected Seroprevalence (95% Confidence Interval) | Test Manufacturers \| Names \| Isotypes | Test Sens, Spec (Source) | Corrected Seroprevalence (95% Credible Interval) |
|---|---|---|---|---|---|---|---|---|---|---|---|---|---|
| Murhekar (Indian Council of Medical Research)[554] | 2020-05-11 to 2020-06-04 | | National | Low | 28000 | Mean 45.3 (SD 15.2) | 51.4 | Probability | Household and community samples | 0.6% (0.3-1.0%) | Cadila Healthcare Limited; EUROIMMUN \| COVID Kavach-Anti-SARS-CoV-2 IgG Antibody Detection ELISA, Euroimmun Anti-SARS-CoV-2 ELISA IgG \| IgG | 75.6%, 97.6% (Used Bastos SR/MA data; no sens, spec, or author adjustment available) | 0.0% (0.0-0.1%) |
| Kher (Indian Council of Medical Research)[555] | 2020-05-11 to 2020-05-25 | Uttarakhand, Kashmir, Rajasthan, Jharkhand, Punjab, Gujarat, Haryana, Assam, Telangana, Maharashtra, Madhya Pradesh, Jammu, Chhattisgarh, West Bengal, Tamil Nadu, Bihar, Uttar Pradesh, Himachal Pradesh, Kerala, Karnataka, Andhra Pradesh, Odisha | National | Moderate | 26400 | NR | NR | Probability | Household and community samples | 0.7% | ICMR-National Institute of Virology \| NR \| IgG | 75.6%, 97.6% (Used Bastos SR/MA data; no sens, spec, or author adjustment available) | 0.0% (0.0-0.1%) |
| **Pakistan** | | | | | | | | | | | | | |
| Nisar (Aga Khan University)[556] | 2020-08-17 to 2020-08-22 | Sindh \| Karachi | Sublocal | Moderate | 500 | Mean 27.1 (SD 17.7) | 57.8 | Probability | Household and community samples | 21.5% (15.6-28.0%) | Roche \| Elecsys Anti-SARS-Cov2 | 100.0%, 99.8% (Used author-adjusted estimate) | 21.5% (18.0-25.2%) |

| Article Author (Study Organization) | Sampling Dates (YMD) | Location | Geographic Scope | Overall Risk of Bias | Sample Size | Age | Female (%) | Sampling method | Sample frame | Uncorrected Seroprevalence (95% Confidence Interval) | Test Manufacturers \| Names \| Isotypes | Test Sens, Spec (Source) | Corrected Seroprevalence (95% Credible Interval) |
|---|---|---|---|---|---|---|---|---|---|---|---|---|---|
| | | | | | | | | | | | Immunoassay \| IgG, IgM | | |
| Nisar (Aga Khan University)[556] | 2020-08-17 to 2020-08-22 | Sindh \| Karachi | Sublocal | Moderate | 501 | Mean 26 (SD 16.7) | 58.9 | Probability | Household and community samples | 12.8% (8.3-17.7%) | Roche \| Elecsys Anti-SARS-Cov2 Immunoassay \| IgG, IgM | 100.0%, 99.8% (Used author-adjusted estimate) | 12.8% (10.1-15.9%) |
| Younas (National Institute of Blood Diseases & Bone Marrow Transplantation)[558] | 2020-05-15 to 2020-07-15 | Sindh \| Karachi | Local | High | 370 | Mean 30.6 (SD 6.3) | 0.0 | Non-probability | Blood donors | 34.6% | Roche Diagnostics \| Elecsys Anti-SARS-Cov2 Immunoassay \| IgG, IgA, IgM | 0.6%, 1.0% (FINDDx / MUHC independent evaluation) | - |
| Nisar (Aga Khan University)[556] | 2020-06-25 to 2020-07-11 | Sindh \| Karachi | Sublocal | Moderate | 504 | Mean 24.3 (SD 16.7) | 60.5 | Probability | Household and community samples | 8.7% (5.1-13.1%) | Roche \| Elecsys Anti-SARS-Cov2 Immunoassay \| IgG, IgM | 100.0%, 99.8% (Used author-adjusted estimate) | 8.7% (6.3-11.2%) |
| Nisar (Aga Khan University)[556] | 2020-06-25 to 2020-07-11 | Sindh \| Karachi | Sublocal | Moderate | 500 | Mean 25.9 (SD 16.7) | 55.0 | Probability | Household and community samples | 15.1% (9.4-21.7%) | Roche \| Elecsys Anti-SARS-Cov2 Immunoassay \| IgG, IgM | 100.0%, 99.8% (Used author-adjusted estimate) | 15.1% (12.1-18.3%) |
| Nisar (Aga Khan University)[556] | 2020-04-15 to 2020-04-25 | Sindh \| Karachi | Sublocal | Moderate | 500 | Mean 28.5 (SD 17.9) | 58.6 | Probability | Household and community samples | 0.2% (0.0-0.7%) | Roche \| Elecsys Anti-SARS-Cov2 Immunoassay \| IgG, IgM | 100.0%, 99.8% (Used author-adjusted estimate) | 0.2% (0.0-0.9%) |
| Nisar (Aga Khan University)[556] | 2020-04-15 to 2020-04-25 | Sindh \| Karachi | Sublocal | Moderate | 500 | Mean 26.2 (SD 17.9) | 48.8 | Probability | Household and community samples | 0.4% (0.0-1.3%) | Roche \| Elecsys Anti-SARS-Cov2 Immunoassay \| IgG, IgM | 100.0%, 99.8% (Used author-adjusted estimate) | 0.4% (0.1-1.3%) |
| **Southeast Asia, East Asia, and Oceania** | | | | | | | | | | | | | |
| **China** | | | | | | | | | | | | | |
| Wang (Capital Medical University )[563] | 2020-01-01 to 2020-08-31 | Beijing | Sublocal | Moderate | 19797 | <1 (n = 2461) ; 1-<3 (n = 4383) ; 3-<6 | 54.4 | Non-probability | Residual sera | 0.1% | Beijing Innovita Biological Technology \| | 0.5%, 1.0% (FINDDx / MUHC | - |

| Article Author (Study Organization) | Sampling Dates (YMD) | Location | Geographic Scope | Overall Risk of Bias | Sample Size | Age | Female (%) | Sampling method | Sample frame | Uncorrected Seroprevalence (95% Confidence Interval) | Test Manufacturers \| Names \| Isotypes | Test Sens, Spec (Source) | Corrected Seroprevalence (95% Credible Interval) |
|---|---|---|---|---|---|---|---|---|---|---|---|---|---|
| | | | | | | (n=4334) ; >= 6 (n = 8619) | | | | | Innovita 2019-nCoV Ab Test (Colloidal Gold) \| IgG | independent evaluation) | |
| He (Renmin Hospital of Wuhan University)[565] | 2020-05-15 to 2020-05-29 | Hubei \| Wuhan | Local | Unclear | 4454 | Median 36.89 (SD 10.811) | 50.2 | Non-probability | Household and community samples | 2.9% | Livzon Diagnostics Inc. \| Diagnostic Kit for IgM/IgG Antibody \| IgG | 0.5%, 0.6% (FINDDx / MUHC independent evaluation) | - |
| Liu (Huazhong University of Science and Technology)[566] | 2020-03-27 to 2020-05-26 | Hubei \| Wuhan | Local | High | 35040 | Median 36 (IQR 30-45) | 50.7 | Non-probability | Household and community samples | 3.9% (3.7-4.1%) | China; Bioscience; Tianjin \| MCLIA kits \| IgG, IgM | 0.6%, 1.0% (FINDDx / MUHC independent evaluation) | - |
| Pan (Wuhan University)[567] | 2020-05-15 to 2020-05-26 | Hubei \| Wuhan | Local | Moderate | 61437 | Median 48 (IQR 32-64) | 51.1 | Probability | Household and community samples | 2.4% (2.3-2.5%) | Ltd; Nanjing Vazyme Medical technology Co. \| a 2019-nCoV IgG/IgM detection kit (catalogue number: C6603C; Colloidal Gold-Based \| IgG, IgM | 91.5%, 97.0% (Test developer / manufacturer evaluation) | 0.6% (0.0-1.7%) |
| Chang (Beijing Hospital)[570] | 2020-01-15 to 2020-04-30 | Hubei \| Wuhan | Local | High | 17794 | Median 33 (IQR 19-47) | 37.7 | Non-probability | Blood donors | 2.3% (2.1-2.5%) | Wantai Biological Pharmacy Enterprise \| Wantai SARS CoV-2 Total Antibody ELISA \| IgG, IgA, IgM | 0.9%, 1.0% (FINDDx / MUHC independent evaluation) | - |
| Liu (Zhongnan Hospital of Wuhan University)[571] | 2020-02-29 to 2020-04-29 | Hubei \| Wuhan | Local | Moderate | 19555 | Mean 41.6 (95% CI 41.4-41.8) | 50.2 | Non-probability | Household and community samples | 4.6% (4.3-4.9%) | YHLO \| SARS-CoV-2 IgM/IgG CLIA kits \| IgG | 0.7%, 1.0% (FINDDx / MUHC independent evaluation) | - |
| Liu (Zhongnan Hospital of | 2020-02-29 to 2020-04-29 | Hubei \| Wuhan | Local | Moderate | 1616 | Mean 53.3 (95% CI 52.4-54.2) | 56.2 | Non-probability | Residual sera | 1.0% (0.5-1.5%) | YHLO \| SARS-CoV-2 | 0.7%, 1.0% (FINDDx / MUHC | - |

| Article Author (Study Organization) | Sampling Dates (YMD) | Location | Geographic Scope | Overall Risk of Bias | Sample Size | Age | Female (%) | Sampling method | Sample frame | Uncorrected Seroprevalence (95% Confidence Interval) | Test Manufacturers \| Names \| Isotypes | Test Sens, Spec (Source) | Corrected Seroprevalence (95% Credible Interval) |
|---|---|---|---|---|---|---|---|---|---|---|---|---|---|
| Wuhan University)[571] | | | | | | | | | | | IgM/IgG CLIA kits \| IgG | independent evaluation) | |
| Ling (Chinese Center for Disease Control and Prevention)[573] | 2020-03-25 to 2020-04-28 | Hubei \| Wuhan | Local | Moderate | 18712 | Median 40 (Range 4-81) | 39.1 | Non-probability | Household and community samples | 3.3% (3.0-3.5%) | Innovita \| 2019-nCoV Ab Test (Colloidal Gold) \| IgG, IgM | 0.6%, 1.0% (FINDDx / MUHC independent evaluation) | - |
| Wang (Peking University)[576] | 2020-04-15 to 2020-04-18 | Hebei \| Beijing | Local | Moderate | 2184 | Mean 42.3 (SD 19.5) | 54.3 | Probability | Household and community samples | 0.2% | Innovita Biological Technology Co Ltd; Wondfo Biotech Co. \| WONDFO SARS-CoV-2 Antibody Test, Tangshan Colloidal Gold Immunochromatography Assay \| IgG, IgM | 66.0%, 96.6% (Used Bastos SR/MA data; no sens, spec, or author adjustment available) | 0.1% (0.0-0.2%) |
| Wu (Wuhan University)[577] | 2020-04-03 to 2020-04-15 | Hubei \| Wuhan | Sublocal | High | 381 | NR | NR | Non-probability | Residual sera | 10.5% | Innovita Biological Technology Co. Ltd \| 2019-nCoV Ab Test \| IgG, IgM | 0.6%, 1.0% (FINDDx / MUHC independent evaluation) | - |
| Xu (Institute of Blood Transfusion)[579] | 2020-03-23 to 2020-04-02 | Guangdong \| Guangzhou | Local | High | 2199 | Median 34 (Range 18 to 59 ) | 32.3 | Probability | Blood donors | 0.3% | Lizhu Diagnostics; Wantai Biological Pharmacy Enterprise \| Wantai SARS CoV-2 Total Antibody ELISA \| IgG, IgA | 75.6%, 97.6% (Used Bastos SR/MA data; no sens, spec, or author adjustment available) | 0.1% (0.0-0.2%) |
| To (Queen Mary Hospital)[587] | 2020-03-04 to 2020-03-17 | Hubei | Regional | Moderate | 452 | Median 41 (NR) | 58.6 | Non-probability | Household and community samples | 2.9% | Author-designed \| NR \| IgG | 73.3%, 100.0% (Test developer / manufacturer evaluation) | 3.5% (0.6-6.5%) |
| Chang (Beijing Hospital)[570] | 2020-02-02 to 2020-03-15 | Guangdong \| Shenzhen | Local | High | 6810 | Median 36 (IQR 19-53) | 35.0 | Non-probability | Blood donors | 0.0% (0.0-0.1%) | Wantai Biological Pharmacy | 0.9%, 1.0% (FINDDx / MUHC | - |

| Article Author (Study Organization) | Sampling Dates (YMD) | Location | Geographic Scope | Overall Risk of Bias | Sample Size | Age | Female (%) | Sampling method | Sample frame | Uncorrected Seroprevalence (95% Confidence Interval) | Test Manufacturers \| Names \| Isotypes | Test Sens, Spec (Source) | Corrected Seroprevalence (95% Credible Interval) |
|---|---|---|---|---|---|---|---|---|---|---|---|---|---|
| | | | | | | | | | | | Enterprise \| Wantai SARS CoV-2 Total Antibody ELISA \| IgG, IgA, IgM | independent evaluation) | |
| Chang (Beijing Hospital)[570] | 2020-01-23 to 2020-03-15 | Hebei \| Shijiazhuang | Local | High | 13540 | Median 40 (IQR 33-48) | 29.5 | Non-probability | Blood donors | 0.0% (0.0-0.0%) | Wantai Biological Pharmacy Enterprise \| Wantai SARS CoV-2 Total Antibody ELISA \| IgG, IgA, IgM | 0.9%, 1.0% (FINDDx / MUHC independent evaluation) | - |
| Zhang (Shengjing Hospital of China Medical University)[590] | 2020-01-21 to 2020-02-16 | Liaoning \| Shenyang | Regional | High | 225 | Median 35 (Range 1-86) | 71.1 | Non-probability | Residual sera | 0.4% | Shenzhen Yahuilong Biotechnology Co. \| Chemiluminescence Detection Kit \| IgG | 0.9%, 1.0% (FINDDx / MUHC independent evaluation) | - |
| Zhang (Shengjing Hospital of China Medical University)[590] | 2020-01-21 to 2020-02-16 | Liaoning \| Shenyang | Regional | High | 222 | Median 50 (Range 27-85) | 72.1 | Non-probability | Residual sera | 0.9% | Shenzhen Yahuilong Biotechnology Co. \| Chemiluminescence Detection Kit \| IgG | 0.9%, 1.0% (FINDDx / MUHC independent evaluation) | - |
| To (Queen Mary Hospital)[587] | 2020-02-01 to 2020-02-13 | Hong Kong | Regional | High | 233 | NR | NR | Non-probability | Residual sera | 0.4% (0.0-2.4%) | Author-designed \| NR \| IgG | 73.3%, 100.0% (Test developer / manufacturer evaluation) | 0.8% (0.0-2.6%) |
| To (Queen Mary Hospital)[587] | 2020-01-01 to 2020-01-31 | Hong Kong | Regional | High | 580 | NR | NR | Non-probability | Residual sera | 2.6% (1.5-4.2%) | Author-designed \| NR \| IgG | 73.3%, 100.0% (Test developer / manufacturer evaluation) | 3.2% (0.8-5.7%) |
| Hallowell (US Centers for Disease Control and Prevention)[591] | 2020-01-28 to 2020-01-28 | Hubei \| Wuhan | Sublocal | High | 186 | Median 42.0 (NR) | 47.8 | Non-probability | Household and community samples | 0.5% | SeraCare Life Sciences \| HRP-conjugated anti human IgM/IgA/IgG \| IgG, IgA, IgM | 75.6%, 97.6% (Used Bastos SR/MA data; no sens, spec, or author adjustment available) | 0.6% (0.0-2.4%) |

| Article Author (Study Organization) | Sampling Dates (YMD) | Location | Geographic Scope | Overall Risk of Bias | Sample Size | Age | Female (%) | Sampling method | Sample frame | Uncorrected Seroprevalence (95% Confidence Interval) | Test Manufacturers \| Names \| Isotypes | Test Sens, Spec (Source) | Corrected Seroprevalence (95% Credible Interval) |
|---|---|---|---|---|---|---|---|---|---|---|---|---|---|
| **Malaysia** | | | | | | | | | | | | | |
| Sam (University Malaya)[592] | 2020-01-29 to 2020-06-06 | Selangor \| Kuala Lumpur | Regional | High | 588 | NR | 62.6 | Non-probability | Residual sera | 0.4% (0.0-0.9%) | Author-designed \| N/A - Author-designed \| IgG | 97.0%, 100.0% (Test developer / manufacturer evaluation) | 0.4% (0.0-1.0%) |
| **Sub-Saharan Africa** | | | | | | | | | | | | | |
| **Congo** | | | | | | | | | | | | | |
| Batchi-Bouyou (Fondation Congolaise pour la Recherche Médicale)[596] | 2020-04-01 to 2020-07-31 | Brazzaville | Local | High | 754 | Mean 36.7 (SD 13.07) | 38.6 | Non-probability | Household and community samples | 19.7% | Wuhan UNniscience Biotechnology; Salofa Oy manufacturer; Finland \| UNsiences COVID-19 IgG/IgM rapid test, Sienna™ \| IgG, IgM | 66.0%, 96.6% (Used Bastos SR/MA data; no sens, spec, or author adjustment available) | 26.0% (21.3-30.8%) |
| **Ethiopia** | | | | | | | | | | | | | |
| Kempen (Kuwait University)[597] | 2020-05-18 to 2020-05-21 | Addis Ababa | Local | High | 99 | Mean 37.8 (SARS-CoV-2 IgG negative), 33.3 (SARS-CoV-2 IgG positive) (NR) | 55.6 | Non-probability | Residual sera | 3.0% (0.6-8.6%) | Abbott \| Abbott IgG Architect \| IgG | 0.4%, 1.0% (FINDDx / MUHC independent evaluation) | - |
| Alemu (Ministry of Health, Ethiopia)[598] | 2020-04-23 to 2020-04-28 | Addis Ababa | Local | Moderate | 301 | Median 30 (SD 10.9) | 37.5 | Probability | Household and community samples | 8.8% (5.5-11.6%) | China; Zhejiang Orient Gene Biotech Co Ltd; Zhejiang; Huzhou \| Immunolateral flow immunoassay kit \| IgG, IgM | 87.9%, 100.0% (Used author-adjusted estimate) | 8.8% (5.9-12.2%) |
| **Keyna** | | | | | | | | | | | | | |
| Adetifa (KEMRI-Wellcome | 2020-04-30 to 2020-09-30 | | National | Low | 9922 | 15-24 n = 2,763 25-34 n = 3,902 35-44 | 19.2 | Non-probability | Blood donors | 9.4% (8.8-9.9%) | N/A - in-house \| N/A - in-house \| IgG | 92.7%, 99.0% (Author-reported | 9.0% (8.1-10.0%) |

| Article Author (Study Organization) | Sampling Dates (YMD) | Location | Geographic Scope | Overall Risk of Bias | Sample Size | Age | Female (%) | Sampling method | Sample frame | Uncorrected Seroprevalence (95% Confidence Interval) | Test Manufacturers \| Names \| Isotypes | Test Sens, Spec (Source) | Corrected Seroprevalence (95% Credible Interval) |
|---|---|---|---|---|---|---|---|---|---|---|---|---|---|
| Trust Research Programme)[599] | | | | | | n = 2,261 45-54 n = 794 55-64 n = 202 | | | | | | independent evaluation) | |
| **Nigeria** | | | | | | | | | | | | | |
| Majiya (Ibrahim Badamasi Babangida University)[601] | 2020-06-26 to 2020-06-30 | Niger State | Regional | Moderate | 185 | NR | 44.3 | Probability | Household and community samples | 25.4% | Cambridge Network \| COVID-19 IgG and IgM Rapid Test \| IgG | 100.0%, 100.0% (Author-reported independent evaluation) | 19.5% (0.5-31.9%) |
| **South Africa** | | | | | | | | | | | | | |
| Hsiao ( University of Cape Town)[604] | 2020-06-15 to 2020-08-07 | Cape Town | Local | High | 2791 | < 20 (n = 151) ; 20-50 (n = 2349) ; >50 (n = 291) | 80.3 | Non-probability | Residual sera | 40.2% (38.4-42.1%) | Roche Diagnostics \| ElecsysTM anti-SARS-CoV-2 assay \| IgG, IgM | 0.8%, 1.0% (FINDDx / MUHC independent evaluation) | 59.8% (57.9-61.8%) |

#### S2 Table. Characteristics and primary results of studies reporting population-specific seroprevalence estimates

| Article Author (Study Organization) | Sampling Dates (YMD) | Location | Geographic Scope | Overall Risk of Bias | Sample Size | Age | Female (%) | Sampling method | Sample frame | Uncorrected Seroprevalence (95% Confidence Interval) | Test Manufacturers \| Names \| Isotypes | Test Sens, Spec (Source) | Corrected Seroprevalence (95% Credible Interval) |
|---|---|---|---|---|---|---|---|---|---|---|---|---|---|
| **Central Europe, Eastern Europe, and Central Asia** | | | | | | | | | | | | | |
| **Armenia** | | | | | | | | | | | | | |
| Grigoryan (Ministry of Health of the Republic of Armenia)[1] | 2020-06-29 | Yerevan \| Yerevan | Local | High | 380 | NR | NR | Non-probability | Health care workers and caregivers | 16.1% | NR \| NR \| NR | 100.0%, 99.8% (Test developer / manufacturer evaluation) | 14.4% (9.4-19.3%) |
| **Croatia** | | | | | | | | | | | | | |
| Vince (Croatian Football Federation)[3] | 2020-05-29 to 2020-07-31 | | Sublocal | Moderate | 305 | Mean 28.5 (SD 9.1) | 0.0 | Non-probability | Non-essential workers and unemployed persons | 20.0% | Euroimmun Medizinische Labordiagnostika AG \| Anti-SARS-CoV-2 IgA, Anti-SARS-CoV-2 IgG S and Anti-SARS-CoV-2 IgG NCP \| IgG, IgA | 75.6%, 97.6% (Used Bastos SR/MA data; no sens, spec, or author adjustment available) | 24.1% (18.3-30.8%) |
| Lapić (Zagreb University)[4] | 2020-05-01 to 2020-05-31 | Zagreb | Sublocal | Moderate | 1678 | Median 43 (NR) | 78.2 | Non-probability | Health care workers and caregivers | 0.6% | Abbott Laboratories \| Abbott SARS-CoV-2 IgG assay \| IgG | 0.4%, 1.0% (FINDDx / MUHC independent evaluation) | - |
| Vilibic-Cavlek (University of Zagreb)[5] | 2020-04-25 to 2020-05-24 | | National | High | 592 | 20-65 | 74.3 | Non-probability | Health care workers and caregivers | 2.7% | Vircell Spain S.L.U \| NR \| IgG | 0.7%, 1.0% (FINDDx / MUHC independent evaluation) | - |
| Jerkovic (University of Split)[6] | 2020-04-23 to 2020-04-28 | | Local | High | 1494 | Median 46 (Range 19-79) | 11.9 | Non-probability | Essential non-healthcare workers | 1.3% (0.8-2.0%) | AMP Diagnostics \| AMP Rapid Test SARS-CoV-2 IgG/IgM \| IgG, IgM | 91.8%, 96.4% (Test developer / manufacturer evaluation) | 0.3% (0.0-1.1%) |
| **Czechia** | | | | | | | | | | | | | |

| Article Author (Study Organization) | Sampling Dates (YMD) | Location | Geographic Scope | Overall Risk of Bias | Sample Size | Age | Female (%) | Sampling method | Sample frame | Uncorrected Seroprevalence (95% Confidence Interval) | Test Manufacturers \| Names \| Isotypes | Test Sens, Spec (Source) | Corrected Seroprevalence (95% Credible Interval) |
|---|---|---|---|---|---|---|---|---|---|---|---|---|---|
| Malickova (Clinical and Research Center for Inflammatory Bowel Disease)[8] | Until 2020-04-15 | Central Bohemian \| Prague | Sublocal | Unclear | 92 | Median 45 (IQR 38-57 ) | 71.7 | Non-probability | Health care workers and caregivers | 2.2% | EUROIMMUN \| Euroimmun Anti-SARS-CoV-2 ELISA IgG \| IgG | 0.8%, 1.0% (FINDDx / MUHC independent evaluation) | - |
| **Estonia** | | | | | | | | | | | | | |
| Veerus (University of Tartu)[10] | 2020-05-04 to 2020-06-10 | | National | High | 433 | Mean 31 (SD 5.89) | 100.0 | Non-probability | Pregnant or parturient women | 0.5% | Abbott \| SARS-CoV-2 IgG Architect \| IgG | 0.4%, 1.0% (FINDDx / MUHC independent evaluation) | - |
| **Poland** | | | | | | | | | | | | | |
| Ciechanowicz (Central ClinicalHospital MSWiA)[13] | 2020-04-01 to 2020-10-31 | Warsaw | Sublocal | High | 61 | Mean 46 (SD 14) | 34.4 | Non-probability | Patients seeking care for non-COVID-19 reasons | 16.4% | Vircell Microbiologist; Spain:; Granada \| Vircell anti-SARS-CoV-2 IgM, IgA and IgG ELISA; ref. MA1032 and G103 \| IgG, IgA, IgM | 100.0%, 95.0% (Test developer / manufacturer evaluation) | 10.6% (1.0-21.1%) |
| Kasztelewicz (The Children's Memorial Health Institute)[14] | 2020-07-01 to 2020-08-09 | Masovia \| Warsaw | Sublocal | Moderate | 1879 | Median 48 (IQR 38-56) | 85.8 | Non-probability | Health care workers and caregivers | 0.9% | Abbott Laboratories \| Abbott SARS-CoV-2 IgG assay \| IgG | 0.4%, 1.0% (FINDDx / MUHC independent evaluation) | - |
| Gujski (Medical University of Warsaw)[15] | 2020-06-22 to 2020-07-08 | Mazowiecki e Province | Regional | Moderate | 5082 | Mean 39.6 (SD 9) | 33.5 | Probability | Essential non-healthcare workers | 4.3% (3.7-4.9%) | Vircell S.L \| COVID-19 ELISA IgG Kit \| IgG | 0.7%, 1.0% (FINDDx / MUHC independent evaluation) | - |
| **Romania** | | | | | | | | | | | | | |
| MedLife (MedLife)[16] | 2020-03-24 to 2020-04-21 | Nord-Vest, Bucuresti-Ilfov, Sud-Vest Oltenia, Vest, Sud-Mutenia, | National | Unclear | 371 | NR | NR | Non-probability | Health care workers and caregivers | 3.0% | Maglumi; Abbott; YHLO \| Multiple tests used in combination \| IgG, IgM | No data | - |

| Article Author (Study Organization) | Sampling Dates (YMD) | Location | Geographic Scope | Overall Risk of Bias | Sample Size | Age | Female (%) | Sampling method | Sample frame | Uncorrected Seroprevalence (95% Confidence Interval) | Test Manufacturers \| Names \| Isotypes | Test Sens, Spec (Source) | Corrected Seroprevalence (95% Credible Interval) |
|---|---|---|---|---|---|---|---|---|---|---|---|---|---|
| | | Sud-Est, Centru, Nord-Est \| Brasov, Sibiu, Timisoara, Pitesti, Targoviste, Arad, Galati, Braila, Ploiesti, Bucharest, Piatra Neamt | | | | | | | | | | | |
| **Russian Federation** | | | | | | | | | | | | | |
| Krechetova (Ministry of Health of Russia)[24] | 2020-04-15 to 2020-06-15 | Moscow | Sublocal | Unclear | 1589 | NR | NR | Non-probability | Health care workers and caregivers | 8.9% | "NMITs Hematology" Ministry of Health of the Russian Federation \| SARS-CoV-2-IgG-ELISA \| IgG | 75.6%, 97.6% (Used Bastos SR/MA data; no sens, spec, or author adjustment available) | 8.8% (6.4-11.1%) |
| **High Income** | | | | | | | | | | | | | |
| **Argentina** | | | | | | | | | | | | | |
| Silva (Instituto Nacional de Epidemiologia)[28] | 2020-06-03 to 2020-07-06 | Buenos Aires \| Buenos Aires | Sublocal | Moderate | 738 | NR | NR | Probability | Health care workers and caregivers | 0.8% (0.0-8.1%) | CONICET \| COVIDAR IgG ELISA, COVIDAR IgM Test \| IgG, IgM | 75.6%, 97.6% (Used Bastos SR/MA data; no sens, spec, or author adjustment available) | 0.2% (0.0-0.8%) |
| Figar (Minister of Health, Buenos Aires City)[29] | 2020-06-10 to 2020-07-01 | Buenos Aires \| Buenos Aires | Sublocal | High | 60 | NR | NR | Non-probability | Persons living in slums | 36.0% | Laboratio Lemos \| COVIDAR IgG ELISA \| IgG | 95.0%, 100.0% (Test developer / manufacturer evaluation) | 38.4% (24.2-51.8%) |
| Insúa (Hospital General de Niños Pedro de Elizalde)[30] | 2020-06-08 to 2020-06-09 | Province of Buenos Aires \| Autonomous City of Buenos Aires | Sublocal | High | 116 | Mean 45.6 (SD 13.3) | NR | Probability | Health care workers and caregivers | 0.9% (0.1-5.5%) | Snibe (Shenzen New Industries) \| MAGLUMI 2019-nCoV IgM and 2019-nCoV IgG \| IgG, IgM | 95.6%, 96.0% (Test developer / manufacturer evaluation) | 0.8% (0.0-3.4%) |

| Article Author (Study Organization) | Sampling Dates (YMD) | Location | Geographic Scope | Overall Risk of Bias | Sample Size | Age | Female (%) | Sampling method | Sample frame | Uncorrected Seroprevalence (95% Confidence Interval) | Test Manufacturers \| Names \| Isotypes | Test Sens, Spec (Source) | Corrected Seroprevalence (95% Credible Interval) |
|---|---|---|---|---|---|---|---|---|---|---|---|---|---|
| **Australia** | | | | | | | | | | | | | |
| Hicks (Australian National University)[31] | 2020-05-15 to 2020-06-15 | | National | Unclear | 2991 | Mean 54 (Range 15-95) | 51.2 | Non-probability | Patients seeking care for non-COVID-19 reasons | 1.4% | In House \| Dual-antigen IgG ELISA \| IgG, IgA, IgM | 100.0%, 98.9% (Test developer / manufacturer evaluation) | 0.4% (0.0-1.0%) |
| Macartney (University of Sydney)[32] | 2020-01-25 to 2020-04-10 | New South Wales | Regional | Moderate | 173 | NR | NR | Non-probability | Contacts of COVID patients | 2.3% | Author-designed \| N/A - Author-designed \| IgG, IgA, IgM | 91.3%, 98.9% (Test developer / manufacturer evaluation) | 1.3% (0.0-3.9%) |
| **Austria** | | | | | | | | | | | | | |
| Fuereder (Medical University of Vienna)[34] | 2020-04-01 to 2020-06-04 | Vienna \| Vienna | Sublocal | High | 62 | Median 41 (Range 23-59) | 71.0 | Non-probability | Health care workers and caregivers | 3.2% (0.4-11.2%) | Abbott; Roche \| Elecsys Anti-SARS-CoV-2 kit, Abbott Architect SARS-CoV-2 IgG assay \| IgG | 97.8%, 97.8% (Used Bastos SR/MA data; no sens, spec, or author adjustment available) | 1.7% (0.0-6.4%) |
| Fuereder (Medical University of Vienna)[34] | 2020-03-21 to 2020-06-04 | Vienna \| Vienna | Sublocal | High | 84 | Median 61 (NR) | 48.8 | Non-probability | Patients seeking care for non-COVID-19 reasons | 2.4% (0.3-8.3%) | Abbott; Roche \| Elecsys Anti-SARS-CoV-2 kit, Abbott Architect SARS-CoV-2 IgG assay \| IgG | 97.8%, 97.8% (Used Bastos SR/MA data; no sens, spec, or author adjustment available) | 1.8% (0.0-5.9%) |
| Hackner (Karl Landsteiner University of Health Sciences)[35] | 2020-04-01 to 2020-04-30 | | Sublocal | High | 130 | Median 41 (Range 19-64) | 76.9 | Non-probability | Health care workers and caregivers | 2.3% | Epitope Diagnostics \| NR \| IgG, IgM | 0.8%, 0.9% (FINDDx / MUHC independent evaluation) | - |
| Reiter (Medical University of Vienna)[38] | 2020-03-15 to 2020-03-29 | Vienna | Sublocal | High | 235 | Mean 44.2 (SD 11.4) | 70.2 | Non-probability | Health care workers and caregivers | 25.5% (20.4-31.5%) | ImmunoDiagnostics \| ImmunoDiagnostics test system IgG IgM \| IgG, IgM | 75.6%, 97.6% (Used Bastos SR/MA data; no sens, spec, or author adjustment available) | 31.1% (23.9-39.0%) |
| Orth-Höller (Innsbruck Clinical | 2020-03-20 to 2020-03-27 | Tyrol | Regional | High | 377 | Median 51 (NR) | 46.7 | Non-probability | Health care workers and caregivers | 0.3% (0.0-1.5%) | EUROIMMUN \| Euroimmun Anti-SARS- | 0.8%, 1.0% (FINDDx / MUHC | - |

| Article Author (Study Organization) | Sampling Dates (YMD) | Location | Geographic Scope | Overall Risk of Bias | Sample Size | Age | Female (%) | Sampling method | Sample frame | Uncorrected Seroprevalence (95% Confidence Interval) | Test Manufacturers \| Names \| Isotypes | Test Sens, Spec (Source) | Corrected Seroprevalence (95% Credible Interval) |
|---|---|---|---|---|---|---|---|---|---|---|---|---|---|
| Microbiology Laboratory)[39] | | | | | | | | | | | CoV-2 ELISA IgG \| IgG | independent evaluation) | |
| **Belgium** | | | | | | | | | | | | | |
| Duysburgh (Sciensano)[40] | 2020-04-22 to 2020-09-30 | | National | Unclear | 850 | NR | NR | Non-probability | Health care workers and caregivers | 9.5% | Euroimmun Medizinische Labordiagnostika \| EUROIMMUN -Anti-SARS-CoV-2 ELISA (IgG) \| IgG | 0.6%, 0.9% (FINDDx / MUHC independent evaluation) | - |
| Blairon (Iris Hospitals South)[42] | 2020-05-25 to 2020-06-19 | Brussels \| City of Brussels | Local | High | 1485 | Median 47.45 (Males); 43.90 (Females) (Range 46.00–48.84 (Males); 42.80–45.10 (Females)) | 73.1 | Non-probability | Health care workers and caregivers | 14.6% | DiaSorin \| Liaison® SARS- CoV-2 S1/S2 IgG \| IgG | 0.8%, 1.0% (FINDDx / MUHC independent evaluation) | - |
| Berardis (Cliniques universitaires Saint-Luc)[43] | 2020-04-16 to 2020-05-19 | City of Brussels | Local | High | 149 | Mean 24.9 (SD 15) | 49.0 | Non-probability | Patients seeking care for non-COVID-19 reasons | 2.7% | "SNIBE – Shenzhen New Industries Biomedical Engineering Co.; Ltd" \| MAGLUMI 2019-nCov IgG/IgM CLIA \| IgG, IgM | 95.6%, 96.0% (Test developer / manufacturer evaluation) | 1.2% (0.0-4.1%) |
| Martin (Université Libre de Bruxelles)[44] | 2020-04-15 to 2020-05-18 | Brussels \| City of Brussels | Local | High | 326 | Mean 37 (Range 21-66) | 73.3 | Non-probability | Health care workers and caregivers | 11.0% | EUROIMMUN \| Euroimmun Anti-SARS-CoV-2 ELISA IgG \| IgG | 0.4%, 1.0% (FINDDx / MUHC independent evaluation) | - |
| Steensels (ZiekenhuisOost-Limburg)[45] | 2020-04-22 to 2020-04-30 | Limburg \| Genk | Sublocal | High | 3056 | Mean 39.5 (staff with antibodies); 41.3 (staff without antibodies) (SD 13.1 (staff with antibodies); | NR | Non-probability | Health care workers and caregivers | 6.4% (5.5-7.3%) | Multi-G \| COVID-19 IgG/IgM Rapid Test Cassette \| IgG | 0.6%, 1.0% (FINDDx / MUHC independent evaluation) | - |

| Article Author (Study Organization) | Sampling Dates (YMD) | Location | Geographic Scope | Overall Risk of Bias | Sample Size | Age | Female (%) | Sampling method | Sample frame | Uncorrected Seroprevalence (95% Confidence Interval) | Test Manufacturers \| Names \| Isotypes | Test Sens, Spec (Source) | Corrected Seroprevalence (95% Credible Interval) |
|---|---|---|---|---|---|---|---|---|---|---|---|---|---|
| | | | | | | 12.4 (staff without antibodies) ) | | | | | | | |
| Buntinx (University of Leuven)[46] | 2020-04-14 to 2020-04-16 | Limburg \| Lanaken | Sublocal | High | 188 | Mean 86 (NR) | 100.0 | Non-probability | Multiple populations | 18.6% | SureScreen Diagnostics; UK; Derby \| SureScreen Diagnostics Covid-19 IgG/IgM Rapid test cassette \| IgG, IgM | 74.7%, 95.1% (Author-reported independent evaluation) | 17.4% (6.1-27.2%) |
| **Canada** | | | | | | | | | | | | | |
| Brousseau (Institut National de Sante Publique)[50] | 2020-07-06 to 2020-09-24 | Quebec | Regional | High | 2056 | 18-29 ( n = 427) ; 30-39 (n = 643) ; 40-49 (n = 547) ; 50-59 (n = 356) ; 60+ (n = 83) | 76.1 | Non-probability | Health care workers and caregivers | 11.7% | DiaSorin \| LIAISON SARS-CoV-2 S1/S2 IgG \| IgG | 0.8%, 1.0% (FINDDx / MUHC independent evaluation) | - |
| Bardai (Shriners Hospital for Children)[52] | 2020-06-10 to 2020-07-27 | Quebec \| Montreal | Sublocal | High | 199 | Median Patients: 15.6, Accompanying persons: 47.1, Employees 42.5 (IQR Patients 13.4-16.8, accompanying persons 41.4- 50.8, employees 32.5-52.5) | 69.8 | Non-probability | Multiple populations | 11.0% | Omega Diagnostics \| Mologic, Omega Diagnostics \| IgG | 0.9%, 1.0% (FINDDx / MUHC independent evaluation) | - |
| Majdoubi (BC Children's Hospital )[54] | 2020-05-17 to 2020-06-19 | British Columbia \| Vancouver | Local | High | 276 | Mean 42.4 (SD 11.9) | 67.4 | Non-probability | Multiple populations | 0.6% (0.0-2.7%) | Ortho-Clinical Diagnostics; Meso Scale Diagnostics \| VITROS 5600 analyser, 10-plex multiplex assay \| IgG, IgA, IgM | 0.0%, 0.0% (Used author-adjusted estimate) | 0.6% (0.0-1.7%) |

| Article Author (Study Organization) | Sampling Dates (YMD) | Location | Geographic Scope | Overall Risk of Bias | Sample Size | Age | Female (%) | Sampling method | Sample frame | Uncorrected Seroprevalence (95% Confidence Interval) | Test Manufacturers \| Names \| Isotypes | Test Sens, Spec (Source) | Corrected Seroprevalence (95% Credible Interval) |
|---|---|---|---|---|---|---|---|---|---|---|---|---|---|
| Vijh (University of British Columbia)[57] | 2020-05-04 to 2020-05-14 | British Columbia \| Vancouver | Sublocal | High | 291 | Median 86 (residents); 49 (staff) (IQR 15 (residents); 18 (staff)) | 72.2 | Non-probability | Assisted living and long-term care facilities | 39.0% | Abbott; DiaSorin; Ortho Vitros; Siemens \| DiaSorin Liaison IgG, Ortho Vitros Total Antibody, Ortho Vitros IgG, Abbott Architect IgG, Siemens Centaur Total Antibody \| IgG | No data | - |
| **Chile** | | | | | | | | | | | | | |
| Torres (Universidade de Chile)[58] | 2020-05-04 to 2020-05-19 | Santiago \| Vitacura | Sublocal | High | 1009 | Mean 10.8 (SD 4.1) | 46.0 | Probability | Students and Daycares | 9.9% (8.6-11.5%) | Genrui Biotech \| Genrui Biotech IgG/IgM Test Kit (Colloidal Gold) \| IgG, IgM | 91.0%, 95.4% (Test developer / manufacturer evaluation) | 5.2% (1.3-9.0%) |
| Torres (Universidade de Chile)[58] | 2020-05-04 to 2020-05-19 | Santiago \| Vitacura | Sublocal | High | 235 | Mean 42.8 (SD 10.4) | 73.2 | Probability | Essential non-healthcare workers | 16.6% (12.1-21.9%) | Genrui Biotech \| Genrui Biotech IgG/IgM Test Kit (Colloidal Gold) \| IgG, IgM | 91.0%, 95.4% (Test developer / manufacturer evaluation) | 13.3% (7.1-20.0%) |
| **Denmark** | | | | | | | | | | | | | |
| Laursen (Copenhagen University Hospital)[59] | 2020-06-22 to 2020-08-10 | | National | High | 2024 | <40 (n=542); 40-60 (n=1045); 60+ (n=437) | 24.5 | Non-probability | Health care workers and caregivers | 2.8% | Zhuhai; China; Guangzhou Wondfo Biotech; Livson Diagnostics; Gunangdong \| Livzon lateral flow test, WONDFO lateral flow \| IgG, IgM | 66.0%, 96.6% (Used Bastos SR/MA data; no sens, spec, or author adjustment available) | 0.3% (0.0-1.1%) |
| Egerup (Copenhagen University Hospital Hvidovre)[60] | 2020-04-04 to 2020-07-03 | Capital Region of Denmark \| Copenhagen | Sublocal | High | 1206 | Median 31.5 (IQR 28.6-34.7 (antibody positive); | 47.5 | Non-probability | Perinatal | 1.4% | Shenzhen Yhlo Biotech \| iFlash 1800 and SARS-CoV-2 | 94.0%, 99.3% (Test developer / manufacturer evaluation) | 0.6% (0.0-1.6%) |

| Article Author (Study Organization) | Sampling Dates (YMD) | Location | Geographic Scope | Overall Risk of Bias | Sample Size | Age | Female (%) | Sampling method | Sample frame | Uncorrected Seroprevalence (95% Confidence Interval) | Test Manufacturers \| Names \| Isotypes | Test Sens, Spec (Source) | Corrected Seroprevalence (95% Credible Interval) |
|---|---|---|---|---|---|---|---|---|---|---|---|---|---|
| | | | | | | 28.2-33.4 (antibody negative)) | | | | | IgM and IgG kits \| IgG, IgM | | |
| Egerup (Copenhagen University Hospital Hvidovre)[60] | 2020-04-04 to 2020-07-03 | Capital Region of Denmark \| Copenhagen | Sublocal | Moderate | 1188 | Median 33.4 - without antibodies 32.4 - with antibodies (IQR 30.2-37.2 - without antibodies 30.6-35.8 - with antibodies) | NR | Non-probability | Hospital visitors | 2.7% | Shenzhen Yhlo Biotech \| iFlash 1800 and SARS-CoV-2 IgM and IgG kits \| IgG, IgM | 94.0%, 99.3% (Test developer / manufacturer evaluation) | 1.5% (0.0-3.0%) |
| Egerup (Copenhagen University Hospital Hvidovre)[60] | 2020-04-04 to 2020-07-03 | Capital Region of Denmark \| Copenhagen | Sublocal | Moderate | 1313 | Median 31.5 (IQR 28.2–33.4 - Women with antibodies 28.6 - 34.7 - Women without antibodies) | 100.0 | Non-probability | Pregnant or parturient women | 2.1% | Shenzhen Yhlo Biotech \| iFlash 1800 and SARS-CoV-2 IgM and IgG kits \| IgG, IgM | 94.0%, 99.3% (Test developer / manufacturer evaluation) | 1.0% (0.0-2.3%) |
| Jespersen (Aarhus University Hospital)[61] | 2020-05-18 to 2020-06-19 | Central Denmark Region | Regional | Moderate | 17948 | <=29 (n=2032); 30-39 (n=3938); 40-49 (n=4735); 50-59 (n=4666); >=60 (2577) | 85.9 | Non-probability | Health care workers and caregivers | 3.4% (2.4-3.8%) | Wantai Biological Pharmacy Enterprise Co; Beijing; China; Ltd \| SARS-CoV-2 total antibody enzyme-linked immunosorbent assay \| IgG, IgA, IgM | 96.7%, 99.5% (Used author-adjusted estimate) | 3.4% (3.1-3.6%) |
| Storgaard (Aarhus Department of Public Health )[63] | 2020-06-01 to 2020-06-04 | Central Denmark Region \| Aarhus | Local | High | 129 | Median 53 (Range 42-61) | 24.0 | Non-probability | Persons experiencing homelessness | 4.7% (2.1-9.8%) | NR \| NR \| IgG, IgM | 98.9%, 97.6% (Test developer / manufacturer evaluation) | 2.8% (0.0-7.3%) |
| Freiesleben (Copenhagen University Hospital)[64] | 2020-04-14 to 2020-05-21 | Copenhagen \| Copenhagen | Sublocal | High | 36 | Mean 32.96 (SD 5.22) | 100.0 | Non-probability | Pregnant or parturient women | 0.0% | YHLO Biotechnology \| YHLO's SARS-CoV-2 IgM/IgG kits \| IgG, IgM | 97.8%, 97.8% (Used Bastos SR/MA data; no sens, spec, or author | 1.9% (0.0-8.0%) |

| Article Author (Study Organization) | Sampling Dates (YMD) | Location | Geographic Scope | Overall Risk of Bias | Sample Size | Age | Female (%) | Sampling method | Sample frame | Uncorrected Seroprevalence (95% Confidence Interval) | Test Manufacturers \| Names \| Isotypes | Test Sens, Spec (Source) | Corrected Seroprevalence (95% Credible Interval) |
|---|---|---|---|---|---|---|---|---|---|---|---|---|---|
| | | | | | | | | | | | | adjustment available) | |
| Iversen (University of Copenhagen)[66] | 2020-04-15 to 2020-04-22 | Capital Region of Denmark | Regional | Low | 28792 | Mean 44.4 (SD 12.6) | 78.9 | Probability | Health care workers and caregivers | 4.4% (4.1-4.7%) | Livzon Diagnostics \| NR \| IgG, IgM | 73.4%, 99.9% (Used author-adjusted estimate) | 4.4% (4.1-4.6%) |
| Freiesleben (Copenhagen University Hospital)[64] | 2020-02-17 to 2020-04-17 | Copenhagen \| Copenhagen | Sublocal | High | 1019 | Mean 31.71 (SD 4.52) | 100.0 | Non-probability | Pregnant or parturient women | 1.8% | YHLO Biotechnology \| YHLO's iFlash 1800 and SARS-CoV-2 IgM/IgG kits \| IgG, IgM | 97.8%, 97.8% (Used Bastos SR/MA data; no sens, spec, or author adjustment available) | 0.3% (0.0-0.9%) |
| **Finland** | | | | | | | | | | | | | |
| Kantele (University of Helsinki)[68] | 2020-04-22 to 2020-05-15 | Uusimaa \| Helsinki | Sublocal | High | 1095 | Median 38 (IQR 31-48) | 81.7 | Non-probability | Health care workers and caregivers | 6.7% | Euroimmun \| EUROLabwork station SARS-CoV-2 IgG ELISA \| IgG | 0.6%, 0.9% (FINDDx / MUHC independent evaluation) | - |
| **France** | | | | | | | | | | | | | |
| Anna ( Institut Curie)[69] | 2020-04-28 to 2020-07-31 | Île-de-France \| Paris | Local | Moderate | 1847 | Mean 38 (Range 19-75) | 77.4 | Non-probability | Health care workers and caregivers | 11.6% (10.2-13.2%) | Institut Curie - author designed \| N/A - author designed \| IgG | No data | - |
| Dimeglio (Centre Hospitalier Universitaire (CHU) de Toulouse)[70] | 2020-06-10 to 2020-07-10 | Toulouse | Sublocal | High | 8758 | Median 40 (IQR 32-50) | 80.4 | Non-probability | Health care workers and caregivers | 3.2% (2.8-3.5%) | Beijing Wantai Biological Pharmacy Enterprise Co. \| Wantai ELISA kit \| NR | 1.0%, 1.0% (FINDDx / MUHC independent evaluation) | - |
| Roederer (Médecins sans Frontières (Epicentre) )[71] | 2020-06-23 to 2020-07-02 | Ile de France \| Paris | Local | High | 818 | Mean 39 (NR) | 20.4 | Non-probability | Persons experiencing homelessness | 52.0% | N/A - Author Designed \| Luciferase-Linked Immunosorbent Assay N, Luciferase-Linked Immunosorbent Assay S, Pseudo | No data | - |

| Article Author (Study Organization) | Sampling Dates (YMD) | Location | Geographic Scope | Overall Risk of Bias | Sample Size | Age | Female (%) | Sampling method | Sample frame | Uncorrected Seroprevalence (95% Confidence Interval) | Test Manufacturers \| Names \| Isotypes | Test Sens, Spec (Source) | Corrected Seroprevalence (95% Credible Interval) |
|---|---|---|---|---|---|---|---|---|---|---|---|---|---|
| | | | | | | | | | | | Neutralization Test \| IgG | | |
| Pere (Hopital Europeen Georges Pompidou)[72] | 2020-05-02 to 2020-06-26 | Île-de-France \| Paris | Sublocal | High | 3569 | Median 39.6 (NR) | 74.8 | Non-probability | Health care workers and caregivers | 11.9% | Abbott \| Abbott Architect SARS-CoV-2 IgG \| IgG | 0.4%, 1.0% (FINDDx / MUHC independent evaluation) | - |
| Tsatsarisa (Université Paris-Saclay)[73] | 2020-04-29 to 2020-06-26 | Île-de-France \| Paris | Sublocal | High | 529 | Mean 33.7 (SD 4.7) | 100.0 | Non-probability | Pregnant or parturient women | 4.7% (3.0-6.7%) | Abbott \| SARS-CoV-2 IgG Architect assay \| IgG | 0.4%, 1.0% (FINDDx / MUHC independent evaluation) | - |
| Gallais (Strasbourg University Hospital)[74] | 2020-05-07 to 2020-06-26 | Grand-Est \| Strasbourg | Local | High | 11 | Mean 50.6 (SD 7.8) | 45.5 | Non-probability | Contacts of COVID patients | 0.0% | Abbott Laboratories \| Abbott Architect SARS-CoV-2 IgG assay \| IgG | 0.4%, 1.0% (FINDDx / MUHC independent evaluation) | - |
| Mesnil (Rothschild Foundation Hospital)[76] | 2020-06-08 to 2020-06-22 | Paris \| Paris | Sublocal | High | 646 | Mean 39 (Range 28-50) | 73.1 | Non-probability | Health care workers and caregivers | 12.0% | Roche \| Elecsys Anti-SARS-CoV-2 Roche COBAS 6000 \| IgG | 99.5%, 99.8% (Test developer / manufacturer evaluation) | 10.7% (6.2-14.6%) |
| Delmas (Cochin APHP)[77] | 2020-05-14 to 2020-06-17 | Île-de-France \| Paris | Sublocal | High | 4607 | Mean 41.8 (SD 12.6) | 75.1 | Non-probability | Health care workers and caregivers | 11.5% (10.6-12.4%) | Abbott Laboratories \| Abbott-Architect test \| IgG | 0.4%, 1.0% (FINDDx / MUHC independent evaluation) | - |
| Mattern (Paris Saclay University)[79] | 2020-05-04 to 2020-05-31 | Paris \| Paris | Sublocal | High | 249 | Median 33 (IQR 29-36) | 100.0 | Non-probability | Pregnant or parturient women | 8.0% | Ortho-Clinical Diagnostics; Abbott \| Abbott SARS-CoV-2 IgG assay \| IgG | 0.4%, 1.0% (FINDDx / MUHC independent evaluation) | - |
| Bal (Université Claude Bernard Lyon)[80] | 2020-04-10 to 2020-05-28 | Auvergne-Rhône-Alpes \| Lyon | Sublocal | High | 190 | Median 36 (NR) | NR | Non-probability | Health care workers and caregivers | 3.7% | Beijing; China bioMérieux; France; Lyon; Wantai \| Wantai SARS-CoV-2 Ab ELISA kit VIDAS SARS-CoV-2 IgG test \| IgG | No data | - |

| Article Author (Study Organization) | Sampling Dates (YMD) | Location | Geographic Scope | Overall Risk of Bias | Sample Size | Age | Female (%) | Sampling method | Sample frame | Uncorrected Seroprevalence (95% Confidence Interval) | Test Manufacturers \| Names \| Isotypes | Test Sens, Spec (Source) | Corrected Seroprevalence (95% Credible Interval) |
|---|---|---|---|---|---|---|---|---|---|---|---|---|---|
| Fumery (Picardie University)[83] | 2020-03-25 to 2020-05-11 | Somme \| Amiens | Sublocal | High | 146 | Median 38 (Range 17-81) | 49.3 | Non-probability | Patients seeking care for non-COVID-19 reasons | 4.8% | DiaSorin \| Liaison® SARS- CoV-2 S1/S2 IgG \| IgG | 97.4%, 98.5% (Test developer / manufacturer evaluation) | 4.0% (0.3-8.3%) |
| Solodky (Centre Leon Berard)[85] | 2020-03-01 to 2020-04-16 | Auvergne-Rhône-Alpes \| Lyon | Sublocal | High | 85 | NR | NR | Non-probability | Health care workers and caregivers | 5.9% | TODA Pharma \| TODA Coronadiag \| IgG, IgM | 100.0%, 100.0% (Test developer / manufacturer evaluation) | 4.6% (0.1-10.0%) |
| Solodky (Centre Leon Berard)[85] | 2020-03-01 to 2020-04-16 | Auvergne-Rhône-Alpes \| Lyon | Sublocal | High | 244 | NR | NR | Non-probability | Patients seeking care for non-COVID-19 reasons | 5.3% | TODA Pharma \| TODA Coronadiag \| IgG, IgM | 100.0%, 100.0% (Test developer / manufacturer evaluation) | 4.8% (1.0-8.4%) |
| Germain (CHU Lille)[88] | 2019-11-01 to 2020-03-16 | Lille | Local | High | 235 | Median 68yo (IQR 57-79) | 51.9 | Non-probability | Tissue donor | 0.9% | Wantai \| SARS-CoV-21 total Ab ELISA test \| NR | 1.0%, 1.0% (FINDDx / MUHC independent evaluation) | - |
| **Germany** | | | | | | | | | | | | | |
| Armann (Technische Universität Dresden)[91] | 2020-09-15 to 2020-10-13 | Saxony | Regional | Moderate | 1779 | Median 15 (students); 50 (teachers) (IQR 14-16 (students); 36-57 (teachers)) | 55.8 | Non-probability | Multiple populations | 0.7% | 1) DiaSorin LIAISON; 2) Abbott; 3) EUROIMMUN \| 1) Liaison® SARS- CoV-2 S1/S2 IgG, 2)Abbott Architect SARS-CoV-2 IgG assay, 3)Euroimmun Anti-SARS-CoV-2 ELISA IgG \| IgG | No data | - |
| Thielecke (Berlin Institute of Health)[92] | 2020-09-28 to 2020-10-02 | City State of Berlin \| Berlin | Local | Moderate | 672 | Median Children (4.4), Educators (44), Household Members (36) (Range Children (1- | NR | Probability | Multiple populations | 0.1% | Euroimmun AG; Germany \| EUROLabWorkstation anti-SARS-CoV-2 IgG \| IgG | 0.6%, 0.9% (FINDDx / MUHC independent evaluation) | - |

| Article Author (Study Organization) | Sampling Dates (YMD) | Location | Geographic Scope | Overall Risk of Bias | Sample Size | Age | Female (%) | Sampling method | Sample frame | Uncorrected Seroprevalence (95% Confidence Interval) | Test Manufacturers \| Names \| Isotypes | Test Sens, Spec (Source) | Corrected Seroprevalence (95% Credible Interval) |
|---|---|---|---|---|---|---|---|---|---|---|---|---|---|
| | | | | | | 6.3), Educators (18-78), Household Members (0-90)) | | | | | | | |
| Rauber (University Hospital Heidelberg)[94] | 2020-05-05 to 2020-08-06 | Baden-Württemberg \| Heidelberg | Sublocal | High | 219 | Median 56.9 (Range 18.1-78.2) | 40.6 | Non-probability | Patients seeking care for non-COVID-19 reasons | 3.2% | EUROIMMUN ™ \| Euroimmun Anti-SARS-CoV-2 ELISA IgG \| IgG | 0.6%, 0.9% (FINDDx / MUHC independent evaluation) | - |
| Hoffmann (Brandenburg University of Technology Cottbus-Senftenberg)[95] | 2020-07-01 to 2020-07-31 | Brandenburg \| Oberspreewald-Lausitz | Sublocal | High | 156 | Mean 42.7 (very-high-risk group); 46.77 (high-risk group); 44.7 (medium risk group); 45.6 (low-risk group) (SD 11.6 (very-high-risk group); 11.91 (high-risk group); 10.9 (medium risk group); 8.5 (low-risk group)) | 86.5 | Non-probability | Health care workers and caregivers | 1.3% | Author-designed \| NR \| NR | 99.5%, 99.8% (Test developer / manufacturer evaluation) | 1.5% (0.0-3.8%) |
| Finkenzeller (Munich Clinic)[96] | 2020-06-29 to 2020-07-29 | Bavaria \| Tirschenreuth, Wieden | Sublocal | Moderate | 1838 | 18-29 (n=322); 30-39 (n=321); 40-49 (n=396); 50-59 (N=536); 60-69 (n=176); >69 (n=2); | 78.5 | Non-probability | Health care workers and caregivers | 15.1% | Roche Diagnostics \| Elecsys® Anti-SARS-CoV-2 immunassay \| IgG, IgM | 0.8%, 1.0% (FINDDx / MUHC independent evaluation) | - |
| Finkenzeller (Munich Clinic)[96] | 2020-06-29 to 2020-07-29 | Bavaria \| Tirschenreuth, Wieden | Sublocal | High | 986 | 18-29 (n=329); 30-39 (n=227); 40-49 (n=151); 50-59 (N=163); 60-69 | 22.8 | Non-probability | Essential non-healthcare workers | 3.7% | Roche DIagnostics \| Elecsys® Anti-SARS-CoV-2 immunassay \| IgG, IgM | 0.8%, 1.0% (FINDDx / MUHC independent evaluation) | - |

| Article Author (Study Organization) | Sampling Dates (YMD) | Location | Geographic Scope | Overall Risk of Bias | Sample Size | Age | Female (%) | Sampling method | Sample frame | Uncorrected Seroprevalence (95% Confidence Interval) | Test Manufacturers \| Names \| Isotypes | Test Sens, Spec (Source) | Corrected Seroprevalence (95% Credible Interval) |
|---|---|---|---|---|---|---|---|---|---|---|---|---|---|
| | | | | | | (n=34); >69 (n=1); | | | | | | | |
| Brehm (University Medical Center Hamburg-Eppendorf)[98] | 2020-06-22 to 2020-07-17 | Hamburg \| Hamburg | Sublocal | Moderate | 1253 | Median 36 (IQR 29-48) | 74.5 | Non-probability | Health care workers and caregivers | 1.8% (1.0-2.5%) | Euroimmun \| Euroimmun SARS-CoV-2 IgG ELISA \| IgG | 0.6%, 0.9% (FINDDx / MUHC independent evaluation) | - |
| Mack (Saarland University)[99] | 2020-06-16 to 2020-06-30 | | National | High | 1007 | Median 31 (IQR 25-37) | 2.5 | Non-probability | Non-essential workers and unemployed persons | 2.1% (1.4-3.2%) | Roche; EUROimmun; NR \| EUROImmun anti-SARS-CoV-2 ELISA (IgG), Cobas Elecsys Anti-SARS-CoV-2 chemiluminescent immunoassay (CLIA), Neutralizing assay \| IgG | No data | - |
| Epstude (Thuringia Clinic)[100] | 2020-06-15 to 2020-06-30 | Thuringia \| Saalfeld | Sublocal | High | 45 | Mean 47.8 (SD 10.4) | 86.7 | Non-probability | Health care workers and caregivers | 0.0% | PerkinElmer Inc \| Euroimmun Anti-SARS-CoV-2 ELISA IgG \| IgG | 0.4%, 1.0% (FINDDx / MUHC independent evaluation) | - |
| Epstude (Thuringia Clinic)[100] | 2020-06-15 to 2020-06-30 | Thuringia \| Saalfeld | Sublocal | High | 20 | Mean 39.4 (SD 10.2) | 70.0 | Non-probability | Health care workers and caregivers | 5.0% | PerkinElmer Inc \| Euroimmun Anti-SARS-CoV-2 ELISA IgG \| IgG | 0.4%, 1.0% (FINDDx / MUHC independent evaluation) | - |
| Bahrs (Jena University Hospital)[102] | 2020-05-19 to 2020-06-19 | Thuringia \| Jena | Sublocal | High | 660 | Median 40.5 (IQR 32-49) | 73.6 | Non-probability | Health care workers and caregivers | 2.7% | Roche; Epitope Diagnostics \| EDI Novel Coronavirus SARS-CoV-2 IgG ELISA kit, Elecsys Anti-SARS-CoV-2 kit \| IgG | No data | - |

| Article Author (Study Organization) | Sampling Dates (YMD) | Location | Geographic Scope | Overall Risk of Bias | Sample Size | Age | Female (%) | Sampling method | Sample frame | Uncorrected Seroprevalence (95% Confidence Interval) | Test Manufacturers \| Names \| Isotypes | Test Sens, Spec (Source) | Corrected Seroprevalence (95% Credible Interval) |
|---|---|---|---|---|---|---|---|---|---|---|---|---|---|
| Herzberg (University Hospitals of the Ruhr-University of Bochum)[103] | 2020-04-14 to 2020-06-16 | Schleswig-Holstein \| Hamburg | Sublocal | High | 871 | Mean 40 (SD 14.2) | 75.1 | Non-probability | Health care workers and caregivers | 2.6% | Mikrogen Diagnostik; Euroimmun \| recomWell SARS-CoV2 IgG immunoassay test, anti-SARS-CoV- 2 ELISA (IgG) test \| IgG | 75.6%, 97.6% (Used Bastos SR/MA data; no sens, spec, or author adjustment available) | 0.7% (0.0-2.1%) |
| Malfertheiner (University of Regensburg)[106] | 2020-03-15 to 2020-06-07 | Bavaria \| Regensburg | Sublocal | High | 139 | 18-35 (n = 27) ; 36-50 (n = 43) ; 51-65 (n = 32) | NR | Non-probability | Health care workers and caregivers | 0.0% | EUROIMMUN AG; Roche Diagnostics \| Anti- SARS-CoV-2 IgG and IgA ELISA, Elecsys Anti-SARS-CoV-2 \| IgG, IgA | 75.6%, 97.6% (Used Bastos SR/MA data; no sens, spec, or author adjustment available) | 0.7% (0.0-2.9%) |
| Thieme (Ruhr-University Bochum)[109] | 2020-04-01 to 2020-05-31 | Bochum | Local | High | 99 | Median 51 (IQR 41-62) | 43.4 | Non-probability | Patients seeking care for non-COVID-19 reasons | 3.0% | Euroimmun \| NR \| IgG, IgA | 0.9%, 0.8% (FINDDx / MUHC independent evaluation) | - |
| Erber (Technical University of Munich)[110] | 2020-04-14 to 2020-05-29 | Bavaria \| Munich | Sublocal | High | 4554 | Mean 38.5 (NR) | 70.4 | Non-probability | Health care workers and caregivers | 2.4% (1.9-2.9%) | Unclear \| Unclear which chemiluminescent immunoassay was used. "In all samples with incongruent results, IgG antibodies against SARS-CoV-2 S1 protein were determined using an enzyme linked immunosorbent assay (ELISA) (Euroimmun, Luebeck, Germany), and immunoblot was used to differentiate antibodies against N, S1 | 97.8%, 97.8% (Used Bastos SR/MA data; no sens, spec, or author adjustment available) | 0.3% (0.0-0.8%) |

| Article Author (Study Organization) | Sampling Dates (YMD) | Location | Geographic Scope | Overall Risk of Bias | Sample Size | Age | Female (%) | Sampling method | Sample frame | Uncorrected Seroprevalence (95% Confidence Interval) | Test Manufacturers \| Names \| Isotypes | Test Sens, Spec (Source) | Corrected Seroprevalence (95% Credible Interval) |
|---|---|---|---|---|---|---|---|---|---|---|---|---|---|
| | | | | | | | | | | | and the receptor binding domain of SARS-CoV-2 from those against seasonal coronaviruses 165 (Mikrogen, Neuried, Germany)" \| IgG | | |
| Choi (Charité University)[112] | 2020-03-19 to 2020-05-19 | Berlin \| Berlin | Sublocal | High | 223 | Median 54 (IQR 42.64) | 38.1 | Non-probability | Patients seeking care for non-COVID-19 reasons | 7.1% | Euroimmun Medizinische Labordiagnostika AG \| Euroimmun Analyzer I \| IgG, IgA | 0.9%, 0.8% (FINDDx / MUHC independent evaluation) | - |
| Neumann (University of Bremen)[114] | 2020-03-31 to 2020-05-16 | Bremen | Local | High | 281 | NR | NR | Non-probability | Multiple populations | 2.1% | Euroimmun \| Euroimmun AG ELISA \| IgG, IgA | 0.7%, 0.7% (FINDDx / MUHC independent evaluation) | - |
| Tonshoff (Heidelberg University)[115] | 2020-04-22 to 2020-05-15 | Baden-Württemberg \| Tübingen, Ulm, Freiburg, Heidelberg | Regional | Moderate | 4964 | Median 40 (IQR 36-43) | 61.1 | Non-probability | Multiple populations | 1.7% (1.3-2.0%) | Euroimmun; Roche Elecsys' Mikrogen in-house \| Euroimmun SARS-CoV-2 IgG; Roche Eleccsys Anti-SARS-CoV-2; Mikrogen recomWell EILSA; in-house Luminex-based assay \| IgG | 75.6%, 97.6% (None) | 0.3% (0.1-0.4%) |
| Zöllkau (Universitätsklinikum Jena)[116] | 2020-04-06 to 2020-05-13 | Thuringia \| Jena | Local | High | 180 | NR | 100.0 | Non-probability | Pregnant or parturient women | 0.6% (0.0-1.7%) | Epitope Diagnostics \| NR \| IgG | 0.8%, 1.0% (FINDDx / MUHC independent evaluation) | - |
| Schmidt (University of Hannover)[117] | 2020-04-20 to 2020-04-30 | Lower Saxony \| Hannover | Sublocal | High | 385 | 18-29 (n=55); 30-49 (n=154); 50-64 | 80.0 | Non-probability | Health care workers and caregivers | 2.9% | EUROIMMUN \| Euroimmun Anti-SARS-CoV-2 ELISA IgG \| IgG | 0.8%, 1.0% (FINDDx / MUHC independent evaluation) | - |

| Article Author (Study Organization) | Sampling Dates (YMD) | Location | Geographic Scope | Overall Risk of Bias | Sample Size | Age | Female (%) | Sampling method | Sample frame | Uncorrected Seroprevalence (95% Confidence Interval) | Test Manufacturers \| Names \| Isotypes | Test Sens, Spec (Source) | Corrected Seroprevalence (95% Credible Interval) |
|---|---|---|---|---|---|---|---|---|---|---|---|---|---|
| | | | | | | (n=170); >=65 (n=6) | | | | | | | |
| Korth (University Hospital Essen)[119] | 2020-03-25 to 2020-04-21 | North Rhine-Westphalia \| Essen | Sublocal | High | 316 | Mean 36.7 (Average age of high-risk group); 42.3 (Average age of low-risk group) (SD 10.7 (SD of high-risk group); 13.2 (SD of low-risk group)) | 64.6 | Non-probability | Health care workers and caregivers | 1.6% | EUROIMMUN \| Eurommun Anti-SARS-CoV-2-IgG \| IgG | 0.8%, 1.0% (FINDDx / MUHC independent evaluation) | - |
| Harsch (Thuringia Clinic)[120] | 2020-04-16 to 2020-04-21 | Thuringia \| Saalfeld | Sublocal | High | 18 | Mean 44.9 (Range 21-60) | NR | Non-probability | Health care workers and caregivers | 0.0% | EUROIMMUN \| EUROIMMUN ™ \| IgG | 0.4%, 1.0% (FINDDx / MUHC independent evaluation) | - |
| Behrens ( Hannover Medical School)[121] | 2020-03-23 to 2020-04-17 | Lower Saxony \| Hannover | Local | High | 217 | Mean 36.5 (Range 18-63) | 65.0 | Non-probability | Health care workers and caregivers | 1.4% | EUROIMMUN \| Euroimmun Anti-SARS-CoV-2 ELISA IgG \| IgG | 0.6%, 0.9% (FINDDx / MUHC independent evaluation) | - |
| Kern (Universitätsmedizin Marburg)[122] | 2020-04-09 to 2020-04-16 | Fulda | Sublocal | High | 1780 | 10-19 (n=52, 2.9%), 20-29 (n=422, 23.7%), 30-39 (n=378, 21.2%), 40-49 (n=347, 19.5%), 50-59 (n=431, 24.2%), 60-69 (n=150, 8.4%) | 79.8 | Non-probability | Multiple populations | 1.0% (0.6-1.5%) | EUROIMMUN \| Euroimmun Anti-SARS-CoV-2 IgG IgA ELISA \| IgG | 0.8%, 1.0% (FINDDx / MUHC independent evaluation) | - |
| Kraehling ( Goethe-University Frankfurt)[123] | 2020-04-06 to 2020-04-14 | Hesse \| Frankfurt | Local | High | 998 | <30 (n=115); 30-40 (n=172); 41-50 (n=213); 51-60 (n=399); >60 (n=101) | 21.5 | Non-probability | Essential non-healthcare workers | 0.4% (0.2-1.0%) | Author-designed \| N/A - Author-designed \| IgG | 75.6%, 97.6% (Used Bastos SR/MA data; no sens, spec, or author adjustment available) | 0.1% (0.0-0.5%) |

| Article Author (Study Organization) | Sampling Dates (YMD) | Location | Geographic Scope | Overall Risk of Bias | Sample Size | Age | Female (%) | Sampling method | Sample frame | Uncorrected Seroprevalence (95% Confidence Interval) | Test Manufacturers \| Names \| Isotypes | Test Sens, Spec (Source) | Corrected Seroprevalence (95% Credible Interval) |
|---|---|---|---|---|---|---|---|---|---|---|---|---|---|
| Lackermair (University Hospital Munich)[124] | 2020-04-02 to 2020-04-06 | Bavaria \| Dachau | Regional | High | 151 | Mean 38 (Range 26-47) | 83.4 | Non-probability | Health care workers and caregivers | 2.6% (0.8-7.1%) | EUROIMMUN \| Euroimmun Anti-SARS-CoV-2 ELISA IgG \| IgG | 0.4%, 1.0% (FINDDx / MUHC independent evaluation) | - |
| **Greece** | | | | | | | | | | | | | |
| Tsitsilonis (National and Kapodistrian University of Athens)[126] | 2020-06-15 to 2020-07-15 | Athens | Sublocal | Moderate | 2500 | 18-34 (n=1268); 35-54 (n=916); 55-74 (n=309); Unknown (n=7) | 64.0 | Non-probability | Non-essential workers and unemployed persons | 0.9% (0.3-2.1%) | Roche Diagnostics \| CE-IVD Roche Cobas Elecsys® Anti-SARS-CoV-2 \| NR | 100.0%, 99.8% (Used author-adjusted estimate) | 0.9% (0.6-1.4%) |
| Psichogiou (University of Athens)[127] | 2020-04-13 to 2020-05-15 | Attica \| Athens | Sublocal | Moderate | 1495 | Mean 46.4 (SD 10.3) | 69.7 | Non-probability | Health care workers and caregivers | 1.0% | Genebody Inc \| GeneBody COVID-19 IgM/IgG detection \| IgG, IgM | 74.1%, 100.0% (Test developer / manufacturer evaluation) | 0.8% (0.0-1.7%) |
| Vlachoyiannopoulosa (National and Kapodistrian University of Athens/Laiko University Hospital)[128] | 2020-04-25 to 2020-05-10 | Attica \| Athens | Sublocal | High | 321 | Mean 42.71 (NR) | 67.9 | Non-probability | Health care workers and caregivers | 2.2% | EUROIMMUN; Germany \| EUROIMMUN IgG ELISA \| IgG | 0.6%, 0.9% (FINDDx / MUHC independent evaluation) | - |
| **Italy** | | | | | | | | | | | | | |
| Caponigro (ASST della Valtellina Alto Lario-Oncologia Medica Sondrio)[134] | 2020-03-01 to 2020-10-19 | Lombardy \| Sondrio | Sublocal | Unclear | 220 | NR | NR | Non-probability | Health care workers and caregivers | 11.4% | Nal Von Minden GmbH (Germany) \| NADAL® COVID-19 IgG/IgM Rapid Test (test cassette) REF 243001N-10; \| IgG, IgM | 94.1%, 99.2% (Test developer / manufacturer evaluation) | 10.0% (4.1-16.1%) |
| Russo (University of Campania)[135] | 2020-03-27 to 2020-08-30 | Campania \| Naples | Sublocal | High | 65 | Median 32 (IQR 28-43) | 61.5 | Non-probability | Health care workers and caregivers | 0.0% | Xiamen \| WANTAI SARS-CoV-2 Ab ELISA \| NR | 1.0%, 1.0% (FINDDx / MUHC independent evaluation) | - |

| Article Author (Study Organization) | Sampling Dates (YMD) | Location | Geographic Scope | Overall Risk of Bias | Sample Size | Age | Female (%) | Sampling method | Sample frame | Uncorrected Seroprevalence (95% Confidence Interval) | Test Manufacturers \| Names \| Isotypes | Test Sens, Spec (Source) | Corrected Seroprevalence (95% Credible Interval) |
|---|---|---|---|---|---|---|---|---|---|---|---|---|---|
| Ralli (Eleemosynaria Apostolica)[136] | 2020-04-14 to 2020-07-31 | Lazio \| Rome | Local | High | 173 | Mean 45.6 (Range 10-80) | 29.5 | Non-probability | Multiple populations | 1.8% | Inzek B.V \| Biozek Medical COVID-19 Rapid Test, \| IgG, IgM | 96.0%, 96.0% (Test developer / manufacturer evaluation) | 0.8% (0.0-3.1%) |
| Norsa (Papa Giovani XXII)[138] | 2020-03-04 to 2020-07-10 | Lombardy \| Bergamo | Sublocal | High | 90 | <28 (n=32); 38-47 (n=27); >47 (n=31) | 46.7 | Non-probability | Patients seeking care for non-COVID-19 reasons | 21.1% | VivaChek \| VivaDiag COVID-19 IgM/IgG Rapid Test \| IgG, IgM | 0.7%, 0.9% (FINDDx / MUHC independent evaluation) | - |
| Medas ( University of Cagliari)[140] | 2020-03-31 to 2020-06-30 | Sardinia \| Cagliari | Sublocal | High | 86 | Mean 57.6 (NR) | 62.8 | Non-probability | Patients seeking care for non-COVID-19 reasons | 5.8% | Snibe \| Maglumi platform \| IgG, IgM | 97.8%, 97.8% (Used Bastos SR/MA data; no sens, spec, or author adjustment available) | 3.4% (0.0-8.3%) |
| Berte (University of Cagliari)[141] | 2020-04-15 to 2020-06-15 | Milan, Cagliari, Erlangen | Local | High | 354 | Median 43 (IQR 31-57) | 37.9 | Non-probability | Patients seeking care for non-COVID-19 reasons | 2.3% (0.8-3.8%) | University of Cagliari \| N/A - Author Designed \| IgG, IgA | 97.6%, 95.2% (Test developer / manufacturer evaluation) | 0.7% (0.0-2.4%) |
| Capasso (University of Naples)[142] | 2020-05-11 to 2020-06-15 | Campania \| Naples | Sublocal | High | 235 | Mean 39.4 (SD 10.9) | 54.5 | Non-probability | Health care workers and caregivers | 10.6% | Ltd.; Shanghai Kehua Bio-engineering Co.; China; Shanghai \| NR \| IgG, IgM | 0.7%, 1.0% (FINDDx / MUHC independent evaluation) | - |
| Capasso (University of Naples)[142] | 2020-05-11 to 2020-06-15 | Campania \| Naples | Sublocal | Moderate | 862 | Mean 42.9 (SD 13.3) | 47.8 | Non-probability | Health care workers and caregivers | 1.3% | Ltd.; Shanghai Kehua Bio-engineering Co.; China; Shanghai \| NR \| IgG, IgM | 0.7%, 1.0% (FINDDx / MUHC independent evaluation) | - |
| Capasso (University of Naples)[142] | 2020-05-11 to 2020-06-15 | Campania \| Naples | Sublocal | Moderate | 310 | Mean 42.3 (SD 12.4) | 67.1 | Non-probability | Patients seeking care for non-COVID-19 reasons | 2.9% | Ltd.; Shanghai Kehua Bio-engineering Co.; China; Shanghai \| NR \| IgG, IgM | 0.7%, 1.0% (FINDDx / MUHC independent evaluation) | - |
| Cosma (University of Turin)[144] | 2020-04-16 to 2020-06-04 | Piedmont \| Turin | Local | High | 138 | NR | 100.0 | Non-probability | Pregnant or parturient women | 5.8% | Boditech \| AFIAS™ COVID-19 \| IgG | 66.0%, 96.6% (Used Bastos SR/MA data; no sens, spec, | 3.7% (0.0-9.7%) |

| Article Author (Study Organization) | Sampling Dates (YMD) | Location | Geographic Scope | Overall Risk of Bias | Sample Size | Age | Female (%) | Sampling method | Sample frame | Uncorrected Seroprevalence (95% Confidence Interval) | Test Manufacturers \| Names \| Isotypes | Test Sens, Spec (Source) | Corrected Seroprevalence (95% Credible Interval) |
|---|---|---|---|---|---|---|---|---|---|---|---|---|---|
| | | | | | | | | | | | | or author adjustment available) | |
| Lorenzo (Santagostino Medical Center)[146] | 2020-05-02 to 2020-05-31 | Lombardy \| Milan | Local | High | 119 | Mean 47 (healthcare workers); 79 (family member old); 42 (family member middle age); 11 (family member youth) (SD 18 (health care workers); 6 (family member old); 12 (family member middle age); 7 (family member young); ) | 54.6 | Non-probability | Family of essential workers | 23.5% | Prima Lab \| IgM/IgG serological rapid tests \| IgG, IgM | 0.8%, 0.9% (FINDDx / MUHC independent evaluation) | - |
| Lastrucci (University of Florence)[147] | 2020-05-01 to 2020-05-31 | Prato | Regional | Moderate | 4656 | Median 49 (IQR 38-57) | 67.1 | Non-probability | Multiple populations | 3.0% (2.5-3.5%) | Hangzhou Laihe Biotech Co.; Ltd; Hangzhou China \| LYHER Novel Coronavirus (2019-nCoV) IgM/IgG Antibody Combo Test Kit(Colloidal Gold) \| IgG, IgM | 100.0%, 98.8% (Test developer / manufacturer evaluation) | 1.5% (0.0-2.8%) |
| Papalini (University of Perugia)[148] | 2020-05-04 to 2020-05-31 | Umbria \| Perugia | Local | Unclear | 2843 | NR | NR | Non-probability | Multiple populations | 5.4% | Screen Italia \| SCREEN test COVID-19 \| IgG | 66.0%, 96.6% (Used Bastos SR/MA data; no sens, spec, or author adjustment available) | 3.2% (1.4-4.8%) |

| Article Author (Study Organization) | Sampling Dates (YMD) | Location | Geographic Scope | Overall Risk of Bias | Sample Size | Age | Female (%) | Sampling method | Sample frame | Uncorrected Seroprevalence (95% Confidence Interval) | Test Manufacturers \| Names \| Isotypes | Test Sens, Spec (Source) | Corrected Seroprevalence (95% Credible Interval) |
|---|---|---|---|---|---|---|---|---|---|---|---|---|---|
| Papalini (University of Perugia)[148] | 2020-05-04 to 2020-05-31 | Umbria \| Perugia | Sublocal | High | 270 | Median 52 (Range 10-83) | NR | Non-probability | Patients seeking care for non-COVID-19 reasons | 3.7% | Screen Italia \| SCREEN test COVID-19 \| IgG | 66.0%, 96.6% (Used Bastos SR/MA data; no sens, spec, or author adjustment available) | 1.4% (0.0-4.5%) |
| Sperotto (The University of Udine)[150] | 2020-04-01 to 2020-05-30 | Udine \| Udine | Sublocal | High | 70 | Median 56 (Range 23-73) | 51.4 | Non-probability | Patients seeking care for non-COVID-19 reasons | 1.4% | Cellex \| Cellex qSARS-CoV-2 immunoglobilin G (IgG)/immunoglobulin M (IgM) point-of-care antibodies diagnostic rapid test \| IgG, IgM | 0.8%, 0.9% (FINDDx / MUHC independent evaluation) | - |
| Stellato (Azienda Ospedaliera di Rilievo Nazionale Santobono Pausilipon)[151] | 2020-04-03 to 2020-05-29 | Campania \| Naples | Sublocal | High | 249 | NR | NR | Non-probability | Health care workers and caregivers | 2.0% | VivaDiag; N/A \| NR \| IgG, IgM | 66.0%, 96.6% (Used Bastos SR/MA data; no sens, spec, or author adjustment available) | 0.8% (0.0-3.2%) |
| Stellato (Azienda Ospedaliera di Rilievo Nazionale Santobono Pausilipon)[151] | 2020-04-03 to 2020-05-29 | Campania \| Naples | Sublocal | High | 1397 | NR | NR | Non-probability | Multiple populations | 0.4% | VivaDiag; N/A \| NR \| IgG, IgM | 66.0%, 96.6% (Used Bastos SR/MA data; no sens, spec, or author adjustment available) | 0.1% (0.0-0.4%) |
| Plebani (University Hospital Of Padova)[152] | 2020-02-22 to 2020-05-29 | Veneto | Regional | High | 8285 | Mean 43.2 (SD 11.6) | 71.6 | Non-probability | Health care workers and caregivers | 4.6% (4.1-5.0%) | SNIBE – Shenzhen New Industries Biomedical Engineering Co. \| MAGLUMI 2019-nCov IgG/IgM CLIA \| IgG, IgM | 73.0%, 98.0% (Author-reported independent evaluation) | 3.7% (2.6-4.6%) |
| Poletti (Bruno Kessler Foundation)[153] | 2020-04-16 to 2020-05-25 | Lombardy | Regional | Moderate | 4120 | Median 50 (IQR 30-61) | 75.0 | Non-probability | Contacts of COVID patients | 53.1% | LIAISON® \| LIAISON® SARS-CoV-2 test \| IgG | 0.6%, 1.0% (FINDDx / MUHC independent evaluation) | 46.8% (44.6-48.6%) |

| Article Author (Study Organization) | Sampling Dates (YMD) | Location | Geographic Scope | Overall Risk of Bias | Sample Size | Age | Female (%) | Sampling method | Sample frame | Uncorrected Seroprevalence (95% Confidence Interval) | Test Manufacturers \| Names \| Isotypes | Test Sens, Spec (Source) | Corrected Seroprevalence (95% Credible Interval) |
|---|---|---|---|---|---|---|---|---|---|---|---|---|---|
| Calcagno (University of Torino)[155] | 2020-04-17 to 2020-05-20 | Piemonte \| Torino (Turin) | Local | Moderate | 5444 | Mean 49.4 (SD 10.6) | 74.7 | Non-probability | Health care workers and caregivers | 6.9% | DiaSorin \| LIAISON® SARS-CoV-2 S1/S2 IgG \| IgG | 0.8%, 1.0% (FINDDx / MUHC independent evaluation) | - |
| Carlo (Azienda Ospedaliera-Universitaria Policlinico Riuniti di Foggia)[156] | 2020-03-17 to 2020-05-18 | Province of Foggia, Apulia \| Foggia | Sublocal | Moderate | 3242 | Mean 46.5 (SD 11.7) | NR | Non-probability | Health care workers and caregivers | 1.9% (1.4-2.3%) | Shenzhen YHLO Biotech \| iFlash1800 SARS-COV-2 IgG & IgM \| IgG, IgM | 97.0%, 98.8% (Test developer / manufacturer evaluation) | 0.9% (0.0-1.9%) |
| Martella (Azienda Usl Toscana Centro Oncology)[157] | 2020-05-11 to 2020-05-17 | Tuscany \| Florence | Sublocal | Unclear | 1145 | NR | NR | Non-probability | Patients seeking care for non-COVID-19 reasons | 2.3% | NR \| Quantitative serologic IgM and IgG test (Qt-ST) \| IgG, IgM | No data | - |
| Cavaliere (Santo Stefano Hospital)[158] | 2020-04-04 to 2020-05-16 | Tuscany \| Prato | Local | High | 134 | NR | 100.0 | Non-probability | Pregnant or parturient women | 4.5% | Acro Biotech Inc. \| COVID-19 IgG/IgM Rapid Test Cassette \| IgG, IgM | 0.4%, 0.9% (FINDDx / MUHC independent evaluation) | - |
| Sandri (IRCCS)[159] | 2020-04-28 to 2020-05-16 | Lombardy \| Milan, Castellanza, Varese | Regional | Moderate | 3985 | Median 42 (Range 21-86) | 66.8 | Non-probability | Health care workers and caregivers | 13.4% | DiaSorin \| Liaison® SARS- CoV-2 S1/S2 IgG \| IgG | 0.8%, 1.0% (FINDDx / MUHC independent evaluation) | - |
| Bianchi (Università degli Studi di Milano)[160] | 2020-04-15 to 2020-05-15 | Lombardy \| Milan | Sublocal | Unclear | 42 | NR | NR | Non-probability | Patients seeking care for non-COVID-19 reasons | 9.5% | Diasorin; Abbott Laboratories \| NR \| IgG | 97.8%, 97.8% (Used Bastos SR/MA data; no sens, spec, or author adjustment available) | 6.6% (0.0-15.1%) |
| Bianchi (Università degli Studi di Milano)[160] | 2020-04-15 to 2020-05-15 | Lombardy \| Milan | Sublocal | Unclear | 34 | NR | NR | Non-probability | Health care workers and caregivers | 5.9% | Diasorin; Abbott Laboratories \| NR \| IgG | 97.8%, 97.8% (Used Bastos SR/MA data; no sens, spec, or author adjustment available) | 3.6% (0.0-11.9%) |

| Article Author (Study Organization) | Sampling Dates (YMD) | Location | Geographic Scope | Overall Risk of Bias | Sample Size | Age | Female (%) | Sampling method | Sample frame | Uncorrected Seroprevalence (95% Confidence Interval) | Test Manufacturers \| Names \| Isotypes | Test Sens, Spec (Source) | Corrected Seroprevalence (95% Credible Interval) |
|---|---|---|---|---|---|---|---|---|---|---|---|---|---|
| Marra (Istituto di Ricovero e Cura a Carattere Scientifico (IRCCS))[162] | 2020-03-30 to 2020-05-11 | Lombardy | Regional | High | 166 | Median 46 (IQR 21) | 71.1 | Non-probability | Multiple populations | 41.6% | PRIMA Lab SA \| 2019-nCoV IgG/ IgM Rapid Test Cassette \| IgG | 0.7%, 0.9% (FINDDx / MUHC independent evaluation) | 59.7% (50.6-69.2%) |
| Baracco (A.S.S.T. Lodi )[164] | 2020-04-23 to 2020-05-05 | Lombardy Region | Sublocal | High | 2415 | Mean 48 (SD 10.0) | 72.6 | Non-probability | Health care workers and caregivers | 16.8% | DiaSorin \| The Liaison® DiaSorin® SARS-CoV-2 S1/S2 IgG test \| IgG | 0.8%, 1.0% (FINDDx / MUHC independent evaluation) | - |
| Denina (University of Turin)[165] | 2020-03-08 to 2020-04-30 | Piedmont | Regional | High | 24 | Mean 13 (Range 6-17) | 66.7 | Non-probability | Patients seeking care for non-COVID-19 reasons | 12.5% | DiaSorin \| Liaison® SARS- CoV-2 S1/S2 IgG \| IgG | 0.8%, 1.0% (FINDDx / MUHC independent evaluation) | - |
| Zambelli (Ospedale Papa Giovanni XXIII)[166] | 2020-04-01 to 2020-04-30 | Lombardy \| Bergamo | Sublocal | High | 560 | Median 65 (Range 19-89) | 60.0 | Non-probability | Patients seeking care for non-COVID-19 reasons | 31.0% | Moers; Germany \| NADAL COVID-19 IgG/IgM Test \| IgG, IgM | 92.1%, 99.2% (Author-reported independent evaluation) | 32.6% (27.9-36.9%) |
| Carozzi (Arezzo Hospital)[168] | 2020-04-01 to 2020-04-30 | Tuscany | Regional | High | 17098 | NR | NR | Non-probability | Health care workers and caregivers | 3.1% | "Zhejiang Orient Gene Biotech Co.; LTD"; Screen Italia S.R.L \| Screen Test COVID-19 2019-nCOV IgG/IgM, COVID-19 IgG / IgM Rapid Test Cassette \| IgG, IgM | 66.0%, 96.6% (Used Bastos SR/MA data; no sens, spec, or author adjustment available) | 0.2% (0.0-0.8%) |
| Bontadi (National Association of Company and Competent Doctors)[169] | 2020-04-11 to 2020-04-29 | Veneto | Local | High | 1267 | Mean 44 (Range 19-69) | 17.1 | Non-probability | Essential non-healthcare workers | 1.6% | NR; MAGLUMI; NR \| Rapid Test (NR), MAGLUMI CLIA, ELISA (NR) \| IgG, IgM | No data | - |

| Article Author (Study Organization) | Sampling Dates (YMD) | Location | Geographic Scope | Overall Risk of Bias | Sample Size | Age | Female (%) | Sampling method | Sample frame | Uncorrected Seroprevalence (95% Confidence Interval) | Test Manufacturers \| Names \| Isotypes | Test Sens, Spec (Source) | Corrected Seroprevalence (95% Credible Interval) |
|---|---|---|---|---|---|---|---|---|---|---|---|---|---|
| Volta (University of Brescia)[171] | 2020-04-27 to 2020-04-27 | Lombardy \| Brescia | Sublocal | High | 76 | Median 46 (IQR 23-69) | 75.0 | Non-probability | Health care workers and caregivers | 11.8% | DiaSorin \| NR \| IgG | 0.8%, 1.0% (FINDDx / MUHC independent evaluation) | - |
| Lahner (University of Rome)[172] | 2020-04-07 to 2020-04-27 | Lazio \| Rome | Sublocal | High | 1084 | Median 46 (Range 19-69) | 60.9 | Non-probability | Health care workers and caregivers | 0.7% | Medical Systems \| 2019-nCoV IgM/IgG CLIA \| IgG | 50.0%, 99.1% (Author-reported independent evaluation) | 0.3% (0.0-1.3%) |
| Landro (Hospital of Bergamo)[170] | Until 2020-04-24 | Lombardy \| Bergamo | Sublocal | Unclear | 3000 | NR | NR | Non-probability | Health care workers and caregivers | 13.0% | NR \| NR \| NR | No data | - |
| Paradiso (Istituto Tumori G. Paolo II)[175] | 2020-03-26 to 2020-04-17 | Puglia \| Bari | Sublocal | High | 606 | Median 47.5 (Range 20-73) | 60.6 | Non-probability | Health care workers and caregivers | 0.3% | Snibe Diagnostic \| MAGLUMI800 TM \| IgG | 0.7%, 1.0% (FINDDx / MUHC independent evaluation) | - |
| Sotgiu (Università degli Studi di Milano )[176] | 2020-04-02 to 2020-04-16 | Lombardy \| Milan | Local | High | 202 | 20-29 (n=27); 30-39 (n=44); 40-49 (n=57); 50-59 (n=51); 60-69 (n=23) | 65.3 | Non-probability | Health care workers and caregivers | 7.4% (3.8-11.0%) | BioMedomics \| BioMedomics IgM-IgG Combined Antibody Rapid Test \| IgG | 0.6%, 1.0% (FINDDx / MUHC independent evaluation) | - |
| Amendola (University of Milan)[177] | 2020-04-15 to 2020-04-15 | Lombardy \| Milan | Sublocal | High | 663 | Median 44 (NR) | 83.7 | Non-probability | Health care workers and caregivers | 5.1% | EUROIMMUN \| Euroimmun Anti-SARS-CoV-2 ELISA IgG \| IgG | 0.8%, 1.0% (FINDDx / MUHC independent evaluation) | - |
| Canetti (San Raffaele Scientific Institute)[178] | 2020-03-15 to 2020-04-15 | Lombardy \| Milan | Sublocal | High | 40 | Median 51 (IQR 34-58) | 77.5 | Non-probability | Health care workers and caregivers | 5.0% | Diasorin; Prima lab SA \| COVID-19 IgG/IgM Rapid Test , DiaSorin LIAISON® SARSCoV- 2 S1/S2 IgG test \| IgG | No data | - |
| Fusco (D. Cotugno' Hospital)[181] | 2020-03-23 to 2020-04-02 | Campania \| Naples | Sublocal | High | 115 | Median 43 (IQR 32-51.5) | 48.7 | Non-probability | Health care workers and caregivers | 1.7% | Snibe Diagnostic \| MAGLUMI 2019-nCoV IgG (CLIA) \| IgG | 0.7%, 1.0% (FINDDx / MUHC independent evaluation) | - |

| Article Author (Study Organization) | Sampling Dates (YMD) | Location | Geographic Scope | Overall Risk of Bias | Sample Size | Age | Female (%) | Sampling method | Sample frame | Uncorrected Seroprevalence (95% Confidence Interval) | Test Manufacturers \| Names \| Isotypes | Test Sens, Spec (Source) | Corrected Seroprevalence (95% Credible Interval) |
|---|---|---|---|---|---|---|---|---|---|---|---|---|---|
| Apolone (Istituto Nazionale Tumori )[184] | 2019-09-01 to 2020-03-10 | | National | High | 959 | <55 (n=75); 55-65 (n=606); >=65 (n=278) | 41.4 | Non-probability | Patients seeking care for non-COVID-19 reasons | 11.6% | Mazzini et al \| In house ELISA \| IgG, IgM | 85.7%, 98.1% (Test developer / manufacturer evaluation) | 10.7% (4.7-16.1%) |
| **Japan** | | | | | | | | | | | | | |
| Oka (Tokyo National Hospital)[185] | 2020-06-01 to 2020-10-30 | Tokyo | Sublocal | High | 93 | NR | NR | Non-probability | Patients seeking care for non-COVID-19 reasons | 0.0% | Roche Diagnostics \| Elecsys Anti-SARS-CoV-2 \| IgG, IgM | 0.8%, 1.0% (FINDDx / MUHC independent evaluation) | - |
| Matthews (Okinawa Institute of Science and Technology)[186] | 2020-08-01 to 2020-08-31 | Okinawa Prefecture \| Okinawa | Sublocal | High | 634 | 20-40 (n=124), 40-60 (n=74), 60+ (n=4) | NR | Non-probability | Multiple populations | 0.0% | N/A - institution designed \| N/A - institution designed \| NR | 92.5%, 100.0% (Test developer / manufacturer evaluation) | 0.1% (0.0-0.6%) |
| Hibino (Medical Corporation Koshikai)[187] | 2020-05-26 to 2020-08-25 | Tokyo | Local | High | 615 | NR | NR | Non-probability | Non-essential workers and unemployed persons | 34.3% | Aurora Biomed \| COVID-19 IgG / IgM Rapid Test Cassette \| IgG, IgM | 0.8%, 1.0% (FINDDx / MUHC independent evaluation) | - |
| Fukuda (Juntendo Univesity)[188] | 2020-07-06 to 2020-08-21 | Kantō \| Tokyo | Sublocal | Moderate | 4147 | 20-29 (n=1461); 30-39 (n=1245); 40-49 (n=776); 50-59 (n=424); 60-69 (N=193); 70+ (n=48) Mean = 36.8 | 63.9 | Non-probability | Health care workers and caregivers | 0.3% (0.2-0.6%) | Roche Diagnostics \| Elecsys Anti-SARS-CoV-2 electrochemiluminescence immunoassay \| IgG, IgA, IgM | 0.6%, 1.0% (FINDDx / MUHC independent evaluation) | - |
| Hibino (Shonan Fujisawa Tokushukai Hospital)[189] | 2020-06-01 to 2020-07-30 | Kanagawa \| Fujisawa | Sublocal | Unclear | 806 | Median 33 (Range 21-83) | 71.6 | Non-probability | Health care workers and caregivers | 0.7% (0.3-1.6%) | Abbott \| Abbott Architect SARS-CoV-2 IgG \| IgG | 0.4%, 1.0% (FINDDx / MUHC independent evaluation) | - |
| Nishida (Toyonaka Municipal Hospital)[192] | 2020-06-12 to 2020-06-19 | Kansai \| Osaka | Sublocal | Moderate | 925 | Mean 40 (SD 11.8) | 80.0 | Non-probability | Health care workers and caregivers | 0.4% (0.2-1.1%) | Abbott \| Abbott Architect SARS-CoV-2 IgG assay \| IgG | 0.4%, 1.0% (FINDDx / MUHC independent evaluation) | - |

| Article Author (Study Organization) | Sampling Dates (YMD) | Location | Geographic Scope | Overall Risk of Bias | Sample Size | Age | Female (%) | Sampling method | Sample frame | Uncorrected Seroprevalence (95% Confidence Interval) | Test Manufacturers \| Names \| Isotypes | Test Sens, Spec (Source) | Corrected Seroprevalence (95% Credible Interval) |
|---|---|---|---|---|---|---|---|---|---|---|---|---|---|
| Chong (Kanenokuma Hospital)[193] | 2020-06-12 to 2020-06-15 | Kyushu \| Fukuoka City | Sublocal | High | 108 | NR | NR | Non-probability | Assisted living and long-term care facilities | 2.8% | Hangzhou Alltest Biotech Co. Ltd. \| All Test®2019-nCoV IgG/IgM RapidTest Casette \| IgG | 0.6%, 1.0% (FINDDx / MUHC independent evaluation) | - |
| Suda (Hokkaido University Graduate School of Medicine)[194] | 2020-05-01 to 2020-05-31 | Hokkaido \| Sapporo | Sublocal | High | 300 | Mean 66.5 (Range 20-84) | 42.0 | Non-probability | Patients seeking care for non-COVID-19 reasons | 0.0% (0.0-0.0%) | Roche Diagnostics \| Elecsys Anti-SARS-Cov2 Immunoassay \| IgG | 0.8%, 1.0% (FINDDx / MUHC independent evaluation) | - |
| Nakamura (Iwate Prefectural Central Hospital)[195] | 2020-05-18 to 2020-05-29 | Honshu \| Morioka | Sublocal | High | 1000 | Mean 40 (SD 11) | 73.6 | Non-probability | Health care workers and caregivers | 0.0% | Abbott Laboratories: Roche \| Abbott Architect® SARS- CoV- 2 IgG Assay; Roche \| IgG | 97.8%, 97.8% (Used Bastos SR/MA data; no sens, spec, or author adjustment available) | 0.1% (0.0-0.3%) |
| Kobashi ( Hirata Central Hospital)[196] | 2020-05-08 to 2020-05-28 | Fukushima Prefecture \| Hirata | Sublocal | High | 637 | Median 44 (IQR 34, 56) | 75.8 | Non-probability | Health care workers and caregivers | 0.9% | YHLO Biotech Shenzhen; China \| iFlash-SARS-CoV-2 IgM/IgG \| IgG | 1.0%, 0.0% (FINDDx / MUHC independent evaluation) | - |
| Denyer (SoftBank)[198] | 2020-05-12 to 2020-05-18 | | Sublocal | Unclear | 44066 | NR | NR | Non-probability | Multiple populations | 0.4% | Innovita; "Zhejiang Orient Gene Biotech Co.; LTD" \| NR \| NR | No data | - |
| Fujita (National Hospital Organization Kyoto Medical Center)[199] | 2020-04-10 to 2020-04-20 | Kansai \| Kyoto | Sublocal | High | 92 | 20-29 (n=30), 30-39 (n=29), 40-49 (n=21), 50+ (n=12) | 64.1 | Non-probability | Health care workers and caregivers | 5.4% | Inc.; DRG international \| COVID-19 IgG ELISA kits \| IgG | 0.8%, 1.0% (FINDDx / MUHC independent evaluation) | - |
| Suzuki (National Center for Global Health and Medicine)[201] | 2020-02-14 to 2020-04-03 | Tokyo | Sublocal | High | 49 | 20-29 (n = 5), 30-39 (n = 25), 40-49 (n = 11), 50-59 (n = 4), 60+ (n =2) | 69.4 | Non-probability | Health care workers and caregivers | 0.0% | N/A - in house \| N/A - in house \| IgG | No data | - |

| Article Author (Study Organization) | Sampling Dates (YMD) | Location | Geographic Scope | Overall Risk of Bias | Sample Size | Age | Female (%) | Sampling method | Sample frame | Uncorrected Seroprevalence (95% Confidence Interval) | Test Manufacturers \| Names \| Isotypes | Test Sens, Spec (Source) | Corrected Seroprevalence (95% Credible Interval) |
|---|---|---|---|---|---|---|---|---|---|---|---|---|---|
| Suda (Hokkaido University Graduate School of Medicine)[194] | 2020-03-01 to 2020-03-31 | Hokkaido \| Sapporo | Sublocal | High | 300 | Mean 68 (Range 20-80) | 49.0 | Non-probability | Patients seeking care for non-COVID-19 reasons | 0.3% (0.0-1.8%) | Roche Diagnostics \| Elecsys Anti-SARS-Cov2 Immunoassay \| IgG | 0.8%, 1.0% (FINDDx / MUHC independent evaluation) | - |
| **Liechtenstein** | | | | | | | | | | | | | |
| Schaffner (Kantonsspital Graunbunden)[202] | 2020-06-02 to 2020-10-06 | | National | Unclear | 1159 | Median 45 (IQR 39-48) | 53.6 | Non-probability | Multiple populations | 7.8% (6.4-9.5%) | Switzerland Diasorin Euroimmun; Switzerland Abbott Diagnostics Baar; Rotkreuz; Roche Diagnostics \| Elecsys®Anti-SARS-CoV-2 S antibody assay Abbott test Diasorin test Euroimmun test Sugentech, Daejeon, Republic ofKorea; SGTi-flex COVID-19 IgM/IgG \| IgG, IgM | 97.6%, 99.8% (Author-reported independent evaluation) | 7.7% (6.2-9.5%) |
| **New Zealand** | | | | | | | | | | | | | |
| Craigie (Southern Community Laboratories)[208] | 2020-06-04 to 2020-08-04 | Southern Region | Regional | High | 1127 | Median 46 (Range 4-90) | 76.0 | Non-probability | Contacts of COVID patients | 1.0% | Abbott \| Abbott Architect SARS-CoV-2 IgG chemiluminescent microparticle immunoassay \| IgG | 0.4%, 1.0% (FINDDx / MUHC independent evaluation) | - |
| **Norway** | | | | | | | | | | | | | |
| Trieu (University of Bergen)[210] | 2020-03-06 to 2020-04-09 | Midhordland \| Bergen | Local | High | 607 | Median 39 (SD 12.6) | 77.1 | Non-probability | Health care workers and caregivers | 5.3% | Sigma-Aldrich \| In house ELISA test \| IgG | 75.6%, 97.6% (Used Bastos SR/MA data; no sens, spec, or author | 3.7% (1.2-6.5%) |

| Article Author (Study Organization) | Sampling Dates (YMD) | Location | Geographic Scope | Overall Risk of Bias | Sample Size | Age | Female (%) | Sampling method | Sample frame | Uncorrected Seroprevalence (95% Confidence Interval) | Test Manufacturers \| Names \| Isotypes | Test Sens, Spec (Source) | Corrected Seroprevalence (95% Credible Interval) |
|---|---|---|---|---|---|---|---|---|---|---|---|---|---|
| | | | | | | | | | | | | adjustment available) | |
| Kuwelker (University of Bergen)[211] | 2020-02-28 to 2020-04-04 | Hordaland \| Bergen | Local | High | 179 | Median 30 (IQR 17-50) | 55.3 | Non-probability | Contacts of COVID patients | 45.3% | N/A - Author Designed \| N/A - Author Designed \| IgG | 75.6%, 97.6% (Used Bastos SR/MA data; no sens, spec, or author adjustment available) | 58.6% (48.8-68.7%) |
| Cox (University of Bergen)[212] | 2020-02-28 to 2020-04-04 | Western Norway \| Bergen | Local | High | 77 | NR | NR | Probability | Contacts of COVID patients | 31.2% | Amant et al. \| N/A - Author-designed \| IgG, IgA, IgM | 75.6%, 97.6% (Used Bastos SR/MA data; no sens, spec, or author adjustment available) | 39.7% (25.8-53.2%) |
| **Portugal** | | | | | | | | | | | | | |
| Figueiredo-Campos (Universidade de Lisboa)[213] | 2020-05-13 to 2020-07-10 | Lisbon | Sublocal | High | 2715 | NR | NR | Non-probability | Non-essential workers and unemployed persons | 2.6% | N/A - author designed \| N/A - author designed \| IgG | 99.0%, 100.0% (Author-reported independent evaluation) | 2.2% (0.2-3.4%) |
| Figueiredo-Campos (University of Lisbon)[214] | 2020-05-13 to 2020-07-10 | Estremadura \| Lisbon | Sublocal | Moderate | 2571 | NR | NR | Non-probability | Non-essential workers and unemployed persons | 1.5% | et al. [Independent third party]; Krammer \| SARS-CoV-2 ELISA \| IgG, IgA, IgM | 94.7%, 98.0% (Author-reported independent evaluation) | 0.5% (0.0-1.3%) |
| Gonçalves (NOVA University of Lisbon)[216] | 2020-06-15 to 2020-06-30 | Lisboa Region \| Lisbon | Sublocal | Moderate | 1636 | 17-76 | 65.9 | Non-probability | Non-essential workers and unemployed persons | 3.0% | NOVA University of Lisbon \| In-House ELISA anti-SARS-CoV-2 assay \| IgG | 94.7%, 99.5% (Test developer / manufacturer evaluation) | 2.0% (0.2-3.5%) |
| **Republic of Korea** | | | | | | | | | | | | | |
| KBS World (Korea Disease Control and | 2020-08-15 to 2020-10-15 | | National | Unclear | 6900 | NR | 0.0 | Non-probability | Essential non-healthcare workers | 0.4% | NR \| NR \| NR | No data | - |

| Article Author (Study Organization) | Sampling Dates (YMD) | Location | Geographic Scope | Overall Risk of Bias | Sample Size | Age | Female (%) | Sampling method | Sample frame | Uncorrected Seroprevalence (95% Confidence Interval) | Test Manufacturers \| Names \| Isotypes | Test Sens, Spec (Source) | Corrected Seroprevalence (95% Credible Interval) |
|---|---|---|---|---|---|---|---|---|---|---|---|---|---|
| Prevention Agency)[217] | | | | | | | | | | | | | |
| Song (Daegu Catholic University Hospital)[219] | 2020-05-25 to 2020-06-05 | North Gyeongsang \| Daegu | Local | High | 198 | Mean 51.7 (Range 18-82) | 50.0 | Non-probability | Multiple populations | 7.6% | Shin Jin Medics Inc \| DIAKEY COVID-19 IgM/IgG Rapid Test Kit \| IgG | 100.0%, 92.0% (Author-reported independent evaluation) | 2.3% (0.0-6.6%) |
| Ko (Sungkyunkwan University School of Medicine)[221] | 2020-04-26 to 2020-05-09 | | National | High | 309 | Mean 31.1 (SD 7.84) | 84.5 | Non-probability | Health care workers and caregivers | 0.3% | PCL Inc \| PCL COVID-19 Total Ab EIA test \| NR | 98.2%, 100.0% (Test developer / manufacturer evaluation) | 0.2% (0.0-1.0%) |
| Ko (Sungkyunkwan University School of Medicine)[221] | 2020-04-26 to 2020-05-09 | | National | High | 123 | Mean 34.9 (SD 10.9) | 75.6 | Non-probability | Health care workers and caregivers | 0.0% | PCL Inc \| PCL COVID-19 Total Ab EIA test \| NR | 98.2%, 100.0% (Test developer / manufacturer evaluation) | 0.6% (0.0-2.5%) |
| **Singapore** | | | | | | | | | | | | | |
| Kurohi (Singapore National Centre for Infectious Diseases)[222] | 2020-02-15 to 2020-04-15 | Singapore | Sublocal | Unclear | 1096 | NR | NR | Non-probability | Health care workers and caregivers | 0.0% | NR \| NR \| NR | No data | - |
| Ng (National Centre for Infectious Diseases, Singapore)[223] | 2020-01-23 to 2020-04-03 | | National | Moderate | 1150 | Median 35 (IQR 26-51) | 54.2 | Probability | Contacts of COVID patients | 3.8% | N/A - author designed \| N/A - author designed \| NR | 98.9%, 100.0% (Test developer / manufacturer evaluation) | 3.1% (0.5-4.9%) |
| **Spain** | | | | | | | | | | | | | |
| Savirón-Cornudella (University of Zaragoza)[224] | 2020-03-31 to 2020-08-31 | Madrid | Sublocal | High | 260 | NR | 100.0 | Non-probability | Pregnant or parturient women | 6.9% | Vircell; Abbott; Biozek \| Biozek COVID-19 IgG/IgM Rapid Test Cassette IgG Abbott reactive Vircell reactive \| IgG, IgM | No data | - |

| Article Author (Study Organization) | Sampling Dates (YMD) | Location | Geographic Scope | Overall Risk of Bias | Sample Size | Age | Female (%) | Sampling method | Sample frame | Uncorrected Seroprevalence (95% Confidence Interval) | Test Manufacturers \| Names \| Isotypes | Test Sens, Spec (Source) | Corrected Seroprevalence (95% Credible Interval) |
|---|---|---|---|---|---|---|---|---|---|---|---|---|---|
| Garde-Noguera (Hospital Arnau de Vilanova de Valencia, Spain)[225] | 2020-05-02 to 2020-06-30 | Valencia \| Valencia | Local | High | 166 | Median 63 (Range 33-86) | 42.2 | Non-probability | Patients seeking care for non-COVID-19 reasons | 1.8% | DiaSorin \| LIASON SARS-CoV-2 S1/S2IgG test and LIASON SARS-CoV-2 IgM test \| IgG, IgM | 75.6%, 97.6% (Used Bastos SR/MA data; no sens, spec, or author adjustment available) | 0.9% (0.0-3.4%) |
| Prados (IVIRMA Global Headquarters)[226] | 2020-04-27 to 2020-06-26 | | National | Unclear | 6140 | NR | 72.8 | Non-probability | Patients seeking care for non-COVID-19 reasons | 0.7% | Epitope Diagnostics \| EDI Novel Coronavirus COVID-19 ELISA kit \| IgM | 0.6%, 1.0% (FINDDx / MUHC independent evaluation) | - |
| Cabezón-Gutiérrez (Hospital General Universitario Gregorio Maranon)[227] | 2020-06-01 to 2020-06-19 | Community of Madrid \| Torrejon de Ardoz | Sublocal | High | 229 | <50 (n=35); 51-70 (n=122); >70 (n=72) | 54.1 | Non-probability | Patients seeking care for non-COVID-19 reasons | 31.4% | Hangzhou Testsea Biotechnology \| Testsealabs IgG/IgM Rapid Test Cassette \| IgG, IgM | 0.8%, 0.9% (FINDDx / MUHC independent evaluation) | - |
| Camba ( Quironsalud Tenerife Hospital)[228] | 2020-04-27 to 2020-06-15 | Canary Islands \| Tenerife | Sublocal | High | 211 | Median 53.9 (NR) | 55.9 | Non-probability | Patients seeking care for non-COVID-19 reasons | 1.9% (0.1-4.8%) | Roche Diagnostics; Unclear \| Elecsys Anti-SARS-CoV-2, solid phase immunochromatographic analysis \| NR | 97.8%, 97.8% (Used Bastos SR/MA data; no sens, spec, or author adjustment available) | 0.8% (0.0-2.7%) |
| Brotons (University Hospital Sant Joan de Deu)[229] | 2020-04-28 to 2020-06-03 | Catalonia \| Barcelona | Local | Moderate | 1084 | Mean Children 5.9; Adults 40 (SD Children 3.7; adults 10.2) | 45.4 | Non-probability | Contacts of COVID patients | 18.0% | Innovita Tangshan Biological Technology Co \| 2019-n-CoV Ab Test \| IgG, IgM | 0.6%, 1.0% (FINDDx / MUHC independent evaluation) | - |
| Perez-Garcia (Hospital Universitario Príncipe de Asturias)[231] | 2020-05-01 to 2020-05-30 | Madrid | Sublocal | High | 2424 | Median 42.6 (HCW without evidence of infection); 50.0 (HCW symptomatic with positive serology); | 80.4 | Non-probability | Health care workers and caregivers | 22.4% | AllTest Biotech \| AllTest COVID-19 IgG/IgM kit \| IgG, IgM | 0.6%, 1.0% (FINDDx / MUHC independent evaluation) | - |

| Article Author (Study Organization) | Sampling Dates (YMD) | Location | Geographic Scope | Overall Risk of Bias | Sample Size | Age | Female (%) | Sampling method | Sample frame | Uncorrected Seroprevalence (95% Confidence Interval) | Test Manufacturers \| Names \| Isotypes | Test Sens, Spec (Source) | Corrected Seroprevalence (95% Credible Interval) |
|---|---|---|---|---|---|---|---|---|---|---|---|---|---|
| | | | | | | 38.6 (HCW asymptomatic with positive serology) (IQR 29.9-54.8 (HCW without evidence of infection); 36.9–57.3 (HCW symptomatic with positive serology); 28.5–52.1 (HCW asymptomatic with positive serology)) | | | | | | | |
| Fabre (Hospital Clínico Universitario Lozano Blesa)[232] | 2020-04-27 to 2020-05-29 | Zaragoza \| Zaragoza | Sublocal | High | 169 | NR | 100.0 | Non-probability | Pregnant or parturient women | 8.3% | Abbott; Guangzhou Wondfo Biotech Co.; Vircell Microbiologists \| Wondfo SARS-CoV-2 Antibody Test , IgG-CMIA; Abbott SARS-CoV-2 IgG Assay; Vircell COVID- 19 VIRCLIA IgM/IgA, \| IgG, IgM | 66.0%, 96.6% (Used Bastos SR/MA data; no sens, spec, or author adjustment available) | 7.3% (1.1-14.0%) |
| Barallat (Institut Català de la Salut)[233] | 2020-05-04 to 2020-05-22 | Barcelona | Local | High | 7563 | Mean 43.8 (SD 12.43) | 76.0 | Non-probability | Health care workers and caregivers | 9.4% | Italy; DiaSorin; Vercelli \| SARS-CoV-2 S1/S2 IgG LIAISON® \| IgG | 0.8%, 1.0% (FINDDx / MUHC independent evaluation) | - |
| Viñuela (University Hospital Gregorio Marañón)[234] | 2020-05-06 to 2020-05-21 | Madrid | Local | High | 100 | Median 32 (Range 16-44) | 100.0 | Non-probability | Pregnant or parturient women | 13.0% | Abbott \| Abbott Architect SARS-CoV-2 IgG assay \| IgG | 0.4%, 1.0% (FINDDx / MUHC independent evaluation) | - |

| Article Author (Study Organization) | Sampling Dates (YMD) | Location | Geographic Scope | Overall Risk of Bias | Sample Size | Age | Female (%) | Sampling method | Sample frame | Uncorrected Seroprevalence (95% Confidence Interval) | Test Manufacturers \| Names \| Isotypes | Test Sens, Spec (Source) | Corrected Seroprevalence (95% Credible Interval) |
|---|---|---|---|---|---|---|---|---|---|---|---|---|---|
| Soriano ( Universidad Autónoma de Madrid)[235] | 2020-04-26 to 2020-05-16 | Community of Madrid \| Madrid | Local | High | 674 | Median 42 (Range 18-89) | 70.2 | Non-probability | Multiple populations | 13.8% | PCL Inc. \| PCL COVID19 IgG/IgM Rapid Gold \| IgG, IgM | 0.9%, 1.0% (FINDDx / MUHC independent evaluation) | - |
| Villalaın (The Universidad Complutense de Madrid)[237] | 2020-02-28 to 2020-05-10 | Madrid | Sublocal | Moderate | 769 | Median 32 (IQR 11) | 100.0 | Non-probability | Pregnant or parturient women | 11.2% | NR \| NR \| IgG | 75.6%, 97.6% (Used Bastos SR/MA data; no sens, spec, or author adjustment available) | 12.0% (8.8-15.3%) |
| Gras-Valentí (COVID-19 Group of the Infections Commission)[238] | 2020-04-24 to 2020-05-08 | Alacant \| Alicante | Sublocal | Unclear | 4179 | NR | 73.3 | Non-probability | Health care workers and caregivers | 6.6% (5.8-7.3%) | NR \| NR \| IgG | 75.6%, 97.6% (Used Bastos SR/MA data; no sens, spec, or author adjustment available) | 5.6% (3.9-7.1%) |
| Crovetto (Universitat de Barcelona)[239] | 2020-04-14 to 2020-05-05 | Barcelona | Sublocal | High | 874 | NR | 100.0 | Non-probability | Pregnant or parturient women | 14.0% | Ortho Clinical Diagnostics; Vircell Microbiologist \| VIRCLIA, VITROS (Ortho) \| IgG, IgA, IgM | No data | - |
| Valdivia (Hospital Clínico Universitario)[240] | 2020-04-13 to 2020-04-30 | Valencia \| Valencia | Local | High | 1153 | NR | NR | Non-probability | Health care workers and caregivers | 3.5% | Snibe Diagnostic \| MAGLUMI 2019-nCov IgG/IgM CLIA \| IgG, IgM | 95.6%, 96.0% (Test developer / manufacturer evaluation) | 0.9% (0.0-2.6%) |
| Galán (Hospital Universitario Fundación Alcorcón)[241] | 2020-04-14 to 2020-04-27 | Madrid | Sublocal | High | 2590 | Mean 43.8 (SD 11.1) | 73.9 | Non-probability | Health care workers and caregivers | 31.6% | Palex \| Diapro \| IgG | 88.9%, 91.8% (Author-reported independent evaluation) | 26.9% (16.9-35.2%) |
| Olalla ( Hospital Costa del Sol )[242] | 2020-04-15 to 2020-04-25 | Marbella | Sublocal | High | 498 | Mean 41.5 (95% CI 40.8-42.3) | 71.1 | Non-probability | Health care workers and caregivers | 2.2% | "Zhejiang Orient Gene Biotech Co.; LTD" \| COVID-19 IgG/IgM Rapid Test \| IgG, IgM | 0.4%, 1.0% (FINDDx / MUHC independent evaluation) | - |

| Article Author (Study Organization) | Sampling Dates (YMD) | Location | Geographic Scope | Overall Risk of Bias | Sample Size | Age | Female (%) | Sampling method | Sample frame | Uncorrected Seroprevalence (95% Confidence Interval) | Test Manufacturers \| Names \| Isotypes | Test Sens, Spec (Source) | Corrected Seroprevalence (95% Credible Interval) |
|---|---|---|---|---|---|---|---|---|---|---|---|---|---|
| Borraz (Hospital San Juan de Dios)[244] | 2020-03-20 to 2020-04-21 | Aragon \| Zaragoza | Sublocal | High | 289 | Median 41.9 (SD 12.7 ) | 77.2 | Non-probability | Health care workers and caregivers | 5.9% | Madrid; Master Labor S.L. \| 2019-nCoV IgG/IgM Rapid Test \| IgG, IgM | 100.0%, 98.0% (Test developer / manufacturer evaluation) | 3.1% (0.0-6.2%) |
| Borraz (Hospital San Juan de Dios)[244] | 2020-03-20 to 2020-04-21 | Aragon \| Zaragoza | Sublocal | High | 229 | Median 74.4 (SD 13.9 ) | 43.2 | Non-probability | Patients seeking care for non-COVID-19 reasons | 7.0% | Madrid; Master Labor S.L. \| 2019-nCoV IgG/IgM Rapid Test \| IgG, IgM | 100.0%, 98.0% (Test developer / manufacturer evaluation) | 4.4% (0.4-8.7%) |
| Dacosta-Urbieta (Hospital Clinico Universitario de Santiago de Compostela)[245] | 2020-04-14 to 2020-04-16 | Galicia \| Santiago de Compostela | Sublocal | High | 175 | NR | NR | Non-probability | Health care workers and caregivers | 4.0% (1.1-6.9%) | Genobio Pharmaceutical (Shanghai; China) \| Virusee Immunochromatographic Rapid Method \| IgG, IgM | No data | - |
| Martin (Gerencia de Atención Primaria del Área de Salud de León, Spain)[246] | 2020-04-01 to 2020-04-15 | Léon \| Léon | Local | High | 676 | Mean 48.5 (SD 12.8) | 74.1 | Non-probability | Health care workers and caregivers | 7.4% (5.5-10.0%) | Hangzhou All Test Biotech Co Ltd \| All Test®2019-nCoV IgG/IgM RapidTest Casette \| IgG, IgM | 0.0%, 0.0% (Used author-adjusted estimate) | 7.4% (5.6-9.6%) |
| Fernandez (Hospital Universitario de Fuenlabrada)[247] | 2020-05-13 to 2020-04-14 | Fuenlabrada | Sublocal | High | 2439 | Mean 42.1 (Range 18-65) | 78.4 | Non-probability | Health care workers and caregivers | 16.9% | Snibe diagnostic \| Maglumi 2000 \| IgG | 0.9%, 1.0% (FINDDx / MUHC independent evaluation) | - |
| **Sweden** | | | | | | | | | | | | | |
| Dopico (Karolinska Institutet)[248] | 2020-03-14 to 2020-12-11 | Uppland \| Stockholm | Local | High | 4100 | NR | NR | Probability | Multiple populations | 14.8% (12.2-18.0%) | NR - In-house \| Anti-SARS-CoV-2 ELISA \| IgG | 0.0%, 0.0% (Used author-adjusted estimate) | 14.8% (13.7-15.9%) |
| Rashid-Abdi (The Hospital of Vastmanland )[249] | 2020-05-04 to 2020-08-19 | Vasteras | Sublocal | High | 131 | Mean 39 (SD 12) | 84.0 | Non-probability | Health care workers and caregivers | 19.1% | Abbott \| Abbott’s SARS-CoV-2 IgG immunoassay, \| IgG | 0.4%, 1.0% (FINDDx / MUHC independent evaluation) | - |

| Article Author (Study Organization) | Sampling Dates (YMD) | Location | Geographic Scope | Overall Risk of Bias | Sample Size | Age | Female (%) | Sampling method | Sample frame | Uncorrected Seroprevalence (95% Confidence Interval) | Test Manufacturers \| Names \| Isotypes | Test Sens, Spec (Source) | Corrected Seroprevalence (95% Credible Interval) |
|---|---|---|---|---|---|---|---|---|---|---|---|---|---|
| Laursen (Copenhagen University Hospital)[59] | 2020-06-22 to 2020-08-10 | | National | High | 1248 | <40 (n=374); 40-60 (n=687); 60+ (n=187) | 67.1 | Non-probability | Health care workers and caregivers | 8.3% | Zhuhai; China; Guangzhou Wondfo Biotech; Livson Diagnostics; Gunangdong \| Livzon lateral flow test, WONDFO lateral flow \| IgG, IgM | 66.0%, 96.6% (Used Bastos SR/MA data; no sens, spec, or author adjustment available) | 7.8% (5.2-10.5%) |
| Lidström (Uppsala University )[252] | 2020-05-27 to 2020-06-25 | Uppsala | Regional | Moderate | 8679 | Mean 45 (IgG negative); 42 (IgG positive) (Range 18-85 (IgG negative); 18-78 (IgG positive)) | 76.7 | Probability | Health care workers and caregivers | 6.6% | Abbott \| Abbott Architect SARS-CoV-2 IgG assay \| IgG | 0.4%, 1.0% (FINDDx / MUHC independent evaluation) | - |
| Hassan (Karolinska University Hospital)[254] | 2020-05-11 to 2020-06-17 | Stockholm | Sublocal | High | 403 | Median 43 (NR) | 52.9 | Non-probability | Assisted living and long-term care facilities | 20.1% | (Luminex Corp) \| FlexMap3D \| IgG | 99.2%, 99.8% (Author-reported independent evaluation) | 20.1% (16.2-24.1%) |
| Rudberg (Danderyd Hospital)[257] | 2020-04-14 to 2020-05-08 | Sodermanland \| Stockholm | Sublocal | Moderate | 2149 | Mean 44 (SD 12) | 84.5 | Non-probability | Health care workers and caregivers | 19.1% | Author designed \| Author designed \| IgG | 99.2%, 99.8% (Test developer / manufacturer evaluation) | 19.0% (17.2-20.8%) |
| Lindahl (Uppsala University )[258] | 2020-04-01 to 2020-04-20 | Stockholm | Local | High | 1005 | 36-50 (n=133/332); 51-65 (n=116/332) | 24.3 | Non-probability | Health care workers and caregivers | 22.9% (20.4-25.7%) | Zhejiang Orient Gene Biotech Co Ltd \| COVID-19 IgG / IgM Rapid Test Cassette \| IgG, IgM | 0.4%, 1.0% (FINDDx / MUHC independent evaluation) | - |
| **Switzerland** | | | | | | | | | | | | | |
| Kahlert (Cantonal Hospital St Gallen)[261] | 2020-06-22 to 2020-08-15 | Canton of St. Gallen | Regional | Moderate | 4664 | Median 38.3 (IQR 29.7-49.5) | 78.3 | Non-probability | Health care workers and caregivers | 3.0% | Switzerland ELISA: Euroimmune; Rotkreuz; ECLIA: Roche Diagnostics; | No data | - |

| Article Author (Study Organization) | Sampling Dates (YMD) | Location | Geographic Scope | Overall Risk of Bias | Sample Size | Age | Female (%) | Sampling method | Sample frame | Uncorrected Seroprevalence (95% Confidence Interval) | Test Manufacturers \| Names \| Isotypes | Test Sens, Spec (Source) | Corrected Seroprevalence (95% Credible Interval) |
|---|---|---|---|---|---|---|---|---|---|---|---|---|---|
| | | | | | | | | | | | Germany \| electro-chemiluminescence immunoassay (ECLIA) and Enzyme-linked Immunosorbent Assay (ELISA) \| IgG, IgA | | |
| Fenwick (Lausanne University Hospital and University of Lausanne)[267] | 2020-05-04 to 2020-06-27 | Vaud | Regional | High | 177 | NR | NR | Non-probability | Contacts of COVID patients | 32.2% | Roche \| Elecsys anti-SARS-CoV-2 assay \| NR | 0.9%, 1.0% (FINDDx / MUHC independent evaluation) | - |
| Dupraz (University of Lausanne)[268] | 2020-05-04 to 2020-06-27 | Vaud | Regional | Moderate | 69 | Mean 47.8 (SD 17) | 56.5 | Probability | Contacts of COVID patients | 17.4% | Switzerland; Lausanne University Hospital \| in-house Luminex S protein trimer IgG Assay \| IgG | 96.7%, 99.2% (Test developer / manufacturer evaluation) | 17.2% (7.3-28.0%) |
| Dupraz (University of Lausanne)[268] | 2020-05-04 to 2020-06-27 | Vaud | Regional | Moderate | 302 | Mean 37 (SD 21.3) | 51.7 | Probability | Contacts of COVID patients | 53.0% | Switzerland; Lausanne University Hospital \| in-house Luminex S protein trimer IgG Assay \| IgG | 96.7%, 99.2% (Test developer / manufacturer evaluation) | 55.3% (47.2-64.0%) |
| Kohler (Cantonal Hospital St. Gallen)[270] | 2020-03-19 to 2020-04-03 | St. Gallen \| St. Gallen | Regional | High | 1012 | Median 38.3 (Range 16.9-64.8) | 75.2 | Non-probability | Health care workers and caregivers | 1.0% | Daejeon; Basel; Yuseong-gu; IL); a chemiluminescence microparticle immunoassay (CMIA; Roche Diagnostics; and an electro-chemiluminescence immunoassay (ECLIA; Sugentech; Abbott Diagnostics; Lake Bluff; | No data | - |

| Article Author (Study Organization) | Sampling Dates (YMD) | Location | Geographic Scope | Overall Risk of Bias | Sample Size | Age | Female (%) | Sampling method | Sample frame | Uncorrected Seroprevalence (95% Confidence Interval) | Test Manufacturers \| Names \| Isotypes | Test Sens, Spec (Source) | Corrected Seroprevalence (95% Credible Interval) |
|---|---|---|---|---|---|---|---|---|---|---|---|---|---|
| | | | | | | | | | | | lateral flow immunochromatographic assay (LFIA; Switzerland); Republic of Korea) \| NR \| IgG | | |
| **The United Kingdom** | | | | | | | | | | | | | |
| Taubel (Richmond Pharmacology Ltd)[273] | 2020-03-19 to 2020-12-02 | England \| London | Local | High | 6061 | 3-89 | 11.3 | Non-probability | Multiple populations | 6.4% | NR \| NR \| IgG | No data | - |
| Lumley (Oxford University Hospitals)[274] | 2020-04-23 to 2020-11-30 | | Local | Moderate | 12541 | Median 38 (IQR 29 to 49) | 73.9 | Probability | Health care workers and caregivers | 10.1% | University of Oxford \| N/A - Author Designed \| IgG | 99.1%, 99.0% (Test developer / manufacturer evaluation) | 8.7% (4.7-11.2%) |
| Abo-Leyah (Ninewells Hospital and Medical School)[280] | 2020-05-28 to 2020-09-02 | Scotland | Regional | High | 2062 | Mean 44.8 (NR) | 81.7 | Non-probability | Health care workers and caregivers | 14.5% | Siemens \| Siemens SARS-CoV-2 antibody assay \| NR | 0.9%, 1.0% (FINDDx / MUHC independent evaluation) | - |
| Harris (Public Health England)[283] | 2020-03-23 to 2020-08-20 | | Sublocal | Moderate | 2247 | NR | NR | Non-probability | Health care workers and caregivers | 12.1% | Euroimmun \| Euroimmun anti-SARS-CoV-2 ELISA (IgG) serology assay \| IgG | 0.6%, 0.9% (FINDDx / MUHC independent evaluation) | - |
| Roarty (Queens University Belfast)[284] | 2020-06-26 to 2020-08-15 | Belfast, London, Glasgow, Manchester, Cardiff | National | Moderate | 849 | 2-15 | NR | Non-probability | Family of essential workers | 7.7% (6.0-9.6%) | Basel; Switzerland; DiaSorin; Roche; Saluggia; Italy \| Elecys ANti-SARS-CoV-2 Total Antibody assay; LIAISON SARS-CoV-2 S1/S2 IgG assay \| IgG | No data | - |

| Article Author (Study Organization) | Sampling Dates (YMD) | Location | Geographic Scope | Overall Risk of Bias | Sample Size | Age | Female (%) | Sampling method | Sample frame | Uncorrected Seroprevalence (95% Confidence Interval) | Test Manufacturers \| Names \| Isotypes | Test Sens, Spec (Source) | Corrected Seroprevalence (95% Credible Interval) |
|---|---|---|---|---|---|---|---|---|---|---|---|---|---|
| Cooper (Cambridge University Hospitals NHS Foundation Trust)[286] | 2020-06-10 to 2020-08-07 | England \| Cambridge | Sublocal | Moderate | 5698 | Median 38 (Range 17-83) | 77.3 | Non-probability | Health care workers and caregivers | 7.2% | Siemens Healthcare Limited \| Centaur XP SARS-Cov-2 Total Antibody assay \| NR | 0.9%, 1.0% (FINDDx / MUHC independent evaluation) | - |
| Black (University Hospitals of Leicester NHS Trust)[287] | 2020-05-29 to 2020-08-03 | England \| Leicester | Sublocal | High | 200 | Mean 45.3 (SD 12) | 75.0 | Non-probability | Health care workers and caregivers | 12.0% | Abbott Laboratories \| Abbott SARS CoV-2 assay \| IgG | 0.4%, 1.0% (FINDDx / MUHC independent evaluation) | - |
| Claudiani (Imperial College London)[289] | 2020-06-01 to 2020-07-27 | England \| London | Sublocal | High | 161 | Median 54 (Range 18-92) | 41.6 | Non-probability | Patients seeking care for non-COVID-19 reasons | 11.2% | Imperial College London \| Imperial Hybrid DABA \| NR | No data | - |
| Martin (University of Leicester)[291] | 2020-05-29 to 2020-07-13 | East Midlands \| Leicester | Sublocal | Moderate | 10662 | Median 44 (IQR 33-53) | 79.8 | Non-probability | Health care workers and caregivers | 10.8% | Abbott \| Abbott SARS-CoV- 2 IgG assay \| IgG | 0.4%, 1.0% (FINDDx / MUHC independent evaluation) | - |
| Hanrath (Newcastle-upon-Tyne Hospitals NHS Trust)[292] | 2020-05-29 to 2020-07-06 | England \| Newcastle-Upon-Tyne | Sublocal | High | 11103 | Median 43 (IQR 32-53) | 77.0 | Non-probability | Health care workers and caregivers | 8.4% | Roche Diagnostics \| Roche Anti-SARS-CoV-2 serology assay \| IgG | No data | - |
| Prendecki (Imperial College London)[293] | 2020-06-01 to 2020-07-03 | Greater London \| London | Sublocal | High | 855 | Median 57 (Range 45-77) | 34.7 | Non-probability | Patients seeking care for non-COVID-19 reasons | 8.1% (6.4-10.1%) | Abbott \| Abbott Architect SARS-CoV-2 IgG assay \| IgG | 68.4%, 100.0% (Author-reported independent evaluation) | 10.0% (2.1-16.0%) |
| Waterfield (Royal Belfast Hospital for Sick Children)[294] | 2020-04-16 to 2020-07-03 | Wales, England, Scotland, Ireland \| Belfast, London, Glasgow, Manchester, Cardiff | National | High | 992 | Median 10.1 (Range 2.03-15.99) | 48.7 | Non-probability | Family of essential workers | 3.1% | DiaSorin \| - Spike protein assays (DiaSorin LIAISON SARS CoV-2 S1/ S2 IgG assay) \| IgG | 0.8%, 1.0% (FINDDx / MUHC independent evaluation) | - |

| Article Author (Study Organization) | Sampling Dates (YMD) | Location | Geographic Scope | Overall Risk of Bias | Sample Size | Age | Female (%) | Sampling method | Sample frame | Uncorrected Seroprevalence (95% Confidence Interval) | Test Manufacturers \| Names \| Isotypes | Test Sens, Spec (Source) | Corrected Seroprevalence (95% Credible Interval) |
|---|---|---|---|---|---|---|---|---|---|---|---|---|---|
| Rowland (Public Health England)[296] | 2020-06-03 to 2020-06-30 | Greater London \| London | Sublocal | High | 327 | 0-39 (n=142); 40-49 (n=149); 60+ (n=36) | 77.7 | Non-probability | Health care workers and caregivers | 47.1% | Abbott \| Abbott Architect nucleocapsid IgG assay \| IgG | 0.4%, 1.0% (FINDDx / MUHC independent evaluation) | 52.2% (46.1-57.4%) |
| Graham (Imperial College London)[297] | 2020-06-01 to 2020-06-30 | Greater London \| London | Sublocal | High | 241 | NR | NR | Non-probability | Assisted living and long-term care facilities | 71.8% (66.0-77.0%) | Abbott \| Abbott Architect nucleocapsid IgG assay \| IgG | 0.4%, 1.0% (FINDDx / MUHC independent evaluation) | - |
| Davis (King's College London)[298] | 2020-06-01 to 2020-06-30 | England \| London | Sublocal | High | 1882 | Median 37 (NR) | 71.1 | Non-probability | Multiple populations | 7.0% | Sure Screen Diagnostics \| SureScreen Diagnostics Rapid COVID-19 IgG/IgM Immunoassay Test Cassette \| IgG, IgM | 94.0%, 96.3% (Author-reported independent evaluation) | 3.6% (2.1-5.2%) |
| Gulraj (Gibraltar Government )[300] | 2020-06-03 to 2020-06-17 | Gibraltar | Regional | Unclear | 1247 | NR | NR | Non-probability | Health care workers and caregivers | 1.4% | NR \| NR \| IgG | No data | - |
| Lumley (University of Oxford)[301] | 2020-04-14 to 2020-06-15 | Oxford | Local | Moderate | 1000 | Median 32 (IQR 28-35) | 100.0 | Non-probability | Pregnant or parturient women | 5.3% (4.0-6.9%) | University of Oxford \| IgG ELISA \| IgG | 99.1%, 99.0% (Test developer / manufacturer evaluation) | 3.7% (0.4-6.2%) |
| Jones (North Bristol NHS Trust)[302] | 2020-01-15 to 2020-06-15 | England \| North Somerset, Bristol, South Gloucestershire | Sublocal | High | 6858 | <=20years (n=114); 21-30 (n=1757); 31-40 (n=1624); 41-50 (n=1536); 51-60 (n=1408); 61-70 (n=402) | 77.8 | Non-probability | Health care workers and caregivers | 9.3% | Abbott laboratories; RocheDiagnostics \| Abbott SARS-CoV-2 IgG chemiluminescent microparticle assay; RocheTMElecsysR©Anti-SARS-CoV-2(IgG/IgM)electro-chemiluminescentimmunoassay \| IgG, IgM | 97.8%, 97.8% (Used Bastos SR/MA data; no sens, spec, or author adjustment available) | 7.4% (6.5-8.3%) |

| Article Author (Study Organization) | Sampling Dates (YMD) | Location | Geographic Scope | Overall Risk of Bias | Sample Size | Age | Female (%) | Sampling method | Sample frame | Uncorrected Seroprevalence (95% Confidence Interval) | Test Manufacturers \| Names \| Isotypes | Test Sens, Spec (Source) | Corrected Seroprevalence (95% Credible Interval) |
|---|---|---|---|---|---|---|---|---|---|---|---|---|---|
| Favara (Cambridge University Hospitals NHS Foundation Trust)[303] | 2020-06-15 to 2020-06-15 | East of England | Regional | High | 434 | Median 41 (Range 19-66 ) | 82.0 | Non-probability | Health care workers and caregivers | 18.4% | Cambridge University Hospitals \| Luminex \| IgG | 84.0%, 100.0% (Test developer / manufacturer evaluation) | 21.6% (15.1-28.4%) |
| Pallett (Chelsea and Westminster Hospital NHS Trust)[304] | 2020-04-08 to 2020-06-12 | England \| London | Local | High | 504 | Mean 42.4 (Range 20.3-72.1) | 59.1 | Non-probability | Health care workers and caregivers | 10.6% (7.6-13.6%) | CTK Biotech; Zhuhai Encode Medical Engineering \| Onsite CTK Biotech COVID-19 split IgG/IgM Rapid Test, Encode SARS-CoV-2 split IgM/IgG One Step Rapid Test Device \| IgG, IgM | 66.0%, 96.6% (Used Bastos SR/MA data; no sens, spec, or author adjustment available) | 11.5% (7.4-16.2%) |
| Baxendale (Royal Papworth Hospital NHS Trust)[305] | 2020-04-20 to 2020-06-10 | England \| Cambridge | Sublocal | High | 500 | Median 42 (IQR 33-51) | 61.6 | Non-probability | Health care workers and caregivers | 14.0% | Addenbrookes Hospital NHS Trust \| Luminex based multiplexed particle flow cytometry assay \| IgG | 97.0%, 100.0% (Author-reported independent evaluation) | 14.2% (10.6-17.7%) |
| Razvi (Gateshead Health NHS Foundation Trust)[307] | 2020-05-28 to 2020-06-08 | England \| Gateshead | Sublocal | High | 2521 | Mean 42.5 (SD 12.6) | 91.9 | Non-probability | Health care workers and caregivers | 19.4% | Roche \| Elecsys Anti-SARS-CoV-2 kit \| IgG, IgM | 0.8%, 1.0% (FINDDx / MUHC independent evaluation) | - |
| Favara (The Queen Elizabeth Hospital)[308] | 2020-06-01 to 2020-06-07 | England \| King's Lynn | Sublocal | High | 70 | NR | NR | Non-probability | Health care workers and caregivers | 21.4% | Unclear \| Luminex SARS-CoV-2 IgG multiplex particle-based flow cytometry assay \| IgG | No data | - |
| Bampoe (University College London)[309] | 2020-05-11 to 2020-06-05 | England \| London | Sublocal | High | 200 | Mean 37 (IQR 30-37) | 83.5 | Non-probability | Health care workers and caregivers | 14.5% (9.9-20.1%) | Abbott \| Abbott Architect SARS-CoV-2 IgG assay \| IgG | 0.4%, 1.0% (FINDDx / MUHC independent evaluation) | - |

| Article Author (Study Organization) | Sampling Dates (YMD) | Location | Geographic Scope | Overall Risk of Bias | Sample Size | Age | Female (%) | Sampling method | Sample frame | Uncorrected Seroprevalence (95% Confidence Interval) | Test Manufacturers \| Names \| Isotypes | Test Sens, Spec (Source) | Corrected Seroprevalence (95% Credible Interval) |
|---|---|---|---|---|---|---|---|---|---|---|---|---|---|
| Grant (Whittington Health NHS Trust)[310] | 2020-05-15 to 2020-06-05 | England \| London | Sublocal | High | 2004 | Mean 40.21 (Range 18-73) | NR | Non-probability | Health care workers and caregivers | 31.6% | Roche Diagnostics \| Elecsys Anti-SARS-Cov2 Immunoassay \| IgG, IgM | 0.8%, 1.0% (FINDDx / MUHC independent evaluation) | - |
| McCafferty (Barts Health NHS Trust )[311] | 2020-05-01 to 2020-06-04 | London | Sublocal | High | 1253 | NR | NR | Non-probability | Patients seeking care for non-COVID-19 reasons | 18.0% | Roche \| Elecsys Anti-SARS-CoV-2 test \| IgG, IgM | 0.8%, 1.0% (FINDDx / MUHC independent evaluation) | - |
| Ladhani (Immunisation and Countermeasures Division)[314] | 2020-05-01 to 2020-05-31 | Belfast, London, Glasgow, Manchester, Cardiff | National | High | 44 | Median 6.3 (seronegative); 9.1 (seropositive) (IQR 3.3-8.6 (seronegative); 5.9-11.6 (seropositive)) | 54.5 | Non-probability | Contacts of COVID patients | 36.2% (9.0-63.4%) | Abbott Commerce \| Abbott SARS-CoV-2 IgG \| IgG | 0.4%, 1.0% (FINDDx / MUHC independent evaluation) | - |
| Khalil (St George's University of London)[315] | 2020-05-15 to 2020-05-28 | England \| London | Sublocal | High | 190 | NR | NR | Non-probability | Health care workers and caregivers | 22.0% | Abbott \| Abbott Architect SARS-CoV-2 IgG assay \| IgG | 0.4%, 1.0% (FINDDx / MUHC independent evaluation) | - |
| Poulikakos (Salford Royal NHS Foundation Trust)[318] | 2020-05-04 to 2020-05-06 | England | Sublocal | High | 281 | NR | 73.0 | Non-probability | Health care workers and caregivers | 6.0% | Snibe Diagnostic \| MAGLUMI 2019-nCoV IgG (CLIA) \| IgG | 0.7%, 1.0% (FINDDx / MUHC independent evaluation) | - |
| Shields (University Hospitals Birmingham NHS Foundation Trust)[321] | 2020-04-24 to 2020-04-25 | England \| Birmingham | Sublocal | High | 516 | Median 42 (IQR 30-51) | 75.2 | Non-probability | Health care workers and caregivers | 24.4% | University of Birmingham Clinical Immunology Service \| ELISA developed inhouse \| IgG, IgA, IgM | 100.0%, 97.8% (Test developer / manufacturer evaluation) | 22.5% (18.6-26.7%) |
| Sikora (Rutherford Health)[322] | 2020-04-14 to 2020-04-24 | England \| Bedlington, Liverpool, Newport, Reading | Local | High | 161 | Mean 43 (NR) | 50.3 | Non-probability | Health care workers and caregivers | 1.2% | Sugentech \| Sugentech SGTi-flex COVID-19 IgM/IgG \| IgG | 66.0%, 96.6% (Used Bastos SR/MA data; no sens, spec, or author | 0.8% (0.0-3.2%) |

| Article Author (Study Organization) | Sampling Dates (YMD) | Location | Geographic Scope | Overall Risk of Bias | Sample Size | Age | Female (%) | Sampling method | Sample frame | Uncorrected Seroprevalence (95% Confidence Interval) | Test Manufacturers \| Names \| Isotypes | Test Sens, Spec (Source) | Corrected Seroprevalence (95% Credible Interval) |
|---|---|---|---|---|---|---|---|---|---|---|---|---|---|
| | | | | | | | | | | | | adjustment available) | |
| Ladhani (Public Health England)[323] | 2020-04-10 to 2020-04-13 | England \| London | Local | High | 394 | 19-39 (n=66); 40-59 (n=113); 60-79 (n=71); 80+ (n=144) | 75.9 | Non-probability | Assisted living and long-term care facilities | 77.9% (73.6-81.7%) | In House: PHE England \| SARS-CoV-2 infected virus lysate assay \| NR | 75.6%, 97.6% (Used Bastos SR/MA data; no sens, spec, or author adjustment available) | 98.4% (94.6-100.0%) |
| Houlihan (University College London Hospitals)[324] | 2020-03-26 to 2020-04-08 | England \| London | Sublocal | High | 181 | Median 34 (IQR 29-44) | 67.4 | Non-probability | Health care workers and caregivers | 45.3% | in-house flow cytometry and ELISA assays developed as part of the Crick-COVID-Consortium \| NR \| IgG, IgA, IgM | No data | - |
| **United States of America** | | | | | | | | | | | | | |
| Godbout (Virginia Commonwealth University)[330] | 2020-07-27 to 2020-10-02 | Virginia \| Richmond | Sublocal | High | 1962 | Mean 37 (Range 19-75) | 77.3 | Non-probability | Health care workers and caregivers | 1.4% | Abbott Laboratories \| Abbott Architect SARS-CoV-2 IgG antibody immunoassay \| IgG | 0.4%, 1.0% (FINDDx / MUHC independent evaluation) | - |
| Jimenez-Kurlander (Memorial Sloan Kettering Cancer Center)[333] | 2020-05-05 to 2020-09-10 | New York \| New York City | Sublocal | High | 257 | Median 6.9 (Range 1-18.6) | 65.0 | Non-probability | Patients seeking care for non-COVID-19 reasons | 7.8% | Abbott \| Abbott architect \| IgG | 0.4%, 1.0% (FINDDx / MUHC independent evaluation) | - |
| Halbrook (University of California Los Angeles)[335] | 2020-05-19 to 2020-08-31 | California | Local | Moderate | 679 | 20-29 (n = 53) 30-39 ( n = 237) 40-49 (n = 182) 50-59 (n = 182) 60+ (n = 15) | 6.0 | Non-probability | Essential non-healthcare workers | 8.1% | NR \| NR \| IgG | 75.6%, 97.6% (Used Bastos SR/MA data; no sens, spec, or author adjustment available) | 7.5% (4.8-10.9%) |
| Halbrook (University of | 2020-04-08 to 2020-08-31 | California | Local | Moderate | 1108 | 20-29 (n = 172) 30-39 (n = 418) 40- | 64.1 | Non-probability | Health care workers and caregivers | 3.9% | NR \| NR \| IgG | 75.6%, 97.6% (Used Bastos SR/MA data; | 2.0% (0.2-3.7%) |

| Article Author (Study Organization) | Sampling Dates (YMD) | Location | Geographic Scope | Overall Risk of Bias | Sample Size | Age | Female (%) | Sampling method | Sample frame | Uncorrected Seroprevalence (95% Confidence Interval) | Test Manufacturers \| Names \| Isotypes | Test Sens, Spec (Source) | Corrected Seroprevalence (95% Credible Interval) |
|---|---|---|---|---|---|---|---|---|---|---|---|---|---|
| California Los Angeles)[335] | | | | | | 49 (n = 235) 50-59 (n = 148) 60+ (n = 55) | | | | | | no sens, spec, or author adjustment available) | |
| Brant-Zawadzki (Hoag Memorial Hospital Presbyterian)[337] | 2020-07-15 to 2020-08-15 | California | Local | High | 92 | Mean 42.05 (SD 8.27) | 13.0 | Non-probability | Essential non-healthcare workers | 4.3% (1.2-10.8%) | Ortho Diagnostics \| Ortho Clinical Diagnostics VITROS Anti-SARS-CoV-2 IgG assay \| IgG | 93.6%, 100.0% (Used author-adjusted estimate) | 4.3% (1.5-10.0%) |
| Brant-Zawadzki (Hoag Memorial Hospital Presbyterian)[337] | 2020-07-15 to 2020-08-15 | California | Local | High | 2754 | Mean 43.06 (SD 12.05) | 72.1 | Non-probability | Health care workers and caregivers | 2.8% (2.2-3.4%) | Ortho Diagnostics \| Ortho Clinical Diagnostics VITROS Anti-SARS-CoV-2 IgG assay \| IgG | 93.6%, 100.0% (Used author-adjusted estimate) | 2.8% (2.2-3.4%) |
| Sabourin (University of Colorado)[338] | 2020-07-15 to 2020-08-15 | Colorado | Local | High | 264 | Median 38 (IQR 32-48) | 54.2 | Non-probability | Essential non-healthcare workers | 4.0% | Exsera BioLabs \| NR \| IgG | 85.0%, 98.6% (Test developer / manufacturer evaluation) | 3.1% (0.4-6.1%) |
| Coffman (University of Louiseville )[340] | 2020-07-01 to 2020-07-31 | Kentucky \| Louiseville | Local | Unclear | 1100 | NR | NR | Non-probability | Health care workers and caregivers | 2.2% | NR \| NR \| NR | No data | - |
| Azzi (Albert Einstein College of Medicine)[343] | 2020-05-03 to 2020-07-29 | New York \| New York City | Sublocal | Moderate | 912 | NR | NR | Non-probability | Patients seeking care for non-COVID-19 reasons | 16.7% | Abbott \| Abbott SARS-CoV-2 IgG antibody test \| IgG | 0.4%, 1.0% (FINDDx / MUHC independent evaluation) | - |
| Nicholson (Scripps Clinic)[344] | 2020-04-14 to 2020-07-28 | California \| San Diego | Local | High | 11993 | NR | NR | Non-probability | Health care workers and caregivers | 0.9% | Diazyme; Abbott Laboratories \| Diazyme DZ-Lite 2019-nCoV IgG (CLIA) Assay, Abbott Architect SARS- CoV-2 IgG assay \| IgG | 97.8%, 97.8% (Used Bastos SR/MA data; no sens, spec, or author adjustment available) | 0.0% (0.0-0.1%) |

| Article Author (Study Organization) | Sampling Dates (YMD) | Location | Geographic Scope | Overall Risk of Bias | Sample Size | Age | Female (%) | Sampling method | Sample frame | Uncorrected Seroprevalence (95% Confidence Interval) | Test Manufacturers \| Names \| Isotypes | Test Sens, Spec (Source) | Corrected Seroprevalence (95% Credible Interval) |
|---|---|---|---|---|---|---|---|---|---|---|---|---|---|
| Haizler-Cohen (Donald and Barbara Zucker School of Medicine)[347] | 2020-05-27 to 2020-07-24 | New York | Regional | High | 1671 | NR | 100.0 | Non-probability | Pregnant or parturient women | 16.1% | Roche \| Roche Elecsys® Anti-SARS-CoV-2 RUO Assay \| IgG | No data | - |
| Klawiter (Massachusetts General Hospital)[348] | 2020-05-29 to 2020-07-23 | Massachusetts \| Boston | Sublocal | High | 143 | NR | NR | Non-probability | Multiple populations | 6.3% | Massachusetts General Hospital \| MGH SARS-CoV-2 IgG ELISA \| IgG | 99.7%, 100.0% (Test developer / manufacturer evaluation) | 6.1% (1.0-10.9%) |
| Klawiter (Massachusetts General Hospital)[348] | 2020-05-29 to 2020-07-23 | Massachusetts \| Boston | Local | High | 227 | NR | NR | Non-probability | Patients seeking care for non-COVID-19 reasons | 3.5% | Massachusetts General Hospital \| MGH SARS-CoV-2 IgG ELISA \| IgG | 99.7%, 100.0% (Test developer / manufacturer evaluation) | 2.7% (0.0-5.3%) |
| Lopez (Harvard Medical School)[351] | 2020-07-15 to 2020-07-20 | Indiana \| St. John Township | Sublocal | Moderate | 753 | 18-35 (n=153); 36-50 (n=287); 51-65 (n=282); 66+ (n=30) | 84.3 | Non-probability | Essential non-healthcare workers | 1.7% (0.3-3.3%) | Abbott Alinity \| Abbott Alinity SARS-CoV-2 IgG antibody test \| IgG | 100.0%, 99.0% (Used author-adjusted estimate) | 1.7% (0.9-2.7%) |
| Reuben (District of Columbia Department of Health (DC Health))[354] | 2020-05-28 to 2020-07-15 | Washington D.C \| Washington | Regional | High | 310 | Mean 42.55 (SD 9.07) | 23.5 | Non-probability | Essential non-healthcare workers | 3.5% | DiaSorin Inc. \| DiaSorin LIAISON® XL assay \| IgG | 0.8%, 1.0% (FINDDx / MUHC independent evaluation) | - |
| Shah (Mayo Clinic)[356] | 2020-05-25 to 2020-07-09 | Minnesota \| Rochester | Sublocal | High | 320 | Mean 36.3 (SD 10.8) | 78.1 | Non-probability | Health care workers and caregivers | 0.6% | Ortho-Clinical Diagnostics; EUROIMMUN; Roche Diagnostics \| Euroimmun anti–SARS-CoV-2 IgG ELISA, Ortho-Clinical Diagnostics anti–SARS-CoV-2 IgG Antibody Immunoassay, Roche Diagnostics | No data | - |

| Article Author (Study Organization) | Sampling Dates (YMD) | Location | Geographic Scope | Overall Risk of Bias | Sample Size | Age | Female (%) | Sampling method | Sample frame | Uncorrected Seroprevalence (95% Confidence Interval) | Test Manufacturers \| Names \| Isotypes | Test Sens, Spec (Source) | Corrected Seroprevalence (95% Credible Interval) |
|---|---|---|---|---|---|---|---|---|---|---|---|---|---|
| | | | | | | | | | | | anti-SARS-CoV-2 Total Antibody Immunoassay \| IgG | | |
| Shah (Mayo Clinic)[356] | 2020-05-25 to 2020-07-09 | Minnesota \| Rochester | Sublocal | High | 248 | Mean 47.1 (SD 11.7) | 87.5 | Non-probability | Health care workers and caregivers | 0.0% | Ortho-Clinical Diagnostics; EUROIMMUN; Roche Diagnostics \| Euroimmun anti–SARS-CoV-2 IgG ELISA, Ortho-Clinical Diagnostics anti–SARS-CoV-2 IgG Antibody Immunoassay, Roche Diagnostics anti-SARS-CoV-2 Total Antibody Immunoassay \| IgG | No data | - |
| Mohr (University of Iowa)[357] | 2020-05-13 to 2020-07-08 | | National | Moderate | 1606 | <30 (n = 376); 31-40 (n = 622); 41-50 (n = 319); 51-60 (n = 222); >61 (n = 67) | 63.4 | Non-probability | Health care workers and caregivers | 1.7% | IL EUROIMMUN; Abbott Laboratories; Chicago \| Architect i2000 ELISA EUROIMMUN \| IgG | No data | - |
| Dimcheff (Veterans Affairs Ann Arbor Healthcare System)[358] | 2020-06-08 to 2020-07-08 | Michigan | Local | Moderate | 1476 | 18-30 (n=111, 7.5%); 31-40 (n=372, 25.2%); 41-50 (n=392, 26.6%); 51-60 (n=406, 27.5%); >60 (n=149, 10.1%); No response (n=46, 3.1%)/ | 64.0 | Non-probability | Health care workers and caregivers | 4.9% (3.8-6.1%) | Abbott \| ARCHITECT i2000SR analyzer \| IgG | 0.4%, 1.0% (FINDDx / MUHC independent evaluation) | - |

| Article Author (Study Organization) | Sampling Dates (YMD) | Location | Geographic Scope | Overall Risk of Bias | Sample Size | Age | Female (%) | Sampling method | Sample frame | Uncorrected Seroprevalence (95% Confidence Interval) | Test Manufacturers \| Names \| Isotypes | Test Sens, Spec (Source) | Corrected Seroprevalence (95% Credible Interval) |
|---|---|---|---|---|---|---|---|---|---|---|---|---|---|
| Freeman (University of Pittsburgh School of Medicine)[362] | 2020-03-15 to 2020-07-04 | Pennsylvania \| Pittsburgh | Sublocal | High | 485 | Mean 10.96 (Range 0.79-18.96) | 46.6 | Non-probability | Patients seeking care for non-COVID-19 reasons | 1.0% (0.3-2.4%) | Euroimmun; Siemens; Beckman Coulter \| Euroimmun Assay, Beckman Coulter SARS-CoV-2 IgG automated Emergency Use Authorization (EUA) assay, Siemens Centaur SARS-CoV-2 Total automated EUA assay \| IgG | 75.6%, 97.6% (Used Bastos SR/MA data; no sens, spec, or author adjustment available) | 0.3% (0.0-1.2%) |
| Barrett (Rutgers Robert Wood Johnson Medical School)[364] | 2020-04-28 to 2020-06-30 | New Jersey \| New Brunswick, Somerville | Sublocal | Moderate | 3907 | 20-39 (N=1703); 40-59 (n=1690); 60+ (n=472) | 73.6 | Non-probability | Health care workers and caregivers | 9.6% | Abbott Laboratories \| Abbott Architect SARS-CoV-2 assay \| IgG | 0.4%, 1.0% (FINDDx / MUHC independent evaluation) | - |
| Wilkins (Northwestern University )[365] | 2020-05-28 to 2020-06-30 | Illinois \| Chicago | Local | Moderate | 6510 | Mean 40.6 (SD 12) | 79.6 | Non-probability | Health care workers and caregivers | 5.3% (4.8-5.9%) | Abbott \| Abbott Architect SARS-CoV-2 IgG assay \| IgG | 0.4%, 1.0% (FINDDx / MUHC independent evaluation) | - |
| Nailescu (Indiana University )[366] | 2020-05-01 to 2020-06-30 | Indiana \| Indianapolis | Local | High | 31 | Median 12 (Range 2-21) | 32.3 | Non-probability | Patients seeking care for non-COVID-19 reasons | 3.2% | Mount Sinai medical center; Abnova; Eli Lilly and Co & IUSM \| (1) Abnova (#KA5826) (2)Indiana University School of Medicine (IUSM) in collaboration with Eli Lilly and Co., Indianapolis, IN, (LSN3832334); (3) Mount Sinai Medical Center's(MSM | 94.4%, 98.5% (Test developer / manufacturer evaluation) | 2.4% (0.0-10.2%) |

| Article Author (Study Organization) | Sampling Dates (YMD) | Location | Geographic Scope | Overall Risk of Bias | Sample Size | Age | Female (%) | Sampling method | Sample frame | Uncorrected Seroprevalence (95% Confidence Interval) | Test Manufacturers \| Names \| Isotypes | Test Sens, Spec (Source) | Corrected Seroprevalence (95% Credible Interval) |
|---|---|---|---|---|---|---|---|---|---|---|---|---|---|
| | | | | | | | | | | | C) 2-Step ELISA \| IgG, IgM | | |
| Rivas (Cedars-Sinai Medical Center)[369] | 2020-05-11 to 2020-06-28 | California \| Los Angeles | Local | Moderate | 6201 | Mean 41.46 (SD 12.01) | 67.9 | Non-probability | Health care workers and caregivers | 3.5% | Abbott \| Architect SARS-CoV-2 IgG chemiluminescent microparticle immunoassay \| IgG | 0.4%, 1.0% (FINDDx / MUHC independent evaluation) | - |
| Patel (Vanderbilt University Medical Center)[370] | 2020-06-02 to 2020-06-27 | Tennessee \| Nashville | Sublocal | High | 230 | NR | NR | Non-probability | Health care workers and caregivers | 3.5% | CDC \| NR \| NR | 96.0%, 99.0% (Test developer / manufacturer evaluation) | 2.4% (0.0-5.5%) |
| Baker (Emory University Hospital)[371] | 2020-04-19 to 2020-06-26 | Atlanta | Local | Moderate | 10275 | <30 (n=1581); 30-39 (n=2953); 40-49 (n=2118); 50-59 (n=1805); >=60 (n=1153) | 72.6 | Non-probability | Health care workers and caregivers | 5.7% (5.2-6.1%) | Emory University \| In house ELISA \| IgG | 97.5%, 98.0% (Test developer / manufacturer evaluation) | 3.0% (0.0-5.3%) |
| McBride (Memorial Sloan Kettering Cancer Center)[372] | 2020-05-07 to 2020-06-25 | New York \| New York City | Sublocal | High | 952 | NR | NR | Non-probability | Patients seeking care for non-COVID-19 reasons | 5.5% | Abbott \| Abbott Architect SARS-CoV-2 IgG assay \| IgG | 0.4%, 1.0% (FINDDx / MUHC independent evaluation) | - |
| Moscola (Northwell Health COVID-19 Research Consortium)[373] | 2020-04-20 to 2020-06-23 | New York \| New York City | Local | High | 40329 | Median 42 (IQR 31.5-54.5) | 73.7 | Non-probability | Health care workers and caregivers | 13.7% | AnshLabs; Roche; DiaSorin; Abbott Laboratories; Ortho Clinical Diagnostics; Euroimmun \| Euroimmun SARS-CoV-2 ELISA, AnshLabs SARS-CoV-2 IgG ELISA, Abbott | No data | - |

| Article Author (Study Organization) | Sampling Dates (YMD) | Location | Geographic Scope | Overall Risk of Bias | Sample Size | Age | Female (%) | Sampling method | Sample frame | Uncorrected Seroprevalence (95% Confidence Interval) | Test Manufacturers \| Names \| Isotypes | Test Sens, Spec (Source) | Corrected Seroprevalence (95% Credible Interval) |
|---|---|---|---|---|---|---|---|---|---|---|---|---|---|
| | | | | | | | | | | | Architect SARS-CoV-2 IgG, Ortho Clinical Diagnostics VITROS Anti-SARSCoV-2 IgG, Ortho Clinical Diagnostics VITROS Immunodiagnostic Products Anti-SARS-CoV-2 Total Reagent Pack and Calibrator, DiaSorin LIAISON SARS-CoV-2 S1/S2 IgG, Roche Elecsys Anti-SARS-CoV-2 \| IgG | | |
| Self (CDC COVID-19 Response Team)[376] | 2020-04-03 to 2020-06-19 | Colorado, Minnesota, Oregon, Utah, Washington, Tennessee, New York, California, North Carolina, Massachusetts, Ohio, Maryland \| Winston-Salem, Murray, Portland, Baltimore, Bronx, Springfield, Los Angeles, Aurora, Nashville, Seattle, Columbus, Boston, Minneapolis | National | Moderate | 3248 | Median 36 (NR) | 65.5 | Non-probability | Health care workers and caregivers | 6.0% | CDC \| NR \| NR | 96.0%, 99.0% (Test developer / manufacturer evaluation) | 4.5% (0.5-6.9%) |

| Article Author (Study Organization) | Sampling Dates (YMD) | Location | Geographic Scope | Overall Risk of Bias | Sample Size | Age | Female (%) | Sampling method | Sample frame | Uncorrected Seroprevalence (95% Confidence Interval) | Test Manufacturers \| Names \| Isotypes | Test Sens, Spec (Source) | Corrected Seroprevalence (95% Credible Interval) |
|---|---|---|---|---|---|---|---|---|---|---|---|---|---|
| Rebeiro (Vanderbilt University)[380] | 2020-05-15 to 2020-06-15 | Tennessee | Regional | High | 11787 | Median 40.3 (IgG Negative); 32.4 (IgG Positive) (IQR 30.9-52.4 (IgG Negative); 26.5-43.4 (IgG Positive)) | NR | Non-probability | Health care workers and caregivers | 1.0% | Abbott \| Abbott ARCHITECT SARS-CoV-2 IgG Assay \| IgG | 0.4%, 1.0% (FINDDx / MUHC independent evaluation) | - |
| Akinbami (Region 2 South Healthcare Coalition)[382] | 2020-05-18 to 2020-06-13 | Michigan \| Detroit | Local | Moderate | 16397 | Mean 42.1 (SD 12.2) | 68.6 | Non-probability | Health care workers and caregivers | 6.9% (6.5-7.3%) | Ortho Clinical Diagnostics \| VITROS Immunodiagnostic Products Anti- SARS-CoV-2 IgG Test \| IgG | 0.8%, 1.0% (FINDDx / MUHC independent evaluation) | - |
| Dora (Veterans Affairs Healthcare System)[383] | 2020-06-05 to 2020-06-12 | California \| Los Angeles | Sublocal | High | 150 | Median 75 (PCR positive); 74 (PCR negative) (NR) | NR | Non-probability | Assisted living and long-term care facilities | 17.3% | DiaSorin \| DiaSorin Liaison \| IgG | 0.8%, 1.0% (FINDDx / MUHC independent evaluation) | - |
| Yogo (Sharp Healthcare)[386] | 2020-05-20 to 2020-06-08 | California \| San Diego | Local | High | 1770 | 18-35 (n=802); 36-45 (n=460); 45-55 (n=323); 56-65 (n=146); >65 (n=39) | 75.5 | Non-probability | Health care workers and caregivers | 2.2% | Roche Diagnostics \| Elecsys Anti-SARS-CoV-2 \| NR | 0.9%, 1.0% (FINDDx / MUHC independent evaluation) | - |
| Hogan (Stanford University)[387] | 2020-04-20 to 2020-06-08 | California \| San Francisco | Local | High | 12418 | Median 39.5 (IQR 32.4-50.3) | 70.7 | Non-probability | Health care workers and caregivers | 0.9% (0.7-1.1%) | Stanford Health Care \| SHC Laboratory - Developed ELISA \| IgG | 75.6%, 97.6% (Used Bastos SR/MA data; no sens, spec, or author adjustment available) | 0.1% (0.0-0.2%) |
| Flannery (University of Pennsylvania)[389] | 2020-04-04 to 2020-06-03 | Pennsylvania \| Philadelphia | Local | Moderate | 1293 | Median 31 (IQR 27-35) | 100.0 | Non-probability | Pregnant or parturient women | 6.2% (4.9-8.0%) | Author-designed \| N/A - Author-designed \| IgG, IgM | 100.0%, 98.9% (Author-reported independent evaluation) | 5.3% (3.6-7.2%) |

| Article Author (Study Organization) | Sampling Dates (YMD) | Location | Geographic Scope | Overall Risk of Bias | Sample Size | Age | Female (%) | Sampling method | Sample frame | Uncorrected Seroprevalence (95% Confidence Interval) | Test Manufacturers \| Names \| Isotypes | Test Sens, Spec (Source) | Corrected Seroprevalence (95% Credible Interval) |
|---|---|---|---|---|---|---|---|---|---|---|---|---|---|
| Coffman (University of Louiseville )[340] | 2020-04-01 to 2020-05-31 | Kentucky \| Louiseville | Local | Unclear | 1372 | NR | NR | Non-probability | Health care workers and caregivers | 1.0% | NR \| NR \| NR | No data | - |
| Addetia (University of Washington)[390] | 2020-05-01 to 2020-05-31 | Washington \| Seattle | Sublocal | High | 120 | NR | 7.5 | Probability | Essential non-healthcare workers | 5.0% | Abbott \| Abbott Architect SARS-CoV-2 IgG \| IgG | 0.4%, 1.0% (FINDDx / MUHC independent evaluation) | - |
| Gray (Truckee Meadows Fire Protection District)[391] | 2020-05-01 to 2020-05-31 | Nevada | Sublocal | High | 132 | NR | NR | Non-probability | Essential non-healthcare workers | 14.0% | NR \| NR \| NR | No data | - |
| Brunner (Bassett Medical Center)[392] | 2020-05-04 to 2020-05-29 | New York \| Upstate New York | Local | High | 764 | Mean 41.8 (seropositive); 43.7 (seronegative) (SD 16.0 (seropositive); 13.7 (seronegative)) | 56.9 | Non-probability | Health care workers and caregivers | 2.0% | Abbott Laboratories \| SARS-CoV-2 IgG Abbott Architect assay \| IgG | 0.4%, 1.0% (FINDDx / MUHC independent evaluation) | - |
| Brunner (Bassett Medical Center)[392] | 2020-05-04 to 2020-05-29 | New York \| Upstate New York | Local | High | 762 | Mean 41.8 (igg+); 43.7 (igg-) (SD 16 (igg+); 13.7(igg-)) | NR | Non-probability | Health care workers and caregivers | 4.5% | Linden; Ortho-Clinical Diagnostics; NJ \| VITROS Anti-SARS-CoV-2 IgG Test \| IgG | 0.8%, 1.0% (FINDDx / MUHC independent evaluation) | - |
| Sims (Oakland University)[394] | 2020-04-13 to 2020-05-28 | Michigan \| Trenton, Wayne, Grosse Pointe, Farmington Hills, Dearborn, Taylor, Canton, Royal Oak, Troy | Sublocal | Moderate | 20614 | Mean 43.1 (SD 13) | 76.3 | Non-probability | Health care workers and caregivers | 8.1% (6.8-8.9%) | EUROIMMUN Lübeck \| EUROIMMUN SARS-CoV-2 IgG assay \| IgG | 98.1%, 99.4% (Used author-adjusted estimate) | 8.1% (7.7-8.5%) |
| Stavert (Cambridge Health Alliance)[399] | 2020-04-13 to 2020-05-24 | Massachusetts \| Cambridge | Sublocal | High | 24 | 11-64 | 45.8 | Non-probability | Patients seeking care for non-COVID-19 reasons | 0.0% | DiaSorin \| DiaSorin Liaison \| IgG | 0.8%, 1.0% (FINDDx / MUHC independent evaluation) | - |

| Article Author (Study Organization) | Sampling Dates (YMD) | Location | Geographic Scope | Overall Risk of Bias | Sample Size | Age | Female (%) | Sampling method | Sample frame | Uncorrected Seroprevalence (95% Confidence Interval) | Test Manufacturers \| Names \| Isotypes | Test Sens, Spec (Source) | Corrected Seroprevalence (95% Credible Interval) |
|---|---|---|---|---|---|---|---|---|---|---|---|---|---|
| Ahmad (The Lundquist Institute )[400] | 2020-04-21 to 2020-05-22 | California | Local | High | 244 | Mean 51.1 (NR) | 50.0 | Non-probability | Multiple populations | 20.9% | Hangzhou Testsea Biotechnology Co. \| One Step SARS-CoV2 (COVID-19) IgG/IgM Rapid Test \| IgG, IgM | 0.8%, 0.9% (FINDDx / MUHC independent evaluation) | - |
| Jackson (Michigan Department of Corrections )[401] | 2020-05-14 to 2020-05-22 | Michigan | Sublocal | Unclear | 1248 | NR | NR | Non-probability | Persons who are incarcerated | 92.0% | NR \| NR \| NR | No data | - |
| Jackson (Michigan Department of Corrections )[401] | 2020-05-14 to 2020-05-22 | Michigan \| Lenox Township | Sublocal | Unclear | 1201 | NR | NR | Non-probability | Persons who are incarcerated | 65.6% | NR \| NR \| NR | No data | - |
| Jackson (Michigan Department of Corrections )[401] | 2020-05-14 to 2020-05-22 | Michigan \| Detroit | Sublocal | Unclear | 57 | NR | NR | Non-probability | Persons who are incarcerated | 35.0% | NR \| NR \| NR | No data | - |
| Jackson (Michigan Department of Corrections )[401] | 2020-05-14 to 2020-05-22 | Michigan | Sublocal | Unclear | 867 | NR | NR | Non-probability | Persons who are incarcerated | 12.2% | NR \| NR \| NR | No data | - |
| Staletovich (Coral Springs City Government)[402] | 2020-05-17 to 2020-05-22 | Florida \| Coral Springs | Local | Unclear | 700 | NR | NR | Non-probability | Essential non-healthcare workers | 0.0% | Cellex \| NR \| NR | No data | - |
| Leidner (Chiles Research Institute)[403] | 2020-04-08 to 2020-05-22 | Oregon \| Seaside, Newberg, Portland, Oregon City, Hood River, Medford, Milwaukie | Regional | High | 10019 | Median 42 (Range 18-82) | 75.7 | Non-probability | Health care workers and caregivers | 2.5% | Author-designed \| N/A - Author-designed \| IgG | 80.0%, 100.0% (Test developer / manufacturer evaluation) | 3.2% (2.5-4.3%) |
| Pritchett (University of Utah)[404] | 2020-04-18 to 2020-05-21 | Utah | Sublocal | High | 39 | Median 45 (Range 23 to 76) | 89.7 | Non-probability | Patients seeking care for non- | 2.6% | in house - ARUP \| NR \| IgG | No data | - |

| Article Author (Study Organization) | Sampling Dates (YMD) | Location | Geographic Scope | Overall Risk of Bias | Sample Size | Age | Female (%) | Sampling method | Sample frame | Uncorrected Seroprevalence (95% Confidence Interval) | Test Manufacturers \| Names \| Isotypes | Test Sens, Spec (Source) | Corrected Seroprevalence (95% Credible Interval) |
|---|---|---|---|---|---|---|---|---|---|---|---|---|---|
| | | | | | | | | | COVID-19 reasons | | | | |
| Shukla (Vincere Cancer Center)[405] | 2020-04-24 to 2020-05-21 | Arizona \| Tempe, Peoria, Phoenix, Glendale, Chandler, Surprise | Regional | Moderate | 3326 | Median 41.3 (IQR 9.6) | 16.4 | Non-probability | Essential non-healthcare workers | 1.5% | Ray Biotech \| RayBiotech Coronavirus (COVID-19) IgG Rapid Test Kit \| IgG | 0.1%, 0.8% (FINDDx / MUHC independent evaluation) | - |
| Rosser (Stanford Health Care)[406] | 2020-04-20 to 2020-05-20 | California \| San Francisco Bay Area | Local | High | 10449 | Mean 40.9 (SD 11.6) | 65.0 | Non-probability | Health care workers and caregivers | 1.3% | Stanford Clinical Laboratory \| anti–SARS-CoV-2 RBD enzyme-linked immunosorbent assay \| IgG | 87.0%, 99.8% (Test developer / manufacturer evaluation) | 0.6% (0.0-1.3%) |
| Geraci (MUSC Health-Lancaster)[409] | 2020-03-16 to 2020-05-20 | South Carolina \| Lancaster | Sublocal | High | 230 | NR | NR | Non-probability | Health care workers and caregivers | 2.2% | Abbott \| NR \| NR | No data | - |
| Tarabichi (Case Western Reserve University)[410] | 2020-04-20 to 2020-05-19 | Ohio \| Cleveland | Local | High | 296 | Mean 44.1 (NR) | 14.2 | Non-probability | Essential non-healthcare workers | 5.4% (3.1-8.6%) | Epitope Diagnostics; Inc \| EDI™ Novel Coronavirus COVID-19 ELISA Kits \| IgG, IgM | 0.8%, 0.9% (FINDDx / MUHC independent evaluation) | - |
| Mughal (Monmouth Medical Centre)[411] | 2020-05-14 to 2020-05-19 | New Jersey \| Long Branch | Sublocal | High | 121 | Mean 39.2 (IQR 28-48.5) | 82.6 | Non-probability | Health care workers and caregivers | 0.8% | SD Biosensor \| Standard Q COVID-19 IgM/IgG Duo rapid immunochromatography test kit \| IgG | 96.2%, 96.6% (Test developer / manufacturer evaluation) | 0.9% (0.0-3.3%) |
| Morris (Emory University)[413] | 2020-04-16 to 2020-05-18 | Georgia \| Atlanta | Sublocal | Unclear | 300 | Mean 41-50 (NR) | 83.0 | Non-probability | Health care workers and caregivers | 4.7% | NR \| NR \| IgG | No data | - |
| Parker-Magyar (Summit Medical Center)[418] | 2020-05-01 to 2020-05-14 | New Jersey \| Long Hill Township | Sublocal | High | 70 | NR | NR | Non-probability | Essential non-healthcare workers | 4.3% | NR \| NR \| NR | No data | - |

| Article Author (Study Organization) | Sampling Dates (YMD) | Location | Geographic Scope | Overall Risk of Bias | Sample Size | Age | Female (%) | Sampling method | Sample frame | Uncorrected Seroprevalence (95% Confidence Interval) | Test Manufacturers \| Names \| Isotypes | Test Sens, Spec (Source) | Corrected Seroprevalence (95% Credible Interval) |
|---|---|---|---|---|---|---|---|---|---|---|---|---|---|
| Iwuji (Texas Tech University)[419] | 2020-05-12 to 2020-05-13 | Texas \| Lubbock | Local | High | 683 | 18-76 | NR | Non-probability | Essential non-healthcare workers | 0.7% | Abbott \| Abbott SARS-CoV-2 IgG \| IgG | 0.4%, 1.0% (FINDDx / MUHC independent evaluation) | - |
| Tilley (University of Southern California)[422] | 2020-04-29 to 2020-05-08 | California \| Los Angeles | Sublocal | Moderate | 790 | NR | 48.2 | Non-probability | Students and Daycares | 4.0% (3.0-5.1%) | EUROIMMUN \| EUROIMMUN Anti-SARS-CoV-2 ELISA (IgG) \| IgG | 90.0%, 100.0% (Used author-adjusted estimate) | 4.0% (2.7-5.5%) |
| Hunter ( Indiana University)[423] | 2020-04-29 to 2020-05-08 | Indiana \| Indianapolis | Regional | High | 734 | Mean 42.8 (NR) | 70.0 | Non-probability | Health care workers and caregivers | 1.6% | Abbott \| Abbott Architect SARS-CoV-2 IgG assay \| IgG | 0.4%, 1.0% (FINDDx / MUHC independent evaluation) | - |
| Racine-Brzostek (Weill Cornell Medicine)[425] | 2020-04-17 to 2020-05-07 | New York \| New York City | Local | Moderate | 2274 | Median 37 (IQR 31-48) | 63.3 | Non-probability | Health care workers and caregivers | 35.4% | ET Healthcare \| SARS-CoV-2 cyclic enhanced fluorescence assay (CEFA) \| IgG, IgM | 0.6%, 1.0% (FINDDx / MUHC independent evaluation) | - |
| Regan (Fellowship Village)[426] | 2020-04-15 to 2020-05-06 | New Jersey \| Basking Ridge | Sublocal | Unclear | 371 | NR | NR | Probability | Assisted living and long-term care facilities | 34.8% | NR \| NR \| IgG, IgM | No data | - |
| Regan (Fellowship Village)[426] | 2020-04-15 to 2020-05-06 | New Jersey \| Basking Ridge | Sublocal | Unclear | 305 | NR | NR | Probability | Assisted living and long-term care facilities | 23.6% | NR \| NR \| IgG, IgM | No data | - |
| Sydney (Jacobi Medical Center)[427] | 2020-04-28 to 2020-05-04 | New York \| New York City | Sublocal | High | 1700 | NR | NR | Non-probability | Health care workers and caregivers | 19.2% | Abbott \| Abbott Architect SARS-CoV-2 IgG assay \| IgG | 0.4%, 1.0% (FINDDx / MUHC independent evaluation) | - |
| Reagan (Medical University South Carolina)[431] | 2020-04-27 to 2020-05-02 | South Carolina \| Charleston | Sublocal | High | 920 | NR | NR | Non-probability | Multiple populations | 2.0% | Abbott \| Not possible to determine \| NR | No data | - |

| Article Author (Study Organization) | Sampling Dates (YMD) | Location | Geographic Scope | Overall Risk of Bias | Sample Size | Age | Female (%) | Sampling method | Sample frame | Uncorrected Seroprevalence (95% Confidence Interval) | Test Manufacturers \| Names \| Isotypes | Test Sens, Spec (Source) | Corrected Seroprevalence (95% Credible Interval) |
|---|---|---|---|---|---|---|---|---|---|---|---|---|---|
| Venugopal (NYC Health + Hospitals/Lincoln)[432] | 2020-03-01 to 2020-05-01 | New York \| New York City | Local | Moderate | 500 | 20-39 (n=230), 40-59 (n=196), 60+ (n=52) | 65.8 | Non-probability | Health care workers and caregivers | 27.0% | Abbott Laboratories \| Abbott Architect SARS-CoV-2 IgG Assay \| IgG | 0.4%, 1.0% (FINDDx / MUHC independent evaluation) | - |
| Jeremias (St Francis Hospital)[433] | 2020-03-01 to 2020-04-30 | New York \| New York City | Sublocal | High | 1699 | Mean 42.8 (SD 13.8) | 74.1 | Non-probability | Health care workers and caregivers | 9.8% | EUROIMMUN ; Abbott \| Euroimmun Anti-SARS-CoV-2 ELISA IgG, SARS-CoV-2 IgG \| IgG | No data | - |
| Cohen (DaVita Clinical Research)[434] | 2020-04-23 to 2020-04-30 | Florida \| Miami | Local | High | 677 | Mean 65.7 (SD +/- 14.5 years) | 41.1 | Non-probability | Patients seeking care for non-COVID-19 reasons | 5.6% (4.0-7.6%) | Diazyme Laboratories Inc \| DZ-Lite SARS-CoV-2 IgG IgM CLIA Kit \| IgG, IgM | 57.1%, 85.0% (Author-reported independent evaluation) | 2.8% (0.0-9.6%) |
| Jeong (Columbia University Medical Center)[435] | 2020-02-01 to 2020-04-30 | New York \| New York City | Sublocal | High | 50 | Median 35 (IQR 31-49) | 42.0 | Non-probability | Health care workers and caregivers | 46.0% | Abbott Laboratories \| SARS-CoV-2 immunoglobulin G antibody test \| IgG | 0.4%, 1.0% (FINDDx / MUHC independent evaluation) | 53.3% (39.9-66.9%) |
| Lewis (Centers for Disease Control )[440] | 2020-03-22 to 2020-04-25 | Utah, Wisconsin \| Milwaukee, Salt Lake City | Local | High | 178 | Median 22 (Range <1-76) | 53.9 | Non-probability | Contacts of COVID patients | 25.8% | CDC \| CDC-developed SARS-CoV-2 ELISA \| NR | 75.6%, 97.6% (Used Bastos SR/MA data; no sens, spec, or author adjustment available) | 31.5% (22.7-40.1%) |
| Payne (US Navy)[442] | 2020-04-20 to 2020-04-24 | Guam | Sublocal | High | 382 | Median 30 (IQR 24-35) | 24.3 | Non-probability | Essential non-healthcare workers | 59.7% | CDC \| CDC Pan-immunoglobulin ELISA \| IgG, IgA, IgM | 96.0%, 99.3% (Test developer / manufacturer evaluation) | 64.6% (56.4-75.8%) |
| Kiefer (Ohio State University College of Medicine)[445] | 2020-04-23 to 2020-04-23 | Ohio \| Columbus | Sublocal | High | 110 | Median 34 (IQR 28.8-45.0) | 100.0 | Non-probability | Health care workers and caregivers | 20.0% | Epitope Diagnostics \| Novel Coronavirus COVID-19 IgG ELISA Kit \| IgG, IgM | 0.8%, 1.0% (FINDDx / MUHC independent evaluation) | - |

| Article Author (Study Organization) | Sampling Dates (YMD) | Location | Geographic Scope | Overall Risk of Bias | Sample Size | Age | Female (%) | Sampling method | Sample frame | Uncorrected Seroprevalence (95% Confidence Interval) | Test Manufacturers \| Names \| Isotypes | Test Sens, Spec (Source) | Corrected Seroprevalence (95% Credible Interval) |
|---|---|---|---|---|---|---|---|---|---|---|---|---|---|
| Morcuende (Columbia University Irving Medical Center)[447] | 2020-03-01 to 2020-04-21 | New York \| New York City | Sublocal | High | 91 | NR | 42.9 | Non-probability | Health care workers and caregivers | 12.1% | "Zhejiang Orient Gene Biotech Co.; LTD" \| COVID-19 IgG / IgM Rapid Test Cassette \| IgG, IgM | 93.1%, 99.2% (Test developer / manufacturer evaluation) | 12.1% (3.7-21.2%) |
| McCormack (Stanford University)[448] | 2020-04-08 to 2020-04-21 | | National | Unclear | 5603 | NR | NR | Non-probability | Non-essential workers and unemployed persons | 0.7% (0.3-1.1%) | NR \| NR \| NR | No data | - |
| Stock (Albert Einstein College of Medicine)[450] | 2020-04-04 to 2020-04-20 | New York \| New York City | Local | High | 98 | Mean 37.6 (SD 10.6) | 50.0 | Non-probability | Health care workers and caregivers | 15.3% | Epitope Diagnostics \| anti-n IgG ELISA \| IgG | 0.8%, 1.0% (FINDDx / MUHC independent evaluation) | - |
| Madsen (University of Utah)[451] | 2020-04-13 to 2020-04-19 | Utah \| Salt Lake City | Sublocal | High | 270 | NR | NR | Non-probability | Health care workers and caregivers | 5.9% | EUROIMMUN \| Eurommun Anti-SARS-CoV-2-IgG \| IgG | 0.8%, 1.0% (FINDDx / MUHC independent evaluation) | - |
| Caban-Martinez (University of Miami)[452] | 2020-04-16 to 2020-04-17 | Florida | Sublocal | High | 203 | 21-30 (n=33); 31-40 (n=51); 41-50 (n=67); 51+ (n=52) | 6.4 | Non-probability | Essential non-healthcare workers | 8.9% | BioMedomics \| Rapid immunoglobulin (Ig)M-IgG combined point-of-care (POC) lateral flow immunoassay \| IgG, IgM | 0.6%, 0.9% (FINDDx / MUHC independent evaluation) | - |
| Goldberg (Harvard Medical School)[456] | 2020-04-01 to 2020-04-06 | Massachusetts | Sublocal | High | 140 | Mean 83 (resident age); 45 (staff) (Range 54-102 (resident); NR (staff)) | NR | Non-probability | Assisted living and long-term care facilities | 10.7% | BioMedomics \| COVID-19 IgM-IgG Dual Antibody Rapid Test \| IgG, IgM | 0.6%, 0.9% (FINDDx / MUHC independent evaluation) | - |
| Mansour (Icahn School of Medicine at | 2020-03-24 to 2020-04-04 | New York \| New York City | Local | High | 285 | Mean 38.36 (NR) | 33.3 | Non-probability | Health care workers and caregivers | 32.6% | NR \| NR \| IgG | 75.6%, 97.6% (Used Bastos SR/MA data; no sens, spec, | 40.9% (33.2-48.6%) |

| Article Author (Study Organization) | Sampling Dates (YMD) | Location | Geographic Scope | Overall Risk of Bias | Sample Size | Age | Female (%) | Sampling method | Sample frame | Uncorrected Seroprevalence (95% Confidence Interval) | Test Manufacturers \| Names \| Isotypes | Test Sens, Spec (Source) | Corrected Seroprevalence (95% Credible Interval) |
|---|---|---|---|---|---|---|---|---|---|---|---|---|---|
| Mount Sinai)[458] | | | | | | | | | | | | or author adjustment available) | |
| McPherson (US Centers for Disease Control and Prevention)[460] | 2020-03-03 to 2020-03-06 | Illinois | Sublocal | High | 59 | Median 39 (IQR 32-54) | 79.7 | Non-probability | Contacts of COVID patients | 0.0% | CDC \| SARS-CoV-2 spike enzyme-linked immunosorbent assay \| IgG, IgA, IgM | 96.0%, 99.0% (Test developer / manufacturer evaluation) | 1.3% (0.0-5.5%) |
| Chu (US Centers for Disease Control and Prevention)[461] | 2020-01-20 to 2020-02-03 | Washington | Regional | High | 32 | Median 45 (Range 0-78) | NR | Non-probability | Contacts of COVID patients | 0.0% | Author-designed \| CDC SARS-CoV-2 ELISA \| NR | 96.0%, 99.0% (Test developer / manufacturer evaluation) | 2.3% (0.0-9.6%) |
| **Latin America and Caribbean** | | | | | | | | | | | | | |
| **Brazil** | | | | | | | | | | | | | |
| Santana (Clínica de Reumatologia)[464] | 2020-03-15 to 2020-08-15 | State of Sao Paulo \| São Paulo | Local | High | 100 | Mean 46.5 (SD 14.2) | 85.0 | Non-probability | Patients seeking care for non-COVID-19 reasons | 21.0% | Snibe Diagnostics \| MAGLUMI anti-SARS-CoV-2 IgG/IgM \| IgG, IgM | 97.8%, 97.8% (Used Bastos SR/MA data; no sens, spec, or author adjustment available) | 20.2% (12.2-28.4%) |
| Silva (Adolfo Lutz Central Institute)[467] | 2020-06-05 to 2020-07-31 | State of Sao Paulo \| Sao Paulo | Sublocal | High | 406 | Median 50 (IQR 40-57) | 72.9 | Non-probability | Multiple populations | 8.6% | Ltd.; Switzerland; China; Roche Diagnostics; Rotkreuz; Guangzhou Wondfo Biotech Co. \| SARS-CoV-2 Wondfo; Elecys Anti-SARS-CoV-2 \| IgG, IgM | No data | - |
| Melo (Federal University of Sergipe)[469] | 2020-06-01 to 2020-06-30 | Sergipe | Regional | High | 471 | NR | NR | Non-probability | Health care workers and caregivers | 13.6% | Boditech Med Inc. \| Ichroma COVID-19 Ab Test \| IgG | 95.8%, 97.0% (Author-reported independent evaluation) | 10.7% (5.8-15.5%) |

| Article Author (Study Organization) | Sampling Dates (YMD) | Location | Geographic Scope | Overall Risk of Bias | Sample Size | Age | Female (%) | Sampling method | Sample frame | Uncorrected Seroprevalence (95% Confidence Interval) | Test Manufacturers \| Names \| Isotypes | Test Sens, Spec (Source) | Corrected Seroprevalence (95% Credible Interval) |
|---|---|---|---|---|---|---|---|---|---|---|---|---|---|
| Costa (Faculdade De Medicina da Universidade)[475] | 2020-05-14 to 2020-05-28 | São Paulo State \| São Paulo | Sublocal | Moderate | 4987 | <30 (n=1512), 31-40 (n=1539). 51-60 (n=636), 61+ (n=150) | 72.9 | Non-probability | Health care workers and caregivers | 14.1% (13.1-15.0%) | Wondfo-China \| Rapid Chromatograph ic Immunoassay \| IgG, IgM | 0.7%, 1.0% (FINDDx / MUHC independent evaluation) | - |
| Silva (Centro de Terapia Oncológica)[482] | 2020-04-09 to 2020-04-29 | Rio de Janeiro \| Petrópolis | Sublocal | High | 61 | NR | NR | Non-probability | Health care workers and caregivers | 4.9% | NR \| NR \| IgG, IgM | 66.0%, 96.6% (Used Bastos SR/MA data; no sens, spec, or author adjustment available) | 3.4% (0.0-11.5%) |
| **Columbia** | | | | | | | | | | | | | |
| Ariza (Pontificia Universidad Javeriana)[486] | 2020-06-25 to 2020-08-10 | Capital District Cundinamar ca \| Bogotá | Sublocal | High | 351 | Median 31.5 (Seronegativ e); 29.4 (Seropositive ) (IQR 27.5-38.6 (Seronegativ e); 26.9-37 (Seropositive )) | 58.7 | Non-probability | Health care workers and caregivers | 8.3% (5.8-11.6%) | Columbia SD Biosensor; Abbott \| Abbott Architect SARS-CoV-2 IgG assay \| IgG | 0.4%, 1.0% (FINDDx / MUHC independent evaluation) | - |
| Idrovo (Universidad Industrial de Santander)[487] | 2020-04-17 to 2020-07-14 | | National | High | 4234 | NR | NR | Non-probability | Non-essential workers and unemployed persons | 4.0% | NR \| NR \| IgG, IgM | 66.0%, 96.6% (Used Bastos SR/MA data; no sens, spec, or author adjustment available) | 1.0% (0.0-2.1%) |
| **Cuba** | | | | | | | | | | | | | |
| Giraldoni (University of Medical Sciences of Cienfuegos)[488] | 2020-03-23 to 2020-04-06 | Cienfuegos \| Aguada de Pasajeros | Local | Unclear | 200 | NR | NR | Non-probability | Contacts of COVID patients | 0.0% | NR \| NR \| IgG, IgM | No data | **215** |
| **Mexico** | | | | | | | | | | | | | |
| Cruz-Arenas (Instituto Nacional de | 2020-08-10 to 2020-09-09 | Mexico City | Sublocal | Moderate | 300 | < 30 (n=41) ; 30-44 (n = 124) ; 45-59 | 65.0 | Probability | Health care workers and caregivers | 11.0% | ; Hangzhou Biotest Biotech Co. Ltd. \| | 1.0%, 1.0% (FINDDx / MUHC | - |

| Article Author (Study Organization) | Sampling Dates (YMD) | Location | Geographic Scope | Overall Risk of Bias | Sample Size | Age | Female (%) | Sampling method | Sample frame | Uncorrected Seroprevalence (95% Confidence Interval) | Test Manufacturers \| Names \| Isotypes | Test Sens, Spec (Source) | Corrected Seroprevalence (95% Credible Interval) |
|---|---|---|---|---|---|---|---|---|---|---|---|---|---|
| Rehabilitacion)[494] | | | | | | ( n = 114) ; 60+ (n = 7) | | | | | COVID-19 IgG/IGm Rapid Test Cassette \| IgG, IgM | independent evaluation) | |
| Díaz-Salazar (Universidad Autonoma de Nuevo Leon)[497] | 2020-07-09 to 2020-07-23 | Nuevo Leon \| Guadalupe | Local | Moderate | 3268 | Median 40 (IQR 3-49) | 38.6 | Non-probability | Multiple populations | 5.7% (4.9-6.6%) | Roche elecsys \| Elecsys Anti-SARS-CoV-2 Roche COBAS e801 \| IgG, IgM | 99.5%, 99.8% (Used author-adjusted estimate) | 5.7% (4.9-6.5%) |
| **Panama** | | | | | | | | | | | | | |
| Villarreal (Instituto de Investigaciones Científicas y Servicios de Alta Tecnología (INDICASAT-AIP))[498] | 2020-04-30 to 2020-07-07 | Colon, Panama | National | High | 351 | Mean 39.5 (SD 11.5) | 67.8 | Non-probability | Health care workers and caregivers | 12.8% (8.6-15.4%) | N/A - Author Designed \| Chinese Academy of Science Test (CAST) \| IgG, IgM | 87.0%, 98.9% (Test developer / manufacturer evaluation) | 12.8% (5.8-18.6%) |
| **Peru** | | | | | | | | | | | | | |
| Álvarez-Antonio (Universidad de Ingeniería y Tecnología)[500] | 2020-07-15 to 2020-08-15 | Iquitos | Local | Low | 1337 | Mean 29.2 (Range 3 months - 89) | NR | Probability | Multiple populations | 68.1% (65.6-70.6%) | China); Zhejiang Orient Gene (Biotech Co LTD \| COVID-19 IgG/IgM Rapid Test Cassette \| IgG, IgM | 95.8%, 100.0% (Used author-adjusted estimate) | 68.1% (65.6-70.6%) |
| **North Africa and Middle East** | | | | | | | | | | | | | |
| **Egypt** | | | | | | | | | | | | | |
| Kassem (Cairo University)[503] | 2020-06-01 to 2020-06-14 | Cairo | Local | High | 74 | Median 32 (Range 23-48) | 59.5 | Non-probability | Health care workers and caregivers | 12.2% | Artron Laboratories \| COVID-19 IgM/IgG antibody rapid diagnostic test \| IgG, IgM | 0.4%, 1.0% (FINDDx / MUHC independent evaluation) | - |
| Mostafa (Ain Shams University)[504] | 2020-04-22 to 2020-05-14 | Cairo Governorate \| Cairo | Local | High | 4040 | 18-24 (n=603); 25-29 (n=1279); 30-39 | 61.5 | Non-probability | Health care workers and caregivers | 1.3% | Artron Laboratories \| Artron One Step COVID-19 | 0.4%, 1.0% (FINDDx / MUHC | - |

| Article Author (Study Organization) | Sampling Dates (YMD) | Location | Geographic Scope | Overall Risk of Bias | Sample Size | Age | Female (%) | Sampling method | Sample frame | Uncorrected Seroprevalence (95% Confidence Interval) | Test Manufacturers \| Names \| Isotypes | Test Sens, Spec (Source) | Corrected Seroprevalence (95% Credible Interval) |
|---|---|---|---|---|---|---|---|---|---|---|---|---|---|
| | | | | | | (n=1057); 40-49 (n=700); >=50 (n=401) | | | | | IgM/IgG Antibody Test \| IgG, IgM | independent evaluation) | |
| **Iran** | | | | | | | | | | | | | |
| Poustchi (Tehran University of Medical Sciences)[505] | 2020-04-17 to 2020-06-02 | Kerman, Shiraz, Esfahan , Urmia, Tehran, Sari, Ahvaz, Zahedan, Qom, Ardabil, Hamedan, Sanandaj, Kermanshah, Mashhad, Gorgan, Tabriz, Babol , Rasht | National | Moderate | 5372 | ≤19 (n = 11); 20–29 (n = 770); 30–39 (n = 2003); 40–49 (n = 1675); 50–59 (n =749); ≥60 (n = 164) | NR | Probability | Multiple populations | 14.9% (13.8-16.0%) | Pishtaz Teb \| PT-SARS-COV-2.IgM-96 and PT-SARS-COV-2.IgG-96 \| IgG, IgM | 66.9%, 98.2% (Author-reported independent evaluation) | 19.4% (14.1-23.7%) |
| Saberian (Tehran University of Medical Sciences)[506] | 2020-03-20 to 2020-05-20 | Tehran \| Tehran | Sublocal | High | 243 | Mean 36.14 (SD 8.70) | 12.3 | Non-probability | Essential non-healthcare workers | 41.6% | KarmaAzmaAndish Co. \| COVID-19 IgM/IgG rapid test \| IgG, IgM | 71.1%, 58.7% (Test developer / manufacturer evaluation) | 13.4% (0.0-46.8%) |
| Armin (High Institute for Research and Education in Transfusion Medicine)[507] | 2020-04-20 to 2020-05-05 | Tehran \| Tehran | Sublocal | High | 475 | <40 (n=238), 40-62 (n=97) | 81.5 | Non-probability | Health care workers and caregivers | 29.4% | Hangzhou AllTest Biotech Co. \| Rapid Test Dipstick \| IgG, IgM | 0.6%, 1.0% (FINDDx / MUHC independent evaluation) | - |
| Vazirinejad (University of Pecs)[508] | 2020-03-01 to 2020-04-30 | Kerman | Local | High | 238 | Mean 30.77 (SD 19.3) | 54.2 | Non-probability | Contacts of COVID patients | 15.6% | NR \| NR \| IgG, IgM | No data | - |
| Ossareh (Iran University of Medical Sciences)[510] | 2020-02-20 to 2020-03-20 | Tehran \| Tehran | Sublocal | High | 178 | Mean 58.9 (SD 16.5) | 31.5 | Non-probability | Multiple populations | 18.0% | Pishtaz Diagnostics Iran \| ELISA Pishtaz Teb Diagnostics Iran \| IgG, IgM | 75.6%, 97.6% (Used Bastos SR/MA data; no sens, spec, or author adjustment available) | 20.7% (13.5-28.7%) |

| Article Author (Study Organization) | Sampling Dates (YMD) | Location | Geographic Scope | Overall Risk of Bias | Sample Size | Age | Female (%) | Sampling method | Sample frame | Uncorrected Seroprevalence (95% Confidence Interval) | Test Manufacturers \| Names \| Isotypes | Test Sens, Spec (Source) | Corrected Seroprevalence (95% Credible Interval) |
|---|---|---|---|---|---|---|---|---|---|---|---|---|---|
| **Kuwait** | | | | | | | | | | | | | |
| Alali (University of Kuwait)[511] | 2020-05-23 to 2020-06-26 | | National | High | 525 | Median 43 (NR) | 0.0 | Probability | Essential non-healthcare workers | 38.1% (34.0-42.3%) | Biozek Medical; Netherlands \| COVID-19 Rapid Test Cassette \| IgG, IgM | 66.0%, 97.0% (Test developer / manufacturer evaluation) | 58.9% (42.4-83.2%) |
| **Libya** | | | | | | | | | | | | | |
| Kammon (Al-Zintan University)[512] | 2020-04-02 to 2020-05-18 | Libya \| Alzintan City | Local | High | 142 | <=40 (n = 78); >40 (n = 64) | 65.5 | Probability | Multiple populations | 4.2% | Guangzhou Wondfo Biotech \| One Step Novel COVID-19 IgM/IgG Antibody Test \| IgG, IgM | 0.4%, 1.0% (FINDDx / MUHC independent evaluation) | - |
| Kammon (Al-Zintan University)[512] | 2020-04-02 to 2020-05-18 | Libya \| Alzintan City | Local | High | 77 | <=40 (n = 59); >40 (n = 18) | 29.9 | Probability | Health care workers and caregivers | 0.0% | Guangzhou Wondfo Biotech \| One Step Novel COVID-19 IgM/IgG Antibody Test \| IgG, IgM | 0.4%, 1.0% (FINDDx / MUHC independent evaluation) | - |
| **Qatar** | | | | | | | | | | | | | |
| Jeremijenko (Cornell University )[514] | 2020-06-21 to 2020-09-09 | | National | High | 4970 | <29 (n=1579), 30-39 (n=1973), 40-49 (n=1040), 50+ (n=339) | 5.0 | Non-probability | Non-essential workers and unemployed persons | 64.4% (63.0-65.7%) | Roche Diagnostics \| Roche Elecsys® Anti-SARS-CoV-2 \| NR | 0.9%, 1.0% (FINDDx / MUHC independent evaluation) | - |
| MohamedH.Al-Thani (Ministry of Public Health)[515] | 2020-07-26 to 2020-09-09 | | National | Moderate | 2641 | Median 35 (Range 18-80) | NR | Probability | Non-essential workers and unemployed persons | 55.3% (53.3-57.3%) | Switzerland; Roche \| Roche Elecsys Anti-SARS-CoV-2 \| NR | 0.9%, 1.0% (FINDDx / MUHC independent evaluation) | 45.3% (42.0-49.7%) |
| Nasrallah (Cornell University - Qatar)[516] | 2020-07-26 to 2020-09-09 | | National | High | 394 | NR | NR | Non-probability | Essential non-healthcare workers | 40.6% (35.9-45.5%) | France; BioMérieux; Marcy-l'Etoile \| | 88.6%, 99.9% (Test developer / | 46.3% (37.0-56.9%) |

| Article Author (Study Organization) | Sampling Dates (YMD) | Location | Geographic Scope | Overall Risk of Bias | Sample Size | Age | Female (%) | Sampling method | Sample frame | Uncorrected Seroprevalence (95% Confidence Interval) | Test Manufacturers \| Names \| Isotypes | Test Sens, Spec (Source) | Corrected Seroprevalence (95% Credible Interval) |
|---|---|---|---|---|---|---|---|---|---|---|---|---|---|
| | | | | | | | | | | | BioMérieux VidasIII \| IgG | manufacturer evaluation) | |
| **Saudi Arabia** | | | | | | | | | | | | | |
| Alharbi (King Fahad General Hospital)[518] | 2020-04-18 to 2020-06-17 | Hejaz \| Madinah | Local | High | 330 | NR | NR | Non-probability | Health care workers and caregivers | 24.2% | Vircell Microbiologists \| IgM/IgA Kit #MA1032, IgG Kit #G1032 \| IgG, IgA, IgM | 75.6%, 97.6% (Used Bastos SR/MA data; no sens, spec, or author adjustment available) | 29.4% (23.2-36.2%) |
| Alserehi (Saudi Center for Disease Prevention and Control)[519] | 2020-05-20 to 2020-05-30 | Qassim, Emirate of the Eastern Province, Baha, Najran, Norther Border, Jazan, Madinah, Riyadh, Makkah, Hail, Aseer | National | Moderate | 12621 | NR | NR | Non-probability | Health care workers and caregivers | 2.4% | Abbott Laboratories \| Abbott Architect SARS-CoV-2 IgG \| IgG | 0.4%, 1.0% (FINDDx / MUHC independent evaluation) | - |
| **Turkey** | | | | | | | | | | | | | |
| Alkurt (University of Health Sciences)[521] | 2020-05-30 to 2020-06-06 | Istanbul, Darica | Local | Moderate | 813 | Mean 34.8 (SD 9.54) | 64.0 | Non-probability | Health care workers and caregivers | 2.7% | Abbott \| Abbott Architect SARS-CoV-2 IgG assay \| IgG | 0.4%, 1.0% (FINDDx / MUHC independent evaluation) | - |
| **South Asia** | | | | | | | | | | | | | |
| **India** | | | | | | | | | | | | | |
| Goenka (Apollo Gleneagles Hospitals)[528] | 2020-08-01 to 2020-08-31 | West Bengal \| Kolkata | Sublocal | High | 117 | <30 (n=57); 31-40 (n=35); 41-50 (n=19); >50 (n=6) | 47.0 | Non-probability | Health care workers and caregivers | 32.5% | Ortho Clinical Diagnostics \| Vitros ECi IgG assay \| IgG | 0.8%, 1.0% (FINDDx / MUHC independent evaluation) | - |
| Goenka (Apollo Gleneagles Hospitals)[532] | 2020-07-12 to 2020-08-23 | West Bengal \| Kolkata | Sublocal | Moderate | 1122 | <30 (n=364); 30-50 (n=665); >50 (n=93) | 34.6 | Probability | Health care workers and caregivers | 11.9% | Ortho Clinical Diagnostics \| Vitros anti-SARS-COV-2 IgG \| IgG | 0.8%, 1.0% (FINDDx / MUHC independent evaluation) | - |

| Article Author (Study Organization) | Sampling Dates (YMD) | Location | Geographic Scope | Overall Risk of Bias | Sample Size | Age | Female (%) | Sampling method | Sample frame | Uncorrected Seroprevalence (95% Confidence Interval) | Test Manufacturers \| Names \| Isotypes | Test Sens, Spec (Source) | Corrected Seroprevalence (95% Credible Interval) |
|---|---|---|---|---|---|---|---|---|---|---|---|---|---|
| Siddiqui (Max Super Specialty Hospital)[534] | 2020-04-15 to 2020-08-15 | NCT Delhi \| New Delhi | Local | High | 780 | 18-20 (n=19); 21-30 (n=239); 31-40 (n=204); 41-50 (n=139); 51-60 (n=74); >60 (n=33) | 29.4 | Non-probability | Multiple populations | 19.5% | Roche Diagnostics \| Elecsys Anti-SARS-CoV-2 kit \| NR | 0.7%, 1.0% (FINDDx / MUHC independent evaluation) | - |
| Kumar (Amrita Institute of Medical Sciences & Research Centre)[540] | 2020-07-11 to 2020-07-24 | Kerala | Sublocal | High | 635 | Mean 34.8 (Range 19-70) | NR | Non-probability | Health care workers and caregivers | 0.2% | Roche Diagnostics \| ElecsysR Anti-SARS- CoV-2 total antibodies assay \| NR | 0.9%, 1.0% (FINDDx / MUHC independent evaluation) | - |
| Malani (Tata Institute for Fundamental Research)[541] | 2020-06-29 to 2020-07-19 | Mumbai | Local | Low | 4202 | 12-24 (n=702), 25-39 (n=1480), 40-60 (n=1695), 61+ (n=325) | 54.3 | Probability | Persons living in slums | 54.1% (52.7-55.5%) | Abbott Laboratories \| Abbott Diagnostics Architect \| IgG | 0.4%, 1.0% (FINDDx / MUHC independent evaluation) | 45.1% (42.7-47.3%) |
| Unclear (Ahmedabad Municipal Corporation)[546] | 2020-06-16 to 2020-07-11 | Gujarat \| Ahmedabad | Local | Unclear | 30054 | NR | NR | Non-probability | Multiple populations | 17.6% | NR \| NR \| NR | No data | - |
| Kumar (Postgraduate Institute of Medical Education and Research)[549] | 2020-06-01 to 2020-06-30 | Maharashtra \| Mumbai | Local | High | 801 | 20-40 (n=413); 40-60 (n=380); >=60 (n=8) | 48.2 | Non-probability | Health care workers and caregivers | 11.1% (9.1-13.5%) | Roche Diagnostics \| Elecsys Anti-SARS-CoV-2 Assay \| NR | 0.9%, 1.0% (FINDDx / MUHC independent evaluation) | - |
| Singhal (Kokilaben Dhirubhai Ambani Hospital and Medical Research Institute)[550] | 2020-06-01 to 2020-06-30 | Maharashtra \| Mumbai | Sublocal | High | 208 | NR | NR | Non-probability | Health care workers and caregivers | 12.5% | 1) Abbott 2) Roche \| Architect, Abbott, USA, and later in the study by Elecsys®, Roche, Switzerland. \| NR | 97.8%, 97.8% (Used Bastos SR/MA data; no sens, spec, or author adjustment available) | 11.1% (6.4-16.0%) |

| Article Author (Study Organization) | Sampling Dates (YMD) | Location | Geographic Scope | Overall Risk of Bias | Sample Size | Age | Female (%) | Sampling method | Sample frame | Uncorrected Seroprevalence (95% Confidence Interval) | Test Manufacturers \| Names \| Isotypes | Test Sens, Spec (Source) | Corrected Seroprevalence (95% Credible Interval) |
|---|---|---|---|---|---|---|---|---|---|---|---|---|---|
| Khan (Government Medical College)[551] | 2020-06-15 to 2020-06-29 | Kashmir | Local | Moderate | 2905 | Mean 38.6 (NR) | 35.8 | Non-probability | Health care workers and caregivers | 2.5% (2.0-3.1%) | Abbott Laboratories \| NR \| IgG | 0.4%, 1.0% (FINDDx / MUHC independent evaluation) | - |
| Rao (Sri Jayadeva Institute of Cardiovascular Sciences and Research)[553] | 2020-05-23 to 2020-06-06 | Karnataka \| Bengaluru | Sublocal | Unclear | 1000 | 30-40 (n=1000) | 45.0 | Non-probability | Health care workers and caregivers | 1.0% | New Life Healthcare \| NR \| IgG | No data | - |
| **Pakistan** | | | | | | | | | | | | | |
| Zaidi (National Institute of Blood Diseases and Bone Marrow Transplant)[557] | 2020-05-15 to 2020-07-15 | Karachi | Local | High | 1675 | 18-60 | NR | Non-probability | Multiple populations | 36.0% | Roche Diagnostics \| electrochemiluminescence immunoassay analyzer method on Cobas e-411 \| IgG, IgA, IgM | 0.6%, 1.0% (FINDDx / MUHC independent evaluation) | - |
| Noor (Hayatabad Medical Complex)[559] | 2020-07-13 to 2020-07-15 | Khyber Pakhtunkhwa \| Peshawar | Sublocal | Moderate | 1011 | Mean 33.25 (COVID-19 receiving hospital); 33.94 (Non-COVID-19 receiving hospital) (SD 8.71 (COVID-19 receiving hospital); 11.77 (Non-COVID-19 receiving hospital)) | 31.9 | Probability | Health care workers and caregivers | 30.8% | Roche \| Elecsys® Anti-SARS-CoV-2 immunoassay \| IgG, IgM | 0.8%, 1.0% (FINDDx / MUHC independent evaluation) | - |
| Haq (Riphah International University)[560] | 2020-06-15 to 2020-06-29 | Khyber Pakhtunkhwa \| Peshawar City | Local | Moderate | 1011 | Mean 33.6 (SD 10.5) | 31.9 | Non-probability | Health care workers and caregivers | 30.7% (27.8-33.6%) | Roche Diagnostics \| Elecsys® Anti-SARS-CoV-2 antibody test \| NR | 0.9%, 1.0% (FINDDx / MUHC independent evaluation) | - |
| Chughtai (Chughtai | 2020-05-20 to 2020-05-30 | Lahore | Local | High | 154 | Mean 27.1 (SD 3.8) | 0.0 | Non-probability | Essential non- | 15.6% | Abbott \| Abbott Architect | 0.4%, 1.0% (FINDDx / | - |

| Article Author (Study Organization) | Sampling Dates (YMD) | Location | Geographic Scope | Overall Risk of Bias | Sample Size | Age | Female (%) | Sampling method | Sample frame | Uncorrected Seroprevalence (95% Confidence Interval) | Test Manufacturers \| Names \| Isotypes | Test Sens, Spec (Source) | Corrected Seroprevalence (95% Credible Interval) |
|---|---|---|---|---|---|---|---|---|---|---|---|---|---|
| Institute of Pathology)[561] | | | | | | | | | healthcare workers | | SARS-CoV-2 IgG assay \| IgG | MUHC independent evaluation) | |
| **Southeast Asia, East Asia, And Oceania** | | | | | | | | | | | | | |
| **China** | | | | | | | | | | | | | |
| Li (Fudan University)[562] | 2020-05-15 to 2020-10-15 | Shanghai Municipality \| Shanghai | Sublocal | High | 6414 | NR | NR | Non-probability | Patients seeking care for non-COVID-19 reasons | 0.3% | Bioscience \| NR \| IgG, IgM | 0.6%, 1.0% (FINDDx / MUHC independent evaluation) | - |
| He (Wuhan University)[564] | 2020-05-13 to 2020-06-10 | Hubei \| Wuhan | Sublocal | High | 1059 | Median 32.69 (NR) | 75.3 | Non-probability | Health care workers and caregivers | 9.3% | Livzon Diagnostics \| Diagnostic Kit for IgM/IgG Antibody to Coronavirus \| IgG | 0.7%, 1.0% (FINDDx / MUHC independent evaluation) | - |
| He (Renmin Hospital of Wuhan University)[565] | 2020-05-15 to 2020-05-29 | Hubei \| Wuhan | Sublocal | Unclear | 4614 | Median 36.86 (SD 10.644) | 70.3 | Non-probability | Health care workers and caregivers | 3.3% | Livzon Diagnostics Inc. \| Diagnostic Kit for IgM/IgG Antibody \| IgG | 0.5%, 0.6% (FINDDx / MUHC independent evaluation) | - |
| Tong (Nanjing University Medical School)[568] | 2020-05-12 to 2020-05-15 | Hubei \| Wuhan | Local | High | 191 | Mean 32 (Range 24-58) | 70.2 | Non-probability | Health care workers and caregivers | 0.0% | Shenzhen YHLO Biotech Co \| NR \| IgG | 0.7%, 1.0% (FINDDx / MUHC independent evaluation) | - |
| Liang (The First Affiliated Hospital of Guangzhou Medical University)[569] | 2020-01-20 to 2020-04-30 | Guangdong \| Guangzhou | Local | High | 8782 | Median 54 (IQR 44-62) | 51.6 | Non-probability | Multiple populations | 0.6% | Shenzhen YHLO Biotech Co \| SARS-CoV-2 IgM/IgG CLIA kits \| IgG, IgM | 97.8%, 97.8% (Used Bastos SR/MA data; no sens, spec, or author adjustment available) | 0.0% (0.0-0.1%) |
| Liang (The First Affiliated Hospital of Guangzhou Medical University)[569] | 2020-01-19 to 2020-04-30 | Hubei \| Wuhan | Local | High | 8272 | Median 55 (IQR 38-67) | 50.0 | Non-probability | Multiple populations | 2.1% | Innovita \| 2019-nCoV Ab Test (Colloidal Gold) \| IgG, IgM | 0.6%, 1.0% (FINDDx / MUHC independent evaluation) | - |

| Article Author (Study Organization) | Sampling Dates (YMD) | Location | Geographic Scope | Overall Risk of Bias | Sample Size | Age | Female (%) | Sampling method | Sample frame | Uncorrected Seroprevalence (95% Confidence Interval) | Test Manufacturers \| Names \| Isotypes | Test Sens, Spec (Source) | Corrected Seroprevalence (95% Credible Interval) |
|---|---|---|---|---|---|---|---|---|---|---|---|---|---|
| Liu (Zhongnan Hospital of Wuhan University)[571] | 2020-02-29 to 2020-04-29 | Hubei \| Wuhan | Local | Moderate | 3832 | Mean 37.1 (95% CI 36.7-37.4) | 32.3 | Non-probability | Health care workers and caregivers | 4.0% (3.4-4.7%) | YHLO \| SARS-CoV-2 IgM/IgG CLIA kits \| IgG | 0.7%, 1.0% (FINDDx / MUHC independent evaluation) | - |
| Lei ( Tongji Medical College)[572] | 2020-02-17 to 2020-04-28 | Hubei \| Wuhan | Local | High | 11766 | NR | NR | Non-probability | Multiple populations | 0.5% | YHLO \| YHLO Biotech SARS-CoV-2 IgM/IgG Test \| IgG, IgM | 66.7%, 99.5% (Test developer / manufacturer evaluation) | 0.3% (0.0-0.7%) |
| Want (Huazhong University of Science and Technology)[574] | 2020-01-23 to 2020-04-24 | Hubei \| Xiangyang, Wuhan | Local | Unclear | 299 | Median 5.2 (Range 2 months - 16 years) | 58.9 | Non-probability | Patients seeking care for non-COVID-19 reasons | 0.0% | NR \| NR \| IgG, IgM | 97.8%, 97.8% (Used Bastos SR/MA data; no sens, spec, or author adjustment available) | 0.2% (0.0-1.0%) |
| Liu (Sun Yat-sen University)[575] | 2020-02-07 to 2020-04-21 | Hubei \| Wuhan | Local | High | 420 | Mean 35.8 (NR) | 68.1 | Non-probability | Health care workers and caregivers | 0.0% | YHLO \| SARS-CoV-2 IgM/IgG CLIA kits \| IgG | 1.0%, 1.0% (FINDDx / MUHC independent evaluation) | - |
| Wu (Wuhan University)[577] | 2020-04-03 to 2020-04-15 | Hubei \| Wuhan | Sublocal | High | 1021 | NR | NR | Non-probability | Essential non-healthcare workers | 9.6% | Innovita Biological Technology Co. Ltd \| 2019-nCoV Ab Test \| IgG, IgM | 0.6%, 1.0% (FINDDx / MUHC independent evaluation) | - |
| Xu (National Clinical Research Centre for Kidney Disease)[578] | 2020-03-09 to 2020-04-10 | Hubei, Sichuan, Guangdong \| Chengdu, Wuhan, Guangzhou | National | High | 10449 | Median 50 (IQR 35-49 ) | 52.3 | Non-probability | Multiple populations | 0.8% | Bioscience \| Magnetic Chemiluminescence Enzyme Immunoassay Kit \| IgG, IgM | 97.8%, 97.8% (Used Bastos SR/MA data; no sens, spec, or author adjustment available) | 0.0% (0.0-0.1%) |
| Xu (National Clinical Research Centre for Kidney Disease)[578] | 2020-03-09 to 2020-04-10 | Hubei, Sichuan, Guangdong \| Jinzhou, Chongqing, Foshan, Chengdu, Honghu, Wuhan, Guangzhou | National | High | 6919 | Median 41 (IQR 31-56 ) | 63.1 | Non-probability | Multiple populations | 2.5% | Bioscience \| Magnetic Chemiluminescence Enzyme Immunoassay Kit \| IgG, IgM | 97.8%, 97.8% (Used Bastos SR/MA data; no sens, spec, or author adjustment available) | 0.3% (0.0-0.8%) |

| Article Author (Study Organization) | Sampling Dates (YMD) | Location | Geographic Scope | Overall Risk of Bias | Sample Size | Age | Female (%) | Sampling method | Sample frame | Uncorrected Seroprevalence (95% Confidence Interval) | Test Manufacturers \| Names \| Isotypes | Test Sens, Spec (Source) | Corrected Seroprevalence (95% Credible Interval) |
|---|---|---|---|---|---|---|---|---|---|---|---|---|---|
| Zhang (Huazhong University of Science and Technology)[580] | 2020-01-23 to 2020-03-27 | Hubei \| Wuhan | Sublocal | High | 44 | Median 6 (Range 1-15) | 38.6 | Non-probability | Patients seeking care for non-COVID-19 reasons | 0.0% | NR \| NR \| IgG, IgM | 97.8%, 97.8% (Used Bastos SR/MA data; no sens, spec, or author adjustment available) | 1.6% (0.0-6.6%) |
| Zhou (Zhongnan Hospital of Wuhan University)[581] | 2020-03-16 to 2020-03-25 | Hubei \| Wuhan | Sublocal | Moderate | 3674 | 18-30 (n=1378); 31-50 (n=1656); >50 (n=640) | 67.7 | Non-probability | Health care workers and caregivers | 1.7% | Shenzhen Yahuilong Biotechnology Co. \| COVID-19 IgM/IgG chemiluminescence test kit \| IgG | 0.9%, 1.0% (FINDDx / MUHC independent evaluation) | - |
| Li (National Center for Medical Service Administration)[582] | 2020-02-10 to 2020-03-22 | Hubei \| Wuhan | Sublocal | High | 300 | Mean 46.3 (SD 10.3) | NR | Non-probability | Contacts of COVID patients | 47.7% | Innovita Biological Technology Co. Ltd \| NR \| IgG, IgM | 0.6%, 1.0% (FINDDx / MUHC independent evaluation) | 52.2% (46.7-57.6%) |
| Tang ( Tongji Medical College)[583] | 2020-03-08 to 2020-03-21 | Hubei \| Wuhan | Local | Moderate | 1027 | Mean 60.3 (SD 13.4) | 39.7 | Non-probability | Patients seeking care for non-COVID-19 reasons | 4.6% | Ltd."; "Tangshan Yingnuote Biotechnology Enterprise Co. \| Tangshan Colloidal Gold Immunochromatography Assay \| IgG, IgM | 0.9%, 1.0% (FINDDx / MUHC independent evaluation) | - |
| Tu (Wuhan Institute of Virology)[584] | 2020-03-19 to 2020-03-20 | Hubei \| Wuhan | Sublocal | High | 325 | NR | NR | Probability | Health care workers and caregivers | 24.3% | Zhu Hai Livzon Diagnostics Ins \| Zhu Hai Livzon ELISA \| IgG | 0.7%, 1.0% (FINDDx / MUHC independent evaluation) | - |
| Zhang (Shenzhen Second People's Hospital)[585] | 2020-02-29 to 2020-03-18 | Guangdong \| Shenzhen | Local | High | 1589 | Mean 36.4 (Range 11-89) | NR | Non-probability | Non-essential workers and unemployed persons | 0.4% | Shenzhen Sciarray Biotech Ltd \| shenzhen ELISA \| IgG | 75.6%, 97.6% (Used Bastos SR/MA data; no sens, spec, or author adjustment available) | 0.1% (0.0-0.3%) |

| Article Author (Study Organization) | Sampling Dates (YMD) | Location | Geographic Scope | Overall Risk of Bias | Sample Size | Age | Female (%) | Sampling method | Sample frame | Uncorrected Seroprevalence (95% Confidence Interval) | Test Manufacturers \| Names \| Isotypes | Test Sens, Spec (Source) | Corrected Seroprevalence (95% Credible Interval) |
|---|---|---|---|---|---|---|---|---|---|---|---|---|---|
| Xiong (Huazhong University of Science and Technology)[586] | 2020-02-12 to 2020-03-17 | Hubei \| Wuhan | Local | Unclear | 797 | Median 31 (Range 23-53) | 88.5 | Non-probability | Health care workers and caregivers | 4.4% | NR \| NR \| IgG | 75.6%, 97.6% (Used Bastos SR/MA data; no sens, spec, or author adjustment available) | 2.5% (0.6-4.8%) |
| Zhao (Renmin Hospital of Wuhan University)[588] | 2020-01-14 to 2020-02-21 | Hubei \| Wuhan | Sublocal | High | 1060 | NR | NR | Non-probability | Health care workers and caregivers | 8.3% | Shenzhen YHLO Biotech Co \| NR \| IgG, IgM | 97.8%, 97.8% (Used Bastos SR/MA data; no sens, spec, or author adjustment available) | 6.3% (4.6-8.2%) |
| Chen (Nanjing University Medical School)[589] | 2020-02-19 to 2020-02-19 | Jiangsu \| Nanjing | Local | High | 105 | Median 30 (IQR 26-62) | 79.0 | Non-probability | Health care workers and caregivers | 17.1% | Author-designed \| N/A - Author-designed \| IgG, IgM | 93.3%, 100.0% (Test developer / manufacturer evaluation) | 17.5% (10.1-26.2%) |
| Zhang (Shengjing Hospital of China Medical University)[590] | 2020-01-21 to 2020-02-16 | Liaoning \| Shenyang | Regional | High | 63 | Median 40 (Range 25-61) | 88.9 | Non-probability | Health care workers and caregivers | 0.0% | Shenzhen Yahuilong Biotechnology Co. \| Chemiluminescence Detection Kit \| IgG | 0.9%, 1.0% (FINDDx / MUHC independent evaluation) | - |
| **Thailand** | | | | | | | | | | | | | |
| Nopsopon (Chulalongkorn University)[593] | 2020-04-08 to 2020-06-26 | | National | Moderate | 857 | Median 37 (IQR 27-45) | 74.7 | Non-probability | Multiple populations | 5.5% (4.1-7.2%) | Baiya Phytopharm \| Baiya Rapid COVID-19 IgG/IgM Test Kit \| IgM | 94.1%, 98.0% (Author-reported independent evaluation) | 3.5% (0.4-6.1%) |
| Nopsopon (Chulalongkorn University)[594] | 2020-04-17 to 2020-05-17 | Ranong | Sublocal | Moderate | 844 | Median 42 (IQR 32-50) | 71.7 | Non-probability | Health care workers and caregivers | 0.8% (0.4-1.7%) | Baya Phytopharm; Thailand \| Baya Rapid IgG/IgM test kit \| IgG, IgM | 94.1%, 98.0% (Test developer / manufacturer evaluation) | 0.3% (0.0-0.9%) |
| **Vietnam** | | | | | | | | | | | | | |

| Article Author (Study Organization) | Sampling Dates (YMD) | Location | Geographic Scope | Overall Risk of Bias | Sample Size | Age | Female (%) | Sampling method | Sample frame | Uncorrected Seroprevalence (95% Confidence Interval) | Test Manufacturers \| Names \| Isotypes | Test Sens, Spec (Source) | Corrected Seroprevalence (95% Credible Interval) |
|---|---|---|---|---|---|---|---|---|---|---|---|---|---|
| Chau (Oxford University Clinical Research Unit)[595] | 2020-08-23 to 2020-08-30 | Đông Nam Bộ \| Ho Chi Minh City | Sublocal | High | 408 | Median 32 (Range 20-60) | 64.7 | Non-probability | Health care workers and caregivers | 0.0% | Roche Diagnostics \| Elecsys Anti–SARS– CoV– 2 assay \| NR | 0.9%, 1.0% (FINDDx / MUHC independent evaluation) | - |
| **Sub-Saharan Africa** | | | | | | | | | | | | | |
| **Malawi** | | | | | | | | | | | | | |
| Chibwana (Malawi-Liverpool-Wellcome Trust Clinical Research Programme)[600] | 2020-05-22 to 2020-06-19 | Southern Region Malawi \| Blantyre | National | High | 500 | Median 31 (Range 20-64) | 52.8 | Non-probability | Health care workers and caregivers | 16.8% | Omega diagnostics \| COVID-19 IgG ELISA \| IgG | 88.1%, 93.2% (Test developer / manufacturer evaluation) | 11.3% (1.9-19.1%) |
| **Nigeria** | | | | | | | | | | | | | |
| Asuquo (Joseph Ukpo Hospitals and Research Institutes)[602] | 2020-06-17 to 2020-06-24 | Cross River State \| Calabar | Sublocal | High | 66 | NR | 42.4 | Non-probability | Multiple populations | 26.0% | Healgen Scientific \| COVID-19 IgG / IgM Rapid Test Cassette \| IgG | 1.0%, 1.0% (FINDDx / MUHC independent evaluation) | - |
| **South Africa** | | | | | | | | | | | | | |
| Shaw (Stellenbosch University)[603] | 2020-08-17 to 2020-09-04 | Cape Town | Sublocal | High | 405 | Median 38 (Range 18-69) | 53.6 | Non-probability | Essential non-healthcare workers | 23.7% | Abbott Laboratories \| Abbott SARS-CoV-2 IgG assay \| IgG | 0.4%, 1.0% (FINDDx / MUHC independent evaluation) | - |
| **Togo** | | | | | | | | | | | | | |
| Halatoko (Universite de Lome)[605] | 2020-04-23 to 2020-05-08 | Lome | Local | High | 955 | Median 36 (IQR 32-43) | 28.4 | Non-probability | Non-essential workers and unemployed persons | 0.9% (0.4-1.8%) | "Hangzhou Clongene Biotech Co; Ltd." \| Lungene Rapid Test \| IgG, IgM | 0.7%, 0.9% (FINDDx / MUHC independent evaluation) | - |

#### S3 Table. Risk of bias results for each included study

| Article Author (Study Organization) | Sampling End Date (YMD) | Item 1: Appropriate sample frame* | Item 2: Probability sampling method | Item 3: Adequate sample size | Item 4: Subjects & setting described | Item 5: Good coverage of sample | Item 6: Sens > 90% & spec > 95% | Item 7: Same tests for all subjects | Item 8: Appropriate analysis | Item 9: Adequate response rate | Overall Risk of Bias† |
|---|---|---|---|---|---|---|---|---|---|---|---|
| **Central Europe, Eastern Europe, and Central Asia** | | | | | | | | | | | |
| **Armenia** | | | | | | | | | | | |
| Grigoryan | 2020-06-29 | Yes | Unclear | No | No | Unclear | Yes | Yes | No | Unclear | High |
| **Bulgaria** | | | | | | | | | | | |
| Tsaneva-Damyanova | 2020-04-20 | Yes | No | No | No | Unclear | No | Yes | No | Unclear | High |
| **Croatia** | | | | | | | | | | | |
| Vince | 2020-07-31 | Yes | No | No | Yes | Unclear | Yes | Yes | No | Yes | Moderate |
| Lapic | 2020-05-31 | No | Unclear | Yes | Yes | Yes | Yes | Yes | No | Unclear | Moderate |
| Vilibic-Cavlek | 2020-05-24 | Yes | No | No | Yes | Yes | Unclear | Yes | No | Unclear | High |
| Jerkovic | 2020-04-28 | Yes | No | Yes | Yes | No | Yes | Yes | No | Unclear | High |
| **Czechia** | | | | | | | | | | | |
| Bloomfield | 2020-08-19 | Yes | No | No | Yes | Unclear | Yes | Yes | No | Unclear | High |
| Malickova | 2020-04-15 | Yes | Unclear | No | Yes | Yes | Unclear | Yes | No | Unclear | Unclear |
| **Estonia** | | | | | | | | | | | |
| Jogi | 2020-07-31 | Yes | Yes | Yes | Yes | Yes | Yes | Yes | No | No | Moderate |
| Jogi | 2020-07-31 | Yes | Yes | Yes | Yes | Yes | Yes | Yes | No | No | Moderate |
| Veerus | 2020-06-10 | No | No | No | No | Unclear | Yes | Yes | No | Unclear | High |
| **Georgia** | | | | | | | | | | | |
| Tsertsvadze | 2020-05-27 | Yes | No | Yes | Yes | Yes | Yes | Yes | Yes | Yes | Moderate |

| Article Author (Study Organization) | Sampling End Date (YMD) | Item 1: Appropriate sample frame* | Item 2: Probability sampling method | Item 3: Adequate sample size | Item 4: Subjects & setting described | Item 5: Good coverage of sample | Item 6: Sens > 90% & spec > 95% | Item 7: Same tests for all subjects | Item 8: Appropriate analysis | Item 9: Adequate response rate | Overall Risk of Bias† |
|---|---|---|---|---|---|---|---|---|---|---|---|
| **Hungary** | | | | | | | | | | | |
| Merkely | 2020-05-16 | Yes | Yes | Yes | Yes | Yes | Unclear | Yes | No | No | Moderate |
| **Poland** | | | | | | | | | | | |
| Ciechanowicz | 2020-10-31 | Yes | No | No | Yes | Yes | Yes | Yes | No | Yes | High |
| Kasztelewicz | 2020-08-09 | Yes | No | Yes | Yes | Unclear | Yes | Yes | No | Yes | Moderate |
| Gujski | 2020-07-08 | Yes | Yes | Yes | Yes | Unclear | Yes | Yes | No | No | Moderate |
| **Romania** | | | | | | | | | | | |
| MedLife (MedLife) | 2020-04-21 | Yes | Unclear | No | No | Unclear | Unclear | No | No | Unclear | Unclear |
| **Russian Federation** | | | | | | | | | | | |
| Popova | 2020-07-19 | Yes | Yes | Yes | Yes | Unclear | Unclear | Yes | No | Unclear | Moderate |
| Popova | 2020-07-11 | Yes | Yes | Yes | Yes | Unclear | Unclear | Yes | Unclear | Unclear | Unclear |
| Popova | 2020-06-26 | Yes | Yes | Yes | Yes | Unclear | Unclear | Yes | No | Yes | High |
| Barchuk | 2020-06-26 | Yes | Yes | Yes | Yes | Yes | Yes | Yes | Yes | No | Low |
| Popova | 2020-06-20 | Yes | Yes | Yes | No | Unclear | Unclear | Yes | No | Yes | High |
| Popova | 2020-06-20 | Yes | Yes | Yes | No | Unclear | Unclear | Unclear | No | No | High |
| Tickle | 2020-06-18 | Yes | Yes | Yes | No | Unclear | Unclear | Unclear | No | Unclear | High |
| Krechetova | 2020-06-15 | No | Unclear | Yes | Unclear | Unclear | Unclear | Yes | No | Unclear | Unclear |
| Kiselyov | 2020-06-10 | Yes | Unclear | Yes | No | Unclear | Yes | Yes | No | Unclear | Moderate |
| **High-income** | | | | | | | | | | | |
| **Andorra** | | | | | | | | | | | |
| Royo-Cebrecos | 2020-05-28 | Yes | No | Yes | Yes | Yes | Yes | Yes | No | Yes | Moderate |

| Article Author (Study Organization) | Sampling End Date (YMD) | Item 1: Appropriate sample frame* | Item 2: Probability sampling method | Item 3: Adequate sample size | Item 4: Subjects & setting described | Item 5: Good coverage of sample | Item 6: Sens > 90% & spec > 95% | Item 7: Same tests for all subjects | Item 8: Appropriate analysis | Item 9: Adequate response rate | Overall Risk of Bias[†] |
|---|---|---|---|---|---|---|---|---|---|---|---|
| **Argentina** | | | | | | | | | | | |
| Munoz | 2020-07-16 | Yes | Yes | No | Yes | Yes | Yes | Yes | No | Yes | Moderate |
| Silva | 2020-07-06 | Yes | Yes | Yes | No | Yes | Yes | Yes | No | Yes | Moderate |
| Figar | 2020-07-01 | Yes | No | No | No | Unclear | No | Yes | No | Unclear | High |
| Insua | 2020-06-09 | Yes | Yes | No | No | Unclear | Yes | Yes | No | Unclear | High |
| **Australia** | | | | | | | | | | | |
| Hicks | 2020-06-15 | No | Unclear | Yes | Yes | Unclear | Yes | Yes | No | Unclear | Unclear |
| Macartney | 2020-04-10 | Yes | No | Yes | No | Yes | Yes | Yes | No | Unclear | Moderate |
| **Austria** | | | | | | | | | | | |
| Ladage | 2020-06-15 | Yes | Yes | Yes | Yes | No | Unclear | Yes | No | No | High |
| Fuereder | 2020-06-04 | Yes | No | No | Yes | Unclear | Unclear | Yes | No | Unclear | High |
| Fuereder | 2020-06-04 | Yes | No | No | Yes | Unclear | Unclear | Yes | No | Unclear | High |
| Hackner | 2020-04-30 | No | No | No | Yes | Unclear | No | Yes | No | Unclear | High |
| Knabl | 2020-04-27 | Yes | Yes | Yes | Yes | Yes | Yes | Yes | No | Yes | Moderate |
| Wagner | 2020-04-17 | No | Unclear | Yes | Yes | Yes | Unclear | Yes | No | Unclear | High |
| Reiter | 2020-03-29 | Yes | No | No | Yes | Unclear | Unclear | Yes | No | Yes | High |
| Orth-Holler | 2020-03-27 | Yes | No | No | Yes | No | No | Yes | No | Unclear | High |
| **Belgium** | | | | | | | | | | | |
| Duysburgh | 2020-09-30 | Yes | Unclear | Yes | No | Unclear | Unclear | Yes | No | Unclear | Unclear |
| Herzog | 2020-07-04 | No | No | Yes | Yes | Yes | No | Yes | Yes | Unclear | Moderate |
| Blairon | 2020-06-19 | Yes | No | Yes | Yes | Yes | Unclear | Yes | No | No | High |

| Article Author (Study Organization) | Sampling End Date (YMD) | Item 1: Appropriate sample frame* | Item 2: Probability sampling method | Item 3: Adequate sample size | Item 4: Subjects & setting described | Item 5: Good coverage of sample | Item 6: Sens > 90% & spec > 95% | Item 7: Same tests for all subjects | Item 8: Appropriate analysis | Item 9: Adequate response rate | Overall Risk of Bias† |
|---|---|---|---|---|---|---|---|---|---|---|---|
| Herzog | 2020-06-13 | No | No | Yes | Yes | Yes | No | Yes | Yes | Unclear | Moderate |
| Herzog | 2020-05-25 | No | No | Yes | Yes | Yes | No | Yes | Yes | Unclear | Moderate |
| Berardis | 2020-05-19 | Yes | No | No | Yes | Yes | No | Yes | No | Unclear | High |
| Martin | 2020-05-18 | Yes | No | No | Yes | Unclear | Unclear | Yes | No | No | High |
| Steensels | 2020-04-30 | No | No | Yes | No | Yes | Yes | Yes | Yes | Yes | High |
| Herzog | 2020-04-26 | No | No | Yes | Yes | Yes | No | Yes | Yes | Unclear | Moderate |
| Buntinx | 2020-04-16 | Yes | No | No | No | No | No | Yes | No | Yes | High |
| Herzog | 2020-04-05 | No | No | Yes | Yes | Yes | No | Yes | Yes | Unclear | Moderate |
| **Canada** | | | | | | | | | | | |
| Canadian Blood Services | 2020-11-25 | No | Yes | Yes | Yes | Yes | Yes | Yes | Yes | Yes | Moderate |
| Canadian Blood Services | 2020-10-31 | No | No | Yes | Yes | Yes | Yes | Yes | Yes | Yes | Moderate |
| Manny | 2020-10-23 | Yes | No | No | Yes | Unclear | No | Yes | No | Unclear | High |
| Brousseau | 2020-09-24 | No | No | Yes | Yes | Yes | No | Yes | No | No | High |
| Bolotin | 2020-08-21 | No | No | Yes | No | Unclear | No | Yes | No | Unclear | High |
| Bolotin | 2020-07-31 | No | No | Yes | No | Unclear | No | Yes | No | Unclear | High |
| Bardai | 2020-07-27 | Yes | No | No | Yes | No | Unclear | Yes | No | Unclear | High |
| Hema-Quebec | 2020-07-09 | No | Yes | Yes | No | Unclear | Unclear | Unclear | No | Unclear | Unclear |
| Bolotin | 2020-06-30 | No | No | Yes | No | Unclear | No | Yes | No | Unclear | High |
| Majdoubi | 2020-06-19 | No | No | No | Yes | Unclear | Yes | Yes | No | Unclear | High |

| Article Author (Study Organization) | Sampling End Date (YMD) | Item 1: Appropriate sample frame* | Item 2: Probability sampling method | Item 3: Adequate sample size | Item 4: Subjects & setting described | Item 5: Good coverage of sample | Item 6: Sens > 90% & spec > 95% | Item 7: Same tests for all subjects | Item 8: Appropriate analysis | Item 9: Adequate response rate | Overall Risk of Bias† |
|---|---|---|---|---|---|---|---|---|---|---|---|
| Canadian Blood Services | 2020-06-18 | No | Unclear | Yes | Yes | Yes | Yes | Yes | Yes | Yes | Moderate |
| Bolotin | 2020-05-30 | No | No | Yes | No | Unclear | No | Yes | No | Unclear | High |
| Skowronski | 2020-05-27 | No | Yes | Yes | Yes | Yes | No | Yes | No | Unclear | Moderate |
| Vijh | 2020-05-14 | Yes | No | No | Yes | Yes | Yes | Yes | No | No | High |
| Bolotin | 2020-04-30 | No | No | Yes | No | Unclear | No | Yes | No | Unclear | High |
| Skowronski | 2020-03-13 | No | Yes | Yes | Yes | Yes | No | Yes | No | Unclear | Moderate |
| **Chile** | | | | | | | | | | | |
| Torres | 2020-05-19 | No | Yes | Yes | Yes | Yes | Unclear | Yes | No | Yes | High |
| Torres | 2020-05-19 | No | Yes | No | Yes | Yes | Unclear | Yes | No | Yes | High |
| **Denmark** | | | | | | | | | | | |
| Laursen | 2020-08-10 | No | No | Yes | Yes | Yes | Unclear | Yes | No | Unclear | High |
| Egerup | 2020-07-03 | No | No | Yes | Yes | Unclear | No | Yes | No | Unclear | High |
| Egerup | 2020-07-03 | No | No | Yes | No | Unclear | No | Yes | No | Yes | Moderate |
| Egerup | 2020-07-03 | No | No | Yes | Yes | Unclear | No | Yes | No | Yes | Moderate |
| Jespersen | 2020-06-30 | No | Unclear | No | No | Unclear | Yes | Yes | No | Unclear | High |
| Jespersen | 2020-06-30 | No | Unclear | No | No | Unclear | Yes | Yes | No | Unclear | High |
| Pedersen | 2020-06-26 | No | No | Yes | No | Unclear | Yes | Yes | Yes | Unclear | Moderate |
| Jespersen | 2020-06-19 | No | No | Yes | Yes | Yes | Yes | Yes | No | Yes | Moderate |
| Pedersen | 2020-06-19 | No | Yes | Yes | Yes | Unclear | Yes | Yes | Yes | No | Moderate |
| Storgaard | 2020-06-04 | Yes | No | No | Yes | No | Yes | Yes | No | Unclear | High |

| Article Author (Study Organization) | Sampling End Date (YMD) | Item 1: Appropriate sample frame* | Item 2: Probability sampling method | Item 3: Adequate sample size | Item 4: Subjects & setting described | Item 5: Good coverage of sample | Item 6: Sens > 90% & spec > 95% | Item 7: Same tests for all subjects | Item 8: Appropriate analysis | Item 9: Adequate response rate | Overall Risk of Bias[†] |
|---|---|---|---|---|---|---|---|---|---|---|---|
| Freiesleben | 2020-05-21 | No | No | No | Yes | No | Unclear | Yes | No | Yes | High |
| Erikstrup | 2020-05-03 | No | Yes | Yes | Yes | Yes | Yes | Yes | No | Yes | Moderate |
| Iversen | 2020-04-22 | Yes | Yes | Yes | Yes | Yes | No | Yes | Yes | Yes | Low |
| Iversen | 2020-04-22 | No | No | Yes | No | Unclear | Yes | Yes | No | Unclear | High |
| Freiesleben | 2020-04-17 | No | No | Yes | Yes | No | Unclear | Yes | No | Yes | High |
| **Faroe Islands** | | | | | | | | | | | |
| Petersen | 2020-05-01 | Yes | Yes | Yes | Yes | Yes | Yes | Yes | Yes | Yes | Low |
| **Finland** | | | | | | | | | | | |
| Kantele | 2020-05-15 | Yes | No | Yes | Yes | Unclear | No | Yes | No | Yes | High |
| **France** | | | | | | | | | | | |
| Anna | 2020-07-31 | Yes | No | Yes | Yes | Unclear | Yes | Yes | No | Unclear | Moderate |
| Dimeglio | 2020-07-10 | No | No | Yes | Yes | Unclear | Unclear | Yes | No | No | High |
| Roederer | 2020-07-02 | Yes | No | Yes | Yes | Yes | Unclear | Yes | No | Unclear | High |
| Pere | 2020-06-26 | Yes | No | Yes | Yes | Unclear | Unclear | Yes | No | Unclear | High |
| Tsatsarisa | 2020-06-26 | Yes | No | No | Yes | Yes | Yes | Yes | No | Unclear | High |
| Gallais | 2020-06-26 | No | No | No | Yes | Yes | Yes | Yes | No | Unclear | High |
| Carrat | 2020-06-23 | Yes | Yes | Yes | Yes | Unclear | Yes | Yes | No | No | Moderate |
| Mesnil | 2020-06-22 | No | No | Yes | Yes | Unclear | Unclear | Yes | No | Yes | High |
| Delmas | 2020-06-17 | Yes | No | Yes | No | Unclear | Unclear | Yes | No | Yes | High |
| Capai | 2020-06-15 | No | No | Yes | Yes | Unclear | No | Yes | No | Yes | Moderate |
| Mattern | 2020-05-31 | Yes | No | No | No | No | Unclear | Yes | No | Unclear | High |

| Article Author (Study Organization) | Sampling End Date (YMD) | Item 1: Appropriate sample frame* | Item 2: Probability sampling method | Item 3: Adequate sample size | Item 4: Subjects & setting described | Item 5: Good coverage of sample | Item 6: Sens > 90% & spec > 95% | Item 7: Same tests for all subjects | Item 8: Appropriate analysis | Item 9: Adequate response rate | Overall Risk of Bias† |
|---|---|---|---|---|---|---|---|---|---|---|---|
| Bal | 2020-05-28 | No | Unclear | No | Unclear | Unclear | Unclear | Yes | No | Unclear | High |
| Vu | 2020-05-17 | No | No | Yes | Yes | Yes | Yes | Yes | Yes | Unclear | Moderate |
| Cohen | 2020-05-12 | No | No | Yes | Yes | Unclear | Yes | Yes | No | Unclear | High |
| Fumery | 2020-05-11 | No | No | No | Yes | Unclear | Yes | Yes | No | Yes | High |
| Fontanet | 2020-04-30 | Yes | Yes | Yes | Yes | Yes | Yes | Yes | No | No | Moderate |
| Solodky | 2020-04-16 | Yes | No | No | No | No | Unclear | Yes | No | Unclear | High |
| Solodky | 2020-04-16 | Yes | No | No | No | Unclear | Unclear | Yes | No | Unclear | High |
| Vu | 2020-04-12 | No | No | Yes | Yes | Yes | Yes | Yes | Yes | Unclear | Moderate |
| Gallian | 2020-04-07 | No | Yes | Yes | Yes | Unclear | Unclear | Yes | No | Unclear | High |
| Grzelak | 2020-03-24 | No | No | No | No | Unclear | Unclear | Yes | No | Unclear | High |
| Germain | 2020-03-16 | No | No | No | Yes | Yes | Yes | Yes | No | Unclear | High |
| Vu | 2020-03-15 | No | No | Yes | Yes | Yes | Yes | Yes | Yes | Unclear | Moderate |
| **Germany** | | | | | | | | | | | |
| Fuest | 2020-11-18 | Yes | Unclear | Yes | No | Unclear | Unclear | Yes | No | Unclear | Unclear |
| Robert Koch Institute | 2020-11-05 | No | Yes | Yes | No | Yes | Unclear | Yes | No | Unclear | Moderate |
| Armann | 2020-10-13 | Yes | Unclear | Yes | Yes | Yes | Yes | Yes | No | Unclear | Moderate |
| Thielecke | 2020-10-02 | Yes | Yes | Yes | Yes | Unclear | Unclear | Yes | No | Unclear | Moderate |
| Schwarz | 2020-09-01 | Yes | No | Yes | No | Unclear | Unclear | No | No | Unclear | High |
| Rauber | 2020-08-06 | No | No | No | Yes | Unclear | Yes | Yes | No | Unclear | High |
| Hoffmann | 2020-07-31 | Yes | No | No | Yes | Unclear | Yes | Yes | No | Unclear | High |
| Finkenzeller | 2020-07-29 | Yes | No | Yes | Yes | Unclear | Yes | Yes | No | Yes | Moderate |

| Article Author (Study Organization) | Sampling End Date (YMD) | Item 1: Appropriate sample frame* | Item 2: Probability sampling method | Item 3: Adequate sample size | Item 4: Subjects & setting described | Item 5: Good coverage of sample | Item 6: Sens > 90% & spec > 95% | Item 7: Same tests for all subjects | Item 8: Appropriate analysis | Item 9: Adequate response rate | Overall Risk of Bias† |
|---|---|---|---|---|---|---|---|---|---|---|---|
| Finkenzeller | 2020-07-29 | Unclear | Unclear | Yes | Yes | Unclear | Yes | Yes | No | No | High |
| Laub | 2020-07-22 | Yes | No | Yes | Yes | Unclear | Yes | Yes | No | Yes | Moderate |
| Brehm | 2020-07-17 | Yes | No | Yes | Yes | Yes | Yes | Yes | No | Unclear | Moderate |
| Mack | 2020-06-30 | No | No | Yes | Yes | Unclear | No | Yes | No | Unclear | High |
| Epstude | 2020-06-30 | No | No | No | Yes | Yes | Unclear | Yes | No | Unclear | High |
| Epstude | 2020-06-30 | No | No | No | Yes | Yes | Unclear | Yes | No | Unclear | High |
| Aziz | 2020-06-30 | No | No | Yes | Yes | No | Yes | Yes | No | Yes | High |
| Aziz | 2020-06-30 | No | No | No | Yes | No | Yes | Yes | No | Unclear | High |
| Bahrs | 2020-06-19 | No | No | Yes | Yes | Unclear | Yes | Yes | No | No | High |
| Herzberg | 2020-06-16 | No | No | Yes | Yes | Unclear | Yes | No | No | Yes | High |
| Runkel | 2020-06-15 | No | Unclear | Yes | Yes | Unclear | Unclear | Yes | No | Unclear | High |
| Santos-Hovener | 2020-06-09 | Yes | Yes | Yes | Yes | Yes | No | Yes | Yes | Yes | Low |
| Malfertheiner | 2020-06-07 | Yes | No | No | Unclear | Unclear | Unclear | Unclear | No | Unclear | High |
| Herrmann | 2020-06-04 | Unclear | Unclear | No | No | Unclear | Yes | Yes | No | Unclear | High |
| Fischer | 2020-06-03 | No | No | Yes | No | No | Yes | Yes | No | Unclear | High |
| Thieme | 2020-05-31 | Yes | No | No | Yes | Yes | Unclear | Yes | No | Yes | High |
| Erber | 2020-05-29 | No | No | Yes | Yes | Yes | Unclear | Yes | No | No | High |
| Weis | 2020-05-22 | Yes | No | Yes | Yes | Yes | Yes | Yes | No | Yes | Moderate |
| Choi | 2020-05-19 | Yes | Unclear | No | Yes | Unclear | Unclear | Yes | No | Unclear | High |
| Weis | 2020-05-16 | Yes | Yes | Yes | Yes | Yes | Yes | Yes | No | Yes | Moderate |
| Neumann | 2020-05-16 | Yes | Unclear | No | No | Unclear | Yes | Yes | No | Unclear | High |

| Article Author (Study Organization) | Sampling End Date (YMD) | Item 1: Appropriate sample frame* | Item 2: Probability sampling method | Item 3: Adequate sample size | Item 4: Subjects & setting described | Item 5: Good coverage of sample | Item 6: Sens > 90% & spec > 95% | Item 7: Same tests for all subjects | Item 8: Appropriate analysis | Item 9: Adequate response rate | Overall Risk of Bias† |
|---|---|---|---|---|---|---|---|---|---|---|---|
| Tonshoff | 2020-05-15 | Unclear | Unclear | Unclear | Unclear | Unclear | Unclear | Unclear | Unclear | Unclear | Moderate |
| Zollkau | 2020-05-13 | No | Unclear | No | Yes | Unclear | No | Yes | No | Unclear | High |
| Schmidt | 2020-04-30 | Yes | No | No | Yes | Unclear | Yes | Yes | No | Yes | High |
| Reisinger | 2020-04-22 | Yes | No | No | Yes | Unclear | Unclear | Yes | No | Unclear | High |
| Korth | 2020-04-21 | No | No | No | No | Unclear | Unclear | Yes | No | Unclear | High |
| Harsch | 2020-04-21 | Yes | No | No | No | Unclear | Yes | Yes | No | No | High |
| Behrens | 2020-04-17 | No | Unclear | No | No | Unclear | No | Yes | No | Unclear | High |
| Kern | 2020-04-16 | Yes | Unclear | Yes | Yes | Unclear | Yes | Yes | No | Unclear | High |
| Kraehling | 2020-04-14 | No | No | Yes | Yes | No | Yes | Yes | No | Unclear | High |
| Lackermair | 2020-04-06 | No | No | No | Yes | No | Unclear | Yes | No | Unclear | High |
| Streeck | 2020-04-06 | Yes | Yes | Yes | Yes | Unclear | Yes | Yes | Yes | Yes | Low |
| **Greece** | | | | | | | | | | | |
| Tsitsilonis | 2020-07-15 | Yes | No | Yes | Yes | Yes | Yes | Yes | Yes | Unclear | Moderate |
| Psichogiou | 2020-05-15 | No | No | Yes | Yes | Yes | No | Yes | Yes | Yes | Moderate |
| Vlachoyiannopoulosa | 2020-05-10 | Yes | Unclear | No | Yes | Unclear | No | Yes | No | Unclear | High |
| Bogogiannidou | 2020-04-30 | No | No | Yes | Yes | Yes | No | Yes | Yes | Unclear | Moderate |
| **Iceland** | | | | | | | | | | | |
| Gudbjartsson | 2020-06-12 | No | Yes | Yes | Unclear | Yes | Yes | Yes | No | Unclear | Moderate |
| **Ireland** | | | | | | | | | | | |
| Igoe | 2020-07-16 | Yes | Yes | Yes | No | Unclear | Yes | Yes | No | No | Moderate |
| **Israel** | | | | | | | | | | | |

| Article Author (Study Organization) | Sampling End Date (YMD) | Item 1: Appropriate sample frame* | Item 2: Probability sampling method | Item 3: Adequate sample size | Item 4: Subjects & setting described | Item 5: Good coverage of sample | Item 6: Sens > 90% & spec > 95% | Item 7: Same tests for all subjects | Item 8: Appropriate analysis | Item 9: Adequate response rate | Overall Risk of Bias† |
|---|---|---|---|---|---|---|---|---|---|---|---|
| Rabinowitz | 2020-08-15 | Yes | Yes | Yes | No | Yes | Unclear | Unclear | No | Unclear | Moderate |
| Efrati | 2020-06-02 | Yes | Yes | Yes | No | Yes | Unclear | Unclear | No | Unclear | Moderate |
| **Italy** | | | | | | | | | | | |
| Caponigro | 2020-10-19 | Unclear | Unclear | No | No | Unclear | Yes | Yes | No | Unclear | Unclear |
| Russo | 2020-08-30 | No | No | No | Yes | No | Yes | Yes | No | Unclear | High |
| Ralli | 2020-07-31 | Yes | No | No | Yes | Unclear | Unclear | Yes | No | Unclear | High |
| Italian Ministry of Health | 2020-07-15 | Yes | No | Yes | No | Unclear | Unclear | Unclear | No | Unclear | Unclear |
| Norsa | 2020-07-10 | No | No | No | Yes | Yes | Unclear | Yes | No | Yes | High |
| Cento | 2020-07-05 | No | No | Yes | Yes | No | Yes | Yes | No | Yes | Moderate |
| Medas | 2020-06-30 | No | No | No | Yes | Unclear | Unclear | Yes | No | Yes | High |
| Berte | 2020-06-15 | No | No | No | Yes | Unclear | Yes | Yes | No | Unclear | High |
| Berte | 2020-06-15 | No | No | No | Yes | Unclear | Yes | Yes | No | Unclear | High |
| Capasso | 2020-06-15 | Yes | No | No | Yes | Yes | No | Yes | No | Yes | High |
| Capasso | 2020-06-15 | No | No | Yes | Yes | Yes | No | Yes | No | Yes | Moderate |
| Capasso | 2020-06-15 | Yes | No | No | Yes | Yes | No | Yes | No | Yes | Moderate |
| Pagani | 2020-06-07 | Yes | Yes | Yes | Yes | Yes | Unclear | Yes | No | Unclear | Moderate |
| Cosma | 2020-06-04 | Yes | No | No | No | Unclear | Unclear | Yes | No | Unclear | High |
| Amante | 2020-06-03 | Yes | Unclear | Yes | No | Yes | Unclear | Unclear | No | Unclear | Moderate |
| Lorenzo | 2020-05-31 | Yes | No | No | No | Unclear | Unclear | Yes | No | Unclear | High |
| Lastrucci | 2020-05-31 | Yes | No | Yes | Yes | Yes | Yes | Yes | No | Yes | Moderate |

| Article Author (Study Organization) | Sampling End Date (YMD) | Item 1: Appropriate sample frame* | Item 2: Probability sampling method | Item 3: Adequate sample size | Item 4: Subjects & setting described | Item 5: Good coverage of sample | Item 6: Sens > 90% & spec > 95% | Item 7: Same tests for all subjects | Item 8: Appropriate analysis | Item 9: Adequate response rate | Overall Risk of Bias† |
|---|---|---|---|---|---|---|---|---|---|---|---|
| Papalini | 2020-05-31 | Unclear | Unclear | Yes | No | Unclear | No | Yes | No | Unclear | Unclear |
| Papalini | 2020-05-31 | No | No | No | Yes | Unclear | No | Yes | No | Unclear | High |
| Fiore | 2020-05-31 | No | No | Yes | Yes | Unclear | No | Yes | No | Yes | High |
| Sperotto | 2020-05-30 | Yes | No | No | Yes | Unclear | Yes | Yes | No | Unclear | High |
| Stellato | 2020-05-29 | No | No | No | No | No | Unclear | No | No | Unclear | High |
| Stellato | 2020-05-29 | No | No | Yes | No | No | Unclear | No | No | Unclear | High |
| Plebani | 2020-05-29 | Yes | Unclear | Yes | Yes | Yes | No | Yes | No | Unclear | High |
| Poletti | 2020-05-25 | Yes | No | Yes | Yes | Unclear | Yes | Yes | No | Yes | Moderate |
| Alessi | 2020-05-24 | Yes | No | Yes | Yes | No | No | Yes | No | No | High |
| Calcagno | 2020-05-20 | Yes | No | Yes | Yes | Unclear | Yes | Yes | No | Yes | Moderate |
| Carlo | 2020-05-18 | Yes | No | Yes | No | Unclear | Yes | Yes | No | Unclear | Moderate |
| Martella | 2020-05-17 | No | No | Yes | No | Unclear | Unclear | Unclear | No | Unclear | Unclear |
| Cavaliere | 2020-05-16 | Yes | No | No | No | Unclear | Unclear | Yes | No | Yes | High |
| Sandri | 2020-05-16 | Yes | No | Yes | Yes | Yes | Yes | Yes | No | Yes | Moderate |
| Bianchi | 2020-05-15 | No | Unclear | No | No | Unclear | Unclear | No | No | Unclear | Unclear |
| Bianchi | 2020-05-15 | No | Unclear | No | No | Unclear | Unclear | No | No | Unclear | Unclear |
| Stefanelli | 2020-05-15 | Yes | Yes | Yes | Yes | Yes | Yes | Yes | No | Yes | Low |
| Marra | 2020-05-11 | No | No | No | Yes | Unclear | Unclear | Yes | No | Unclear | High |
| Guerriero | 2020-05-08 | Yes | Yes | Yes | Yes | Unclear | Unclear | Yes | No | Yes | Moderate |
| Baracco | 2020-05-05 | No | No | Yes | Yes | Yes | Unclear | Yes | No | Yes | High |
| Baracco | 2020-05-05 | Yes | Yes | Yes | Yes | Yes | Unclear | Yes | No | Unclear | Moderate |

| Article Author (Study Organization) | Sampling End Date (YMD) | Item 1: Appropriate sample frame* | Item 2: Probability sampling method | Item 3: Adequate sample size | Item 4: Subjects & setting described | Item 5: Good coverage of sample | Item 6: Sens > 90% & spec > 95% | Item 7: Same tests for all subjects | Item 8: Appropriate analysis | Item 9: Adequate response rate | Overall Risk of Bias† |
|---|---|---|---|---|---|---|---|---|---|---|---|
| Denina | 2020-04-30 | Yes | No | No | Yes | Unclear | Unclear | No | No | Unclear | High |
| Zambelli | 2020-04-30 | Yes | No | No | Yes | Unclear | Unclear | Yes | No | Unclear | High |
| Vena | 2020-04-30 | No | No | Yes | Yes | Yes | Yes | No | No | Unclear | High |
| Carozzi | 2020-04-30 | Yes | No | Yes | No | Unclear | No | No | No | Unclear | High |
| Bontadi | 2020-04-29 | No | No | Yes | Yes | Unclear | Unclear | No | No | No | High |
| Landro | 2020-04-28 | Yes | Unclear | Yes | No | Unclear | Unclear | Unclear | No | Unclear | Unclear |
| Volta | 2020-04-27 | No | No | No | Yes | Yes | Unclear | Yes | No | Yes | High |
| Lahner | 2020-04-27 | No | Yes | Yes | No | Yes | No | Yes | No | Unclear | High |
| Landro | 2020-04-24 | Yes | Unclear | Yes | No | Unclear | Unclear | Unclear | No | Unclear | Unclear |
| Loconsole | 2020-04-21 | Yes | No | Yes | Yes | Unclear | No | Yes | No | Unclear | High |
| Cito | 2020-04-19 | Yes | Yes | Yes | Yes | Yes | Yes | Yes | No | Yes | Moderate |
| Paradiso | 2020-04-17 | No | No | Yes | Yes | Unclear | Yes | Yes | No | Yes | High |
| Sotgiu | 2020-04-16 | Yes | No | No | Yes | Yes | No | Yes | No | Unclear | High |
| Amendola | 2020-04-15 | No | No | Yes | No | Unclear | Unclear | Yes | No | Yes | High |
| Canetti | 2020-04-15 | No | No | No | Yes | Unclear | Unclear | Yes | No | Unclear | High |
| Valenti | 2020-04-08 | No | No | Yes | Yes | Unclear | Yes | Yes | No | Unclear | High |
| Percivalle | 2020-04-06 | No | Unclear | No | No | Yes | Yes | Yes | No | Unclear | High |
| Fusco | 2020-04-02 | No | No | No | Yes | Yes | Unclear | Yes | No | Unclear | High |
| Milani | 2020-03-31 | Yes | No | No | No | No | Unclear | Yes | No | Yes | High |
| Pancrazzi | 2020-03-21 | Yes | No | No | No | No | No | Yes | No | Unclear | High |
| Apolone | 2020-03-10 | No | No | Yes | Yes | Yes | Unclear | Yes | No | Unclear | High |

| Article Author (Study Organization) | Sampling End Date (YMD) | Item 1: Appropriate sample frame* | Item 2: Probability sampling method | Item 3: Adequate sample size | Item 4: Subjects & setting described | Item 5: Good coverage of sample | Item 6: Sens > 90% & spec > 95% | Item 7: Same tests for all subjects | Item 8: Appropriate analysis | Item 9: Adequate response rate | Overall Risk of Bias† |
|---|---|---|---|---|---|---|---|---|---|---|---|
| **Japan** | | | | | | | | | | | |
| Oka | 2020-10-30 | No | No | No | No | No | Yes | Yes | No | Unclear | High |
| Matthews | 2020-08-31 | No | No | Yes | No | Unclear | Yes | Yes | No | No | High |
| Hibino | 2020-08-25 | Yes | No | No | Yes | Yes | Unclear | Yes | No | No | High |
| Fukuda | 2020-08-21 | Yes | No | Yes | Yes | Yes | Unclear | Yes | No | Yes | Moderate |
| Hibino | 2020-07-30 | Yes | No | Yes | Yes | Unclear | Unclear | Yes | No | Unclear | Unclear |
| Nawa | 2020-07-05 | Yes | Yes | Yes | Yes | Yes | Yes | Yes | No | No | Moderate |
| Okuda | 2020-06-28 | No | Yes | Yes | No | Unclear | Unclear | Unclear | No | Unclear | Unclear |
| Nishida | 2020-06-19 | Yes | No | Yes | Yes | No | Yes | Yes | No | Yes | Moderate |
| Chong | 2020-06-15 | Yes | No | No | No | Yes | Unclear | Yes | No | No | High |
| Suda | 2020-05-31 | No | No | No | Yes | Yes | Yes | Yes | No | Yes | High |
| Nakamura | 2020-05-29 | Yes | Unclear | Yes | Yes | Unclear | Yes | Yes | No | Unclear | High |
| Kobashi | 2020-05-28 | No | Unclear | Yes | Yes | Yes | Unclear | Yes | No | Unclear | High |
| Takita | 2020-05-20 | No | No | Yes | Yes | Unclear | No | Yes | No | Unclear | High |
| Denyer | 2020-05-18 | Yes | No | Yes | No | Unclear | Unclear | Unclear | Unclear | Unclear | Unclear |
| Fujita | 2020-04-20 | No | No | No | Yes | Yes | Unclear | Unclear | No | Unclear | High |
| Doi | 2020-04-07 | No | Yes | Yes | No | Unclear | Yes | Yes | No | Unclear | High |
| Suzuki | 2020-04-03 | No | No | No | Yes | Unclear | Unclear | Yes | No | Unclear | High |
| Suda | 2020-03-31 | No | No | No | Yes | Yes | Yes | Yes | No | Yes | High |
| **Liechtenstein** | | | | | | | | | | | |
| Schaffner | 2020-10-06 | Unclear | Unclear | Yes | Yes | Unclear | Yes | Yes | No | Unclear | Unclear |

| Article Author (Study Organization) | Sampling End Date (YMD) | Item 1: Appropriate sample frame* | Item 2: Probability sampling method | Item 3: Adequate sample size | Item 4: Subjects & setting described | Item 5: Good coverage of sample | Item 6: Sens > 90% & spec > 95% | Item 7: Same tests for all subjects | Item 8: Appropriate analysis | Item 9: Adequate response rate | Overall Risk of Bias† |
|---|---|---|---|---|---|---|---|---|---|---|---|
| **Luxembourg** | | | | | | | | | | | |
| Snoeck | 2020-05-05 | Yes | Yes | Yes | Yes | Yes | No | Yes | No | Unclear | Moderate |
| **Monaco** | | | | | | | | | | | |
| Binacchi | 2020-06-16 | Unclear | No | Yes | No | Unclear | Unclear | Unclear | No | No | Unclear |
| **Netherlands** | | | | | | | | | | | |
| Vos | 2020-05-11 | Yes | Yes | No | Yes | Unclear | No | Yes | Yes | Unclear | Low |
| Vos | 2020-05-11 | Yes | Yes | Yes | Yes | Unclear | No | Yes | Yes | Unclear | Low |
| Slot | 2020-04-15 | No | Yes | Yes | No | Yes | Yes | Yes | No | Yes | Moderate |
| Westerhuis | 2020-04-07 | No | No | Yes | No | Unclear | Unclear | Yes | No | Unclear | High |
| Westerhuis | 2020-03-06 | No | No | Yes | No | Unclear | Unclear | Yes | No | Unclear | High |
| **New Zealand** | | | | | | | | | | | |
| Craigie | 2020-08-04 | Yes | No | Yes | Yes | Yes | No | No | No | Unclear | High |
| **Norway** | | | | | | | | | | | |
| Tunheim | 2020-05-17 | No | No | Yes | Yes | Yes | No | Yes | Yes | Unclear | Moderate |
| Trieu | 2020-04-09 | Yes | No | Yes | Yes | Yes | Unclear | Yes | No | Unclear | High |
| Kuwelker | 2020-04-04 | No | No | No | Yes | Unclear | Unclear | Yes | No | No | High |
| Cox | 2020-04-04 | Yes | No | No | No | Unclear | Unclear | Yes | No | Unclear | High |
| **Portugal** | | | | | | | | | | | |
| Figueiredo-Campos | 2020-07-10 | No | No | Yes | No | Unclear | Yes | Yes | No | Unclear | High |
| Figueiredo-Campos | 2020-07-10 | Yes | No | Yes | No | Unclear | Yes | Yes | No | Unclear | Moderate |
| Kislaya | 2020-07-08 | No | No | Yes | Yes | Yes | Yes | Yes | Yes | Unclear | Moderate |

| Article Author (Study Organization) | Sampling End Date (YMD) | Item 1: Appropriate sample frame* | Item 2: Probability sampling method | Item 3: Adequate sample size | Item 4: Subjects & setting described | Item 5: Good coverage of sample | Item 6: Sens > 90% & spec > 95% | Item 7: Same tests for all subjects | Item 8: Appropriate analysis | Item 9: Adequate response rate | Overall Risk of Bias† |
|---|---|---|---|---|---|---|---|---|---|---|---|
| Gonclaves | 2020-06-30 | Yes | No | Yes | Yes | Unclear | Yes | Yes | No | Unclear | Moderate |
| **Republic of Korea** | | | | | | | | | | | |
| KBS World | 2020-10-15 | Unclear | Unclear | Yes | No | Unclear | Unclear | Unclear | Unclear | Unclear | Unclear |
| KBS World | 2020-10-15 | Unclear | Unclear | Yes | No | Unclear | Unclear | Unclear | Unclear | Unclear | Unclear |
| Ji-Ho | 2020-07-09 | No | Unclear | Yes | No | Unclear | Unclear | Unclear | No | Unclear | Unclear |
| Song | 2020-06-05 | No | No | No | Yes | Unclear | No | Yes | No | Unclear | High |
| Noh | 2020-05-29 | No | No | Yes | Yes | Unclear | Yes | Yes | No | Unclear | Moderate |
| Ko | 2020-05-09 | Yes | No | No | Yes | Unclear | No | Yes | No | Unclear | High |
| Ko | 2020-05-09 | Yes | No | No | Yes | Unclear | No | Yes | No | Unclear | High |
| **Singapore** | | | | | | | | | | | |
| Kurohi | 2020-04-15 | Yes | Unclear | Yes | No | Unclear | Unclear | Unclear | No | Unclear | Unclear |
| Kurohi | 2020-04-15 | Yes | Unclear | Yes | No | Unclear | Unclear | Unclear | No | Unclear | Unclear |
| Ng | 2020-04-03 | Yes | Yes | Yes | Yes | Unclear | Yes | Yes | No | No | Moderate |
| **Spain** | | | | | | | | | | | |
| SavirÃ³n-Cornudella | 2020-08-31 | Yes | No | No | No | Unclear | No | Yes | No | Unclear | High |
| Garde-Noguera | 2020-06-30 | Yes | No | No | Yes | Unclear | Unclear | Yes | No | Unclear | High |
| Prados | 2020-06-26 | Yes | Unclear | Yes | No | Unclear | Unclear | Yes | No | Unclear | Unclear |
| Cabezon-Gutierrez | 2020-06-19 | No | No | No | Yes | No | Yes | Yes | No | Unclear | High |
| Camba | 2020-06-15 | Yes | No | No | Yes | Unclear | Yes | Yes | No | Unclear | High |
| Brotons | 2020-06-03 | Yes | No | Yes | Yes | Unclear | Unclear | Yes | No | Unclear | Moderate |
| Flores | 2020-06-03 | Yes | Yes | Yes | No | Yes | Unclear | Unclear | No | Yes | Moderate |

| Article Author (Study Organization) | Sampling End Date (YMD) | Item 1: Appropriate sample frame* | Item 2: Probability sampling method | Item 3: Adequate sample size | Item 4: Subjects & setting described | Item 5: Good coverage of sample | Item 6: Sens > 90% & spec > 95% | Item 7: Same tests for all subjects | Item 8: Appropriate analysis | Item 9: Adequate response rate | Overall Risk of Bias[†] |
|---|---|---|---|---|---|---|---|---|---|---|---|
| Perez-Garcia | 2020-05-30 | No | No | Yes | Yes | Unclear | No | Yes | No | Unclear | High |
| Fabre | 2020-05-29 | No | Unclear | No | No | No | Unclear | Yes | No | Unclear | High |
| Barallat | 2020-05-22 | Yes | No | Yes | Yes | Yes | Unclear | Yes | No | Yes | High |
| Vinuela | 2020-05-21 | Yes | No | No | Yes | Yes | Unclear | Yes | No | Yes | High |
| Soriano | 2020-05-16 | No | Yes | Yes | Yes | No | Unclear | Yes | No | Yes | High |
| Pollan | 2020-05-11 | Yes | Yes | Yes | Yes | Yes | Yes | Yes | Yes | Yes | Low |
| Villalain | 2020-05-10 | Yes | Yes | Yes | Yes | Yes | Unclear | Yes | No | Unclear | Moderate |
| Gras-Valenti | 2020-05-08 | No | No | Yes | No | No | Unclear | Unclear | No | Yes | Unclear |
| Crovetto | 2020-05-05 | No | No | Yes | No | No | Unclear | No | No | Unclear | High |
| Valdivia | 2020-04-30 | No | Unclear | Yes | No | Unclear | No | Yes | No | Unclear | High |
| Galan | 2020-04-27 | No | No | Yes | Yes | Unclear | Unclear | Yes | No | Yes | High |
| Olalla | 2020-04-25 | Yes | No | No | Yes | Yes | Unclear | Yes | No | Unclear | High |
| Montenegro | 2020-04-24 | Yes | Yes | No | Yes | Yes | Unclear | Unclear | No | No | High |
| Borraz | 2020-04-21 | No | No | No | Yes | Unclear | Yes | Yes | No | Unclear | High |
| Borraz | 2020-04-21 | No | No | No | Yes | Unclear | Yes | Yes | No | Unclear | High |
| Dacosta-Urbieta | 2020-04-16 | Yes | No | No | No | Unclear | Yes | Yes | No | Unclear | High |
| Martin | 2020-04-15 | Yes | No | Yes | Yes | Unclear | No | Yes | No | Yes | High |
| Fernandez | 2020-04-14 | No | No | Yes | Yes | Unclear | Unclear | Yes | No | Yes | High |
| **Sweden** | | | | | | | | | | | |
| Dopico | 2020-12-11 | No | Yes | Yes | No | Unclear | Unclear | Yes | No | Unclear | High |
| Rashid-Abdi | 2020-08-19 | Yes | No | No | Yes | Yes | Yes | Yes | No | Unclear | High |

| Article Author (Study Organization) | Sampling End Date (YMD) | Item 1: Appropriate sample frame* | Item 2: Probability sampling method | Item 3: Adequate sample size | Item 4: Subjects & setting described | Item 5: Good coverage of sample | Item 6: Sens > 90% & spec > 95% | Item 7: Same tests for all subjects | Item 8: Appropriate analysis | Item 9: Adequate response rate | Overall Risk of Bias† |
|---|---|---|---|---|---|---|---|---|---|---|---|
| Laursen | 2020-08-10 | No | No | Yes | Yes | Yes | Unclear | Yes | No | Unclear | High |
| The Local | 2020-07-22 | Yes | No | Yes | No | Unclear | Unclear | Unclear | No | Unclear | High |
| Region | 2020-07-15 | Yes | No | Yes | No | Unclear | Unclear | Unclear | No | Unclear | High |
| Lidstrom | 2020-06-25 | Yes | Yes | Yes | Yes | Yes | Unclear | Yes | No | Yes | Moderate |
| Lundkvist | 2020-06-18 | Yes | Yes | No | Yes | Unclear | Yes | Yes | No | Unclear | High |
| Lundkvist | 2020-06-18 | Yes | Yes | No | Yes | Unclear | Yes | Yes | No | Unclear | High |
| Hassan | 2020-06-17 | No | No | No | Yes | No | Yes | Yes | No | Yes | High |
| Swedish Public Health Agency | 2020-06-14 | No | No | Yes | No | Unclear | Yes | Yes | No | Unclear | High |
| Swedish Public Health Agency | 2020-06-07 | No | No | Yes | No | Unclear | Yes | Yes | No | Unclear | High |
| Swedish Public Health Agency | 2020-05-31 | No | No | Yes | No | Unclear | Yes | Yes | No | Unclear | High |
| Swedish Public Health Agency | 2020-05-24 | No | No | Yes | No | Unclear | Yes | Yes | No | Unclear | High |
| Roxhed | 2020-05-21 | Yes | Yes | Yes | Yes | Unclear | Yes | Yes | No | No | Moderate |
| Swedish Public Health Agency | 2020-05-17 | No | No | Yes | No | Unclear | Yes | Yes | No | Unclear | High |
| Swedish Public Health Agency | 2020-05-10 | No | No | Yes | No | Unclear | Yes | Yes | No | Unclear | High |
| Rudberg | 2020-05-08 | Yes | No | Yes | Yes | Yes | Yes | Yes | No | No | Moderate |
| Swedish Public Health Agency | 2020-05-03 | No | No | Yes | No | Unclear | Yes | Yes | No | Unclear | High |
| Swedish Public Health Agency | 2020-04-26 | No | No | Yes | No | Unclear | Yes | Yes | No | Unclear | High |

| Article Author (Study Organization) | Sampling End Date (YMD) | Item 1: Appropriate sample frame* | Item 2: Probability sampling method | Item 3: Adequate sample size | Item 4: Subjects & setting described | Item 5: Good coverage of sample | Item 6: Sens > 90% & spec > 95% | Item 7: Same tests for all subjects | Item 8: Appropriate analysis | Item 9: Adequate response rate | Overall Risk of Bias† |
|---|---|---|---|---|---|---|---|---|---|---|---|
| Lindahl | 2020-04-20 | Yes | No | Yes | No | Unclear | Yes | Yes | No | Unclear | High |
| **Switzerland** | | | | | | | | | | | |
| Ulyte | 2020-11-19 | Yes | Yes | Yes | Yes | Yes | Yes | Yes | Yes | No | Low |
| Anker | 2020-10-14 | Yes | Yes | No | Yes | Yes | Yes | Unclear | Yes | No | Moderate |
| Kahlert | 2020-08-15 | No | No | Yes | Yes | Yes | Unclear | Yes | No | No | Moderate |
| Universita della Svizzera Italiana | 2020-07-31 | Unclear | Yes | Yes | No | Unclear | Unclear | Unclear | No | Unclear | Unclear |
| Emmenegger | 2020-07-15 | No | No | Yes | Yes | Unclear | Yes | Yes | No | Unclear | Moderate |
| Emmenegger | 2020-07-15 | No | No | Yes | Yes | Unclear | Yes | Yes | No | Unclear | Moderate |
| Ulyte | 2020-07-09 | Yes | Yes | No | No | Unclear | Yes | Yes | Yes | Unclear | Moderate |
| Bi | 2020-06-30 | Yes | No | Yes | Yes | No | Yes | Yes | No | No | Moderate |
| Richard | 2020-06-30 | Yes | Yes | Yes | Yes | No | Yes | Yes | Yes | No | Low |
| Fenwick | 2020-06-27 | Yes | Unclear | No | No | Unclear | Yes | Yes | No | Unclear | High |
| Fenwick | 2020-06-27 | Yes | Yes | No | No | Unclear | Yes | Yes | No | Unclear | High |
| Dupraz | 2020-06-27 | Yes | Yes | No | Yes | Unclear | Yes | Yes | Yes | Unclear | Moderate |
| Dupraz | 2020-06-27 | Yes | Yes | No | Yes | Unclear | Yes | Yes | Yes | Unclear | Moderate |
| Posfay-Barbe | 2020-04-30 | No | No | No | Yes | Unclear | Unclear | Yes | No | Unclear | High |
| Emmenegger | 2020-04-30 | No | No | Yes | Yes | Unclear | Yes | Yes | No | Unclear | Moderate |
| Emmenegger | 2020-04-30 | No | No | Yes | Yes | Unclear | Yes | Yes | No | Unclear | Moderate |
| Kohler | 2020-04-03 | Yes | No | Yes | Yes | Yes | Unclear | No | No | Unclear | High |
| Emmenegger | 2020-03-31 | No | No | Yes | Yes | Unclear | Yes | Yes | No | Unclear | Moderate |

| Article Author (Study Organization) | Sampling End Date (YMD) | Item 1: Appropriate sample frame* | Item 2: Probability sampling method | Item 3: Adequate sample size | Item 4: Subjects & setting described | Item 5: Good coverage of sample | Item 6: Sens > 90% & spec > 95% | Item 7: Same tests for all subjects | Item 8: Appropriate analysis | Item 9: Adequate response rate | Overall Risk of Bias† |
|---|---|---|---|---|---|---|---|---|---|---|---|
| **The United Kingdom** | | | | | | | | | | | |
| Office for National Statistics | 2021-01-14 | Yes | Yes | Yes | No | Unclear | Yes | Yes | No | Unclear | Moderate |
| Office for National Statistics | 2021-01-14 | Yes | Yes | No | No | Unclear | Yes | Yes | No | Unclear | High |
| Office for National Statistics | 2021-01-14 | Yes | Yes | Yes | No | Unclear | Yes | Yes | No | Unclear | Moderate |
| Office for National Statistics | 2021-01-14 | Yes | Yes | Yes | No | Unclear | Yes | Yes | No | Unclear | Moderate |
| Office for National Statistics | 2020-12-17 | Yes | Yes | Yes | No | Unclear | Yes | Yes | No | Unclear | Moderate |
| Office for National Statistics | 2020-12-17 | Yes | Yes | No | No | Unclear | Yes | Yes | No | Unclear | High |
| Office for National Statistics | 2020-12-17 | Yes | Yes | Yes | No | Unclear | Yes | Yes | No | Unclear | Moderate |
| Office for National Statistics | 2020-12-17 | Yes | Yes | Yes | No | Unclear | Yes | Yes | No | Unclear | Moderate |
| Taubel | 2020-12-02 | No | No | Yes | Yes | Unclear | Unclear | Yes | No | Unclear | High |
| Lumley | 2020-11-30 | Yes | No | Yes | Yes | Unclear | Yes | Yes | No | Unclear | Moderate |
| Office for National Statistics | 2020-11-19 | Yes | Yes | No | No | Unclear | Yes | Yes | No | Unclear | High |
| Office for National Statistics | 2020-11-19 | Yes | Yes | No | No | Unclear | Yes | Yes | No | Unclear | High |
| Office for National Statistics | 2020-11-19 | Yes | Yes | No | No | Unclear | Yes | Yes | No | Unclear | High |
| Office for National Statistics | 2020-11-19 | Yes | Yes | Yes | No | Unclear | Yes | Yes | No | Unclear | Moderate |

| Article Author (Study Organization) | Sampling End Date (YMD) | Item 1: Appropriate sample frame* | Item 2: Probability sampling method | Item 3: Adequate sample size | Item 4: Subjects & setting described | Item 5: Good coverage of sample | Item 6: Sens > 90% & spec > 95% | Item 7: Same tests for all subjects | Item 8: Appropriate analysis | Item 9: Adequate response rate | Overall Risk of Bias† |
|---|---|---|---|---|---|---|---|---|---|---|---|
| Office for National Statistics | 2020-10-22 | Yes | Yes | Yes | No | Unclear | Yes | Yes | No | Unclear | Moderate |
| Office for National Statistics | 2020-10-22 | Yes | Yes | No | No | Unclear | Yes | Yes | No | Unclear | High |
| Office for National Statistics | 2020-10-22 | Yes | Yes | No | No | Unclear | Yes | Yes | No | Unclear | High |
| Office for National Statistics | 2020-10-22 | Yes | Yes | No | No | Unclear | Yes | Yes | No | Unclear | High |
| Ward | 2020-09-28 | Yes | Yes | Yes | Yes | Unclear | No | Yes | Yes | No | Moderate |
| Office for National Statistics | 2020-09-24 | Yes | Yes | No | No | Unclear | Yes | Yes | No | Unclear | High |
| Office for National Statistics | 2020-09-24 | Yes | Yes | Yes | No | Unclear | Yes | Yes | No | Unclear | Moderate |
| Public Health England | 2020-09-18 | No | Yes | Yes | No | Unclear | No | Yes | Yes | Unclear | Moderate |
| Abo-Leyah | 2020-09-02 | No | Yes | No | No | Yes | Unclear | Yes | No | Unclear | High |
| Abo-Leyah | 2020-09-02 | Yes | No | Yes | Yes | Yes | Unclear | Yes | No | Unclear | High |
| Public Health England | 2020-08-30 | No | Yes | Yes | No | Unclear | No | Yes | Yes | Unclear | Moderate |
| Office for National Statistics | 2020-08-27 | Yes | Yes | Yes | No | Unclear | Yes | Yes | No | Unclear | Moderate |
| Harris | 2020-08-20 | Yes | No | Yes | Yes | Unclear | Unclear | No | No | Unclear | Moderate |
| Public Health England | 2020-08-16 | No | Yes | Yes | No | Yes | Yes | Yes | No | Unclear | Moderate |
| Public Health England | 2020-08-16 | No | Yes | Yes | No | Yes | Yes | Yes | Yes | Unclear | Low |
| Roarty | 2020-08-15 | Yes | No | Yes | No | Unclear | No | Yes | No | Unclear | Moderate |

| Article Author (Study Organization) | Sampling End Date (YMD) | Item 1: Appropriate sample frame* | Item 2: Probability sampling method | Item 3: Adequate sample size | Item 4: Subjects & setting described | Item 5: Good coverage of sample | Item 6: Sens > 90% & spec > 95% | Item 7: Same tests for all subjects | Item 8: Appropriate analysis | Item 9: Adequate response rate | Overall Risk of Bias† |
|---|---|---|---|---|---|---|---|---|---|---|---|
| Ward | 2020-08-13 | Yes | Yes | Yes | Yes | Unclear | No | Yes | Yes | No | Moderate |
| Public Health Scotland | 2020-08-09 | No | No | Yes | Yes | Yes | No | Yes | Yes | Unclear | Moderate |
| Public Health Scotland | 2020-08-09 | No | No | Yes | Yes | Yes | No | Yes | No | Unclear | Moderate |
| Cooper | 2020-08-07 | Yes | No | Yes | Yes | Unclear | Yes | Yes | No | Yes | Moderate |
| Public Health England | 2020-08-05 | Yes | Yes | No | No | Yes | Yes | Yes | Yes | Yes | Moderate |
| Black | 2020-08-03 | No | No | No | Yes | Unclear | Unclear | Yes | No | Unclear | High |
| Office for National Statistics | 2020-07-30 | Yes | Yes | Yes | No | Unclear | Yes | Yes | No | Unclear | Moderate |
| Public Health England | 2020-07-28 | No | Yes | Yes | No | Unclear | No | Yes | Yes | Unclear | Moderate |
| Claudiani | 2020-07-27 | Yes | No | No | Yes | Yes | Unclear | Yes | No | Unclear | High |
| Ward | 2020-07-13 | Yes | Yes | Yes | Yes | No | No | Yes | Yes | Yes | Low |
| Martin | 2020-07-13 | Yes | No | Yes | Yes | Yes | Yes | Yes | No | Yes | Moderate |
| Hanrath | 2020-07-06 | Yes | No | Yes | Yes | Unclear | Unclear | Yes | No | Yes | High |
| Prendecki | 2020-07-03 | No | No | Yes | Yes | Unclear | Unclear | Yes | No | Unclear | High |
| Waterfield | 2020-07-03 | Yes | No | Yes | Yes | Yes | No | Yes | No | Unclear | High |
| Office for National Statistics | 2020-07-02 | Yes | Yes | Yes | No | Unclear | Yes | Yes | No | Unclear | Moderate |
| Rowland | 2020-06-30 | No | No | No | Yes | No | Yes | Yes | No | Unclear | High |
| Public Health England | 2020-06-30 | No | Yes | Yes | No | Unclear | Yes | Yes | Yes | Unclear | Moderate |
| Graham | 2020-06-30 | No | No | No | No | Unclear | Yes | Yes | No | Unclear | High |

| Article Author (Study Organization) | Sampling End Date (YMD) | Item 1: Appropriate sample frame* | Item 2: Probability sampling method | Item 3: Adequate sample size | Item 4: Subjects & setting described | Item 5: Good coverage of sample | Item 6: Sens > 90% & spec > 95% | Item 7: Same tests for all subjects | Item 8: Appropriate analysis | Item 9: Adequate response rate | Overall Risk of Bias† |
|---|---|---|---|---|---|---|---|---|---|---|---|
| Davis | 2020-06-30 | No | No | Yes | Yes | No | Yes | Yes | No | No | High |
| Public Health England | 2020-06-28 | No | Yes | Yes | No | Unclear | No | Yes | Yes | Unclear | Moderate |
| Gulraj | 2020-06-17 | Yes | Unclear | Yes | No | Unclear | Unclear | Unclear | No | Unclear | Unclear |
| Lumley | 2020-06-15 | Yes | Yes | Yes | Yes | Unclear | Yes | Yes | Unclear | Yes | Moderate |
| Jones | 2020-06-15 | Yes | No | Yes | Yes | Unclear | Unclear | No | No | No | High |
| Favara | 2020-06-15 | Yes | No | No | Yes | Unclear | No | Yes | No | Unclear | High |
| Pallett | 2020-06-12 | Yes | No | No | Yes | No | No | Yes | No | No | High |
| Baxendale | 2020-06-10 | No | No | Yes | Yes | Unclear | Yes | Yes | No | Unclear | High |
| Arif | 2020-06-08 | No | No | No | Yes | Yes | Unclear | Unclear | No | Unclear | High |
| Razvi | 2020-06-08 | Yes | No | Yes | Yes | No | Unclear | Yes | No | No | High |
| Favara | 2020-06-07 | Yes | Yes | No | No | Unclear | No | Yes | No | Yes | High |
| Bampoe | 2020-06-05 | Yes | Unclear | No | Yes | No | Yes | Yes | No | Unclear | High |
| Grant | 2020-06-05 | Yes | No | Yes | No | Yes | Yes | Yes | No | No | High |
| McCafferty | 2020-06-04 | Yes | No | Yes | No | No | Yes | Yes | No | Unclear | High |
| Office for National Statistics | 2020-06-04 | Yes | Yes | Yes | No | Unclear | Yes | Yes | No | Unclear | Moderate |
| Wells | 2020-06-02 | No | Unclear | No | Yes | Unclear | Yes | Yes | No | No | High |
| Ladhani | 2020-05-31 | Yes | No | No | Yes | Yes | Unclear | Yes | No | Unclear | High |
| Public Health England | 2020-05-31 | No | Yes | Yes | No | Unclear | No | Yes | Yes | Unclear | Moderate |
| Public Health England | 2020-05-31 | Yes | Yes | Yes | No | Yes | Yes | Yes | No | Unclear | Moderate |

| Article Author (Study Organization) | Sampling End Date (YMD) | Item 1: Appropriate sample frame* | Item 2: Probability sampling method | Item 3: Adequate sample size | Item 4: Subjects & setting described | Item 5: Good coverage of sample | Item 6: Sens > 90% & spec > 95% | Item 7: Same tests for all subjects | Item 8: Appropriate analysis | Item 9: Adequate response rate | Overall Risk of Bias† |
|---|---|---|---|---|---|---|---|---|---|---|---|
| Khalil | 2020-05-28 | No | No | No | No | Unclear | Yes | Yes | No | Unclear | High |
| Hughes | 2020-05-24 | No | No | Yes | Yes | Unclear | Yes | Yes | Yes | Unclear | Moderate |
| Thompson | 2020-05-18 | No | Yes | Yes | Yes | No | Yes | Yes | No | Unclear | Moderate |
| Poulikakos | 2020-05-06 | Yes | No | No | No | No | Unclear | Yes | No | Unclear | High |
| Public Health England | 2020-04-30 | Yes | Yes | Yes | No | Yes | Yes | Yes | Yes | Unclear | Moderate |
| Public Health England | 2020-04-30 | Yes | Yes | No | No | Yes | Yes | Yes | Yes | Unclear | Moderate |
| Public Health England | 2020-04-26 | No | Yes | Yes | No | Unclear | No | Yes | Yes | Unclear | Moderate |
| Shields | 2020-04-25 | Yes | No | No | Yes | Yes | Yes | Yes | No | Unclear | High |
| Public Health England | 2020-04-24 | No | Yes | Yes | No | Unclear | No | Yes | Yes | Unclear | Moderate |
| Sikora | 2020-04-24 | Yes | No | No | No | Unclear | Unclear | Yes | No | Unclear | High |
| Public Health England | 2020-04-20 | No | Yes | Yes | No | Unclear | No | Yes | Yes | Unclear | Moderate |
| Public Health England | 2020-04-16 | No | Yes | Yes | No | Unclear | No | Yes | Yes | Unclear | Moderate |
| Public Health England | 2020-04-13 | No | Yes | Yes | No | Unclear | No | Yes | Yes | Unclear | Moderate |
| Ladhani | 2020-04-13 | Yes | No | No | Yes | Unclear | Unclear | Yes | No | Yes | High |
| Houlihan | 2020-04-08 | Yes | No | No | Yes | Yes | Unclear | Yes | No | Unclear | High |
| Public Health England | 2020-04-03 | No | Yes | Yes | No | Unclear | No | Yes | Yes | Unclear | Moderate |
| Public Health England | 2020-03-31 | Yes | Yes | No | No | Yes | Yes | Yes | Yes | Unclear | Moderate |

| Article Author (Study Organization) | Sampling End Date (YMD) | Item 1: Appropriate sample frame* | Item 2: Probability sampling method | Item 3: Adequate sample size | Item 4: Subjects & setting described | Item 5: Good coverage of sample | Item 6: Sens > 90% & spec > 95% | Item 7: Same tests for all subjects | Item 8: Appropriate analysis | Item 9: Adequate response rate | Overall Risk of Bias† |
|---|---|---|---|---|---|---|---|---|---|---|---|
| Public Health England | 2020-03-31 | Yes | Yes | No | No | Yes | Yes | Yes | Yes | Unclear | Moderate |
| Public Health England | 2020-03-27 | No | Yes | Yes | No | Unclear | No | Yes | Yes | Unclear | Moderate |
| **United States of America** | | | | | | | | | | | |
| Maleck | 2020-12-14 | Yes | Yes | Yes | No | Unclear | Unclear | Yes | No | No | Moderate |
| Craver | 2020-12-09 | Yes | Yes | Yes | No | No | Unclear | Unclear | Unclear | Unclear | Unclear |
| Stout | 2020-12-06 | No | No | Yes | Yes | Unclear | Yes | Yes | No | Unclear | High |
| Jolley | 2020-11-30 | Yes | No | Yes | No | Unclear | Unclear | Unclear | Unclear | Unclear | Unclear |
| Coffman | 2020-11-16 | Yes | Yes | Yes | No | Unclear | Unclear | Unclear | Unclear | No | Unclear |
| Coffman | 2020-11-16 | Yes | No | Yes | No | Unclear | Unclear | Unclear | Unclear | Unclear | Unclear |
| Godbout | 2020-10-02 | No | No | Yes | Yes | Yes | Unclear | Yes | No | Yes | High |
| Bajema | 2020-09-24 | No | No | Yes | No | Unclear | Yes | No | Yes | Unclear | Moderate |
| Bajema | 2020-09-24 | No | No | Yes | No | Unclear | Yes | No | Yes | Unclear | Moderate |
| Bajema | 2020-09-24 | No | No | Yes | No | Unclear | Yes | No | Yes | Unclear | Moderate |
| Bajema | 2020-09-24 | No | No | Yes | No | Unclear | Yes | No | Yes | Unclear | Moderate |
| Bajema | 2020-09-24 | No | No | Yes | No | Unclear | Yes | No | Yes | Unclear | Moderate |
| Bajema | 2020-09-24 | No | No | Yes | No | Unclear | Yes | No | Yes | Unclear | Moderate |
| Bajema | 2020-09-24 | No | No | No | No | Unclear | Yes | No | Yes | Unclear | High |
| Bajema | 2020-09-24 | No | No | Yes | No | Unclear | Yes | No | Yes | Unclear | Moderate |
| Bajema | 2020-09-24 | No | No | Yes | No | Unclear | Yes | No | Yes | Unclear | Moderate |
| Bajema | 2020-09-24 | No | No | No | No | Unclear | Yes | No | Yes | Unclear | High |

| Article Author (Study Organization) | Sampling End Date (YMD) | Item 1: Appropriate sample frame* | Item 2: Probability sampling method | Item 3: Adequate sample size | Item 4: Subjects & setting described | Item 5: Good coverage of sample | Item 6: Sens > 90% & spec > 95% | Item 7: Same tests for all subjects | Item 8: Appropriate analysis | Item 9: Adequate response rate | Overall Risk of Bias† |
|---|---|---|---|---|---|---|---|---|---|---|---|
| Bajema | 2020-09-24 | No | No | Yes | No | Unclear | Yes | No | Yes | Unclear | Moderate |
| Bajema | 2020-09-24 | No | No | Yes | No | Unclear | Yes | No | Yes | Unclear | Moderate |
| Bajema | 2020-09-24 | No | No | No | No | Unclear | Yes | No | Yes | Unclear | High |
| Bajema | 2020-09-23 | No | No | Yes | No | Unclear | Yes | No | Yes | Unclear | Moderate |
| Bajema | 2020-09-23 | No | No | Yes | No | Unclear | Yes | No | Yes | Unclear | Moderate |
| Bajema | 2020-09-23 | No | No | Yes | No | Unclear | Yes | No | Yes | Unclear | Moderate |
| Bajema | 2020-09-23 | No | No | Yes | No | Unclear | Yes | No | Yes | Unclear | Moderate |
| Bajema | 2020-09-23 | No | No | No | No | Unclear | Yes | No | Yes | Unclear | High |
| Bajema | 2020-09-23 | No | No | Yes | No | Unclear | Yes | No | Yes | Unclear | Moderate |
| Bajema | 2020-09-23 | No | No | Yes | No | Unclear | Yes | No | Yes | Unclear | Moderate |
| Bajema | 2020-09-23 | No | No | No | No | Unclear | Yes | No | Yes | Unclear | High |
| Bajema | 2020-09-23 | No | No | Yes | No | Unclear | Yes | No | Yes | Unclear | Moderate |
| Bajema | 2020-09-22 | No | No | No | No | Unclear | Yes | No | Yes | Unclear | High |
| Bajema | 2020-09-22 | No | No | Yes | No | Unclear | Yes | No | Yes | Unclear | Moderate |
| Bajema | 2020-09-22 | No | No | Yes | No | Unclear | Yes | No | Yes | Unclear | Moderate |
| Bajema | 2020-09-22 | No | No | Yes | No | Unclear | Yes | No | Yes | Unclear | Moderate |
| Bajema | 2020-09-22 | No | No | Yes | No | Unclear | Yes | No | Yes | Unclear | Moderate |
| Bajema | 2020-09-22 | No | No | Yes | No | Unclear | Yes | No | Yes | Unclear | Moderate |
| Bajema | 2020-09-22 | No | No | Yes | No | Unclear | Yes | No | Yes | Unclear | Moderate |
| Fountain Hills Times | 2020-09-21 | Yes | Yes | No | No | Unclear | Unclear | Unclear | Unclear | Unclear | Unclear |
| Bajema | 2020-09-19 | No | No | Yes | No | Unclear | Yes | No | Yes | Unclear | Moderate |

| Article Author (Study Organization) | Sampling End Date (YMD) | Item 1: Appropriate sample frame* | Item 2: Probability sampling method | Item 3: Adequate sample size | Item 4: Subjects & setting described | Item 5: Good coverage of sample | Item 6: Sens > 90% & spec > 95% | Item 7: Same tests for all subjects | Item 8: Appropriate analysis | Item 9: Adequate response rate | Overall Risk of Bias† |
|---|---|---|---|---|---|---|---|---|---|---|---|
| Bajema | 2020-09-19 | No | No | Yes | No | Unclear | Yes | No | Yes | Unclear | Moderate |
| Bajema | 2020-09-18 | No | No | Yes | No | Unclear | Yes | No | Yes | Unclear | Moderate |
| Bajema | 2020-09-18 | No | No | Yes | No | Unclear | Yes | No | Yes | Unclear | Moderate |
| Bajema | 2020-09-18 | No | No | Yes | No | Unclear | Yes | No | Yes | Unclear | Moderate |
| Bajema | 2020-09-18 | No | No | Yes | No | Unclear | Yes | No | Yes | Unclear | Moderate |
| Bajema | 2020-09-18 | No | No | Yes | No | Unclear | Yes | No | Yes | Unclear | Moderate |
| Bajema | 2020-09-18 | No | No | Yes | No | Unclear | Yes | No | Yes | Unclear | Moderate |
| Bajema | 2020-09-18 | No | No | Yes | No | Unclear | Yes | No | Yes | Unclear | Moderate |
| Bajema | 2020-09-18 | No | No | Yes | No | Unclear | Yes | No | Yes | Unclear | Moderate |
| Bajema | 2020-09-18 | No | No | Yes | No | Unclear | Yes | No | Yes | Unclear | Moderate |
| Bajema | 2020-09-18 | No | No | Yes | No | Unclear | Yes | No | Yes | Unclear | Moderate |
| Bajema | 2020-09-18 | No | No | Yes | No | Unclear | Yes | No | Yes | Unclear | Moderate |
| Bajema | 2020-09-17 | No | No | Yes | No | Unclear | Yes | No | Yes | Unclear | Moderate |
| Bajema | 2020-09-17 | No | No | Yes | No | Unclear | Yes | No | Yes | Unclear | Moderate |
| Bajema | 2020-09-17 | No | No | Yes | No | Unclear | Yes | No | Yes | Unclear | Moderate |
| Bajema | 2020-09-16 | No | No | Yes | No | Unclear | Yes | No | Yes | Unclear | Moderate |
| Bajema | 2020-09-16 | No | No | Yes | No | Unclear | Yes | No | Yes | Unclear | Moderate |
| Bajema | 2020-09-15 | No | No | Yes | No | Unclear | Yes | No | Yes | Unclear | Moderate |
| Bajema | 2020-09-15 | No | No | Yes | No | Unclear | Yes | No | Yes | Unclear | Moderate |
| Bajema | 2020-09-14 | No | No | Yes | No | Unclear | Yes | No | Yes | Unclear | Moderate |
| Bajema | 2020-09-12 | No | No | Yes | No | Unclear | Yes | No | Yes | Unclear | Moderate |

| Article Author (Study Organization) | Sampling End Date (YMD) | Item 1: Appropriate sample frame* | Item 2: Probability sampling method | Item 3: Adequate sample size | Item 4: Subjects & setting described | Item 5: Good coverage of sample | Item 6: Sens > 90% & spec > 95% | Item 7: Same tests for all subjects | Item 8: Appropriate analysis | Item 9: Adequate response rate | Overall Risk of Bias† |
|---|---|---|---|---|---|---|---|---|---|---|---|
| Bajema | 2020-09-11 | No | No | Yes | No | Unclear | Yes | No | Yes | Unclear | Moderate |
| Bajema | 2020-09-10 | No | No | Yes | No | Unclear | Yes | No | Yes | Unclear | Moderate |
| Bajema | 2020-09-10 | No | No | Yes | No | Unclear | Yes | No | Yes | Unclear | Moderate |
| Bajema | 2020-09-10 | No | No | Yes | No | Unclear | Yes | No | Yes | Unclear | Moderate |
| Bajema | 2020-09-10 | No | No | Yes | No | Unclear | Yes | No | Yes | Unclear | Moderate |
| Bajema | 2020-09-10 | No | No | Yes | No | Unclear | Yes | No | Yes | Unclear | Moderate |
| Bajema | 2020-09-10 | No | No | Yes | No | Unclear | Yes | No | Yes | Unclear | Moderate |
| Bajema | 2020-09-10 | No | No | Yes | No | Unclear | Yes | No | Yes | Unclear | Moderate |
| Bajema | 2020-09-10 | No | No | Yes | No | Unclear | Yes | No | Yes | Unclear | Moderate |
| Bajema | 2020-09-10 | No | No | Yes | No | Unclear | Yes | No | Yes | Unclear | Moderate |
| Bajema | 2020-09-10 | No | No | Yes | No | Unclear | Yes | No | Yes | Unclear | Moderate |
| Bajema | 2020-09-10 | No | No | Yes | No | Unclear | Yes | No | Yes | Unclear | Moderate |
| Bajema | 2020-09-10 | No | No | Yes | No | Unclear | Yes | No | Yes | Unclear | Moderate |
| Bajema | 2020-09-10 | No | No | Yes | No | Unclear | Yes | No | Yes | Unclear | Moderate |
| Bajema | 2020-09-10 | No | No | No | No | Unclear | Yes | No | Yes | Unclear | High |
| Bajema | 2020-09-10 | No | No | No | No | Unclear | Yes | No | Yes | Unclear | High |
| Bajema | 2020-09-10 | No | No | Yes | No | Unclear | Yes | No | Yes | Unclear | Moderate |
| Bajema | 2020-09-10 | No | No | Yes | No | Unclear | Yes | No | Yes | Unclear | Moderate |
| Bajema | 2020-09-10 | No | No | Yes | No | Unclear | Yes | No | Yes | Unclear | Moderate |
| Bajema | 2020-09-10 | No | No | Yes | No | Unclear | Yes | No | Yes | Unclear | Moderate |
| Jimenez-Kurlander | 2020-09-10 | Yes | No | No | Yes | Unclear | Unclear | Unclear | No | Unclear | High |

| Article Author (Study Organization) | Sampling End Date (YMD) | Item 1: Appropriate sample frame* | Item 2: Probability sampling method | Item 3: Adequate sample size | Item 4: Subjects & setting described | Item 5: Good coverage of sample | Item 6: Sens > 90% & spec > 95% | Item 7: Same tests for all subjects | Item 8: Appropriate analysis | Item 9: Adequate response rate | Overall Risk of Bias† |
|---|---|---|---|---|---|---|---|---|---|---|---|
| Bajema | 2020-09-09 | No | No | Yes | No | Unclear | Yes | No | Yes | Unclear | Moderate |
| Bajema | 2020-09-09 | No | No | Yes | No | Unclear | Yes | No | Yes | Unclear | Moderate |
| Bajema | 2020-09-09 | No | No | Yes | No | Unclear | Yes | No | Yes | Unclear | Moderate |
| Bajema | 2020-09-09 | No | No | Yes | No | Unclear | Yes | No | Yes | Unclear | Moderate |
| Bajema | 2020-09-09 | No | No | Yes | No | Unclear | Yes | No | Yes | Unclear | Moderate |
| Bajema | 2020-09-09 | No | No | No | No | Unclear | Yes | No | Yes | Unclear | High |
| Bajema | 2020-09-09 | No | No | Yes | No | Unclear | Yes | No | Yes | Unclear | Moderate |
| Bajema | 2020-09-09 | No | No | No | No | Unclear | Yes | No | Yes | Unclear | High |
| Bajema | 2020-09-09 | No | No | Yes | No | Unclear | Yes | No | Yes | Unclear | Moderate |
| Bajema | 2020-09-08 | No | No | Yes | No | Unclear | Yes | No | Yes | Unclear | Moderate |
| Bajema | 2020-09-08 | No | No | Yes | No | Unclear | Yes | No | Yes | Unclear | Moderate |
| Bajema | 2020-09-08 | No | No | Yes | No | Unclear | Yes | No | Yes | Unclear | Moderate |
| Bajema | 2020-09-08 | No | No | Yes | No | Unclear | Yes | No | Yes | Unclear | Moderate |
| Bajema | 2020-09-08 | No | No | Yes | No | Unclear | Yes | No | Yes | Unclear | Moderate |
| Bajema | 2020-09-08 | No | No | Yes | No | Unclear | Yes | No | Yes | Unclear | Moderate |
| Bajema | 2020-09-08 | No | No | Yes | No | Unclear | Yes | No | Yes | Unclear | Moderate |
| Bajema | 2020-09-08 | No | No | Yes | No | Unclear | Yes | No | Yes | Unclear | Moderate |
| Bajema | 2020-09-08 | No | No | Yes | No | Unclear | Yes | No | Yes | Unclear | Moderate |
| Bajema | 2020-09-08 | No | No | Yes | No | Unclear | Yes | No | Yes | Unclear | Moderate |
| Bajema | 2020-09-07 | No | No | Yes | No | Unclear | Yes | No | Yes | Unclear | Moderate |
| Demonbreun | 2020-09-06 | Yes | No | Yes | Yes | Yes | Unclear | No | No | Unclear | High |

| Article Author (Study Organization) | Sampling End Date (YMD) | Item 1: Appropriate sample frame* | Item 2: Probability sampling method | Item 3: Adequate sample size | Item 4: Subjects & setting described | Item 5: Good coverage of sample | Item 6: Sens > 90% & spec > 95% | Item 7: Same tests for all subjects | Item 8: Appropriate analysis | Item 9: Adequate response rate | Overall Risk of Bias† |
|---|---|---|---|---|---|---|---|---|---|---|---|
| Bajema | 2020-09-05 | No | No | Yes | No | Unclear | Yes | No | Yes | Unclear | Moderate |
| Bajema | 2020-09-04 | No | No | Yes | No | Unclear | Yes | No | Yes | Unclear | Moderate |
| Bajema | 2020-09-04 | No | No | Yes | No | Unclear | Yes | No | Yes | Unclear | Moderate |
| Bajema | 2020-09-03 | No | No | Yes | No | Unclear | Yes | No | Yes | Unclear | Moderate |
| Bajema | 2020-09-03 | No | No | Yes | No | Unclear | Yes | No | Yes | Unclear | Moderate |
| Bajema | 2020-09-03 | No | No | Yes | No | Unclear | Yes | No | Yes | Unclear | Moderate |
| Bajema | 2020-09-03 | No | No | Yes | No | Unclear | Yes | No | Yes | Unclear | Moderate |
| Bajema | 2020-09-03 | No | No | Yes | No | Unclear | Yes | No | Yes | Unclear | Moderate |
| Bajema | 2020-09-02 | No | No | Yes | No | Unclear | Yes | No | Yes | Unclear | Moderate |
| Bajema | 2020-09-01 | No | No | Yes | No | Unclear | Yes | No | Yes | Unclear | Moderate |
| Maleck | 2020-08-31 | Yes | Unclear | No | No | Unclear | Yes | Yes | Yes | No | Moderate |
| Halbrook | 2020-08-31 | Yes | Unclear | Yes | Yes | Yes | Unclear | Yes | No | Unclear | Moderate |
| Halbrook | 2020-08-31 | Yes | Unclear | Yes | Yes | Yes | Unclear | Yes | No | Unclear | Moderate |
| Bajema | 2020-08-29 | No | No | Yes | No | Unclear | Yes | No | Yes | Unclear | Moderate |
| Bajema | 2020-08-27 | No | No | Yes | No | Unclear | Yes | No | Yes | Unclear | Moderate |
| Bajema | 2020-08-27 | No | No | Yes | No | Unclear | Yes | No | Yes | Unclear | Moderate |
| Bajema | 2020-08-27 | No | No | Yes | No | Unclear | Yes | No | Yes | Unclear | Moderate |
| Bajema | 2020-08-27 | No | No | Yes | No | Unclear | Yes | No | Yes | Unclear | Moderate |
| Bajema | 2020-08-27 | No | No | Yes | No | Unclear | Yes | No | Yes | Unclear | Moderate |
| Bajema | 2020-08-27 | No | No | Yes | No | Unclear | Yes | No | Yes | Unclear | Moderate |
| Bajema | 2020-08-27 | No | No | Yes | No | Unclear | Yes | No | Yes | Unclear | Moderate |

| Article Author (Study Organization) | Sampling End Date (YMD) | Item 1: Appropriate sample frame* | Item 2: Probability sampling method | Item 3: Adequate sample size | Item 4: Subjects & setting described | Item 5: Good coverage of sample | Item 6: Sens > 90% & spec > 95% | Item 7: Same tests for all subjects | Item 8: Appropriate analysis | Item 9: Adequate response rate | Overall Risk of Bias† |
|---|---|---|---|---|---|---|---|---|---|---|---|
| Bajema | 2020-08-27 | No | No | Yes | No | Unclear | Yes | No | Yes | Unclear | Moderate |
| Bajema | 2020-08-27 | No | No | Yes | No | Unclear | Yes | No | Yes | Unclear | Moderate |
| Bajema | 2020-08-27 | No | No | Yes | No | Unclear | Yes | No | Yes | Unclear | Moderate |
| Bajema | 2020-08-27 | No | No | Yes | No | Unclear | Yes | No | Yes | Unclear | Moderate |
| Bajema | 2020-08-27 | No | No | No | No | Unclear | Yes | No | Yes | Unclear | High |
| Bajema | 2020-08-27 | No | No | Yes | No | Unclear | Yes | No | Yes | Unclear | Moderate |
| Bajema | 2020-08-27 | No | No | No | No | Unclear | Yes | No | Yes | Unclear | High |
| Bajema | 2020-08-27 | No | No | Yes | No | Unclear | Yes | No | Yes | Unclear | Moderate |
| Bajema | 2020-08-27 | No | No | Yes | No | Unclear | Yes | No | Yes | Unclear | Moderate |
| Bajema | 2020-08-27 | No | No | Yes | No | Unclear | Yes | No | Yes | Unclear | Moderate |
| Bajema | 2020-08-27 | No | No | Yes | No | Unclear | Yes | No | Yes | Unclear | Moderate |
| Bajema | 2020-08-26 | No | No | Yes | No | Unclear | Yes | No | Yes | Unclear | Moderate |
| Bajema | 2020-08-26 | No | No | Yes | No | Unclear | Yes | No | Yes | Unclear | Moderate |
| Bajema | 2020-08-26 | No | No | Yes | No | Unclear | Yes | No | Yes | Unclear | Moderate |
| Bajema | 2020-08-26 | No | No | Yes | No | Unclear | Yes | No | Yes | Unclear | Moderate |
| Bajema | 2020-08-26 | No | No | Yes | No | Unclear | Yes | No | Yes | Unclear | Moderate |
| Bajema | 2020-08-26 | No | No | Yes | No | Unclear | Yes | No | Yes | Unclear | Moderate |
| Bajema | 2020-08-26 | No | No | Yes | No | Unclear | Yes | No | Yes | Unclear | Moderate |
| Bajema | 2020-08-26 | No | No | Yes | No | Unclear | Yes | No | Yes | Unclear | Moderate |
| Bajema | 2020-08-26 | No | No | Yes | No | Unclear | Yes | No | Yes | Unclear | Moderate |
| Bajema | 2020-08-26 | No | No | Yes | No | Unclear | Yes | No | Yes | Unclear | Moderate |

| Article Author (Study Organization) | Sampling End Date (YMD) | Item 1: Appropriate sample frame* | Item 2: Probability sampling method | Item 3: Adequate sample size | Item 4: Subjects & setting described | Item 5: Good coverage of sample | Item 6: Sens > 90% & spec > 95% | Item 7: Same tests for all subjects | Item 8: Appropriate analysis | Item 9: Adequate response rate | Overall Risk of Bias† |
|---|---|---|---|---|---|---|---|---|---|---|---|
| Bajema | 2020-08-26 | No | No | Yes | No | Unclear | Yes | No | Yes | Unclear | Moderate |
| Bajema | 2020-08-26 | No | No | No | No | Unclear | Yes | No | Yes | Unclear | High |
| Bajema | 2020-08-26 | No | No | Yes | No | Unclear | Yes | No | Yes | Unclear | Moderate |
| Bajema | 2020-08-26 | No | No | No | No | Unclear | Yes | No | Yes | Unclear | High |
| Bajema | 2020-08-26 | No | No | Yes | No | Unclear | Yes | No | Yes | Unclear | Moderate |
| Bajema | 2020-08-26 | No | No | Yes | No | Unclear | Yes | No | Yes | Unclear | Moderate |
| Bajema | 2020-08-25 | No | No | Yes | No | Unclear | Yes | No | Yes | Unclear | Moderate |
| Bajema | 2020-08-25 | No | No | Yes | No | Unclear | Yes | No | Yes | Unclear | Moderate |
| Bajema | 2020-08-25 | No | No | Yes | No | Unclear | Yes | No | Yes | Unclear | Moderate |
| Bajema | 2020-08-25 | No | No | Yes | No | Unclear | Yes | No | Yes | Unclear | Moderate |
| Bajema | 2020-08-25 | No | No | Yes | No | Unclear | Yes | No | Yes | Unclear | Moderate |
| Bajema | 2020-08-25 | No | No | Yes | No | Unclear | Yes | No | Yes | Unclear | Moderate |
| Bajema | 2020-08-25 | No | No | Yes | No | Unclear | Yes | No | Yes | Unclear | Moderate |
| Bajema | 2020-08-25 | No | No | Yes | No | Unclear | Yes | No | Yes | Unclear | Moderate |
| Bajema | 2020-08-24 | No | No | Yes | No | Unclear | Yes | No | Yes | Unclear | Moderate |
| Bajema | 2020-08-24 | No | No | No | No | Unclear | Yes | No | Yes | Unclear | High |
| Bajema | 2020-08-24 | No | No | Yes | No | Unclear | Yes | No | Yes | Unclear | Moderate |
| Bajema | 2020-08-24 | No | No | No | No | Unclear | Yes | No | Yes | Unclear | High |
| Bajema | 2020-08-21 | No | No | Yes | No | Unclear | Yes | No | Yes | Unclear | Moderate |
| Bajema | 2020-08-19 | No | No | Yes | No | Unclear | Yes | No | Yes | Unclear | Moderate |
| Bajema | 2020-08-18 | No | No | Yes | No | Unclear | Yes | No | Yes | Unclear | Moderate |

| Article Author (Study Organization) | Sampling End Date (YMD) | Item 1: Appropriate sample frame* | Item 2: Probability sampling method | Item 3: Adequate sample size | Item 4: Subjects & setting described | Item 5: Good coverage of sample | Item 6: Sens > 90% & spec > 95% | Item 7: Same tests for all subjects | Item 8: Appropriate analysis | Item 9: Adequate response rate | Overall Risk of Bias† |
|---|---|---|---|---|---|---|---|---|---|---|---|
| Bajema | 2020-08-17 | No | No | Yes | No | Unclear | Yes | No | Yes | Unclear | Moderate |
| Bruckner | 2020-08-16 | Yes | Yes | Yes | Yes | Yes | Yes | Yes | Yes | No | Low |
| Brant-Zawadzki | 2020-08-15 | Yes | No | No | Yes | Unclear | Yes | Yes | No | Unclear | High |
| Brant-Zawadzki | 2020-08-15 | Yes | No | Yes | Yes | Yes | Yes | Yes | No | Unclear | High |
| Sabourin | 2020-08-15 | Yes | No | No | Yes | Unclear | No | Yes | No | Unclear | High |
| Heim | 2020-08-15 | Yes | Yes | Yes | No | Unclear | Unclear | Unclear | Unclear | Unclear | Unclear |
| Bajema | 2020-08-14 | No | No | Yes | No | Unclear | Yes | No | Yes | Unclear | Moderate |
| Bajema | 2020-08-13 | No | No | Yes | No | Unclear | Yes | No | Yes | Unclear | Moderate |
| Bajema | 2020-08-13 | No | No | Yes | No | Unclear | Yes | No | Yes | Unclear | Moderate |
| Bajema | 2020-08-13 | No | No | Yes | No | Unclear | Yes | No | Yes | Unclear | Moderate |
| Bajema | 2020-08-13 | No | No | Yes | No | Unclear | Yes | No | Yes | Unclear | Moderate |
| Bajema | 2020-08-13 | No | No | Yes | No | Unclear | Yes | No | Yes | Unclear | Moderate |
| Bajema | 2020-08-13 | No | No | Yes | No | Unclear | Yes | No | Yes | Unclear | Moderate |
| Bajema | 2020-08-13 | No | No | Yes | No | Unclear | Yes | No | Yes | Unclear | Moderate |
| Bajema | 2020-08-13 | No | No | Yes | No | Unclear | Yes | No | Yes | Unclear | Moderate |
| Bajema | 2020-08-13 | No | No | Yes | No | Unclear | Yes | No | Yes | Unclear | Moderate |
| Bajema | 2020-08-13 | No | No | Yes | No | Unclear | Yes | No | Yes | Unclear | Moderate |
| Bajema | 2020-08-13 | No | No | Yes | No | Unclear | Yes | No | Yes | Unclear | Moderate |
| Bajema | 2020-08-13 | No | No | Yes | No | Unclear | Yes | No | Yes | Unclear | Moderate |
| Bajema | 2020-08-13 | No | No | No | No | Unclear | Yes | No | Yes | Unclear | High |
| Bajema | 2020-08-13 | No | No | Yes | No | Unclear | Yes | No | Yes | Unclear | Moderate |

| Article Author (Study Organization) | Sampling End Date (YMD) | Item 1: Appropriate sample frame* | Item 2: Probability sampling method | Item 3: Adequate sample size | Item 4: Subjects & setting described | Item 5: Good coverage of sample | Item 6: Sens > 90% & spec > 95% | Item 7: Same tests for all subjects | Item 8: Appropriate analysis | Item 9: Adequate response rate | Overall Risk of Bias† |
|---|---|---|---|---|---|---|---|---|---|---|---|
| Bajema | 2020-08-12 | No | No | No | No | Unclear | Yes | No | Yes | Unclear | High |
| Bajema | 2020-08-11 | No | No | No | No | Unclear | Yes | No | Yes | Unclear | High |
| Bajema | 2020-08-11 | No | No | No | No | Unclear | Yes | No | Yes | Unclear | High |
| Bajema | 2020-08-11 | No | No | Yes | No | Unclear | Yes | No | Yes | Unclear | Moderate |
| Bajema | 2020-08-11 | No | No | No | No | Unclear | Yes | No | Yes | Unclear | High |
| Bajema | 2020-08-11 | No | No | No | No | Unclear | Yes | No | Yes | Unclear | High |
| Bajema | 2020-08-11 | No | No | No | No | Unclear | Yes | No | Yes | Unclear | High |
| Bajema | 2020-08-11 | No | No | Yes | No | Unclear | Yes | No | Yes | Unclear | Moderate |
| Bajema | 2020-08-11 | No | No | Yes | No | Unclear | Yes | No | Yes | Unclear | Moderate |
| Bajema | 2020-08-11 | No | No | Yes | No | Unclear | Yes | No | Yes | Unclear | Moderate |
| Bajema | 2020-08-11 | No | No | Yes | No | Unclear | Yes | No | Yes | Unclear | Moderate |
| Bajema | 2020-08-11 | No | No | Yes | No | Unclear | Yes | No | Yes | Unclear | Moderate |
| Bajema | 2020-08-11 | No | No | Yes | No | Unclear | Yes | No | Yes | Unclear | Moderate |
| Bajema | 2020-08-11 | No | No | Yes | No | Unclear | Yes | No | Yes | Unclear | Moderate |
| Bajema | 2020-08-11 | No | No | No | No | Unclear | Yes | No | Yes | Unclear | High |
| Bajema | 2020-08-11 | No | No | Yes | No | Unclear | Yes | No | Yes | Unclear | Moderate |
| Bajema | 2020-08-11 | No | No | Yes | No | Unclear | Yes | No | Yes | Unclear | Moderate |
| Bajema | 2020-08-11 | No | No | Yes | No | Unclear | Yes | No | Yes | Unclear | Moderate |
| Bajema | 2020-08-11 | No | No | No | No | Unclear | Yes | No | Yes | Unclear | High |
| Bajema | 2020-08-11 | No | No | Yes | No | Unclear | Yes | No | Yes | Unclear | Moderate |
| Bajema | 2020-08-11 | No | No | Yes | No | Unclear | Yes | No | Yes | Unclear | Moderate |

| Article Author (Study Organization) | Sampling End Date (YMD) | Item 1: Appropriate sample frame* | Item 2: Probability sampling method | Item 3: Adequate sample size | Item 4: Subjects & setting described | Item 5: Good coverage of sample | Item 6: Sens > 90% & spec > 95% | Item 7: Same tests for all subjects | Item 8: Appropriate analysis | Item 9: Adequate response rate | Overall Risk of Bias† |
|---|---|---|---|---|---|---|---|---|---|---|---|
| Bajema | 2020-08-10 | No | No | Yes | No | Unclear | Yes | No | Yes | Unclear | Moderate |
| Bajema | 2020-08-10 | No | No | Yes | No | Unclear | Yes | No | Yes | Unclear | Moderate |
| Bajema | 2020-08-10 | No | No | Yes | No | Unclear | Yes | No | Yes | Unclear | Moderate |
| Bajema | 2020-08-10 | No | No | No | No | Unclear | Yes | No | Yes | Unclear | High |
| Bajema | 2020-08-10 | No | No | Yes | No | Unclear | Yes | No | Yes | Unclear | Moderate |
| Bajema | 2020-08-08 | No | No | Yes | No | Unclear | Yes | No | Yes | Unclear | Moderate |
| Bajema | 2020-08-07 | No | No | Yes | No | Unclear | Yes | No | Yes | Unclear | Moderate |
| Bajema | 2020-08-07 | No | No | Yes | No | Unclear | Yes | No | Yes | Unclear | Moderate |
| Bajema | 2020-08-05 | No | No | Yes | No | Unclear | Yes | No | Yes | Unclear | Moderate |
| Bajema | 2020-08-05 | No | No | Yes | No | Unclear | Yes | No | Yes | Unclear | Moderate |
| Bajema | 2020-08-04 | No | No | Yes | No | Unclear | Yes | No | Yes | Unclear | Moderate |
| Bajema | 2020-08-03 | No | No | Yes | No | Unclear | Yes | No | Yes | Unclear | Moderate |
| Bajema | 2020-08-03 | No | No | Yes | No | Unclear | Yes | No | Yes | Unclear | Moderate |
| Bajema | 2020-08-02 | No | No | Yes | No | Unclear | Yes | No | Yes | Unclear | Moderate |
| Coffman | 2020-07-31 | Unclear | Unclear | Yes | No | Unclear | Unclear | Unclear | Unclear | Unclear | Unclear |
| Feehan | 2020-07-31 | Yes | Yes | Yes | Yes | Unclear | Yes | Yes | No | Yes | Moderate |
| Vassallo | 2020-07-31 | No | Yes | Yes | Yes | Unclear | Unclear | Yes | No | Unclear | Moderate |
| Azzi | 2020-07-29 | Yes | No | Yes | No | No | Yes | Yes | No | Unclear | Moderate |
| Nicholson | 2020-07-28 | Yes | No | Yes | No | Unclear | Yes | No | No | Unclear | High |
| Spencer | 2020-07-27 | No | No | Yes | No | Unclear | Unclear | Yes | No | Unclear | High |
| Yamaki | 2020-07-27 | No | No | Yes | Yes | Unclear | Yes | Yes | No | Unclear | High |

| Article Author (Study Organization) | Sampling End Date (YMD) | Item 1: Appropriate sample frame* | Item 2: Probability sampling method | Item 3: Adequate sample size | Item 4: Subjects & setting described | Item 5: Good coverage of sample | Item 6: Sens > 90% & spec > 95% | Item 7: Same tests for all subjects | Item 8: Appropriate analysis | Item 9: Adequate response rate | Overall Risk of Bias† |
|---|---|---|---|---|---|---|---|---|---|---|---|
| Haizler-Cohen | 2020-07-24 | Yes | Unclear | Yes | Yes | Unclear | Unclear | Yes | No | Unclear | High |
| Klawiter | 2020-07-23 | No | No | No | No | Unclear | Yes | Yes | No | Unclear | High |
| Klawiter | 2020-07-23 | Yes | No | No | No | Unclear | Yes | Yes | No | Unclear | High |
| Mahajan | 2020-07-22 | Yes | Yes | No | Yes | No | Yes | Yes | No | No | High |
| Mahajan | 2020-07-22 | Yes | Yes | No | Yes | No | Yes | Yes | No | No | High |
| McMillan | 2020-07-20 | Yes | Yes | Yes | No | Unclear | Unclear | Unclear | No | Unclear | Unclear |
| Lopez | 2020-07-20 | Yes | No | Yes | Yes | Yes | Yes | Yes | No | Yes | Moderate |
| Zauzmer | 2020-07-15 | Yes | No | Yes | No | Unclear | Unclear | No | No | Unclear | High |
| Jin | 2020-07-15 | No | No | Yes | Yes | No | Unclear | Yes | No | Unclear | High |
| Reuben | 2020-07-15 | Yes | No | No | Yes | Unclear | Unclear | Yes | No | Yes | High |
| Kaufman | 2020-07-10 | Yes | No | Yes | No | Unclear | Unclear | No | No | Unclear | High |
| Shah | 2020-07-09 | Yes | Unclear | No | Yes | No | Unclear | No | No | Unclear | High |
| Shah | 2020-07-09 | Yes | Unclear | No | Yes | No | Unclear | No | No | Unclear | High |
| Mohr | 2020-07-08 | Yes | No | Yes | Yes | Yes | Yes | Yes | No | Unclear | Moderate |
| Dimcheff | 2020-07-08 | Yes | No | Yes | Yes | Yes | Yes | Yes | No | No | Moderate |
| Kamath | 2020-07-07 | No | No | Yes | Yes | No | Yes | Yes | No | Unclear | High |
| Childs | 2020-07-06 | Yes | No | Yes | No | No | No | Yes | No | Unclear | High |
| Stadlbauer | 2020-07-05 | No | Yes | Yes | Yes | No | Yes | Yes | No | Unclear | High |
| Stadlbauer | 2020-07-05 | No | No | Yes | Yes | No | Yes | Yes | No | Unclear | High |
| Freeman | 2020-07-04 | Yes | No | Yes | Yes | No | Unclear | Yes | No | Unclear | High |
| Walker | 2020-07-01 | Unclear | Unclear | Unclear | Unclear | Unclear | Unclear | Unclear | Unclear | Unclear | Moderate |

| Article Author (Study Organization) | Sampling End Date (YMD) | Item 1: Appropriate sample frame* | Item 2: Probability sampling method | Item 3: Adequate sample size | Item 4: Subjects & setting described | Item 5: Good coverage of sample | Item 6: Sens > 90% & spec > 95% | Item 7: Same tests for all subjects | Item 8: Appropriate analysis | Item 9: Adequate response rate | Overall Risk of Bias† |
|---|---|---|---|---|---|---|---|---|---|---|---|
| Barrett | 2020-06-30 | Yes | No | Yes | Yes | Yes | Yes | Yes | No | Yes | Moderate |
| Wilkins | 2020-06-30 | Yes | No | Yes | Yes | Yes | Yes | Yes | Yes | No | Moderate |
| Nailescu | 2020-06-30 | Yes | No | No | Yes | Yes | Yes | Yes | No | Unclear | High |
| Viglienzoni | 2020-06-30 | Yes | No | No | No | Unclear | Unclear | Yes | No | Unclear | High |
| Samore | 2020-06-30 | Yes | Yes | Yes | Yes | Yes | No | Yes | Yes | No | Moderate |
| Rivas | 2020-06-28 | Yes | No | Yes | Yes | Yes | Yes | Yes | No | No | Moderate |
| Patel | 2020-06-27 | Yes | No | No | Yes | Unclear | Yes | Yes | No | No | High |
| Baker | 2020-06-26 | Yes | No | Yes | Yes | Yes | Yes | Yes | Yes | No | Moderate |
| McBride | 2020-06-25 | Yes | No | Yes | Unclear | Unclear | Unclear | Yes | No | Unclear | High |
| Mahajan | 2020-06-23 | Yes | Yes | No | Yes | Yes | Yes | Yes | No | Yes | Moderate |
| Moscola | 2020-06-23 | Yes | No | Yes | Yes | Yes | No | No | No | Unclear | High |
| Barzin | 2020-06-19 | No | No | Yes | Yes | No | Yes | Yes | Yes | Unclear | Moderate |
| Spectrum News Staff | 2020-06-19 | Yes | No | Yes | No | Yes | Unclear | Unclear | No | No | High |
| Self | 2020-06-19 | Yes | No | Yes | Yes | Unclear | Yes | Yes | No | Unclear | Moderate |
| Dodd | 2020-06-15 | No | Yes | Yes | Yes | Yes | Yes | Yes | No | Yes | Moderate |
| Reifer | 2020-06-15 | No | No | Yes | Yes | Unclear | Unclear | Yes | No | Unclear | High |
| Sutton | 2020-06-15 | No | Yes | Yes | No | Unclear | Yes | Yes | No | Unclear | High |
| Rebeiro | 2020-06-15 | Yes | No | Yes | No | Unclear | Unclear | Yes | No | Unclear | High |
| Anand | 2020-06-15 | No | Yes | Yes | Yes | Yes | Yes | Yes | Yes | Yes | Moderate |
| Akinbami | 2020-06-13 | Yes | No | Yes | Yes | Unclear | Yes | Yes | No | Unclear | Moderate |
| Dora | 2020-06-12 | Yes | No | No | No | Unclear | Yes | Yes | No | Unclear | High |

| Article Author (Study Organization) | Sampling End Date (YMD) | Item 1: Appropriate sample frame* | Item 2: Probability sampling method | Item 3: Adequate sample size | Item 4: Subjects & setting described | Item 5: Good coverage of sample | Item 6: Sens > 90% & spec > 95% | Item 7: Same tests for all subjects | Item 8: Appropriate analysis | Item 9: Adequate response rate | Overall Risk of Bias[†] |
|---|---|---|---|---|---|---|---|---|---|---|---|
| Hynes | 2020-06-10 | Yes | Yes | No | No | Unclear | Unclear | Yes | No | No | High |
| Harden | 2020-06-10 | No | No | Yes | No | Unclear | Unclear | Unclear | No | Unclear | High |
| Yogo | 2020-06-08 | No | No | Yes | Yes | Yes | Yes | Yes | No | No | High |
| Hogan | 2020-06-08 | Yes | No | Yes | Yes | Unclear | Unclear | Yes | No | Yes | High |
| Rudavsky | 2020-06-08 | Yes | Yes | Yes | No | Yes | Unclear | Unclear | No | Unclear | Moderate |
| Barzin | 2020-06-04 | Yes | No | Yes | Yes | Unclear | Yes | Yes | No | Unclear | Moderate |
| Flannery | 2020-06-03 | Yes | Yes | Yes | Yes | Yes | Unclear | Yes | No | Yes | Moderate |
| Coffman | 2020-05-31 | Unclear | Unclear | Yes | No | Unclear | Unclear | Unclear | Unclear | Unclear | Unclear |
| Addetia | 2020-05-31 | No | No | No | No | No | Unclear | Yes | No | Yes | High |
| Gray | 2020-05-31 | Yes | Unclear | No | No | Unclear | Unclear | Yes | No | Unclear | High |
| Brunner | 2020-05-29 | Yes | No | Yes | Yes | Yes | Yes | Yes | No | Unclear | High |
| Brunner | 2020-05-29 | Unclear | Unclear | Yes | Yes | Unclear | Yes | Yes | No | Unclear | High |
| Ohnesorge | 2020-05-29 | Yes | No | No | No | Unclear | Unclear | Unclear | No | Unclear | Unclear |
| Sims | 2020-05-28 | Yes | No | Yes | Yes | Unclear | Yes | Yes | No | No | Moderate |
| Hayden | 2020-05-28 | Yes | No | Yes | No | Unclear | Unclear | Yes | No | Unclear | Unclear |
| Liu | 2020-05-26 | No | No | Yes | No | Unclear | Unclear | Yes | No | Unclear | High |
| Osher | 2020-05-26 | Yes | No | No | No | Unclear | Unclear | Yes | No | Unclear | High |
| Mukherjee | 2020-05-26 | Yes | No | Yes | No | Unclear | Unclear | Unclear | No | Unclear | High |
| Stavert | 2020-05-24 | No | No | No | Yes | Unclear | Unclear | Yes | No | Unclear | High |
| Ahmad | 2020-05-22 | No | Unclear | No | Yes | Unclear | Yes | Yes | No | Unclear | High |
| Jackson | 2020-05-22 | Yes | Yes | Yes | No | Unclear | Unclear | Unclear | Unclear | Unclear | Unclear |

| Article Author (Study Organization) | Sampling End Date (YMD) | Item 1: Appropriate sample frame* | Item 2: Probability sampling method | Item 3: Adequate sample size | Item 4: Subjects & setting described | Item 5: Good coverage of sample | Item 6: Sens > 90% & spec > 95% | Item 7: Same tests for all subjects | Item 8: Appropriate analysis | Item 9: Adequate response rate | Overall Risk of Bias† |
|---|---|---|---|---|---|---|---|---|---|---|---|
| Jackson | 2020-05-22 | Yes | Yes | Yes | No | Unclear | Unclear | Unclear | Unclear | Unclear | Unclear |
| Jackson | 2020-05-22 | Yes | Yes | No | No | Unclear | Unclear | Unclear | Unclear | Unclear | Unclear |
| Jackson | 2020-05-22 | Yes | Yes | Yes | No | Unclear | Unclear | Unclear | Unclear | Unclear | Unclear |
| Staletovich | 2020-05-22 | Yes | Unclear | Yes | No | Unclear | Unclear | Unclear | No | Unclear | Unclear |
| Leidner | 2020-05-22 | Yes | No | Yes | Yes | Yes | No | Yes | No | Unclear | High |
| Pritchett | 2020-05-21 | No | Unclear | No | Yes | Unclear | Unclear | Yes | No | Unclear | High |
| Shukla | 2020-05-21 | Yes | No | Yes | Yes | Unclear | Yes | Yes | No | Unclear | Moderate |
| Rosser | 2020-05-20 | Yes | No | Yes | Yes | Yes | No | Yes | No | Unclear | High |
| Alfini | 2020-05-20 | Yes | No | No | No | Unclear | Unclear | Unclear | No | Unclear | Unclear |
| Alfini | 2020-05-20 | Yes | No | Yes | No | Unclear | Unclear | Unclear | No | Unclear | Unclear |
| Montgomery | 2020-05-20 | No | No | Yes | No | Unclear | Unclear | No | No | Unclear | High |
| Geraci | 2020-05-20 | No | Unclear | No | No | Unclear | Unclear | Yes | No | Unclear | High |
| Tarabichi | 2020-05-19 | Yes | No | No | Yes | Yes | Unclear | Yes | No | Yes | High |
| Mughal | 2020-05-19 | No | No | No | Yes | Unclear | Yes | Yes | No | Yes | High |
| McLaughlin | 2020-05-19 | Yes | No | Yes | Yes | Yes | Yes | Yes | Yes | Yes | Moderate |
| Morris | 2020-05-18 | Yes | Unclear | No | No | Unclear | Unclear | Unclear | No | Unclear | Unclear |
| Feehan | 2020-05-15 | Yes | Yes | Yes | Yes | Unclear | Unclear | Yes | No | Unclear | Moderate |
| Meyer | 2020-05-15 | Yes | No | Yes | No | Unclear | Unclear | Unclear | No | Unclear | High |
| Dietrich | 2020-05-15 | No | No | Yes | Yes | Yes | Unclear | Yes | No | Unclear | High |
| City of Boston | 2020-05-15 | Yes | Yes | Yes | Yes | Yes | Unclear | Yes | Unclear | No | Moderate |
| Parker-Magyar | 2020-05-14 | Yes | No | No | No | Unclear | Unclear | Unclear | No | Unclear | High |

| Article Author (Study Organization) | Sampling End Date (YMD) | Item 1: Appropriate sample frame* | Item 2: Probability sampling method | Item 3: Adequate sample size | Item 4: Subjects & setting described | Item 5: Good coverage of sample | Item 6: Sens > 90% & spec > 95% | Item 7: Same tests for all subjects | Item 8: Appropriate analysis | Item 9: Adequate response rate | Overall Risk of Bias† |
|---|---|---|---|---|---|---|---|---|---|---|---|
| Iwuji | 2020-05-13 | Yes | No | Yes | No | Unclear | Yes | Yes | No | Unclear | High |
| CBS Los Angeles | 2020-05-12 | Yes | Unclear | Yes | No | Unclear | Unclear | Unclear | No | Unclear | Unclear |
| Nesbitt | 2020-05-11 | No | No | Yes | Yes | Unclear | Yes | Yes | No | Unclear | Moderate |
| Tilley | 2020-05-08 | Yes | No | Yes | Yes | Unclear | Yes | Yes | Yes | Unclear | Moderate |
| Hunter | 2020-05-08 | Yes | No | Yes | Yes | No | Yes | Yes | No | Unclear | High |
| Ripperger | 2020-05-07 | Yes | No | Yes | Yes | Unclear | Yes | Yes | No | Unclear | High |
| Racine-Brzostek | 2020-05-07 | Yes | No | Yes | Yes | Yes | Unclear | Yes | No | Unclear | Moderate |
| Regan | 2020-05-06 | Yes | Unclear | No | No | Unclear | Unclear | Unclear | No | Unclear | Unclear |
| Regan | 2020-05-06 | No | Unclear | No | No | Unclear | Unclear | Unclear | No | Unclear | Unclear |
| Sydney | 2020-05-04 | Yes | No | Yes | Yes | Unclear | Unclear | Unclear | No | No | High |
| Bajema | 2020-05-03 | Yes | Yes | Yes | Yes | Yes | Unclear | Yes | Yes | Yes | Low |
| Biggs | 2020-05-03 | Yes | Yes | Yes | Yes | Yes | Yes | Yes | No | Unclear | Moderate |
| Menachemi | 2020-05-03 | Yes | No | No | Yes | Yes | Yes | Yes | No | Unclear | High |
| Reagan | 2020-05-02 | No | Unclear | Yes | No | Unclear | Yes | Yes | No | Unclear | High |
| Venugopal | 2020-05-01 | Yes | No | No | Yes | Yes | Yes | Yes | No | Yes | Moderate |
| Bajema | 2020-05-01 | No | No | Yes | Yes | Unclear | Yes | Yes | Yes | Unclear | Moderate |
| Jeremias | 2020-04-30 | No | No | Yes | Yes | Yes | Unclear | No | No | No | High |
| Cohen | 2020-04-30 | Yes | No | Yes | Yes | Unclear | No | Yes | No | Unclear | High |
| Jeong | 2020-04-30 | No | No | No | Yes | Unclear | Unclear | Yes | No | Yes | High |
| Menachemi | 2020-04-29 | Yes | Yes | Yes | Yes | No | Yes | Yes | No | No | Moderate |
| Chamie | 2020-04-28 | Yes | No | Yes | Yes | Unclear | Yes | Yes | Yes | Unclear | Moderate |

| Article Author (Study Organization) | Sampling End Date (YMD) | Item 1: Appropriate sample frame* | Item 2: Probability sampling method | Item 3: Adequate sample size | Item 4: Subjects & setting described | Item 5: Good coverage of sample | Item 6: Sens > 90% & spec > 95% | Item 7: Same tests for all subjects | Item 8: Appropriate analysis | Item 9: Adequate response rate | Overall Risk of Bias† |
|---|---|---|---|---|---|---|---|---|---|---|---|
| Rosenberg | 2020-04-28 | Yes | No | Yes | Yes | Yes | Yes | Yes | Yes | Yes | Moderate |
| Zou | 2020-04-27 | No | No | No | No | Unclear | Unclear | Yes | No | Unclear | High |
| Bryan | 2020-04-26 | Yes | No | Yes | Yes | Unclear | Yes | Yes | No | Unclear | High |
| Lewis | 2020-04-25 | Yes | No | No | Yes | Yes | Unclear | Yes | No | Yes | High |
| Appa | 2020-04-24 | Yes | No | Yes | Yes | No | Yes | Yes | No | Yes | Moderate |
| Payne | 2020-04-24 | No | No | No | No | Unclear | Unclear | Yes | No | No | High |
| Sassoon | 2020-04-24 | Yes | No | Yes | No | Unclear | Yes | Yes | No | Unclear | High |
| Dingens | 2020-04-24 | No | No | Yes | Yes | Unclear | Unclear | Yes | No | Unclear | High |
| Kiefer | 2020-04-23 | No | No | No | Yes | No | Unclear | Yes | No | No | High |
| Morrison | 2020-04-22 | No | No | Yes | No | Unclear | Yes | Yes | No | Unclear | High |
| Morcuende | 2020-04-21 | No | No | No | Yes | No | Yes | Yes | No | No | High |
| McCormack | 2020-04-21 | Yes | Unclear | Yes | No | Unclear | Unclear | Unclear | No | Unclear | Unclear |
| Paterson | 2020-04-21 | No | Unclear | No | No | Unclear | Unclear | Yes | Unclear | Unclear | Unclear |
| Stock | 2020-04-20 | No | No | No | Yes | No | Unclear | Yes | No | Unclear | High |
| Madsen | 2020-04-19 | No | No | No | No | Unclear | Yes | Yes | No | Yes | High |
| Caban-Martinez | 2020-04-17 | No | No | No | Yes | Yes | No | Yes | No | Unclear | High |
| Naranbhai | 2020-04-15 | Yes | No | No | Yes | Unclear | Yes | Yes | Yes | Unclear | Moderate |
| Sood | 2020-04-14 | Yes | Yes | Yes | Yes | Yes | No | Yes | Yes | No | Moderate |
| Ng | 2020-04-13 | No | Yes | No | Yes | Yes | Yes | Yes | No | Unclear | High |
| Goldberg | 2020-04-06 | Yes | No | No | Yes | Yes | Yes | Yes | No | Yes | High |
| Bendavid | 2020-04-04 | No | No | Yes | Yes | No | No | Yes | Yes | Unclear | High |

| Article Author (Study Organization) | Sampling End Date (YMD) | Item 1: Appropriate sample frame* | Item 2: Probability sampling method | Item 3: Adequate sample size | Item 4: Subjects & setting described | Item 5: Good coverage of sample | Item 6: Sens > 90% & spec > 95% | Item 7: Same tests for all subjects | Item 8: Appropriate analysis | Item 9: Adequate response rate | Overall Risk of Bias[†] |
|---|---|---|---|---|---|---|---|---|---|---|---|
| Mansour | 2020-04-04 | Yes | No | No | Yes | Yes | Unclear | Yes | No | Unclear | High |
| Ng | 2020-03-31 | No | No | Yes | Yes | Yes | Yes | Yes | No | Unclear | Moderate |
| Schuchat | 2020-03-31 | No | No | No | No | Unclear | Unclear | Unclear | No | Unclear | High |
| McPherson | 2020-03-06 | Yes | No | No | Yes | Yes | Yes | Yes | No | No | High |
| Chu | 2020-02-03 | Yes | No | No | No | Unclear | Yes | Yes | No | Yes | High |
| Basavaraju | 2020-01-17 | No | No | Yes | Yes | No | Yes | Yes | No | Unclear | Moderate |
| **Latin America and Caribbean** | | | | | | | | | | | |
| **Brazil** | | | | | | | | | | | |
| Buss | 2020-10-24 | No | No | Yes | No | Yes | Yes | Yes | Yes | Unclear | Moderate |
| Buss | 2020-10-17 | No | No | Yes | No | Yes | Yes | Yes | Yes | Unclear | Moderate |
| Buss | 2020-09-29 | No | No | Yes | No | Yes | Yes | Yes | Yes | Unclear | Moderate |
| Buss | 2020-09-14 | No | No | Yes | No | Yes | Yes | Yes | Yes | Unclear | Moderate |
| Buss | 2020-08-29 | No | No | Yes | No | Yes | Yes | Yes | Yes | Unclear | Moderate |
| Buss | 2020-08-19 | No | No | Yes | No | Yes | Yes | Yes | Yes | Unclear | Moderate |
| Santana | 2020-08-15 | Yes | No | No | Yes | Unclear | Unclear | Yes | No | Unclear | High |
| daSilva | 2020-08-08 | Yes | Yes | Yes | Yes | Yes | Yes | Yes | No | Yes | Moderate |
| Oliveira | 2020-08-04 | No | No | No | Yes | Unclear | Yes | Yes | No | Unclear | High |
| Buss | 2020-07-31 | No | Unclear | Yes | No | Unclear | Unclear | Yes | No | Unclear | Unclear |
| Silva | 2020-07-31 | No | No | No | Yes | Unclear | Unclear | Unclear | No | Unclear | High |
| Paula | 2020-07-31 | No | No | Yes | Yes | Unclear | Unclear | Yes | No | Yes | High |
| Buss | 2020-07-25 | No | No | Yes | No | Yes | Yes | Yes | Yes | Unclear | Moderate |

| Article Author (Study Organization) | Sampling End Date (YMD) | Item 1: Appropriate sample frame* | Item 2: Probability sampling method | Item 3: Adequate sample size | Item 4: Subjects & setting described | Item 5: Good coverage of sample | Item 6: Sens > 90% & spec > 95% | Item 7: Same tests for all subjects | Item 8: Appropriate analysis | Item 9: Adequate response rate | Overall Risk of Bias† |
|---|---|---|---|---|---|---|---|---|---|---|---|
| Buss | 2020-07-15 | No | No | Yes | No | Yes | Yes | Yes | Yes | Unclear | Moderate |
| Melo | 2020-06-30 | Yes | No | No | No | Unclear | Yes | Yes | No | Unclear | High |
| Horta | 2020-06-24 | Yes | Yes | Yes | Yes | No | No | Yes | No | Yes | Moderate |
| Buss | 2020-06-20 | No | No | Yes | No | Yes | Yes | Yes | Yes | Unclear | Moderate |
| Picon | 2020-06-17 | Yes | Yes | Yes | Yes | Yes | No | Yes | Yes | Yes | Moderate |
| Buss | 2020-06-15 | No | No | Yes | No | Yes | Yes | Yes | Yes | Unclear | Moderate |
| Picon | 2020-06-04 | Yes | Yes | Yes | Yes | Yes | No | Yes | Yes | Yes | Moderate |
| Vieira | 2020-05-31 | Yes | Yes | Yes | No | Unclear | No | Yes | No | Unclear | Moderate |
| Sales | 2020-05-31 | Yes | Yes | Yes | Yes | Yes | Unclear | Yes | No | Unclear | Moderate |
| Jr | 2020-05-30 | Unclear | Unclear | No | Yes | Yes | Unclear | Unclear | Unclear | Unclear | Unclear |
| Costa | 2020-05-28 | Yes | No | Yes | Yes | Unclear | Unclear | Yes | No | Yes | Moderate |
| Vieira | 2020-05-24 | Yes | Yes | Yes | No | Unclear | No | Yes | No | Unclear | Moderate |
| Buss | 2020-05-21 | No | No | Yes | No | Yes | Yes | Yes | Yes | Unclear | Moderate |
| Vieira | 2020-05-17 | Yes | Yes | Yes | No | Unclear | No | Yes | No | Unclear | Moderate |
| Chioro | 2020-05-15 | Yes | Yes | Yes | Yes | Unclear | Unclear | Yes | No | Unclear | High |
| Gomes | 2020-05-15 | Yes | Yes | Yes | Yes | Yes | No | Yes | No | Unclear | Moderate |
| Gomes | 2020-05-15 | Yes | Yes | Yes | Yes | Yes | No | Yes | No | Unclear | Moderate |
| Buss | 2020-05-14 | No | No | Yes | No | Yes | Yes | Yes | Yes | Unclear | Moderate |
| Tess | 2020-05-12 | Yes | Yes | No | Yes | Yes | Yes | Yes | No | No | Moderate |
| Watanabe | 2020-05-12 | Yes | Yes | No | No | Unclear | Unclear | Unclear | No | Unclear | Unclear |
| Silveira | 2020-05-11 | Yes | Yes | Yes | Yes | Yes | No | Yes | Yes | Yes | Low |

| Article Author (Study Organization) | Sampling End Date (YMD) | Item 1: Appropriate sample frame* | Item 2: Probability sampling method | Item 3: Adequate sample size | Item 4: Subjects & setting described | Item 5: Good coverage of sample | Item 6: Sens > 90% & spec > 95% | Item 7: Same tests for all subjects | Item 8: Appropriate analysis | Item 9: Adequate response rate | Overall Risk of Bias† |
|---|---|---|---|---|---|---|---|---|---|---|---|
| Vieira | 2020-05-10 | Yes | Yes | Yes | No | Unclear | No | Yes | No | Unclear | Moderate |
| Borges | 2020-05-09 | No | No | Yes | Yes | Yes | Yes | Yes | No | Unclear | High |
| Vieira | 2020-05-03 | Yes | Yes | Yes | No | Unclear | No | Yes | No | Unclear | Moderate |
| Buss | 2020-04-30 | No | No | Yes | No | Yes | Yes | Yes | Yes | Unclear | Moderate |
| Silva | 2020-04-29 | Yes | Unclear | No | No | Unclear | Unclear | Yes | No | Unclear | High |
| Silveira | 2020-04-27 | Yes | Yes | Yes | Yes | Yes | No | Yes | Yes | Yes | Low |
| Filho | 2020-04-27 | No | No | Yes | Yes | Unclear | No | Yes | Yes | Yes | Moderate |
| Vieira | 2020-04-26 | Yes | Yes | Yes | No | Unclear | No | Yes | No | Unclear | Moderate |
| Vieira | 2020-04-19 | Yes | Yes | Yes | No | Unclear | No | Yes | No | Unclear | Moderate |
| Buss | 2020-04-17 | No | No | Yes | No | Yes | Yes | Yes | Yes | Unclear | Moderate |
| Gurgel | 2020-04-15 | No | No | Yes | Yes | Unclear | Yes | Yes | No | Unclear | Moderate |
| Silveira | 2020-04-13 | Yes | Yes | Yes | Yes | Yes | No | Yes | Yes | Yes | Low |
| Buss | 2020-03-21 | No | No | Yes | No | Yes | Yes | Yes | Yes | Unclear | Moderate |
| Buss | 2020-03-12 | No | No | Yes | No | Yes | Yes | Yes | Yes | Unclear | Moderate |
| Buss | 2020-02-29 | No | No | Yes | No | Yes | Yes | Yes | Yes | Unclear | Moderate |
| Buss | 2020-02-13 | No | No | Yes | No | Yes | Yes | Yes | Yes | Unclear | Moderate |
| **Columbia** | | | | | | | | | | | |
| Mattar | 2020-09-01 | Yes | Yes | Yes | No | Unclear | Yes | Yes | No | Unclear | Moderate |
| Ariza | 2020-08-10 | Yes | No | No | Yes | Unclear | Yes | Yes | No | No | High |
| Idrovo | 2020-07-14 | No | No | Yes | No | No | Unclear | Unclear | No | No | High |
| **Cuba** | | | | | | | | | | | |

| Article Author (Study Organization) | Sampling End Date (YMD) | Item 1: Appropriate sample frame* | Item 2: Probability sampling method | Item 3: Adequate sample size | Item 4: Subjects & setting described | Item 5: Good coverage of sample | Item 6: Sens > 90% & spec > 95% | Item 7: Same tests for all subjects | Item 8: Appropriate analysis | Item 9: Adequate response rate | Overall Risk of Bias[†] |
|---|---|---|---|---|---|---|---|---|---|---|---|
| Giraldoni | 2020-04-06 | No | No | No | Yes | Unclear | Unclear | Unclear | No | Unclear | Unclear |
| **Domninican Republic** | | | | | | | | | | | |
| Paulino-Ramirez | 2020-06-15 | No | Yes | Yes | Yes | Unclear | No | Yes | No | Unclear | High |
| **Ecuador** | | | | | | | | | | | |
| Brutto | 2020-06-30 | No | Yes | No | Yes | Yes | Yes | Yes | No | Yes | High |
| Brutto | 2020-05-25 | Yes | No | Yes | Yes | Yes | Yes | Yes | No | Yes | Moderate |
| **French Guiana** | | | | | | | | | | | |
| Flamand | 2020-07-23 | No | No | No | Yes | Yes | Yes | Yes | No | Unclear | Moderate |
| **Honduras** | | | | | | | | | | | |
| Nunez | 2020-06-23 | Yes | Yes | Yes | Yes | Yes | Yes | Yes | No | Unclear | Moderate |
| **Mexico** | | | | | | | | | | | |
| Cruz-Arenas | 2020-09-09 | Yes | Yes | No | Yes | Yes | Unclear | Yes | No | Unclear | Moderate |
| Martinez-Acuna | 2020-08-30 | No | No | Yes | Yes | Yes | Unclear | Yes | No | Unclear | High |
| Remes-Troche | 2020-07-31 | No | Yes | Yes | Yes | Unclear | Unclear | Yes | No | Unclear | High |
| Diaz-Salazar | 2020-07-23 | Yes | Yes | Yes | Yes | Yes | Yes | Yes | No | Yes | Moderate |
| **Panama** | | | | | | | | | | | |
| Villarreal | 2020-07-07 | No | No | No | Yes | Unclear | No | Yes | No | Unclear | High |
| Villarreal | 2020-07-07 | No | No | No | Yes | No | No | Yes | No | Unclear | High |
| **Paraguay** | | | | | | | | | | | |
| Ultimahora | 2020-12-15 | Yes | Unclear | Yes | No | No | Unclear | Unclear | Unclear | Yes | Unclear |
| **Peru** | | | | | | | | | | | |

| Article Author (Study Organization) | Sampling End Date (YMD) | Item 1: Appropriate sample frame* | Item 2: Probability sampling method | Item 3: Adequate sample size | Item 4: Subjects & setting described | Item 5: Good coverage of sample | Item 6: Sens > 90% & spec > 95% | Item 7: Same tests for all subjects | Item 8: Appropriate analysis | Item 9: Adequate response rate | Overall Risk of Bias† |
|---|---|---|---|---|---|---|---|---|---|---|---|
| Alvarex-Antonio | 2020-08-15 | Yes | Yes | Yes | Yes | Yes | Yes | Yes | Yes | Yes | Low |
| Peruvian Government | 2020-07-09 | Yes | Yes | Yes | No | Unclear | No | Yes | No | Unclear | High |
| Angulo-Bazan | 2020-05-02 | Yes | No | No | Yes | Unclear | Yes | Yes | No | Unclear | Moderate |
| **North Africa and Middle East** | | | | | | | | | | | |
| **Egypt** | | | | | | | | | | | |
| Kassem | 2020-06-14 | No | No | No | Yes | Unclear | Yes | Yes | No | No | High |
| Mostafa | 2020-05-14 | Yes | No | Yes | Yes | Yes | No | Yes | No | Unclear | High |
| **Iran** | | | | | | | | | | | |
| Poustchi | 2020-06-02 | Yes | Yes | Yes | Yes | Yes | No | Yes | Yes | Yes | Low |
| Poustchi | 2020-06-02 | Yes | Yes | Yes | No | Unclear | No | Yes | Yes | Yes | Moderate |
| Saberian | 2020-05-20 | Yes | No | No | Yes | Unclear | No | Yes | No | Unclear | High |
| Armin | 2020-05-05 | Yes | No | No | Yes | Unclear | Yes | Yes | No | Unclear | High |
| Vazirinejad | 2020-04-30 | Yes | No | No | No | Unclear | Unclear | Yes | No | No | High |
| Shakiba | 2020-04-19 | Yes | Yes | No | Yes | Unclear | No | Yes | Yes | No | Moderate |
| Ossareh | 2020-03-20 | No | Unclear | No | Yes | Unclear | Unclear | Yes | No | Unclear | High |
| **Kuwait** | | | | | | | | | | | |
| Alali | 2020-06-26 | No | Yes | No | Yes | Unclear | No | Yes | No | Unclear | High |
| **Libya** | | | | | | | | | | | |
| Kammon | 2020-05-18 | Yes | Yes | No | Yes | Yes | No | Yes | No | Yes | High |
| Kammon | 2020-05-18 | No | Unclear | No | Yes | Yes | No | Yes | No | Yes | High |

| Article Author (Study Organization) | Sampling End Date (YMD) | Item 1: Appropriate sample frame* | Item 2: Probability sampling method | Item 3: Adequate sample size | Item 4: Subjects & setting described | Item 5: Good coverage of sample | Item 6: Sens > 90% & spec > 95% | Item 7: Same tests for all subjects | Item 8: Appropriate analysis | Item 9: Adequate response rate | Overall Risk of Bias† |
|---|---|---|---|---|---|---|---|---|---|---|---|
| Elfakhri | 2020-05-07 | Yes | Yes | Yes | Yes | Yes | No | Yes | No | Unclear | Moderate |
| **Qatar** | | | | | | | | | | | |
| Jeremijenko | 2020-09-09 | Yes | No | Yes | Yes | Unclear | Unclear | Yes | No | Unclear | High |
| MohamedH.Al-Thani | 2020-09-09 | Yes | Yes | Yes | Yes | Yes | Yes | Yes | No | Unclear | Moderate |
| Nasrallah | 2020-09-09 | No | Unclear | No | No | Unclear | No | Yes | No | Unclear | High |
| **Saudi Arabia** | | | | | | | | | | | |
| Mahallawi | 2020-07-15 | No | No | Yes | Yes | Yes | Unclear | Yes | No | Unclear | High |
| Alharbi | 2020-06-17 | Yes | No | No | No | Unclear | No | Yes | No | Unclear | High |
| Alserehi | 2020-05-30 | Yes | No | Yes | No | Unclear | Unclear | Yes | No | Unclear | Moderate |
| **Turkey** | | | | | | | | | | | |
| Sozcu | 2020-06-25 | Yes | Unclear | Yes | No | Unclear | Unclear | Unclear | No | Unclear | Unclear |
| Alkurt | 2020-06-06 | Yes | No | Yes | Yes | Yes | Yes | Yes | No | Unclear | Moderate |
| **South Asia** | | | | | | | | | | | |
| **India** | | | | | | | | | | | |
| Khan | 2020-11-04 | Unclear | Unclear | Unclear | Unclear | Unclear | Unclear | Unclear | Unclear | Unclear | Low |
| Kumar | 2020-10-24 | Yes | Yes | Yes | No | Unclear | Unclear | Unclear | No | Unclear | Unclear |
| Sharma | 2020-10-21 | Yes | Yes | Yes | Yes | Yes | Yes | Yes | Yes | Unclear | Low |
| Ramanath | 2020-10-18 | Yes | Yes | Yes | No | Unclear | Unclear | Unclear | Unclear | Unclear | Unclear |
| Banerjee | 2020-10-17 | Yes | Yes | Yes | No | Yes | Yes | Yes | Yes | No | Moderate |
| Babu | 2020-09-16 | Yes | No | Yes | Yes | Unclear | Yes | Yes | Yes | Unclear | Moderate |
| Sharma | 2020-09-07 | Yes | Yes | Yes | Yes | Yes | Yes | Yes | Yes | Unclear | Low |

| Article Author (Study Organization) | Sampling End Date (YMD) | Item 1: Appropriate sample frame* | Item 2: Probability sampling method | Item 3: Adequate sample size | Item 4: Subjects & setting described | Item 5: Good coverage of sample | Item 6: Sens > 90% & spec > 95% | Item 7: Same tests for all subjects | Item 8: Appropriate analysis | Item 9: Adequate response rate | Overall Risk of Bias† |
|---|---|---|---|---|---|---|---|---|---|---|---|
| Goenka | 2020-08-31 | No | No | No | Yes | Unclear | Unclear | Yes | No | Yes | High |
| Kshatri | 2020-08-31 | Yes | Yes | Yes | Yes | Yes | Unclear | Yes | Yes | No | High |
| Singh | 2020-08-30 | Unclear | Unclear | Yes | No | Unclear | Unclear | Unclear | No | Unclear | Unclear |
| Mohanan | 2020-08-29 | Yes | Yes | Yes | Yes | Yes | No | Yes | Yes | No | Moderate |
| Goenka | 2020-08-23 | Yes | Yes | Yes | Yes | Unclear | Yes | Yes | No | No | Moderate |
| Dixit | 2020-08-19 | Yes | Yes | Yes | Yes | Yes | Unclear | Yes | No | Yes | Moderate |
| Siddiqui | 2020-08-15 | No | No | Yes | Yes | Unclear | Yes | Yes | No | Unclear | High |
| Ray | 2020-08-08 | No | No | No | Yes | Yes | No | Yes | No | Unclear | High |
| Satpati | 2020-08-08 | Yes | Yes | No | Yes | Yes | Yes | Yes | No | Unclear | Moderate |
| Bedi | 2020-08-07 | Yes | Unclear | Yes | No | Yes | Unclear | Unclear | Unclear | Unclear | Unclear |
| Sharma | 2020-08-07 | Yes | Yes | Yes | Yes | Yes | Yes | Yes | Yes | Unclear | Low |
| Dey | 2020-08-07 | Unclear | Unclear | Yes | No | Unclear | Yes | Yes | Yes | Unclear | Unclear |
| Ghose | 2020-08-05 | Yes | Yes | Yes | Yes | Yes | Unclear | Yes | Yes | Yes | Moderate |
| Kumar | 2020-07-24 | Yes | No | Yes | Yes | Yes | Unclear | Yes | No | Unclear | High |
| Malani | 2020-07-19 | Yes | Yes | Yes | Yes | Yes | Yes | Yes | Yes | Unclear | Low |
| Malani | 2020-07-19 | Yes | Yes | Yes | Yes | Yes | Yes | Yes | Yes | Unclear | Low |
| Husain | 2020-07-19 | No | Yes | Yes | No | Unclear | Unclear | Yes | No | Unclear | High |
| India.com News Desk | 2020-07-19 | Yes | Yes | Yes | No | No | Unclear | Unclear | No | Unclear | High |
| Khan | 2020-07-15 | No | No | Yes | Yes | Yes | Yes | Yes | Yes | Yes | Moderate |
| Mishra | 2020-07-15 | No | Yes | Yes | No | Unclear | Unclear | Yes | No | Unclear | High |

| Article Author (Study Organization) | Sampling End Date (YMD) | Item 1: Appropriate sample frame* | Item 2: Probability sampling method | Item 3: Adequate sample size | Item 4: Subjects & setting described | Item 5: Good coverage of sample | Item 6: Sens > 90% & spec > 95% | Item 7: Same tests for all subjects | Item 8: Appropriate analysis | Item 9: Adequate response rate | Overall Risk of Bias† |
|---|---|---|---|---|---|---|---|---|---|---|---|
| Unclear | 2020-07-11 | Yes | No | Yes | No | Unclear | Unclear | Unclear | No | Unclear | Unclear |
| Saxena | 2020-07-10 | Yes | Yes | Yes | No | Unclear | Yes | Yes | No | Unclear | Moderate |
| Dey | 2020-07-10 | Unclear | Unclear | Yes | No | Unclear | Yes | Yes | Yes | Unclear | Unclear |
| Prakash | 2020-07-07 | No | No | Yes | Yes | Unclear | Yes | Yes | No | Unclear | High |
| Kumar | 2020-06-30 | No | Unclear | Yes | Yes | Unclear | Yes | Yes | No | Unclear | High |
| Singhal | 2020-06-30 | No | No | No | No | No | Yes | No | No | No | High |
| Khan | 2020-06-29 | Yes | No | Yes | Yes | Unclear | Yes | Yes | No | No | Moderate |
| Kumar | 2020-06-12 | Yes | Unclear | Yes | No | Unclear | Unclear | Unclear | No | Unclear | Unclear |
| Rao | 2020-06-06 | Yes | Unclear | Yes | No | Unclear | Unclear | Yes | No | Unclear | Unclear |
| Murhekar | 2020-06-04 | Yes | Yes | Yes | Yes | Yes | Yes | Yes | Yes | Yes | Low |
| Kher | 2020-05-25 | Yes | Yes | Yes | No | Yes | Unclear | Yes | No | Unclear | Moderate |
| **Pakistan** | | | | | | | | | | | |
| Nisar | 2020-08-22 | Yes | Yes | No | Yes | Yes | Yes | Yes | No | No | Moderate |
| Nisar | 2020-08-22 | Yes | Yes | No | Yes | Yes | Yes | Yes | No | No | Moderate |
| Zaidi | 2020-07-15 | No | No | Yes | No | Unclear | Unclear | Yes | No | Unclear | High |
| Younas | 2020-07-15 | No | No | No | Yes | No | Unclear | No | No | Unclear | High |
| Noor | 2020-07-15 | Yes | Yes | Yes | Yes | Unclear | No | Yes | No | Yes | Moderate |
| Nisar | 2020-07-11 | Yes | Yes | No | Yes | Yes | Yes | Yes | No | No | Moderate |
| Nisar | 2020-07-11 | Yes | Yes | No | Yes | Yes | Yes | Yes | No | No | Moderate |
| Haq | 2020-06-29 | Yes | No | Yes | Yes | Unclear | Yes | Yes | No | Unclear | Moderate |
| Chughtai | 2020-05-30 | Yes | No | No | Yes | No | Yes | Yes | No | Unclear | High |

| Article Author (Study Organization) | Sampling End Date (YMD) | Item 1: Appropriate sample frame* | Item 2: Probability sampling method | Item 3: Adequate sample size | Item 4: Subjects & setting described | Item 5: Good coverage of sample | Item 6: Sens > 90% & spec > 95% | Item 7: Same tests for all subjects | Item 8: Appropriate analysis | Item 9: Adequate response rate | Overall Risk of Bias† |
|---|---|---|---|---|---|---|---|---|---|---|---|
| Nisar | 2020-04-25 | Yes | Yes | No | Yes | Yes | Yes | Yes | No | No | Moderate |
| Nisar | 2020-04-25 | Yes | Yes | No | Yes | Yes | Yes | Yes | No | No | Moderate |
| **Southeast Asia, East Asia, an Oceania** | | | | | | | | | | | |
| **China** | | | | | | | | | | | |
| Li | 2020-10-15 | Yes | Unclear | Yes | No | Unclear | Unclear | Yes | No | Unclear | High |
| Wang | 2020-08-31 | No | No | Yes | Yes | Yes | Unclear | Yes | No | Unclear | Moderate |
| He | 2020-06-10 | No | No | Yes | Yes | Unclear | No | Yes | No | Unclear | High |
| He | 2020-05-29 | Yes | Unclear | Yes | Yes | Unclear | Unclear | Yes | No | Unclear | Unclear |
| He | 2020-05-29 | Yes | Unclear | Yes | Yes | Unclear | Unclear | Yes | No | Unclear | Unclear |
| Liu | 2020-05-26 | Yes | Unclear | Yes | Yes | Unclear | No | Yes | No | Unclear | High |
| Pan | 2020-05-26 | Yes | Yes | Yes | Yes | Unclear | Yes | Yes | No | Unclear | Moderate |
| Tong | 2020-05-15 | Yes | Unclear | No | No | No | Unclear | Yes | No | Unclear | High |
| Liang | 2020-04-30 | Yes | No | Yes | Yes | Unclear | No | Yes | No | Yes | High |
| Liang | 2020-04-30 | Yes | Unclear | Yes | Yes | Unclear | No | Yes | No | Yes | High |
| Chang | 2020-04-30 | No | Yes | Yes | Yes | Yes | Unclear | Yes | No | Unclear | High |
| Liu | 2020-04-29 | Yes | No | Yes | Yes | Yes | Yes | Yes | No | Yes | Moderate |
| Liu | 2020-04-29 | Yes | Unclear | Yes | Yes | Yes | Yes | Yes | No | Yes | Moderate |
| Liu | 2020-04-29 | Yes | No | Yes | Yes | Yes | Yes | Yes | No | Yes | Moderate |
| Lei | 2020-04-28 | No | No | Yes | No | Unclear | No | Yes | No | Unclear | High |
| Ling | 2020-04-28 | Yes | No | Yes | Yes | Yes | No | Yes | Yes | Unclear | Moderate |
| Want | 2020-04-24 | Yes | Unclear | No | Yes | Yes | Unclear | Unclear | Unclear | Unclear | Unclear |

| Article Author (Study Organization) | Sampling End Date (YMD) | Item 1: Appropriate sample frame* | Item 2: Probability sampling method | Item 3: Adequate sample size | Item 4: Subjects & setting described | Item 5: Good coverage of sample | Item 6: Sens > 90% & spec > 95% | Item 7: Same tests for all subjects | Item 8: Appropriate analysis | Item 9: Adequate response rate | Overall Risk of Bias† |
|---|---|---|---|---|---|---|---|---|---|---|---|
| Liu | 2020-04-21 | No | No | No | Yes | Unclear | Yes | Yes | No | Unclear | High |
| Wang | 2020-04-18 | Yes | Yes | Yes | Yes | Yes | No | Unclear | No | Unclear | Moderate |
| Wu | 2020-04-15 | Yes | Unclear | No | No | Unclear | Unclear | Yes | No | Unclear | High |
| Wu | 2020-04-15 | No | Unclear | Yes | No | Unclear | Unclear | Yes | No | Unclear | High |
| Xu | 2020-04-10 | Yes | No | Yes | Yes | Unclear | Unclear | Yes | No | Unclear | High |
| Xu | 2020-04-10 | Yes | No | Yes | Yes | Unclear | Unclear | Yes | No | Unclear | High |
| Xu | 2020-04-02 | No | Yes | Yes | Yes | No | Unclear | Yes | No | Unclear | High |
| Zhang | 2020-03-27 | No | No | No | Yes | Yes | Unclear | Yes | No | Unclear | High |
| Zhou | 2020-03-25 | Yes | No | Yes | Yes | Yes | Unclear | Yes | No | Yes | Moderate |
| Li | 2020-03-22 | Unclear | No | No | No | Unclear | Yes | Yes | No | Unclear | High |
| Tang | 2020-03-21 | Yes | Yes | Yes | Yes | Yes | No | Yes | No | Yes | Moderate |
| Tu | 2020-03-20 | Yes | No | No | No | Unclear | Unclear | Yes | No | Unclear | High |
| Zhang | 2020-03-18 | Yes | No | Yes | No | Unclear | Unclear | Yes | No | Unclear | High |
| Xiong | 2020-03-17 | Unclear | Unclear | Yes | Yes | Unclear | Unclear | Yes | No | Unclear | Unclear |
| To | 2020-03-17 | Yes | Yes | Yes | No | Yes | No | Yes | No | Unclear | Moderate |
| Chang | 2020-03-15 | No | Yes | Yes | Yes | Yes | Unclear | Yes | No | Unclear | High |
| Chang | 2020-03-15 | No | Yes | Yes | Yes | Yes | Unclear | Yes | No | Unclear | High |
| Zhao | 2020-02-21 | Yes | No | Yes | No | Unclear | Unclear | Yes | No | Yes | High |
| Chen | 2020-02-19 | No | Yes | No | Yes | Yes | Yes | Yes | No | Yes | High |
| Zhang | 2020-02-16 | No | No | No | Yes | Unclear | Yes | Yes | No | Unclear | High |
| Zhang | 2020-02-16 | No | No | No | Yes | Unclear | Yes | Yes | No | Unclear | High |

| Article Author (Study Organization) | Sampling End Date (YMD) | Item 1: Appropriate sample frame* | Item 2: Probability sampling method | Item 3: Adequate sample size | Item 4: Subjects & setting described | Item 5: Good coverage of sample | Item 6: Sens > 90% & spec > 95% | Item 7: Same tests for all subjects | Item 8: Appropriate analysis | Item 9: Adequate response rate | Overall Risk of Bias† |
|---|---|---|---|---|---|---|---|---|---|---|---|
| Zhang | 2020-02-16 | No | No | No | Yes | Unclear | Yes | Yes | No | Unclear | High |
| To | 2020-02-13 | No | No | No | No | Unclear | No | Yes | No | Unclear | High |
| To | 2020-01-31 | No | No | Yes | No | Unclear | No | Yes | No | Unclear | High |
| Hallowell | 2020-01-28 | No | Yes | No | Yes | Yes | Unclear | Yes | No | Yes | High |
| **Malaysia** | | | | | | | | | | | |
| Sam | 2020-06-06 | No | No | Yes | No | Unclear | Yes | Yes | No | Unclear | High |
| **Thailand** | | | | | | | | | | | |
| Nopsopon | 2020-06-26 | Yes | No | Yes | Yes | Yes | Yes | Yes | No | Unclear | Moderate |
| Nopsopon | 2020-05-17 | No | No | Yes | Yes | Yes | Yes | Yes | No | Yes | Moderate |
| **Vietnam** | | | | | | | | | | | |
| Chau | 2020-08-30 | Yes | No | No | Yes | Unclear | Yes | Yes | No | No | High |
| **Sub-Saharan Africa** | | | | | | | | | | | |
| **Congo** | | | | | | | | | | | |
| Batchi-Bouyou | 2020-07-31 | Unclear | No | Yes | Yes | Unclear | Yes | Yes | No | Unclear | High |
| **Ethiopia** | | | | | | | | | | | |
| Kempen | 2020-05-21 | No | No | No | Yes | Yes | Yes | Yes | No | Unclear | High |
| Alemu | 2020-04-28 | Yes | Yes | No | Yes | Unclear | No | Yes | No | Unclear | Moderate |
| **Kenya** | | | | | | | | | | | |
| Adetifa | 2020-09-30 | No | Yes | Yes | Yes | Yes | Yes | Yes | Yes | Unclear | Low |
| **Malawi** | | | | | | | | | | | |
| Chibwana | 2020-06-19 | Yes | No | No | Yes | Yes | No | Yes | No | Unclear | High |

| Article Author (Study Organization) | Sampling End Date (YMD) | Item 1: Appropriate sample frame* | Item 2: Probability sampling method | Item 3: Adequate sample size | Item 4: Subjects & setting described | Item 5: Good coverage of sample | Item 6: Sens > 90% & spec > 95% | Item 7: Same tests for all subjects | Item 8: Appropriate analysis | Item 9: Adequate response rate | Overall Risk of Bias[†] |
|---|---|---|---|---|---|---|---|---|---|---|---|
| **Nigeria** | | | | | | | | | | | |
| Majiya | 2020-06-30 | Yes | Yes | No | Yes | Yes | Yes | Yes | No | Unclear | Moderate |
| Asuquo | 2020-06-24 | Unclear | Yes | No | No | No | Unclear | Yes | No | Unclear | High |
| **South Africa** | | | | | | | | | | | |
| Shaw | 2020-09-04 | No | No | No | Yes | Unclear | Yes | Yes | No | No | High |
| Hsiao | 2020-08-07 | No | No | Yes | Yes | Unclear | Yes | Yes | No | Unclear | High |
| **Togo** | | | | | | | | | | | |
| Halatoko | 2020-05-08 | Yes | Yes | Yes | Yes | Unclear | No | Yes | No | Unclear | High |

#### S4 Table. Summary of unadjusted meta-analysis results

| Factor | Reference Group | Comparison Group | Number of Studies | Risk Ratio (95% CI)[a] | Heterogeneity ($I^2$) |
|---|---|---|---|---|---|
| Age | Adults (18-64) | Youth (0-17) | 82 | 0.91 (0.83-1.00) | 86.5% |
| | Adults (18-64) | Seniors (65+) | 127 | 0.84 (0.77-0.92) | 92.7% |
| Sex/Gender | Female | Male | 129 | 1.02 (0.98-1.06) | 87.1% |
| Race | Caucasian | Black | 19 | 2.46 (2.14-2.83) | 87.7% |
| | Caucasian | Asian | 17 | 1.77 (1.44-2.17) | 90.8% |
| | Caucasian | Indigenous | 8 | 2.00 (1.14-3.50) | 81.1% |
| | Caucasian | Multiple/other | 18 | 1.55 (1.37-1.76) | 63.5% |
| Close contact with COVID-19 patients | Individuals with no close contact | Individuals with close contact | 35 | 1.97 (1.32-2.92) | 98.4% |
| | Health care workers with no close contact | Health care workers with close contact | 44 | 1.57 (1.33-1.85) | 94.8% |
| Health care worker status | Non-health care workers and caregivers | Health care workers and caregivers | 19 | 1.24 (0.99-1.54) | 95.2% |

[a]Using adjusted seroprevalence estimates. Abbreviations: CI= confidence interval.

#### S5 Table. Summary of serological tests used in included seroprevalence studies

| Manufacturer | Test Name | Isotype | No. of studies |
|---|---|---|---|
| Abbott Laboratories | Abbott Architect SARS-CoV-2 IgG | IgG | 124 |
| | Abbott Alinity SARS-COV-2 IgG | IgG | 1 |
| ACRO Biotech Inc. | Acro Biotech COVID-19 Rapid POC test | IgM | 2 |
| | | IgG | 1 |
| | Not available | IgG; IgM | 2 |
| Artron Laboratories Inc. | One Step Novel Coronavirus (COVID-19) IgM/IgG Antibody Test | IgG; IgM | 4 |
| | | IgG | 1 |
| | | IgM | 1 |
| Aurora Biomed Inc | COVID-19 IgG/IgM Rapid Test | IgG; IgM | 1 |
| Beijing Wantai Biological Pharmacy Enterprise Co., Ltd | Wantai SARS-CoV-2 Ab ELISA | Total Ig | 9 |
| | Wantai SARS-CoV-2 IgM ELISA | IgM | 2 |
| | Wantai SARS-CoV-2 Ab ELISA | IgG | 1 |
| BioMedomics, Inc. | COVID-19 IgM-IgG Dual Antibody Rapid Test | IgG | 4 |
| | | IgM | 4 |
| | | IgG; IgM | 3 |
| Cellex Inc. | COVID-19 IgG/IgM Rapid Test Cassette | IgG; IgM | 1 |
| CTK Biotech, Inc. | OnSite COVID-19 IgG/IgM Rapid Test | IgG; IgM | 1 |
| DiaSorin SpA | Liaison SARS-CoV-2 S1/S2 IgG | IgG | 19 |
| DRG International, Inc. | Novel Coronavirus COVID-19 IgG | IgG | 1 |
| Euroimmun AG | Anti-SARS-CoV-2 ELISA (IgG) (manual, automated) | IgG | 35 |
| | EI SARS-Cov-2 IgG assay | IgG | 34 |
| | Anti-SARS-CoV-2 ELISA (IgA) (manual, automated) | IgA | 6 |
| | Anti-SARS-CoV-2 ELISA (IgA) (manual, automated); Anti-SARS-CoV-2 ELISA (IgG) (manual, automated) | IgA; IgG | 7 |
| | Euroimmun EIA | IgA | 4 |
| | Euroimmun | IgG | 1 |
| Epitope Diagnostics, Inc. | EDI™ Novel Coronavirus COVID-19 IgG ELISA Kit | IgG | 7 |
| | EDI™ Novel Coronavirus COVID-19 IgM ELISA Kit | IgM | 3 |
| | EDI™ Novel Coronavirus COVID-19 IgM ELISA Kit; EDI™ Novel Coronavirus COVID-19 IgG ELISA Kit | IgG; IgM | 2 |
| ET Healthcare | Pylon COVID-19 IgG/IgM assay | IgG | 1 |
| | | IgG; IgM | 1 |
| | | IgM | 1 |
| Guangzhou Wondfo Biotech Co., Ltd | Wondfo SARS-CoV-2 Antibody Test (lateral flow method) | IgG; IgM | 12 |
| | Finecare SARS-CoV-2 Antibody test | Total Ig | 4 |
| Hanghzhou AllTest Biotech Co., Ltd | 2019-nCoV IgG/IgM Rapid Test Cassette | IgG | 2 |
| | | IgG; IgM | 2 |
| | | IgM | 1 |
| Hangzhou Biotest Biotech Co., Ltd | COVID-19 IgG/IgM Rapid Test Cassette | IgG; IgM | 2 |
| Hangzhou Clongene Biotech Co., Ltd | 2019-nCoV IgG/IgM Rapid Test | IgG | 1 |
| | | IgG; IgM | 1 |
| | | IgM | 1 |
| Healgen | COVID-19 IgG/IgM Rapid Test Cassette | IgG | 1 |
| | | IgG; IgM | 1 |
| | | IgM | 1 |
| ImmunoDiagnostics limited | SARS-CoV-2 NP IgG ELISA Kit | IgG | 1 |
| | SARS-CoV-2 NP IgM ELISA Kit | IgM | 1 |
| Innovita Biological Technology Co. Ltd | 2019-nCoV Ab Test (Colloidal Gold) (IgM/IgG Whole Blood/Serum/Plasma Combo) | IgG; IgM | 6 |
| | | IgG | 3 |
| | SARS-CoV-2 IgG Ab Rapid Test | IgM | 2 |
| Mikrogen GmbH | recomWell SARS-CoV-2 IgG ELISA | IgG | 1 |
| Mologic Ltd | SARS-CoV-2 ELISA Serology | IgG | 1 |
| Multi-G B.V. | COVID-19 IgG/IgM Ab Test Cassette (Whole Blood/Serum/Plasma) | IgG | 1 |
| nal von minden GmbH | NADAL® COVID-19 IgG/IgM Test | IgG | 1 |
| | | IgM | 1 |
| Ortho Clinical Diagnostics | VITROS® Immunodiagnostic Products Anti-SARS-CoV-2 Total | Total Ig | 5 |
| | VITROS® Immunodiagnostic Products Anti-SARS-CoV-2 IgG | IgG | 12 |

| Manufacturer | Test Name | Isotype | No. of studies |
|---|---|---|---|
| PCL Inc. | PCL COVID19 IgG/IgM Rapid Gold | IgG; IgM | 1 |
| Phamatech | COVID19 RAPID TEST | IgG; IgM | 1 |
| Premier Biotech | COVID-19 IgG/IgM Rapid Test Cassette | IgG; IgM | 2 |
| PRIMA Lab S.A. | PRIMA COVID-19 IgG/IgM Rapid Test | IgG; IgM | 3 |
| | | IgG | 2 |
| RayBiotech | Novel Coronavirus IgG antibody test kit (colloidal gold method) | IgG | 1 |
| Roche Diagnostics, Basel, Switzerland | Elecsys Anti-SARS-CoV-2 | Total Ig | 17 |
| | | IgG; IgM | 20 |
| | Anti-SARS-CoV-2 ELISA (IgA) (manual, automated) | IgA; IgG; IgM | 4 |
| Shanghai Kehua Bio-engineering Co., Ltd | Diagnostic kit for SARS-CoV-2 IgM/IgG Antibody (Colloidal Gold) | IgG; IgM | 3 |
| Shenzhen Yahuilong Biotechnology Co Ltd | 2019-nCoV IgG/IgM Antibody Detection Kit | IgG | 4 |
| | | IgM | 4 |
| Shenzhen Yhlo Biotech Co. Ltd | SARS-CoV-2 IgM and IgG chemiluminescence immunoassay | IgG | 7 |
| | | IgM | 4 |
| | iFlash-SARS-CoV-2 IgG | IgG | 2 |
| | SARS-CoV-2 IgM/IgG CLIA kits | IgG | 1 |
| | | IgM | 1 |
| | iFlash-SARS-CoV-2 IgM | IgM | 1 |
| Siemens | Atellica® IM SARS-CoV-2 Total (COV2T) | Total Ig | 4 |
| Snibe Co., Ltd (Shenzhen New Industries Biomedical Engineering Co., Ltd) | MAGLUMI 2019-nCoV IgG (CLIA) | IgG | 10 |
| | MAGLUMI 2019-nCoV IgM (CLIA) | IgM | 7 |
| Sugentech, Inc. | SGTi-flex COVID-19 IgM/IgG (manual) | IgG; IgM | 1 |
| SureScreen Diagnostics | COVID-19 IgG/IgM Rapid Test Cassette | IgG | 1 |
| Tangshan Yingnuote Biotechnology Enterprise Co., Ltd. | Colloidal Gold Immunochromatography Assay (CGIA) kit | IgG; IgM | 1 |
| TestsealabsE | SARS-COV-2-IgG/IgM Test Cassette | IgG; IgM | 2 |
| | | IgG | 1 |
| | | IgM | 1 |
| UCP Biosciences | Coronavirus IgG/IgM Antibody (COVID-19) Test Cassette | IgG; IgM | 2 |
| | | IgG | 1 |
| | | IgM | 1 |
| Vircell S.L. | COVID-19 ELISA IgG | IgG | 3 |
| | COVID-19 ELISA IgM+IgA | IgA; IgM | 3 |
| VivaChek Biotech (Hangzhou) Co., Ltd | VivaDiag COVID-19 IgM/IgG Rapid Test | IgG; IgM | 3 |
| Wadsworth Center, New York State Department of Health | New York SARS-CoV Microsphere Immunoassay for Antibody Detection | Total Ig | 1 |
| Xiamen Wantai Kairui Biotechnology | Not available | Total Ig | 7 |
| Zhejiang Orient Gene Biotech | COVID-19 IgG/IgM Rapid Test | IgG | 9 |
| | | IgG; IgM | 8 |
| | | IgM | 7 |
| Zhuhai Livzon Diagnostics Inc | 2019-nCoV IgG/IgM Antibody Detection Kit | IgG | 4 |
| | | IgM | 4 |
| | | IgG; IgM | 2 |
| | Diagnostic Kit for IgM / IgG Antibody to Coronavirus (SARS-CoV-2) (Lateral Flow) | IgG | 2 |
| | | IgG; IgM | 2 |
| No independent evaluation available | Multiple tests used | - | 275 |
| | Author designed or validated test, not in Finddx database | - | 139 |
| | Not enough test information reported to match | - | 89 |
| | Commercially available but not in the Finddx database | - | 66 |
| | Evaluations for separate isotypes, review authors report combined positivity rate (and/or isotype) | - | 12 |
| | Evaluation for combined isotype, study examined individual isotypes | - | 9 |

### S6 Table. Summary of meta-regression results

| | Analysis using uncorrected seroprevalence estimates | | | Analysis using corrected seroprevalence estimates | | |
|---|---|---|---|---|---|---|
| Study Characteristics | Change in Seroprevalence (coefficient β)[a,b] (95% CI) | Risk ratio (95%CI) | p-value | Change in Seroprevalence (coefficient β)[a,c] (95% CI) | Risk ratio (95%CI) | p-value |
| **Risk of Bias** | | | | | | |
| Low | Reference | - | - | Reference | - | - |
| Moderate | -0.02 (-0.33, 0.30) | 0.98 (0.72, 1.35) | 0.92 | -0.51 (-0.83, -0.20) | 0.60 (0.44, 0.82) | 0.001 |
| High | 0.02 (-0.30, 0.35) | 1.02 (0.74, 1.42) | 0.89 | -0.43 (-0.76, -0.10) | 0.65 (0.47, 0.90) | 0.01 |
| Unclear | -0.30 (-0.69, 0.08) | 0.74 (0.50, 1.09) | 0.12 | -0.96 (-1.49, -0.43) | 0.38 (0.22, 0.65) | <0.001 |
| **Population Group** | | | | | | |
| Household and community sample | Reference | - | - | Reference | - | - |
| Blood donors | -0.15 (-0.35, 0.06) | 0.86 (0.70, 1.06) | 0.16 | 0.04 (-0.20, 0.27) | 1.04 (0.82, 1.31) | 0.77 |
| Residual sera | -0.33 (-0.49, -0.16) | 0.72 (0.61, 0.85) | <0.001 | -0.12 (-0.30, 0.06) | 0.89 (0.74, 1.06) | 0.19 |
| **Estimate Grade** | | | | | | |
| National | Reference | - | - | Reference | - | - |
| Regional | -0.02 (-0.23, 0.19) | 0.98 (0.79, 1.20) | 0.84 | 0.50 (0.26, 0.74) | 1.65 (1.30, 2.09) | <0.001 |
| Local | 0.43 (0.22, 0.64) | 1.54 (1.25, 1.90) | <0.001 | 0.76 (0.52, 1.00) | 2.14 (1.68, 2.72) | <0.001 |
| Sublocal | 0.50 (0.16, 0.85) | 1.66 (1.17, 2.34) | 0.004 | 0.66 (0.21, 1.11) | 1.93 (1.23, 3.03) | 0.004 |
| **Cases per 100,000 population** | | | | | | |
| Cases per 100,000 | 0.43 (0.31, 0.55) | 1.54 (1.37, 1.73) | <0.001 | 0.30 (0.18, 0.43) | 1.35 (1.19, 1.53) | <0.001 |
| **GBD Region** | | | | | | |
| High-Income | Reference | - | - | Reference | - | - |
| North Africa and Middle East | 0.65 (-0.03, 1.32) | 1.91 (0.97, 3.75) | 0.06 | -0.22 (-0.91, 0.47) | 0.80 (0.40, 1.60) | 0.53 |
| Southeast Asia, East Asia, and Oceania | -1.28 (-1.67, -0.88) | 0.28 (0.19, 0.41) | <0.001 | -1.74 (-2.41, -1.07) | 0.18 (0.09, 0.34) | <0.001 |
| Central Europe, Eastern Europe, and Central Asia | 0.45 (0.04, 0.87) | 1.57 (1.04, 2.38) | 0.03 | 1.04 (0.56, 1.52) | 2.83 (1.75, 4.55) | <0.001 |
| Latin America and Caribbean | 0.39 (0.16, 0.63) | 1.48 (1.17, 1.87) | <0.001 | 1.00 (0.73, 1.26) | 2.71 (2.07, 3.54) | <0.001 |
| South Asia | 1.20 (0.93, 1.47) | 3.30 (2.52, 4.33) | <0.001 | 1.04 (0.74, 1.35) | 2.84 (2.09, 3.85) | <0.001 |
| Sub-Saharan Africa | 1.34 (0.75, 1.93) | 3.81 (2.11, 6.89) | <0.001 | 1.61 (1.06, 2.16) | 5.01 (2.89, 8.69) | <0.001 |

[a]The regression coefficient β refers to the change in the log seroprevalence of antibodies to SARS-CoV-2 given changes in the covariate. [b]Details of uncorrected model: Intercept coefficient β -3.45 (95%CI -3.76, -3.13); Mixed-Effects Model (k = 570); tau^2 0.4759 (SE = 0.1331); I^2=99.64%; R^2=21.26%. Test of Moderators (coefficients 2:16): QM(df = 15) = 263.86, p-val < 0.0001. [c]Details of corrected model: Intercept coefficient β -3.45 (95%CI -3.78, -3.12); Mixed-Effects Model (k = 417); tau^2 =0.3574(SE =0.0791); I^2=98.94%; R^2=63.97%. Test of Moderators (coefficients 2:16): QM(df = 15) = 322.6942, p-val < 0.0001. Abbreviations: B = beta; CI = confidence interval; exp = exponentiated; GBD = global burden of disease.

**Supplementary figures**

**S1 Figure. Map of serosurvey distribution by global burden of disease region**

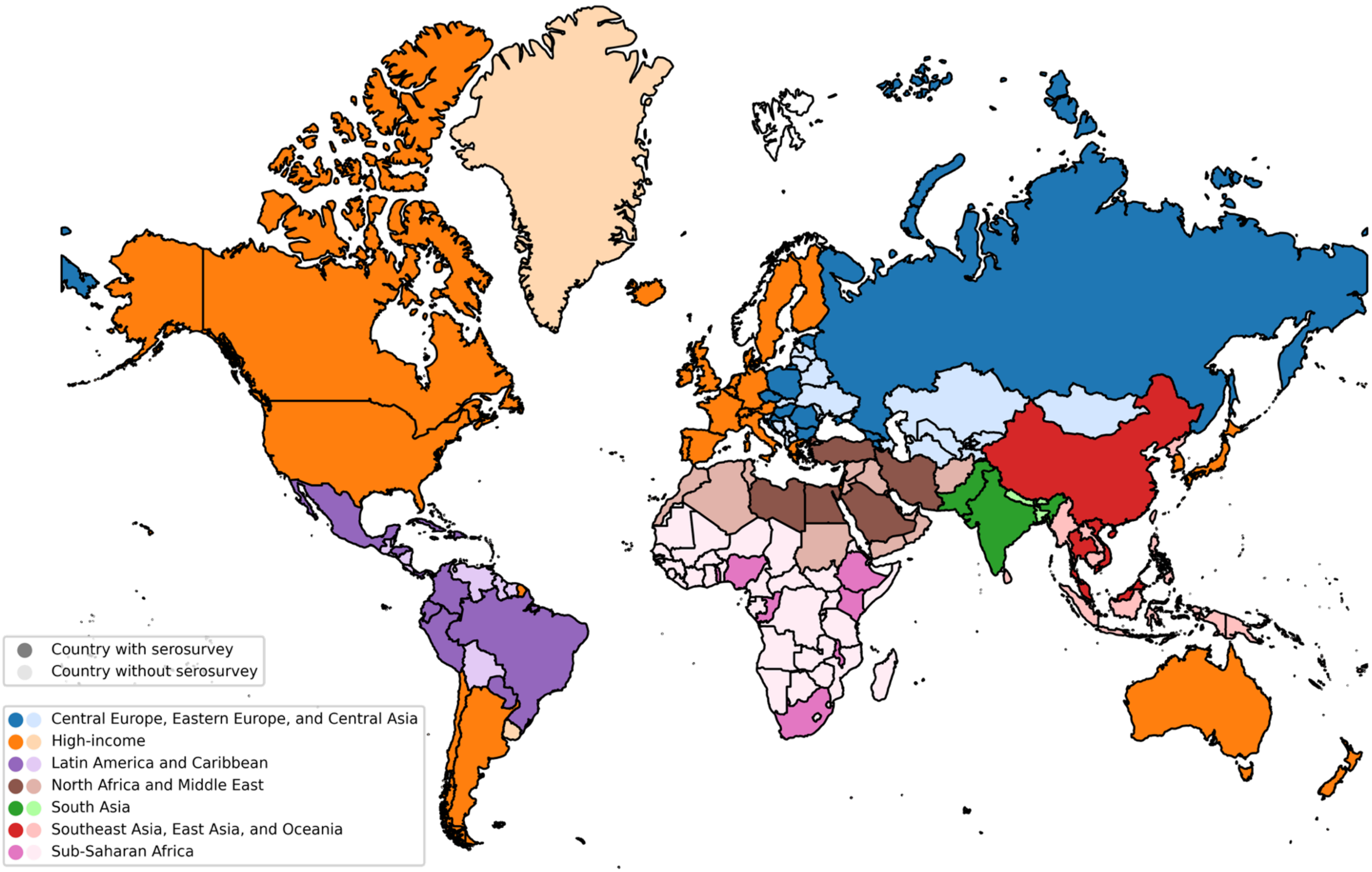

The number of countries reporting any serosurvey in each GBD region were: Central Europe, Eastern Europe, and Central Asia (n=26); in High Income regions (n=747); in Latin America and Caribbean (n=69); in North Africa and Middle East (n=21); in South Asia (n=49); in Southeast Asia, East Asia, and Oceania (n=46); and in Sub-Saharan Africa (n=10).

**S2 Figure. Seroprevalence to cumulative case incidence ratios using cumulative incidence nine days prior to the serosurvey end date**

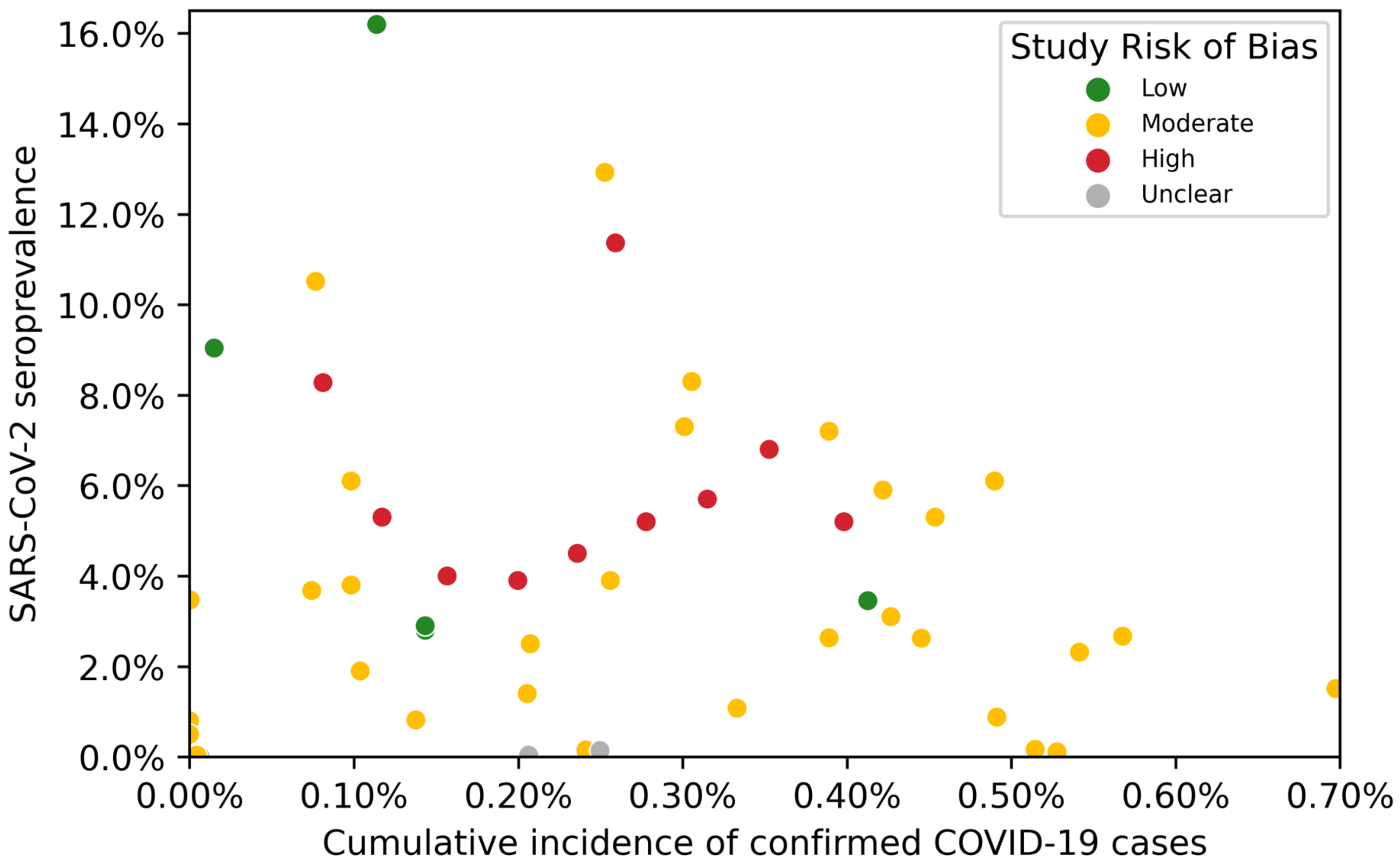

47. Canadian Blood Services CBS. COVID-19 Seroprevalence Report 4 – January 12, 2021. 2021 Jan. Available: N/A

63. Storgaard SF, Eiset AH, Abdullahi F, Wejse C. First wave of COVID-19 did not reach the homeless population in Aarhus. Dan Med J. 2020;67.

203. Snoeck C. Prevalence of SARS-CoV-2 infection in the Luxembourgish population: The CON-VINCE study. Pre-print.

287. Black JT, Haroon U, Barnes SC, Mayaleh S, Bagul A. Seroprevalence of SARS-CoV-2 IgG antibodies in the current COVID-19 pandemic amongst co-workers at a UK renal transplant centre. Transplant International. n/a. doi:10.1111/tri.13791

483. Filho LA, Szwarcwald CL. Seroprevalence of anti-SARS-CoV-2 among blood donors in Rio de Janeiro, Brazil. : 10.

579. Xu R, Huang J, Duan C, Liao Q, Shan Z, Wang M, et al. Low prevalence of antibodies against SARS-CoV-2 among voluntary blood donors in Guangzhou, China. Journal of Medical Virology. n/a. doi:10.1002/jmv.26445

592. Sam I-C, Chong YM, Tan CW, Chan YF. Low postpandemic wave SARS-CoV-2 seroprevalence in Kuala Lumpur and Selangor, Malaysia. Journal of Medical Virology. n/a. doi:10.1002/jmv.26426
